# Immune Correlates Analysis of the mRNA-1273 COVID-19 Vaccine Efficacy Trial

**DOI:** 10.1101/2021.08.09.21261290

**Authors:** Peter B. Gilbert, David C. Montefiori, Adrian McDermott, Youyi Fong, David Benkeser, Weiping Deng, Honghong Zhou, Christopher R. Houchens, Karen Martins, Lakshmi Jayashankar, Flora Castellino, Britta Flach, Bob C. Lin, Sarah O’Connell, Charlene McDanal, Amanda Eaton, Marcella Sarzotti-Kelsoe, Yiwen Lu, Chenchen Yu, Bhavesh Borate, Lars W. P. van der Laan, Nima Hejazi, Chuong Huynh, Jacqueline Miller, Hana M. El Sahly, Lindsey R. Baden, Mira Baron, Luis De La Cruz, Cynthia Gay, Spyros Kalams, Colleen F. Kelley, Mark Kutner, Michele P. Andrasik, James G. Kublin, Lawrence Corey, Kathleen M. Neuzil, Lindsay N. Carpp, Rolando Pajon, Dean Follmann, Ruben O. Donis, Richard A. Koup, on behalf of the Immune Assays; Moderna, Inc.; Coronavirus Vaccine Prevention Network (CoVPN)/Coronavirus Efficacy (COVE); and United States Government (USG)/CoVPN Biostatistics Teams

**Affiliations:** The Vaccine and Infectious Disease Division, Fred Hutchinson Cancer Research Center, Seattle, WA; The Vaccine Research Center; The Biostatistics Research Branch; National Institute of Allergy and Infectious Diseases, National Institutes of Health, Bethesda, MD; the Department of Surgery and Duke Human Vaccine Institute, Duke University Medical Center, Durham, NC; The Department of Biostatistics and Bioinformatics, Rollins School of Public Health, Emory University, Atlanta, GA; The Biomedical Advanced Research and Development Authority, Washington, DC; Graduate Group in Biostatistics, University of Berkeley, Berkeley, CA; Moderna, Inc., Cambridge, MA; Baylor College of Medicine, Houston, TX; Brigham and Women’s Hospital, Boston, MA; Palm Beach Research Center, West Palm Beach, FL; Keystone Vitalink Research, Greenville, SC; University of North Carolina, Chapel Hill, NC; Vanderbilt University Medical Center, Nashville, TN; Division of Infectious Diseases, Department of Medicine, Emory University School of Medicine and the Grady Health System, Atlanta, GA; Suncoast Research Group, Miami, FL; The Center for Vaccine Development and Global Health, University of Maryland School of Medicine, Baltimore, MD

## Abstract

**Background:** In the Coronavirus Efficacy (COVE) trial, estimated mRNA-1273 vaccine efficacy against coronavirus disease-19 (COVID-19) was 94%. SARS-CoV-2 antibody measurements were assessed as correlates of COVID-19 risk and as correlates of protection.

**Methods:** Through case-cohort sampling, participants were selected for measurement of four serum antibody markers at Day 1 (first dose), Day 29 (second dose), and Day 57: IgG binding antibodies (bAbs) to Spike, bAbs to Spike receptor-binding domain (RBD), and 50% and 80% inhibitory dilution pseudovirus neutralizing antibody titers calibrated to the WHO International Standard (cID50 and cID80). Participants with no evidence of previous SARS-CoV-2 infection were included. Cox regression assessed in vaccine recipients the association of each Day 29 or 57 serologic marker with COVID-19 through 126 or 100 days of follow-up, respectively, adjusting for risk factors.

**Results:** Day 57 Spike IgG, RBD IgG, cID50, and cID80 neutralization levels were each inversely correlated with risk of COVID-19: hazard ratios 0.66 (95% CI 0.50, 0.88; p=0.005); 0.57 (0.40, 0.82; p=0.002); 0.42 (0.27, 0.65; p<0.001); 0.35 (0.20, 0.61; p<0.001) per 10-fold increase in marker level, respectively, multiplicity adjusted P-values 0.003-0.010. Results were similar for Day 29 markers (multiplicity adjusted P-values <0.001-0.003). For vaccine recipients with Day 57 reciprocal cID50 neutralization titers that were undetectable (<2.42), 100, or 1000, respectively, cumulative incidence of COVID-19 through 100 days post Day 57 was 0.030 (0.010, 0.093), 0.0056 (0.0039, 0.0080), and 0.0023 (0.0013, 0.0036). For vaccine recipients at these titer levels, respectively, vaccine efficacy was 50.8% (−51.2, 83.0%), 90.7% (86.7, 93.6%), and 96.1% (94.0, 97.8%). Causal mediation analysis estimated that the proportion of vaccine efficacy mediated through Day 29 cID50 titer was 68.5% (58.5, 78.4%).

**Conclusions:** Binding and neutralizing antibodies correlated with COVID-19 risk and vaccine efficacy and likely have utility in predicting mRNA-1273 vaccine efficacy against COVID-19.

**Trial registration number:** COVE ClinicalTrials.gov number, NCT04470427

## Introduction

Multiple vaccines have demonstrated efficacy against coronavirus disease-19 (COVID-19) in phase 3 trials^1^ and have been authorized for emergency use.^2,3^ However, the manufacturing challenges posed by the global demand for doses, the need for affordable and widely accessible options that are safe and effective in diverse populations, the lack of efficacy data in important populations (e.g., pediatrics, pregnancy and immunocompromised patients), and the emergence of more transmissible viral variants, highlight the need for a large armamentarium of effective COVID-19 vaccines.^4,5^ The identification and validation of a correlate of protection^6-8^ could expedite the clinical evaluation and regulatory approval process for existing vaccines for new populations, for vaccine regimen modifications, and for new vaccines.

The Coronavirus Efficacy (COVE) phase 3 trial of the mRNA-1273 COVID-19 vaccine showed estimated vaccine efficacy against COVID-19 of 94.1%,^9^ leading to the US Food and Drug Administration’s Emergency Use Authorization of mRNA-1273 for prevention of COVID-19 in adults,^10^ and more than 130 million doses have been administered in the United States.^11^ The mRNA-1273 vaccine has been shown to be highly effective in the elderly and in essential and frontline workers, including healthcare workers,^12^ and to have non-inferior immunogenicity in adolescents vs. adults.^13^

Neutralizing antibodies (nAbs) or binding antibodies (bAbs) have been established as a correlate of protection/surrogate marker for vaccines against many viral diseases.^7^ The Spike protein^14^ and its receptor binding domain (RBD)^15^ are targets for nAbs produced by SARS-CoV-2 infection and by vaccination.^16-18^ The hypothesis that antibodies, whether elicited by natural infection or by Spike protein-based vaccines, are a correlate of protection against COVID-19 is supported by diverse lines of evidence,^19-30^ including evidence supporting a mechanistic correlate of protection that can be derived from passive transfer of antibodies in experimental challenge studies and from studies of human monoclonal antibodies.^21,28-30^ For the mRNA-1273 vaccine, multiple SARS-CoV-2 antibody markers (IgG bAbs to Spike, IgG bAbs to Spike RBD, 50% (ID50) inhibitory dilution nAb titer, and angiotensin-converting enzyme 2 (ACE2)-binding inhibition) each correlated with protection, defined as reduced SARS-CoV-2 replication after challenge, in vaccinated rhesus macaques.^30^ Here we assessed three of these same SARS-CoV-2 antibody markers (IgG bAbs to Spike, IgG bAbs to Spike RBD, ID50 nAb titer), as well as 80% inhibitory dilution (ID80) nAb titer, as correlates of risk of COVID-19 and as correlates of mRNA-1273 vaccine protection against COVID-19 in the COVE trial.

## Methods

### Trial design and COVID-19 endpoint evaluated for correlates

From July 27, 2020 to October 23, 2020, 30,415 participants were randomized (1:1 ratio) to receive two injections of mRNA-1273 (100 μg) or placebo at Day 1 and at Day 29.^9^ Correlates were evaluated against the protocol-specified primary endpoint, referred to as “COVID-19”: first occurrence of acute symptomatic COVID-19 with virologically-confirmed SARS-CoV-2 infection in participants with no evidence of previous SARS-CoV-2 infection.^9^ Correlates analyses included COVID-19 endpoints diagnosed during two time periods: “intercurrent endpoints” starting 7 days post Day 29 visit through 6 days post Day 57 visit and “post Day 57 endpoints” starting 7 days post Day 57 visit and any time during follow-up while participants were still blinded to randomization assignment (Figure S1). The data cutoff date was March 26, 2021.

### Laboratory methods

A solid-phase electrochemiluminescence S-binding IgG immunoassay was developed for simultaneous quantitative detection of IgG antibodies to three distinct SARS-CoV-2 antigens in human serum. This validated assay uses custom plates manufactured by Meso Scale Discovery (Rockville, Maryland) provided as 4-plex, detecting SARS-CoV-2 prefusion stabilized Spike protein (S-2P), RBD, and Nucleocapsid, with a bovine serum albumin control ligand. The Spike and RBD markers were converted to International Units for assessment as immune correlates. Neutralizing antibodies were measured in a formally validated assay as a function of reductions in luciferase reporter gene expression after a single round of infection with SARS-CoV-2.D614G Spike-pseudotyped virus in 293T/ACE2 cells as described.^31^ ID50 and ID80 nAb titers were calculated based on a dose-response curve and were calibrated to the WHO anti-SARS-CoV-2 immunoglobulin International Standard^32,33^ and the resulting calibrated ID50 titer (cID50) and calibrated ID80 titer (cID80) assessed as immune correlates. Supplementary Text 1 and Table S1 provide additional details on both immunoassays.

### Statistical methods

All data analyses were pre-specified in the Statistical Analysis Plan (Supplementary Material), summarized here. A case-cohort sampling design was used for selecting participants for measurement of antibody markers at Day 1, 29, 57,^34^ comprising a random sample of all participants, stratified by cross-classified randomization arm, baseline SARS-CoV-2 positivity, protocol randomization strata (Age below 65 years At-Risk, Age below 65 years Not At-Risk, Age above 65 years), and underrepresented minority status, and all other participants who acquired an intercurrent or post Day 57 COVID-19 endpoint until database lock. At-Risk comprised six self-reported health/comorbidities.^9^ Analyses were based on the case-cohort set defined by the subset of selected participants with no evidence of previous SARS-CoV-2 infection and per-protocol received both doses without major protocol violations (Figure S2). Each participant had two inverse probability sampling (IPS) weights calculated, for Day 29 and Day 57 marker correlates analyses. Day 29 and Day 57 marker correlates analyses excluded participants with any evidence of SARS-CoV-2 infection before 7 days post Day 29 and Day 57 visit, respectively, because natural infection before the serology may have modulated the antibody response.

For each marker, values below the assay limit of detection (LOD) were set to LOD/2, and values above the upper limit of quantitation (ULOQ) were set to ULOQ. Positive responses for Spike IgG and RBD IgG were defined by IgG > 10.8424 IU/ml and 14.0858 IU/ml, respectively. Positive responses for cID50 and cID80 were defined by value > LOD (LOD = 2.42 and 15.02, respectively). All analyses were done reproducibly based on R scripts hosted at the Github code repository CoVPN/correlates_reporting, which are publicly available with application to a mock COVE trial data set.

The positive response rate for each marker and analyzed group was estimated by an IPS-weighted average, with 95% confidence interval (CI) calculated with an IPS-weighted Clopper-Pearson method. Differences in response rates with 95% CIs were calculated by the Wilson-Score method without continuity correction.^35^ Geometric mean concentration (GMC) or titer (GMT) and their ratios between cases vs. non-cases were estimated with 95% CIs, with hypothesis tests, based on the t-distribution with IPS weighting. Reverse cumulative distribution function curves were estimated by IPS-weighted nonparametric maximum likelihood.

### Covariate-adjustment

All correlates analyses adjusted for baseline variables prognostic for COVID-19: At-Risk status, membership in a community of color, and the logit of predicted COVID-19 risk score built from machine learning of placebo arm participants (Supplementary Text 2, Tables S2-S5, Figures S3-S4).

### Correlates of risk in vaccine recipients

Each antibody marker at Day 57 was assessed as a correlate of risk for COVID-19 starting 7 days post Day 57 visit; similarly each Day 29 marker was assessed as a correlate of risk for COVID-19 starting 7 days post Day 29 visit. For each marker, without replacement IPS-weighted Cox regression was applied to estimate the covariate-adjusted hazard ratio of COVID-19 across tertiles of the marker or per 10-fold increase in the quantitative marker level with 95% CIs and Wald-based p-values. The Cox model fits were used to estimate marker-conditional cumulative incidence of COVID-19 marginalizing over baseline variables, with 95% CIs computed using the bootstrap. The Cox models were fit using the *survey* R package.^36^ Additional analyses estimated the same quantity with a generalized additive model with degrees of freedom selected by generalized cross-validation,^37^ and by nonparametric threshold regression.^38^

### Controlled vaccine efficacy and mediation

Vaccine efficacy by marker level was estimated by a causal inference approach implemented using the IPS-weighted Cox regression described above for the vaccine arm and unweighted Cox regression for the placebo arm, marginalizing over the baseline variables for each randomization arm.^39^ A sensitivity analysis was conducted to assess robustness of findings to unmeasured confounding of the effect of the antibody marker on COVID-19 (Supplementary Text 3).^39^ For each antibody marker with at least 10% of vaccinated participants with value the same as for placebo recipients (i.e., negative or undetectable), the proportion of vaccine efficacy mediated by the antibody marker was estimated.^40^

### Hypothesis testing

All p-values were two-sided. For each set of hypothesis tests for Day 29 and Day 57 marker correlates of risk − which included eight Cox model Wald-tests for correlates of risk including both tertiles of markers and quantitative markers − Westfall-Young multiplicity adjustment^41^ was applied to obtain q-values and family-wise error rate p-values.

### Software and data quality assurance

The analysis was implemented in R version 4.0.3; the code used for analysis was verified using mock data.

## Results

### Participant demographics

Tables 1, S6, and S7 summarize the case-cohort set data and demographics (N = 1010 vaccine, N = 137 placebo in the immunogenicity subcohort). Thirty-four percent of case-cohort participants were age 65 or over, 40% were At-Risk, 47% were female, 32% were Hispanic or Latino, 46% White and Non-Hispanic, and 54% communities of color, with 18% Black or African American.

**Table 1:** Anti-Spike and anti-RBD IgG response rates and Geometric Mean Concentrations (GMCs) and pseudovirus calibrated neutralization titer cID50 and cID80 response rates and Geometric Mean Titers (GMTs) by COVID-19 outcome status. Analysis based on baseline negative per-protocol vaccine recipients in the case-cohort set.

| Visit for Marker | Marker | COVID-19 Cases* |  |  | Non-Cases |  |  | Comparison |  |
| --- | --- | --- | --- | --- | --- | --- | --- | --- | --- |
|  |  | N | Response Rate | GMC or GMT | N | Response Rate | GMC or GMT | Response Rate Difference | Ratio of GM (Cases/Non-Cases) |
| Day 29 | Anti Spike IgG (IU/ml) | 46 | 97.8%<br>(85.4%, 99.7%) | 183<br>(126, 266) | 1005 | 98.6%<br>(97.4%, 99.2%) | 318<br>(292, 347) | -0.01<br>(-0.13, 0.01) | 0.57<br>(0.39, 0.84) |
| Day 29 | Anti RBD IgG (IU/ml) | 46 | 97.8%<br>(85.4%, 99.7%) | 207<br>(147, 293) | 1005 | 98.4%<br>(97.2%, 99.1%) | 327<br>(302, 354) | -0.01<br>(-0.13, 0.02) | 0.63<br>(0.44, 0.90) |
| Day 29 | Pseudovirus-nAb cID50 | 46 | 65.2%<br>(50.1%, 77.8%) | 7.6<br>(5.4, 10.8) | 1005 | 81.7%<br>(78.8%, 84.3%) | 13.0<br>(11.9, 14.1) | -0.17<br>(-0.32, -0.04) | 0.59<br>(0.41, 0.84) |
| Day 29 | Pseudovirus-nAb cID80 | 46 | 43.5%<br>(29.7%, 58.4%) | 18.0<br>(13.3, 24.2) | 1005 | 63.9%<br>(60.4%, 67.3%) | 29.0<br>(27.1, 31.0) | -0.2<br>(-0.35, -0.05) | 0.62<br>(0.46, 0.84) |
| Day 57 | Anti Spike IgG (IU/ml) | 36 | 100.0%<br>(100.0%, 100.0%) | 1890<br>(1449, 2465) | 1005 | 99.4%<br>(98.2%, 99.8%) | 2652<br>(2457, 2863) | 0.01<br>(0, 0.02) | 0.71<br>(0.54, 0.94) |
| Day 57 | Anti RBD IgG (IU/ml) | 36 | 100.0%<br>(100.0%, 100.0%) | 2744<br>(2056, 3664) | 1005 | 99.4%<br>(98.3%, 99.8%) | 3937<br>(3668, 4227) | 0.01<br>(0, 0.02) | 0.70<br>(0.52, 0.94) |
| Day 57 | Pseudovirus-nAb cID50 | 36 | 100.0%<br>(100.0%, 100.0%) | 160<br>(117, 220) | 1005 | 98.7%<br>(97.6%, 99.3%) | 247<br>(231, 264) | 0.01<br>(0.01, 0.02) | 0.65<br>(0.47, 0.90) |
| Day 57 | Pseudovirus-nAb cID80 | 36 | 97.2%<br>(81.6%, 99.6%) | 332<br>(248, 444) | 1005 | 98.3%<br>(97.1%, 99.1%) | 478<br>(450, 508) | -0.01<br>(-0.17, 0.02) | 0.69<br>(0.52, 0.93) |

\*Cases for Day 29 marker correlates analyses are baseline negative per-protocol vaccine recipients with the symptomatic infection COVID-19 primary endpoint diagnosed starting 7 days after the Day 29 study visit. Cases for Day 57 marker correlates analyses are baseline negative per-protocol vaccine recipients with the symptomatic infection COVID-19 primary endpoint diagnosed starting 7 days after the Day 57 study visit.
N is the number of participants with Day 1, 29, 57 antibody marker data included in data analyses (for Non-Cases and for COVID-19 Cases for Day 57 marker correlates analyses). For COVID-19 Cases for Day 29 marker correlates analyses, N is the number with Day 1, 29 antibody marker data included in analyses (Day 57 marker data are not used in Day 29 marker correlates analyses).
GM, geometric mean; GMC, geometric mean concentration; GMT, geometric mean titer.
cID50 and cID80: ID50 and ID80 neutralizing antibody (nAb) titers calibrated to the WHO International Standard.

### COVID-19 endpoints

For Day 57 marker correlates analyses, 47 vaccine recipients had a COVID-19 endpoint. For Day 29 marker correlates analyses, an additional 8 intercurrent COVID-19 endpoints occurred. Overall cumulative incidence from 7 to 126 days post Day 29 was 0.005 in vaccine recipients compared to 0.065 in placebo recipients. Average follow-up of vaccine recipients was 81 days (starting 7 days post Day 57 visit) for Day 57 marker correlates analyses and 109 days (starting 7 days post Day 29 visit) for Day 29 marker correlates analyses.

### Antibody marker distributions

At Day 57, almost 100% of vaccine recipients had positive/detectable antibody response by all four markers (Table 1). This was also true at Day 29 for Spike IgG and RBD IgG, whereas cID50 and cID80 titers were detectable in 82% and 64% of vaccine recipients, respectively. Each marker was moderately correlated between the Day 29 and Day 57 time points (Spearman rank r = 0.527 to 0.625, Figure S5). Median (interquartile range) days from dose one to Day 29 was 28 (28 to 30) and from Day 29 to Day 57 was 28 (28 to 30). The Spike IgG and RBD IgG markers were tightly correlated (Spearman rank r = 0.942, 0.969 at Day 29, 57; Figures S6, S7), as were the cID50 and cID80 markers (r = 0.968, 0.961 at Day 29, 57; Figures S6, S7). Accordingly, some results only show Spike IgG and cID50 for brevity; results were similar for RBD IgG and cID80 (Supplementary Appendix). Each binding antibody marker was correlated with each neutralization marker at each time point (r = 0.734 to 0.804).

Figures 1 and S8 show the Day 29 and Day 57 marker distributions by case/non-case status in vaccine recipients (Figure S9 shows analogous plots for placebo recipients), with Figures S10 and S11 showing marker values by participant age. For all eight markers the geometric mean was lower in cases, with 95% CIs about geometric mean ratios (cases/non-cases) below one (Table 1). Figures S12 and S13 show reverse cumulative distribution function curves of the eight markers, in the context of the overall vaccine efficacy estimates.^42^ Figure S14 shows the marker values of vaccine breakthrough cases by timing of COVID-19 endpoint diagnosis.

**Figure 1:**
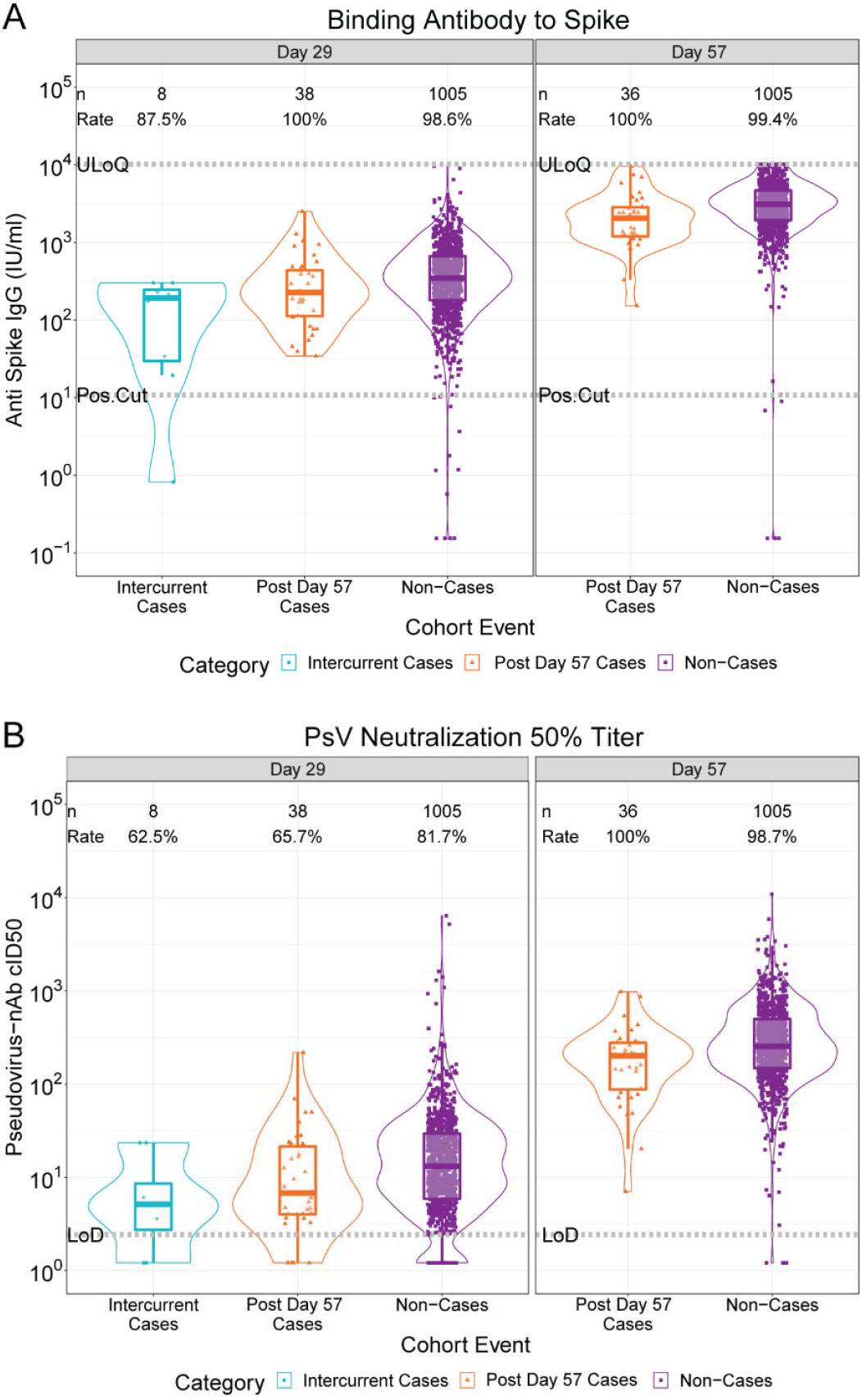
(A) Anti-Spike IgG concentration and (B) pseudovirus neutralization cID50 titer by COVID-19 outcome status. Data points are from baseline negative per-protocol vaccine recipients selected into the case-cohort set. The violin plots contain interior box plots with upper and lower horizontal edges the 25^th^ and 75^th^ percentiles of antibody level and middle line the 50^th^ percentile. Each side shows a rotated probability density (estimated by a kernel density estimator with a default Gaussian kernel) of the data. Positive response rates were computed with inverse probability of sampling weighting. Pos.Cut, Positivity cut-off. LoD, limit of detection. ULoQ, upper limit of quantitation; ULOQ = 10,919 for cID50 (above all data points). Post Day 57 cases are COVID-19 endpoints starting 7 days post Day 57 visit; Intercurrent cases are COVID-19 endpoints starting 7 days post Day 29 visit through 6 days post Day 57 visit. cID50: ID50 nAb titer calibrated to the WHO International Standard.

### Correlates of risk

Figure 2 shows COVID-19 cumulative incidence curves for subgroups of vaccine recipients defined by tertile of Day 57 IgG Spike and cID50 (A, B) (results for IgG RBD and cID80 in Figure S15). A dose response was observed for each marker, with estimated hazard ratios for upper vs. lower tertiles between 0.20 and 0.31 and 95% CIs and multiplicity-adjusted p-values indicating significant inverse correlations with risk (Figure 2C). Results were similar for Day 29 markers, with smaller hazard ratios and smaller p-values indicating strengthened evidence for correlates of risk (Figures S16, S17).

**Figure 2.**
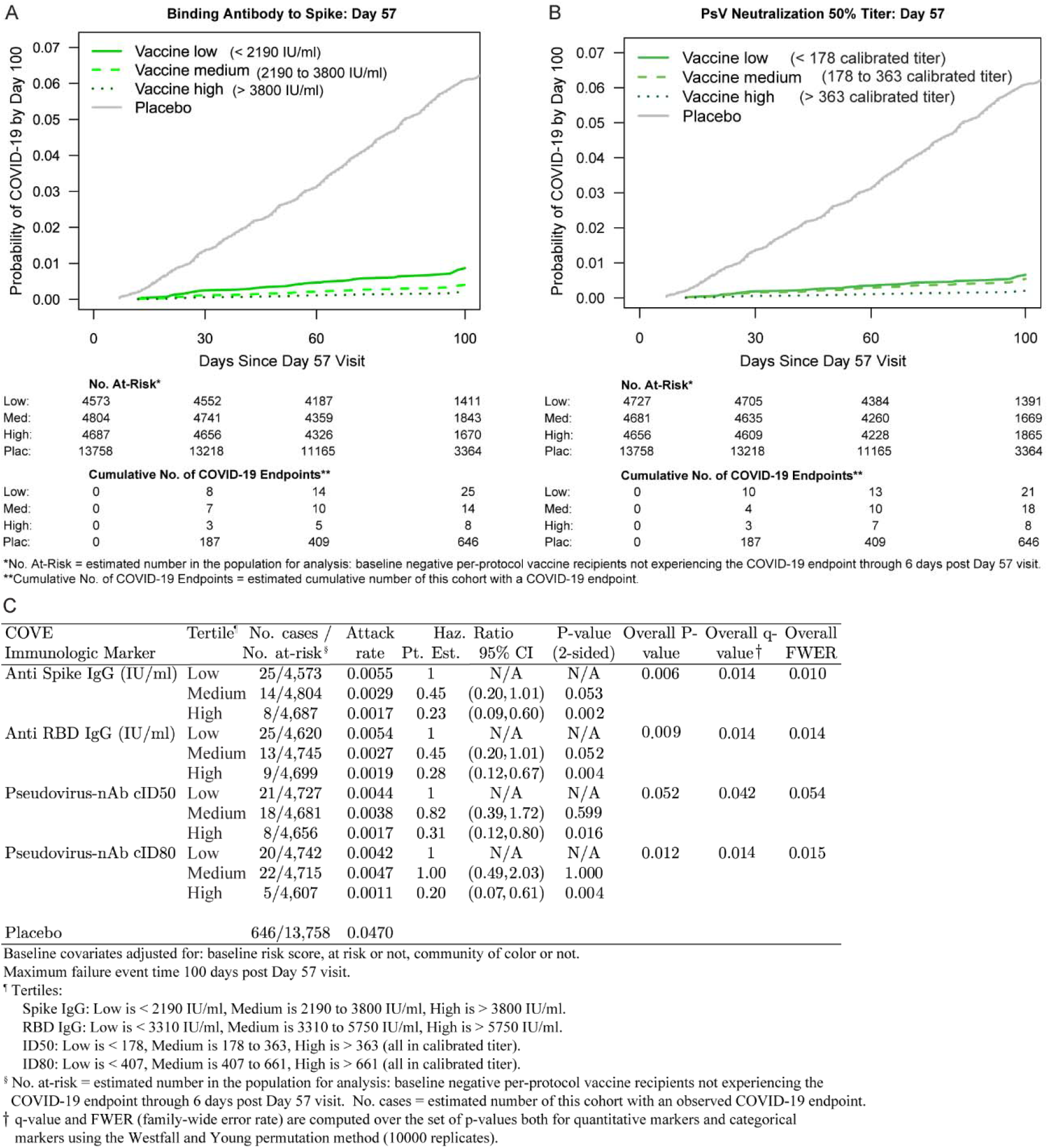
Covariate-adjusted cumulative incidence of COVID-19 by Low, Medium, High tertile of Day 57 IgG concentration or pseudovirus neutralization titer. (A) Anti-Spike IgG concentration; (B) cID50 titer; (C) IgG (Spike, RBD) and (cID50, cID80). The overall p-value is from a generalized Wald test of whether the hazard rate of COVID-19 differed across the Low, Medium, and High subgroups. cID50, cID80: ID50 or ID80 nAb titer calibrated to the WHO International Standard.

For quantitative Day 57 markers, the estimated hazard ratio per 10-fold increase in marker value ranged between 0.35 and 0.66 (Figure 3A), with 95% CIs below one and multiplicity-adjusted p-values indicating significant associations. Generally, similar results were obtained across pre-specified subgroups of vaccine recipients (Figure 3B and 3C, Figure S18).

**Figure 3.**
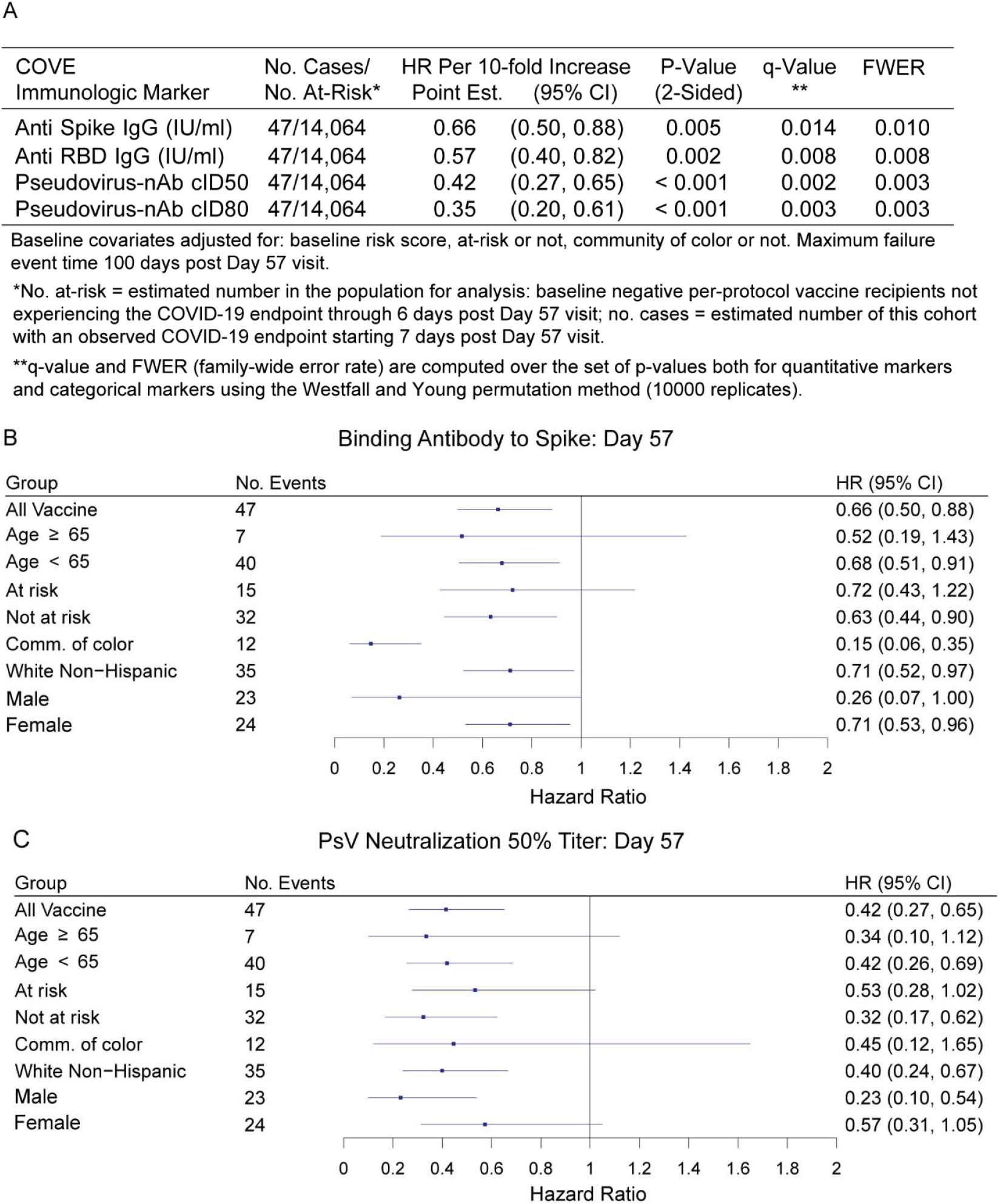
Covariate-adjusted hazard ratios of COVID-19 per 10-fold increase in each Day 57 antibody marker in baseline negative per-protocol vaccine recipients overall and in subgroups. (A) Inferences for IgG (Spike, RBD) and (cID50, cID80); (B) Forest plots for Spike IgG; (C) Forest plots for cID50. cID50, cID80: ID50 or ID80 neutralizing antibody titer calibrated to the WHO International Standard. Comm. of color = all participants other than White Non-Hispanic.

The estimated cumulative incidence of COVID-19 by end of follow-up (100 days post Day 57) for the entire vaccine group was 0.0033 (95% CI 0.0022, 0.0045). Based on nonparametric threshold regression this rate decreased across vaccinated subgroups with Day 57 cID50 titer above a given threshold, with estimates of 0.00088 (0.00, 0.0020) at cID50 titer above 500 and zero COVID-19 endpoints at cID50 titer above 1000 (Figure 4A, Figure 1B). In contrast, 5% of non-cases had cID50 titer above 1000 (Figure 1B). Based on the Cox model, the COVID-19 rate for vaccine recipients with undetectable Day 57 cID50 titer (value < LOD) was 0.030 (0.010, 0.093), and decreased to 0.0056 (0.0039, 0.0080) at titer of 100 and to 0.0023 (0.0013, 0.0036) at titer of 1000 (Figure 4B). Results using an alternative model (generalized additive) also supported inverse correlates of risk for all markers (Figures S19, S20).

**Figure 4.**
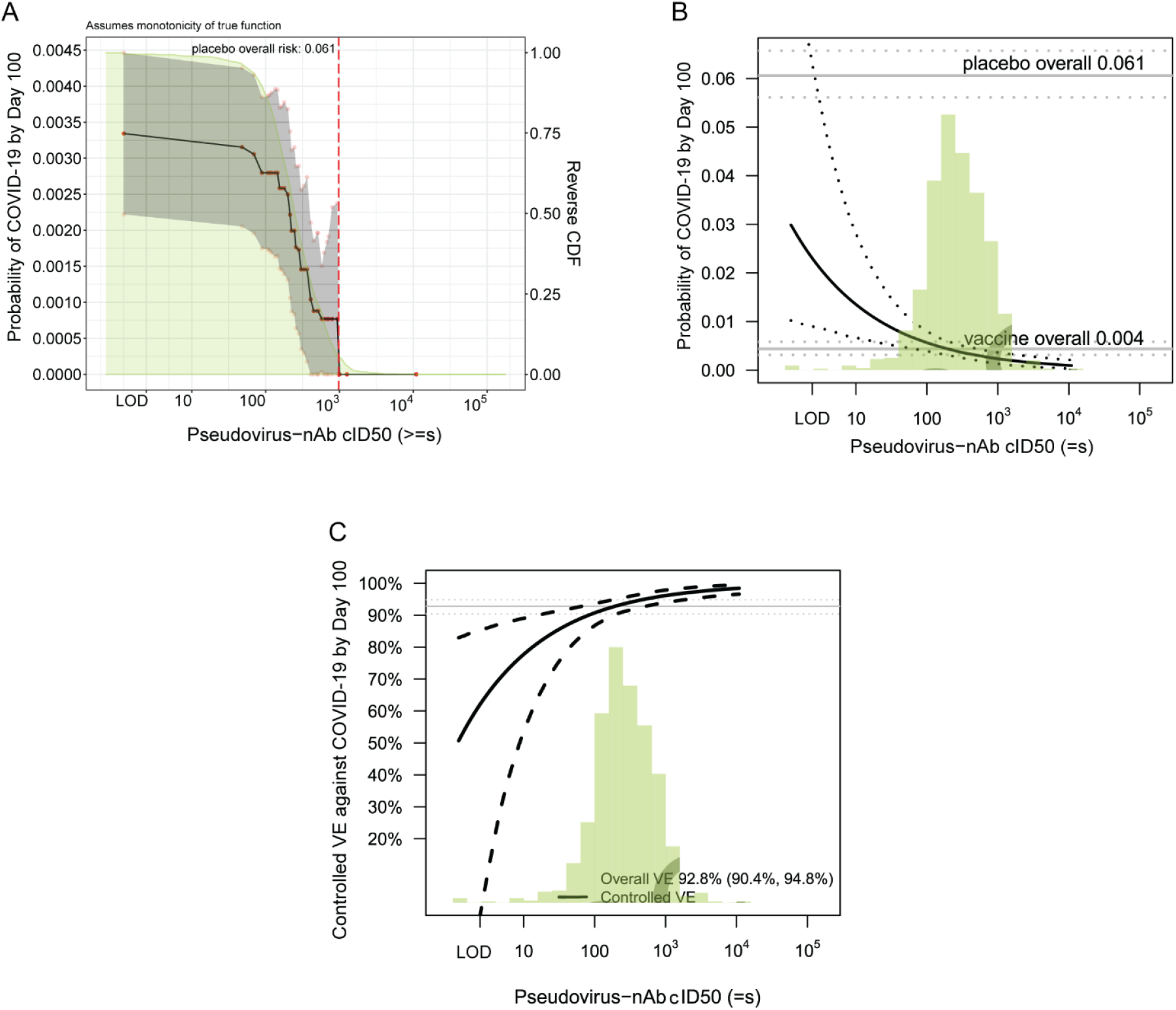
(A) Covariate-adjusted cumulative incidence of COVID-19 by 100 days post Day 57 by subgroups defined by Day 57 cID50 level above a threshold, with reverse cumulative distribution function of Day 57 cID50 level overlaid in green. The gray shaded area is pointwise 95% confidence intervals (CIs). The upper boundary of the green shaded area is the estimate of the reverse cumulative distribution function (CDF) of the marker in baseline SARS-CoV-2 negative per-protocol vaccine recipients. (B) Covariate-adjusted cumulative incidence of COVID-19 by 100 days post Day 57 by Day 57 cID50 level. The dotted lines indicate bootstrap point-wise 95% CIs. (C) Vaccine efficacy by Day 57 cID50 level, estimated using the method of Gilbert, Fong, and Carone.^39^ The dashed lines indicate bootstrap point-wise 95% CIs. In (B) and (C), covariate adjustment was based on an inverse probability sampling-weighted Cox model; the green histograms are an estimate of the density of Day 57 cID50 level in baseline negative per-protocol vaccine recipients. LOD, limit of detection. cID50: ID50 nAb titer calibrated to the WHO International Standard.

### Correlates of protection

Vaccine efficacy estimates (obtained using the method of Gilbert, Fong, and Carone^39^) increased with Day 57 cID50 neutralization titer: At undetectable (<LOD) Day 57 cID50 titer, vaccine efficacy was 50.8% (−51.2, 83.0%). At Day 57 cID50 titer of 100, vaccine efficacy increased to 90.7% (86.7, 93.6%), and at Day 57 cID50 titer of 1000, vaccine efficacy further increased to 96.1% (94.0, 97.8%) (Figure 4C). Figures S21-S27 show these results for the other seven antibody markers. The sensitivity analysis allowing unmeasured confounding (Figure S28C) yielded estimates of vaccine efficacy of 89.8% (68.8, 96.5%) at undetectable Day 57 cID50 titer, 94.9% (92.6, 96.8%) at Day 57 cID50 titer of 500, and 95.8% (93.4, 97.6%) at Day 57 cID50 titer of 1000 (see Figures S28 and S29 for results for the other seven markers). The sensitivity analysis based on E-values^43^ of the vaccine recipient tertile subgroups supported the finding that vaccine efficacy was generally higher for the upper tertile subgroup, indicating some robustness to potential unmeasured confounding (Table S8).

Day 29 cID50 and cID80 titers could be assessed as mediators of VE by the Benkeser et al. method,^40^ given 18% and 36% of vaccine recipients had undetectable titer, respectively. An estimated 68.5% (58.5, 78.4%) of VE was mediated by Day 29 cID50 titer and 48.5% (34.5, 62.4%) by Day 29 cID80 titer (Table S9).

## Discussion

For recipients of two doses of mRNA-1273 in the COVE trial, all four antibody markers at Day 29 and at Day 57 were inverse correlates of risk of COVID-19 occurrence through 3-4 months post second dose. Based on any of the antibody markers, COVID-19 risk was about 10 times lower for vaccine recipients with antibodies in the top 10% of values compared to those with negative/undetectable values (the same value as in baseline negative placebo recipients).

The correlates of protection analyses found that estimated vaccine efficacy against COVID-19 increased with Day 29 and with Day 57 antibody levels, where for the latter estimated vaccine efficacy was about 45-60% for vaccine recipients with negative/undetectable binding or neutralizing antibody response (Figure 4C; Figures S21C, S22C, S23C) and increased to above 98% at highest antibody levels. Positive vaccine efficacy for the vaccinated subgroup with negative/undetectable antibody level implies lack of full mediation of vaccine efficacy through the antibody marker,^39^ and formal causal analysis estimated that 68% of the overall vaccine efficacy was mediated through Day 29 cID50 titer. To interpret the mediation result: if neutralizing antibodies circulating on Day 29 could be removed but the other consequences of vaccination remained, overall vaccine efficacy would be expected to change from 92.3% to 56%, a 68% reduction (on the log scale) of overall vaccine efficacy. In comparison, hemagglutination inhibition titer against B/Victoria influenza virus mediated an estimated 57% of inactivated influenza vaccine efficacy against virologically confirmed B/Victoria influenza illness.^44^ As hemagglutination inhibition titer has been used to guide vaccine strain selection and approval, this defines a benchmark for a potential level of mediation relevant for supporting vaccine approval decisions.^45^ The very high overall vaccine efficacy and the small number of vaccine recipients with negative/undetectable antibody levels in COVE limited the ability to identify a high percentage of neutralization-mediated vaccine efficacy. Similar analyses of trials with lower vaccine efficacy and lower neutralization response rates may be able to detect a greater percentage of vaccine efficacy mediated through neutralizing antibody titers.

An interpretation also consistent with our results is that neutralization as a biological function did fully mediate the vaccine efficacy in COVE, but the cID50 and cID80 markers had inadequate sensitivity to quantify low-level neutralization below the detection limit that was partially protective. This hypothesis is supported by the finding that passive transfer of purified IgG from mRNA-1273-immunized rhesus macaques was sufficient to protect golden Syrian hamsters from disease after SARS-CoV-2 challenge.^30^ Yet, it is more likely that additional immune responses are needed to fully explain the observed vaccine efficacy, given that the vaccine provided more than 90% efficacy by Day 29,^9^ 18% of vaccine recipients had undetectable neutralization response at Day 29 based on cID50 (Figure 1, Figure S8), and the vaccine elicited other immune responses by Day 29 in a high proportion of vaccine recipients.^46^ In addition to markers measuring distinct immunological functions, markers not measured fully in serum (e.g., mucosal markers) and/or anamnestic responses not fully represented by a single time point measurement may be needed to fully explain vaccine efficacy.

The immune correlates results of the AZD12222 trial^25^ for Spike IgG and RBD IgG can be quantitatively compared to the COVE results by virtue of the same MSD assay platform, conversion of IgG concentration to WHO International Units/ml, and the same antibody measurement time: 4 weeks post second dose. Estimated ChAdOx1 nCoV-19 vaccine efficacy was 70% and 90% at Spike IgG levels of 113 (95% CI < LOD = 0.31, 245) and 899 (369, NC) IU/ml, respectively, and at RBD IgG levels of 165 (< LOD = 1.59, 452) and 2360 (723, NC) IU/ml, respectively (NC = not calculated).^25^ In comparison, estimated mRNA-1273 vaccine efficacy was 70% and 90% at Day 57 Spike IgG levels of 1 (< LOD = 0.31, 170) and 298 (1, 1786) IU/ml, respectively, and at Day 57 RBD IgG levels of 8 (< LOD = 1.59, 243) and 775 (29, 2819) IU/ml, respectively. These results are suggestive that lower binding antibody levels in COVE are associated with the same level of vaccine efficacy.

Pseudovirus neutralization results can also be compared between the trials using calibrated ID50 titers, where estimated ChAdOx1 nCoV-19 vaccine efficacy was 70% and 90% at cID50 titer of 8 (< LOD = 2.42, 26) and 140 (43, NC), compared to COVE results at cID50 titer of 4 (< LOD = 2.42, 22) and 83 (16, 188). The neutralization assays employed for AZD12222 and COVE used the D614 and D614G pseudovirus, respectively, such that this comparison is affected by any difference in neutralization sensitivity of these two strains. As a sensitivity analysis, we multiplied the AZD12222 cID50 values by a second conversion factor, 2.19, which scales the values as if D614G had been used in the assay (2.19 was calculated from the validation study that used both strains, Supplementary Text 1). The cID50 titers at 70% and 90% ChAdOx1 nCoV-19 vaccine efficacy are 18 (< LOD = 2.42, 57) and 307 (95, NC), respectively.

With the caveats of different study endpoints/hosts, the COVE results are also consistent with results on Spike IgG and neutralizing antibody titers as correlates of protection against SARS-CoV-2 replication in mRNA-1273-vaccinated rhesus macaques. For instance, all macaques with Spike IgG > 336 IU/ml at 4 weeks post second dose were protected from >10,000 subgenomic RNA copies/ml in bronchoalveolar lavages,^30^ and in COVE, Day 57 Spike IgG of 336 IU/ml corresponded to 90% vaccine efficacy against COVID-19 (Figure S22).

Together with evidence from other studies, the current results support that neutralization titer is a potential surrogate marker for mRNA-1273 vaccination against COVID-19 that can be considered as a primary endpoint for basing certain accelerated approval decisions. The total body of evidence may support an immunogenicity non-inferiority approach to bridging vaccine efficacy of refined or novel mRNA vaccines, which has been proposed for adding vaccine spike variants and boosters.^47^ One open question challenging this approach is whether titers to different strains have different relationships with vaccine efficacy. A more challenging objective is bridging vaccine efficacy of new candidate vaccines in different platforms from mRNA. When correlates results are available from several phase 3 vaccine efficacy trials of different vaccine platforms, it will be possible to conduct validation analyses of how well an antibody marker can be used to predict vaccine efficacy across platforms.

We also note that the Day 29 markers correlated with COVID-19 at least as strongly as the Day 57 markers. If a Day 29 immune marker in recipients of two mRNA-1273 doses becomes established as a correlate of protection, it could be a more practical surrogate marker than a Day 57 marker by virtue of being measured earlier and because trials would not need participants to come back for a Day 57 study visit.

Limitations of this correlates study include the lack of data for assessing correlates against other outcomes besides COVID-19 (e.g., severe COVID-19, asymptomatic SARS-CoV-2 infection, infection regardless of symptomology, viral shedding), the lack of assessment of correlates for recipients of only one mRNA-1273 dose, and the fact that almost all COVID-19 cases resulted from infections with viruses bearing genetically and antigenically similar Spike as compared to the ancestral Spike in the vaccine, implying that it was not possible to assess robustness of correlates to SARS-CoV-2 variants of concern. However, the paucity of divergent circulating virus is also a strength in affording a clear interpretation as correlates against COVID-19 caused by variants genetically close to the vaccine. Another strength is the use of validated assays with all results reported in WHO International Units or calibrated to WHO International Standards, facilitating comparison of results from other studies and vaccine platforms. An additional strength is the racial and ethnic diversity of the trial participants and large number of diverse participants sampled for immunogenicity measurements.^48^

This study evaluated antibody markers measured at two specific time points (Day 29, Day 57) as immune correlates against subsequent COVID-19 occurrence over 3-4 months follow-up, but did not assess how the current level of antibody correlated with instantaneous risk of COVID-19. COVE follows participants for 2 years, enabling future analyses of when and how vaccine efficacy wanes as antibody levels wane and as new variants of SARS-CoV-2 emerge, informing decisions about the timing of a booster or the need to update vaccine composition, and potentially clarifying the mediation of protection. Future correlates analyses that study antibody markers over time in relation to the timing of breakthrough infections of variants with variable neutralization sensitivity might further clarify functional mediation of protection.

In conclusion, the result that all evaluated binding and neutralizing antibody markers strongly inversely correlated with COVID-19 risk, and directly correlated with vaccine efficacy, as well as the result that a substantial proportion of the overall vaccine efficacy was mediated through neutralizing antibody levels, adds evidence toward establishing an immune marker surrogate endpoint for COVID-19 vaccines.

## Supporting information

Statistical Analysis Plan

## Data Availability

As the trial is ongoing, access to patient-level data and supporting clinical documents with qualified external researchers may be available upon request and subject to review once the trial is complete.

## Acknowledgments

We thank the volunteers who participated in the COVE trial. We also thank John Mascola, Matt Hepburn, Robert A. Johnson, Mary Marovich, and Merlin Robb.

