## Supplementary Appendix for "Immune Correlates Analysis of the mRNA-1273 COVID-19 Vaccine Efficacy Trial"

This appendix has been provided by the authors to give readers additional information about their work.

- **Figure S3. (A) Cross-validated receiver operating characteristic curve for the 2 top-performing learners, Superlearner, and the Discrete Superlearner models classifying COVID-19 outcome occurrence in baseline SARS-CoV-2 negative per-protocol placebo recipients, with cross-validated area under the curve (CV-AUC) summarizing classification performance. (B) Receiver operating characteristic curve for the Superlearner upon applying the model built from placebo recipients to baseline SARS-CoV-2 negative per-protocol vaccine recipients, with AUC in parentheses summarizing classification performance. .......... 33**

**Immune Assays Team.** (PubMed listed, and ordered alphabetically by affiliation)

| Affiliation | Team Members |
|---|---|
| Biomedical Advanced Research and Development Authority (BARDA), Washington, DC | Christophe S. Badorrek, Oleg Borisov, Flora Castellino, Brett Chromy, Mark Delvecchio, Ruben O. Donis, Tremel Faison, Corey Hoffman, Christopher Houchens, Tom Hu, Chuong Huynh, Pennie Hylton, Lakshmi Jayashankar, Aparna Kolkehar, James Little, Karen Martins, Jeanne Novak, Jeane Russell, Gregory E. Rutkowski, Carol Sabourin, Evan Sturtevant, Xiaomi Tong, John Treanor, Danielle Turley, Leah Watson |
| Boston Consulting Group, Boston, MA | Gian King, Andrew Li, Najaf Shah, Smruthi Suryaprakash, Jue Xiang Wang |
| Division of AIDS, NIAID, NIH, Bethesda, MD | Patricia D'Souza |
| Duke University, Durham, NC | David Beaumont, Rebecca Beerman, Kendall Bradley, Jiayu Chen, Xiaoju Daniell, Thomas Denny, Elizabeth Domin, Amanda Eaton, Kelsey Engle, Wenhong Feng, Juanfei Gao, Hongmei Gao, Kelli Greene, Sarah Hiles, Marianne Jessup-Cumming, Marcella Sarzotti-Kelsoe, Kristy Long, Kellen Lund, Kaia Lyons, Charlene McDanal, David C. Montefiori, Francesca Suman, Haili Tang, Jin Tong, Olivia Widman |
| Vaccine Research Center, NIAID, NIH, Bethesda, MD | Akua Abrah, Obrimpong Amoa-Awua, Manjula Basappa, Robin Carroll, Erykah Coe, Jevone Fentress, Britta Flach, Supra Gajjala, Nazaire Jean-Baptiste, Richard A. Koup, Bob C. Lin, Adrian McDermott, Christopher Moore, Mursal Naisan, Muhammed Naqvi, Sandeep Narpala, Sarah O'Connell, Abhinaya Srikanth, Clare Whittaker, Weiwei Wu |

**Moderna, Inc. Team.** (PubMed listed, ordered alphabetically)

| Affiliation | Team Members |
|---|---|
| Moderna, Inc., Cambridge, MA | Weiping Deng, Shu Hahn, Jacqueline Miller, Rolando Pajon, Honghong Zhou |

**Coronavirus Vaccine Prevention Network (CoVPN)/Coronavirus Efficacy (COVE) Team.** (PubMed listed, and ordered alphabetically by institution affiliation)

| Affiliation/Funding* | Study Group | Location |
|---|---|---|
| AB Clinical Trials | Atoya Adams, MD, MBA, Eric Miller | Las Vegas, NV |
| Accel Research Sites | Bruce G. Rankin DO, John Hill MD, Steven Shinn MD, Marshall Nash MD | DeLand, FL |
| Advanced Clinical Research | Sinikka L. Green MD, Colleen Jacobsen, Jayasree Krishnankutty, Sikhongi Phungwayo | Cedar Park, TX |
| Alliance for Multispecialty Research | Richard M. Glover, II MD, Drs. Stacy Slechta, Troy Holdeman, Robyn Hartvickson, Amber Grant | Newton, KS |
| Alliance for Multispecialty Research | Terry L. Poling MD, Terry D. Klein MD, Thomas C. Klein MD, Tracy R. Klein MD | Wichita, KS |
| Alliance for Multispecialty Research | William B. Smith MD, Richard L. Gibson MD, Jennifer Winbigler MD, Elizabeth Parker PA | Knoxville,TN |
| Baptist Health Center for Clinical Research | Priyantha N. Wijewardane, MD, Eric Bravo MD, Jeffrey Thessing MD, Michelle Maxwell APRN, Amanda Horn APRN | Little Rock, AR |
| Baylor College of Medicine, NIAID 1UM1AI148575-01S2 | Hana El Sahly MD, Jennifer Whitaker MD, Catherine Mary Healy MD, Christine Akamine MD | Houston, TX |
| Benchmark Research | Laurence Chu, MD, R. Michelle Chouteau, MD | Austin, TX |
| Benchmark Research | Michael J. Cotugno MD, George H. Bauer, Jr. MD | Metairie, LA |
| Benchmark Research | Greg Hachigian MD, Masaru Oshita MD, Michael Cancilla NP, Deborah Murray NP, Kristen Kiersey NP | Sacramento, CA |
| Benchmark Research | William Seger MD, Mohammed Antwi, Allison Green, Anthony Kim | Fort Worth, TX |
| Brigham and Women's Hospital, NIAID UM1AI069412, NCATS UL1RR025758 | Lindsey R Baden MD, Michael Desjardins MD, Jennifer A Johnson MD, Amy Sherman MD, Stephen R Walsh MD | Boston, MA |
| Carolina Institute for Clinical Research | Judith Borger DO, Ryan Starr DO, Scott Syndergaard DO, Nafisa Saleem MD | Fayetteville, NC |
| Centex Studies | Joel Solis MD, Martha Carmen Medina PA-C, Westly Keating PA-C, Edgar Garcia PA-C, Cynthia Bueno PA-C | McAllen, TX |
| Clinical Research Atlanta | Nathan Segall MD, Nathan Segall, Jon Finley, Mildred Stull | Stockbridge, GA |
| Clinical Trials of Texas | Douglas Scott Denham DO, Thomas Weiss MD, Ayoade Avworo DNP, Parke Hedges MD | San Antonio, TX |
| Coastal Carolina Research Center | Cynthia Becher Strout MD, Rica Santiago, Yvonne Davis, Patty Howenstine, Alison Bondell | Mount Pleasant, SC |
| Cornell Clinical Trials Unit - Weill Cornell Uptown & Weill Cornell Chelsea, NIAID UM1AI068619, NCAT UL1TR002384 | Kristin Marks MS MD, Grant Ellsworth, MS, MD, Tina Wang, MD, Timothy Wilkin, MD, MPH, Mary Vogler, MD, Carrie Johnston, MD, MS | New York, NY |
| Covid19 Prevention Network (CoVPN, NIAID-NIH) | Michele P Andrasik, Jessica G Andriesen, Gail Broder, Lawrence Corey, Niles Eaton, Kathleen M Neuzil, Huub G Gelderblom, James G Kublin, Rachael McClennen, Nelson Michael, Merlin Robb, Carrie Sopher | Seattle, WA |
| DM Clinical Research | Vicki E. Miller MD, MPH, Fredric Santiago MD, Blanca Gomez FNP-C, Insiya Valika PA-C, Amy Starr FNP-C | Tomball, TX |
| Emory University – Ponce de Leon Clinical Research Site, NIAID 3UM1AI068614-14S1 | Colleen Kelley MD MPH, Valeria D Cantos MD, Sheetal Kandiah MD MPH, Carlos del Rio MD | Atlanta, GA |
| Emory University – Hope Clinic, NIAID 1UM1AI148576-01 | Nadine Rouphael MD, Paulina Rebolledo, Srilatha Edupuganti, Daniel Sans Graciaa | Decatur, GA |
| Emory University School of Medicine, NIAID 1UM1AI148576-01 | Evan J Anderson MD, Andres Camacho-Gonzalez MD, Satoshi Kamidani MD, Christiana A Rostad MD, Meghan Teherani MD | Atlanta, GA |
| George Washington University, NIAID UM1AI068619 | David Joseph Diemert MD, Elissa Malkin, Marc Siegel, Afsoon Roberts, Gary Simon | Washington, DC |
| Hackensack University Medical Center | Bindu Balani MD, Carolene Stephenson, Steven Sperber, Cristina Cicogna | Hackensack, NJ |
| Henry Ford Health System | Marcus J. Zervos MD, Paul Kilgore MD, MPH, Mayur Ramesh MD, Erica Herc MD, Kate Zenlea MPH | Detroit, MI |
| Hope Research Institute | Abram Burgher MD, Ann Marie Milliken | Phoenix, AZ |
| Hope Research Institute | Joseph D. Davis MD, Brendan Levy, Sandra Kelman | Chandler, AZ |
| Hope Research Institute | Matthew W. Doust MD, Denise Sample, Sandra Erickson | Phoenix, AZ |
| J. Lewis Research | Shane Glade Christensen MD, Christopher Matich, James Longe, John Witbeck | Salt Lake City, UT |
| J. Lewis Research | James Todd Peterson MD, Alexander Clark, Gerald Kelty, Issac Pena-Renteria | Salt Lake City, UT |
| Jacksonville Center for Clinical Research | Michael J. Koren MD, Darlene Bartilucci MD, Jeffery Jacqmein MD, Alpa Patel MD, Carolyn Tran MD | Jacksonville, FL |

| Affiliation/Funding* | Study Group | Location |
|---|---|---|
| Javara | Christina Kennelly MD, Robert Brownlee, Jacob Coleman, Hala Webster | Charlotte, NC |
| Johnson County Clin-Trials | Carlos A. Fierro MD, Natalia Leistner, Amy Thompson, Celia Gonzalez | Lenexa, KS |
| Kaiser Permanente Washington Health Research Institute, NIAID 1UM1AI148373-01 | Lisa A Jackson MD MPH, Janice Suyehira MD | Seattle, WA |
| Laguna Clinical Research Associates | Milton Haber MD, Maria M. Regalado MD, Veronica Procasky RN JD, Alisha Lutat | Laredo, TX |
| Lynn Health Science Institute | Carl P. Griffin MD, Raymond Cornelison, William Schnitz, Shanda Gower | Oklahoma City, OK |
| Lynn Institute of the Rockies | Ripley R. Hollister MD, Jeremy Brown DO, Melody Ronk PA-C | Colorado Springs, CO |
| M3 Wake Research | Wayne Lee Harper MD, Lisa Cohen DO, Lynn Eckert PA-C, Matthew Hong MD | Raleigh, NC |
| MediSync Clinical Research Hattiesburg Clinic | Rambod Rouhbakhsh MD, MBA, Elizabeth Danford MD, John Johnson MD, Richard Calderone MD | Petal, MS |
| Meridian Clinical Research | Shishir Kumar Khetan MD, Oyebisi Olanrewaju AC-CRNP, Nan Zhai NP-C, Kimberly Nieves AC-CRNP, Allison O'Brien AC-CRNP | Rockville, MD |
| Meridian Clinical Research | Paul Simon Bradley MD, Amanda Lilienthal MSN NP-C, Jim Callis PA-C | Savannah, GA |
| Meridian Clinical Research | Adam Benson Brosz MD, Andrea Clement PA, Whitney West APRN, Luke Friesen PA, Paul Cramer APRN | Grand Island, NE |
| Meridian Clinical Research | Frank Steven Eder MD, Ryan Little FNP, Victoria Engler FNP, John Tarbox FNP, Heather Rattenbury-Shaw DO | Binghamton, NY |
| Meridian Clinical Research | David Jon Ensz MD, Tavane Harrison, Allie Oplinger | Dakota Dunes, SD |
| Meridian Clinical Research | Brandon James Essink MD, Jay Meyer MD, Frederick Raiser, III MD, Kimberly Mueller APRN, Roni Gray PA | Omaha, NE |
| Meridian Clinical Research | Keith William Vrbicky MD, Charles Harper MD, Chelsie Nutsch MD, Wendell Lewis III MD, Cathy Laflan MD | Norfolk, NE |
| Meridian Clinical Research | Jordan L. Whatley MD, Nicole Harrell MD, Amie Shannon MD, Crystal Rowell APRN, FNP-C, Christopher Dedon APRN, FNP-C | Baton Rouge, LA |
| NIH | Mamodikoe Makhene MD MPH | Bethesda, MD |
| New Horizons Clinical Research | Gregory Mark Gottschlich MD, Kate Harden PA-C, Melissa Gottschlich PA-C, Mary Smith MSN, FNP-C, Richard Powell MD | Cincinnati, OH |
| Optimal Research | Murray A. Kimmel DO, Simmy Pinto MD | Melbourne, FL |
| Optimal Research | Timothy P. Vachris MD, Mark Hutchens MD, Stephen Daniels DO, Margaret Wells MD | Austin, TX |
| Optimal Research | Mimi Van Der Leden MD, PhD, Peta Gay Jackson Booth MD | Rockville, MD |
| Palm Beach Research Center | Mira Baron MD, Pamela Kane DO, Shannen Seversen PA-C, Mara Kryvicky PA-C, Julia Lord PA-C | West Palm Beach, FL |
| Paradigm Clinical Research Center | Jamshid Saleh MD, Matthew Miles, Rafael Lupercio | Redding, CA |
| Quality of Life Medical & Research Centers | John W. McGettigan Jr. MD, Walter Patton MD, Riemke Brakema MD, Karin Choquette MSN, ABNP-C, Jonlyn McGettigan MSN, RN | Tucson, AZ |
| Rancho Paseo Medical Group | Judith L. Kirstein MD, Marcia Bernard NP | Banning, CA |
| Rapid Medical Research | Mary Beth Manning MD, Joan Rothenberg MD, Toby Briskin MD, Denise Roadman PAC, Sharita Tedder-Edwards FNP | Cleveland, OH |
| Research Centers of America | Howard I. Schwartz MD, Surisday Mederos, Barbara Corral, Jennifer Schwartz, Nelia Sanchez-Crespo | Hollywood, FL |
| Rutgers New Jersey Medical School, NIAID UM1AI068619 | Shobha Swaminathan MD, Amesika Nyaku MD MS, Tilly Varughese MD, Michelle DallaPiazza MD | Newark, NJ |
| Saint Louis University, NIAID 1UM1AI148685-01 | Sharon E Frey MD, Irene Graham MD, Getahun Abate MD PhD MSc, Daniel Hoft MD PhD | St. Louis, MO |
| St. Vincent's Health System | Leland N. Allen III MD, Leslie Anne Edwards MSN, CRNP, William Simpson Davis Jr., MS PA-C, Jessica Maria Mena, PA | Birmingham, AL |
| Suncoast Research Group | Mark E. Kutner MD, Jorge Caso MD, CPI, Maria Hernandez Moran APRN, Marianela Carvajal APRN, Janet Mendez APRN | Miami, FL |
| Sundance Clinical Research | Larkin T. Wadsworth III MD, Horacio Marafioti, Lyly Dang, Jennifer Berry, Lauren Clement | St. Louis, MO |
| Synexus Clinical Research | Michael Ryan Adams MD, Leslie Iverson PA | Murray, UT |
| Synexus Clinical Research | Joseph Lee Newberg MD, Laura Pearlman MS, MD, MBA | Chicago, IL |
| Synexus Clinical Research | Paul Joseph Nugent DO, Leonard Singer | Cincinnati, OH |
| Synexus Clinical Research | Michele Diane Reynolds MD, Jennifer Bashour MD, Robert Schmidt MD | Dallas, TX |
| Synexus Clinical Research | Neil Parmanand Sheth MD, Kenneth Steil DO | Glendale, AZ |

| Affiliation/Funding* | Study Group | Location |
|---|---|---|
| Synexus Clinical Research | Ramy Joseph Toma MD, William Kirby MD, Pink Folmar MD, Samantha Williams NP | Birmingham, AL |
| Synexus Clinical Research | Judith White MD, Robert Meyer MD, Sejal Patel MD, Prity Patel APRN | Orlando, FL |
| Tekton Research | Paul Pickrell MD, Stefanie Mott FNP-C, Carol Ann Linebarger MD, Hussain Malbari MD, David Pampe MD | Austin, TX |
| Texas Center for Drug Development | Veronica G. Fragoso MD, Lisa Holloway MD, Cecilia McKeown-Bragas MD, Teresa Becker MD, Vicki Miller MD | Houston, TX |
| Trial Management Associates | Barton G. Williams MD, William H. Jones MD | Wilmington, NC |
| VA Greater Los Angeles Healthcare System | Michael Lewis MD, Elham Ghadishah, Joseph Yusin, Mai Pham | Los Angeles, CA |
| University of California Los Angeles, NIAID UM1AI068619 | Jesse L Clark MD, Steven Shoptaw PhD, Michele Vertucci PA, NP, Will Hernandez NP | Los Angeles, CA |
| University of California San Diego, NIAID UM1AI068636 | Stephen A. Spector MD, Amaran Moodley MD, Jill Blumenthal MD, Lisa Stangl NP, Karen Deutsch NP | La Jolla, CA |
| University of Chicago | Kathleen M. Mullane DO PharmD, David Pitrak MD, Cheryl Nuss FNP, Judy Pi PharmD | Chicago, IL |
| University of Cincinnati, NIAID UN1AI068619 | Carl Fichtenbaum MD, Margaret Powers-Fletcher PhD, Michelle Saemann RN, Sharon Kohrs RN | Cincinnati, OH |
| University of Colorado Denver, Anschutz Medical Campus, NIAID UM1AI068636 | Thomas B. Campbell MD, Andrew Lauria, Jose Castillo Mancilla, Hillary Dunlevy | Aurora, CO |
| University of Illinois at Chicago – Project WISH, NIAID UM1AI068619 | Richard M Novak MD, Andrea Wendrow, Scott Borgetti, Ben Ladner | Chicago, IL |
| University of Maryland School of Medicine, NIAID 1UM1AI148689-01 | Karen L Kotloff MD, Matthew Laurens, Milagritos Tapia, Lisa Chrisley, Cheryl Young | Baltimore, MD |
| University of Miami, NIAID 3UM1AI068614-14S1 | Susanne Doblecki-Lewis MD, Maria Luisa Alcaide, Jose Gonzales-Zamora, Stephen Morris | Miami, FL |
| University of North Carolina at Chapel Hill, NIAID UM1AI068619 | Cynthia Gay MD MPH, David Wohl MD, Joseph Eron, Jr. MD | Chapel Hill, NC |
| University of Pennsylvania, NIAID 3UM1AI068614-14S1 | Ian Frank MD, Debora Dunbar, David Metzger, Florence Momplaisir | Philadelphia, PA |
| University of Pittsburgh Medical Center, NIAID 1UM1AI148452-01 | Judith Martin MD, Alejandro Hoberman MD, Timothy Shope MD MPH, Gysella Muniz MD | Pittsburgh, PA |
| University of Texas Medical Branch, NIAID 1UM1AI148575-01 | Richard Rupp MD, Amber Stanford PA-C, Megan Berman MD, Laura Porterfield MD | Galveston, TX |
| VA Greater Los Angeles Healthcare System | Michael Lewis MD, Elham Ghadishah, Joseph Yusin, Mai Pham | Los Angeles, CA |
| Vanderbilt University Medical Center, NIAID 1UM1AI148452-01 | Clarence Buddy Creech II MD, Shannon Walker MD, Stephanie Rolsma MD PhD, Robert Samuels, Isaac Thomsen MD | Nashville, TN |
| Vanderbilt University Medical Center, NIAID 3UM1AI068614-14S1 | Spyros Andrews Kalams MD, Greg Wilson MD | Nashville, TN |
| Velocity Clinical Research | Gregg H. Lucksinger MD, Kevin Parks MD, Ryan Israelsen MD, Jaleh Ostovar FNP-C, Kary Kelly FNP-C | Medford, OR |
| Velocity Clinical Research, San Diego | Jeffrey Scott Overcash MD, Hanh Chu, Kia Lee, Karla Zepeda | La Mesa, CA |
| VitaLink Research | Luis I. De La Cruz MD, Steve Clemons, Elizabeth Everette, Suzanna Studdard | Greenville, SC |
| VitaLink Research | Gowdhami Mohan MD, Stefanie Tyson, Alyssa-Kay Peay, Danyel Johnson | Anderson, SC |
| VitaLink Research-Spartanburg | Gregory J. Feldman MD, May-Yin Suen, Jacqueline Muenzner, Joseph Boscia, Farhan Siddiqui | Spartanburg, SC |
| Wake Forest University Health Sciences | John Sanders MD,PhD, James Peacock MD, Julio Nasim MD | Winston Salem, NC |
| WR-Clinical Research Center of Nevada | Michael L. Levin MD, Julie Hussey MSN APRN FNP-C, Marcy Kulic MD | Las Vegas, NV |
| WR-ClinSearch | Mark Montgomery McKenzie MD, Teresa Deese, Erica Osmundsen, Christy Sweet | Chattanooga, TN |
| WR-Global Medical Research | Valentine Mbepson Ebuh MD MA MSc, Elwaleed Elnagar MD, Georgette Ebuh DNP APRN FNP-C, Genevieve Iwuala FNP | Dallas, TX |
| WR-Medical Center for Clinical Research | Laurie J. Han-Conrad MD, Todd Simmons MD, Denis Tarakjian MD | San Diego, CA |

*Funding of institutions by the National Institute of Allergy and Infectious Diseases (NIAID) and/or research support by the National Center for Advancing Translational Science (NCATS) as indicated. All other institutions were funded by Office of the Assistant Secretary for Preparedness and Response, Biomedical Advanced Research and Development Authority. The content of this publication is solely the responsibility of the authors and does not necessarily represent the official views of the funding sources.

### CoVPN/COVE Team (cont'd): COVE Trial Investigators and Study Teams

| Principal Investigator | Study Team | Institution | Location |
|---|---|---|---|
| Atoya Adams, MD, MBA | Miriah Campbell, Eric Miller, Daisy Langarica, Alia Bober, Diana Giraldo | AB Clinical Trials | Las Vegas, NV |
| Michael Ryan Adams, MD | Leslie Iverson, Andryelle Toledo, Melinda Bullington, Alicia Hanten, Carolyn Taylor, Shannon Wright, Chase Carnahan, Rachel Law, Natalie Smith, Julie Taylor, Jared-Robert Blake, Stefanie Vasconez, Courtney Jensen | Synexus Clinical Research | Murray, UT |
| Leland N. Allen III, MD | Leslie Anne Edwards, William Simpson Davis, Jr., Ronald Meza, Jordan Stauffer, John Farringer, Faith Holmes, Rhonda Buzbee, Cristina Velez, Huse Lisa, Lisa Huse, Camelia Speegle, Gregory Prestage, Mary Perez, Jessica Space, Matthew Todd, Jessica McDowell, Marha Bunnell-Pollak, Jackie Ziegler, Jasmine Ali, Dumitru Sirbu, Kellie Williams, Logan Sawyer, Richelle Chambliss, Samantha Blackmon, Stephanie Brennan, Tiffany Gibbs, Alexandria Anderson, Caitlin Roll, Candace Robinson, Zachary McCoy, Jessica Bartlett, Kimberly Cornelison, Chris Bovell, Vincent Baglini, Christy Greenhalgh, Jessica Maria Mena, David House, Matt Honold, Esteban Zurita | St. Vincent's Health System | Birmingham, AL |
| Evan J. Anderson, MD | Kathleen Stephens, Francine Dyer, Maya Stagg, Aaliyah Carron, Austin Lu, Julia Barton, Sy Tran, Leisa Bower, Esther Park, Jianguo Xu, Rebecca Gonzalez, Vy Ngo, Mike Shepard, Lezly Roxxette Zepeda, Karen Sytsma, Sandra Rojas-Honan, Felicia Glover, Susan Rogers, Theda Gibson, Christina A. Rostad, Andres Camacho-Gonzalez, Teresa Ball, Satoshi Kamidani, Mehgan Farah Teherani, Vikash Patel, Etza Peters, Peggy Kettle, Lisa Macoy, Cindy Lubbers, Amber Samuel, Laila Hussaini, Kathryn Zaks, Caroline Ciric, Meg Taylor, Oliver Smith, Amy Muchinsky, Sydney Biccum, Laura Clegg, Dean Kleinhenz, Angelle Ijeoma, Hannah Huston | Emory University School of Medicine | Atlanta, GA |
| Lindsey Baden, MD | Xhoi Mitre, Jon Gothing, Bruce Bausk, Jessica Cauley, Natalie Izaguirre, Lewis Novack, Michael Seaman, Katherine Yanosick, Henry Rutherford, Junghyun Kim, Dominique Betterbed, Kathleen Garvey, Lauren Clore, Alexander Mills, Deepesh Duwadi, Alessandra Setaro, Kyl Bowman, Kevin McManus, Sidali Beriane, Fadi Ghantous, Christy Lavine, Jasper Ophel, Joseph Sapiente, Jessica Dorning, Tessa Speidel, Lauren Garneau, Robert Dannemiller, Kirquenique Rolle, Mulika Chhorn, Bailey McCarthy, Hana Flaxman, Milenko Tanasijevic, Cameron Nutt, Javier Barria, Andre Avila-Paz, Buteau Malhaika, Tong Alexandra, Tenaizus Woods, Bethany Evans, Hannah Jin, LaKeisha Gandy, Stephanie St. Pierre, Carolyn Darcy, Michael Corrado, James Maguire, Adetoun Okenla, Tamara Roldon Sevilla, David Kubiak, Cassandre Titus, Movita Harrigan, Maria Alvarado, Rose Theodat, Amy Sherman, Laura Platt, Kirsten Goodman, Laura Nicholson, Wilfredo Matias, Emily Koleske, Ruth Rodriguez, Nicole Taikeff, Jun Bai Park Chang, Julia Klopfer, Phoebe Cunningham, Elizabeth Sampson, Karen Magsipoc, Maureen Macgowan, Lauren Donahue, Haley Schram, Noah Abasciano, Megan Powell, Janet Morgan, Yazed Alsowaida, Olivia Riccardi, Neha Limaye, Virginia Loudermilk, Austin Kim, Kevin Zinchuk, Caitlin Grant, Charles Kelly, David Mellace, Jamie Myers, Erika Gribb, Jose Licona, Monica Feeley, Stephen R Walsh, Jennifer A Johnson, Ann Woolley, Alexis Liakos, Jane Kleinjan, Jon Gothing, Nicolas Issa, Michael Desjardins, Raphael Dolin, Alka Patel, Opeyemi Talabi, Christin Price, Paulette Chandler, Elizabeth W Karlson, Allison P Moriarty | Brigham and Women's Hospital | Boston, MA |
| Bindu Balani, MD | Smith Kerowyn, Sergio Garcia, Charo Valdez, Shelly Chin, Caitlin DiBello, Silvia Lara, Chika Ekwegharir, Abena Roberts, Abimbola Coker, Marie-Therese Estanbouli, Greg Eskinazi, Michael Tortoriello, Jay Elkareh, Meral Karakoc, Olga Spathis, Patrice Hassoun, Carolene Stephenson, Steven Sperber, Kaur Harveen, Cristina Cicogna, Ciaran Mannion | Hackensack University Medical Center | Hackensack, NJ |
| Mira Baron, MD | Pamela Kane, Maria Bermudez, Shannen Seversen, Mara Kryvicky, Julia Lord, Terri Barr, Daisy Acevedo, Elena Acosta, Delta Anderson, Alexandra Arango, Anne Bauer, Joshua Egbehor, Tim Flanary, Audrey Haber, Carol Henao, Patti Isaacson, Peter Jacob, Sakaiya Jackson, Karen Kodes, Ludovic La-Branche, Kimarie Lee-Russell, Carol Liso, Cristina Liso, Stephanie Morse, Michelle Navarrette, Christy Norcross, Nora Norcross, Annette Pitts, Mary Sergalis, David Scott, Tytiana Spearman, Danielle Theodore, Brian Thomas, Jennifer Torres | Palm Beach Research Center | West Palm Beach, FL |
| Judith Borger, DO | Jennifer Angell, Nicole Austin, Deanna Benz, Lucian Cappoli, Nicole Davis, Lynn Eckert, Kathryn Hostetter, Stephanie Keating, Jeanette Mangual-Coughlin, Avia McClain-Stocker, Ifeanyi Momodu, Cheryl Norris, Brennan Opanasenko, Stacey Saldua, Nafisa Saleem, Amy Sheets, Ryan Starr, Scott Syndergaard, Jennifer Thomas, Michelle Wallace, Jeffery Pemberton, Mitchell Arildsen, Dan Tomita | Carolina Institute for Clinical Research | Fayetteville, NC |
| Paul Simon Bradley, MD | Taja Adams, Stephanie Ailey, Kira Bell, Shanice Bennett, Vincent Bernades, Jim Callis, Bounphone Chanthavong. Taryn Collett, Anne Crouch, Shannon Davis, Morgan Deal, Mimi Duncan, Brandon Essink, Laura Falcone, Debra Gabrielson, Brooke Halpern, Anyfa Hanna, Cassie Heisey, Dawn Kalloniatis, Andrew Kimball, Jeanette Lee, Amanda Lilienthal, Ginny McNew, Crystal Neely, Kay Lynn Olmsted, Nicole Osborn, Chevon Roberts, Pechoka Sanders, Cynthia Seedorf, Kathryn Stoddard, Jonathan Whelan, Stella Yoon | Meridian Clinical Research | Savannah, GA |

| Principal Investigator | Study Team | Institution | Location |
|---|---|---|---|
| Adam Benson Brosz, MD | Rhonda Richter, Debra Gabrielson, Kayla Flege, Ashley Bell, Karen Jo Johnson, Paul Cramer, Jessica Stanton, Andrea Clement, Whitney West, Laura Falcone, Amanda Friesz, Kathy Osborne, Summer Tophoj, Kimber Breeden, Susan Newman, Douglas Herbek, Lindsey Mettenbrink, Luke Friesen, Alison Pierce | Meridian Clinical Research | Grand Island, NE |
| Abram Burgher, MD | Stephanie Catanzaro, Shauna Harrell, Magen Hess, Nate Alderson, Bettie D'Nise Corcoran, Norma Frederick, Adrian Alejo, Brian DeCraene, Karen Wakefield, Scarlett Hammett, Susan DeCraene, Ann Marie Milliken, Neil Pearson, Donald Terral Harper | Hope Research Institute | Phoenix, AZ |
| Thomas B. Campbell, MD | Andrew Lauria, Jenelynn Kimble, Steven Johnson, Matin Krsak, Andrew Monte, Patrisha Adkins, Michelle Barron, Suzanne Fiorillo, Amy Harrison, Anderson Victoria, Nga Le, Sara Berech, Jose Castillo-Mancilla, Kristine Erlandson, Laurel Ware, Josie Marshall, Stephen Bartlett, Hillary Dunlevy | University of Colorado Denver, Anschutz Medical Campus | Aurora, CO |
| Shane Glade Christensen, MD | Christopher Mickelson, Jessica Shaw, Emily Raming, Amy Nelson, Gabrielle Lewis, Jenessa Folsom, Mikaela Jones, Dylan Owen, Rachel Pugmire, Jennifer Bradley, Annjanette Kemp, Krista Marti, Allyson Christensen, Madison Ellis, Holly Anderson, Emily Bloomquist, Ross Brunetti, Thomas Conner. Jr., Gina Cox, Diana Grazulis, Wesley Lewis, James Longe, Christopher Matich, Bryan Nelson, Sarah Scott, John Witbeck, Stephen Wood | J. Lewis Research | Salt Lake City, UT |
| Laurence Chu, MD | Jennifer Bacchi, Maria Barrientes, Lamar Box, Christian Casas, R. Michelle Chouteau, Katherine Davis, Tambra Dora, Cindy Duran, Pamela Fidler, Ruth Fitch, Brooke Harris, Isaiah Knight, Jennifer Leyva, Michelle Listz, Jennifer Montes, Javier Perez, Jessica Ruff, Dean Skiles, Sean Turnbow, Francesca Vigil, Breana Wade, Kelly Weber | Benchmark Research | Austin, TX |
| Jesse L. Clark, MD | Sandy MacNicoll, Somaieh Talebi, Timothy Hall, Steven Shoptaw, Emery Chang, Michael Li, David Goodman, Paul Adamson, Oladunni Adeyiga, Inez Bentancourt, Susan Reed, Christopher Blades, Jasmin Tavares, Demetria Villanueva, Simone Riley, Jonathan Veloz, Schuyler Thomas, Will Hernandez, Jennifer Baughman, Mitchell Stern, Michele Vertucci | University of California, Los Angeles | Los Angeles, CA |
| Michael J. Cotugno, MD | Kyra Lawson, Kim Harper, Edwin Adamson, George H. Bauer Jr., Julie Bilich, Brenda Lawson, Brandon Illickal, Lois Eaglin, Heather Salisbury, Jeff Segner | Benchmark Research | Metairie, LA |
| Clarence Buddy Creech II, MD | Shanda Phillips, Naomi Kown, Katherine Sokolow, Wendy Winn, Katherine Wright, Shannon Walker, Stephanie Rolsma, Anna Gallion, April Hanlotxomphou, Deborah Myers, Robert Adkisson, Natalia Jimenez, Cindy Trimmer, Roberta Winfrey, Matthew Donio, John Oleis, Donna Torr, Shelly McGehee, Robert Samuels, Sandra Yoder, Eric Brady, Isaac Thomsen, Madeleine Guy, Emma Alexander, Lana Howard, Krisha Alexander, Shane Moore, Tacora Wright, Tara Evans, Ursula Powell, Jenna Caserta, Valerie Mitchell, Meryk Moore, Melissa Lehman, Diane Anders, Constance Dotye, Crystal Rice, Lamar Bowman, Sherri Hails, Monique Bennett, Nicki Soper, Leigh Howard | Vanderbilt University Medical Center | Nashville, TN |
| Joseph D. Davis, MD | Sandra Kelman, Sandra Braden, Sabrina Bolland, Mia Munoz, Jose Barocio, Brendan Levy, Dhwani Shah, Neil Pearson, Stephanie Catanzaro, Nathan Alderson, Susan DeCraene, Maureen Godfrey, Skyla Clark | Hope Research Institute | Chandler, AZ |
| Luis I. De La Cruz, MD | Amy Ford, Taylor Wilson, Cindy Smith, Austin Lambert, Erin Zeiler, Kaelyn Rowland, Marlee Smith, Suzanna Studdard, Zandra Hamilton, Meredith Benfield, Sara Poff, David Godwin, Elizabeth Everette, Steven Clemons, Kayla Peay, Stephanie Gilreath | VitaLink Research | Greenville, SC |
| Douglas Scott Denham, DO | Thomas Weiss, Parke Hedges, Ayoade Avworo, Kay Scroggins, Leisel Koerber, Antonio Gutierrez, Nathan Cortez, Andrea Gomez, Darlington Akahara, Michelle Smith, Kristy Trevino, Beatriz Herrera, Shaiane Dickerson, Kerry de Jesus, Matthew Korte, Cynthia Ramos, Reanna Martinez, Erica Leal, Shakera Flores, Paul Esparza, Brian Hemming, Melinda Axton, D'Andre White, Terri Perez, Carolina Coronado, Rebecca Many, Clayton Stone, Kimberly Evans, Anshumaan Maharaj, Stephen Brick, Steffanie Barrera, Staci Poettgen, Dawn Killian, Gerardo Pena, Karol Perez, Victoria Hernandez, Kevin Martinez, Amy Griffith, Nolan Payton, Quincey Hogue, Jamie Padilla, Emily Mendez, Lily Hays, Maristelle Co, Nicholas Trinidad, Ismael Rodriguez, Amy Lewis, Cindi Nellis, Lele Simmons, Marissa Johnson | Clinical Trials of Texas | San Antonio, TX |
| David Joseph Diemert, MD | Linda Witkin, Aimee Desrosiers, DeEnna Wedding, Bertran Walton, LaKeisha Queen, Ryan Mouton, Caroline Thoreson, Manya Magnus, Jennifer Wald, Erika Faust, Nicholas Heredia, Robbie Kattappuram, Hira Qadir, Chelsea Ware, Hannah Yellin, Kegan Dasher, Daniel Mullen, Jeanne Jordan, Taylor Ladson, Madison Lintner, Kaitlyn Macnair, Bitana Saintilma, Kelly Thomas, Samantha Walker, Neha Rampally, Madhu Balachandran, Elissa Malkin, David Parenti, Hana Akselrod, Marc Siegel, Gary Simon, Afsoon Roberts, Aileen Chang | George Washington University | Washington, DC |

| Principal Investigator | Study Team | Institution | Location |
|---|---|---|---|
| Susanne Doblecki-Lewis, MD | Maria Luisa Alcaide, Jose Gonzalez-Zamora, Stephen Morris, Yimy Puerto, Annie Salvarrey, Claudia Balgas, Claudia Santos, Katherine King, Brahian Steven Erazo, Mayra Fernandez, Leopoldo Cordova-Garcia, Elisa Corzo-Sanchez, Edgar Fernandez, Loreta Padron, Stefani Ann Butts, Kenia Moreno, Juan Casuso, Maria de Pilar Valanzasca, Thomas Tanner, Marilyn Fernandez, Mary Aloise, Inza Patton, Vivian Pastrana, Sendy Puerto, Irma Barreto Ojeda, Junlin Long, Barbara Huang, Gilianne Narcisse, Vanessa Perez | University of Miami | Miami, FL |
| Matthew W. Doust, MD | Denise Sample, Sandra Erickson, Nate Alderson, Adrian Alejo, Stephanie Catanzaro, Susan DeCraene, Cassie Enricco, Sandra Erickson, Alex Guereque, Shauna Harrell, Shana Harshell, Stephanie Junker, Stephanie Laufenberg, Madison Mikulak, Makayla Morra, Nicole Olson, Neil Pearson, Jasmin Redden, Monique Romo, Denise Sample, Dhwani Shah, Sahara Vega, Emma Kar | Hope Research Institute | Phoenix, AZ |
| Valentine Mbepson Ebuh, MD | Elwaleed Elnagar, Georgette Ebuh, Genevieve Iwuala, Catina Adams, Marissa Cervenka, Ezgar Del Real, Shraddha Dubal, Elwaleed Elnagar, Jenifer Fiatte, Kathy Harrell, Genevieve Iwuala, Vicki Martinez, Robert Miranda, Brennan Opanasenko, Destiny Robinson, Liz Ruiz, Amy Sheets, Shoniece Wallace | WR-Global Medical Research | Dallas, TX |
| Frank Steven Eder, MD | Ryan Little, Victoria Engler, John Tarbox, Heather Rattenbury-Shaw, Deborah Hubish, Jessie Taylor, Debra Gabrielson, Jessica Fellows, Jennifer Molstead, Kathe Olmstead, Ashley Conover, Tammy Kohn, Chelsea Briar, Corrine Young, Collen McVannan, Kelli Quick, Shaylynne Hubanks, Kimber Breeden, Ann Marie Sampson, Traci Hull, Tarin Gordon, Susan Owen, Kate Macarak, Tonya Rackett, Jacob Blattstein, Partidge Jane Aton, Nicole Croft, Carolyn Grausgruber, Rebecca Miller, Ryan Little, Victoria Engler, John Tarbox, Heather Rattenbury-Shaw, Nathan Kimball, Courtney Heisey, Ginny McNew, Abigail Wine, Cindi VanKuren, Jared Frick, Tammy Dennis, Andrew Kimball | Meridian Clinical Research | Binghamton, NY |
| Hana M. El Sahly, MD | Jennifer A. Whitaker, C. Mary Healy, Christine Akamine, Wendy A Keitel, Robert L Atmar, Annette Nagel, Sandra Francisco, Thea Marie Cordero, Janet Brown, Jennifer Christensen, Caroline Doughty-Skierski, Connie Rangel, Carrie Kibler, Coni Cheesman, Lisreina Toro, Chanei Henry, Chianti Wade Bowers, Pedro Piedra, Kathy Bosworth, Kayla Burrell, Jesus Banay, Tykel Eddy, Trent Davis, Shetel Anassi, Yvette Rugeley, Olga Rybina-Willis | Baylor College of Medicine | Houston, TX |
| David Jon Ensz, MD | Pamela Allen, Taylor Bergh, Kimber Breeden, Avery Dunn, Brandon Essink, Debra Gabrielson, Rylea Gulick, Tavane Harrison, Courtney Heisey, Andrew Kimball, Shelby Klaschen, Jessica Knight, Makayla Langston, Meagan Miller, Allie Oplinger, Heather Persinger, Alison Pierce, Kathryn Stoddard, Kayla Sturgeon, Jamie Thompson, Melissa Wiseman | Meridian Clinical Research | Dakota Dunes, SD |
| Brandon James Essink, MD | Jay Meyer, Frederick Raiser, Kimberly Mueller, Roni Gray, Riley Brockman, Tabitha Campbell, Carrie Essink, Laura Falcone, Roni Gray, Linda Layton, Jay Meyer, Kimberly Mueller, Tiffany Nemecek, Frederick "Fritz" Raiser, III, Jessica Satorie, Chelsea Steinmetz, Nicole Osborn, Cassie Heisey, Maria Nguyen | Meridian Clinical Research | Omaha, NE |
| Gregory J. Feldman, MD | May-Yin Suen, Brittany Cooksey, Madison Fowler, Sarah Chynoweth, Gary Clemons, Laura Jolly, Charlie Jordan, Heather Allison, Steve Clemons, Amber Brittany Belcher, Allison Kelly, Marsha Gossett, Wendy Taylor, Amy Witt, Kendal Nelson, Jeffrey Witt, Jacqueline Muenzner, Elizabeth Everette, Supinder Channa, Allison Ayers, Joseph Boscia, Farhan Siddiqui | VitaLink Research-Spartanburg | Spartanburg, SC |
| Carl J. Fichtenbaum, MD | Maggie Powers-Fletcher, Michelle Saemann, Sharon Kohrs, Kimberly Mullins, Lindsay Davis, Moises Huaman, Angela Snyder, Kristin Weghorn, Brenda Miller, Elizabeth Costea, Lisa Schira, Romana Saeed, Helen Shelton, Kathleen Ballman, Laura Browning-Cho, Sherry Donaworth, Chris Goddard, Jeanine Goodin, Elizabeth Niederegger, Lisa Hachey, Tamara Maus, Pam Fletcher, Makayla Bishop, Victoria Straughn, Shaina Horner, Carrie Christofield, Dana Burns, Jason Mayes, Kelly Windholtz, Lisa Proffitt, Faizan Qureshi, Michelle O'Neil, Arustamyan Lisa, Sarah Trentman, Eva Whitehead, Jennifer Baer, Linda Hinds, Jaasiel Chapman, D'Vaughn House, Gary Frazier, Judy Houston, Lisa Altenau, Mary Burns, Dorice Smith, Justin Ragle, Eric Mueller, Cynthia Nypaver, Jaime Robertson, Anissa Moussa, Geronimo Feria Garzon, Sierra Bennett, Marlena Petrie | University of Cincinnati | Cincinnati, OH |
| Carlos A. Fierro, MD | Mazen Zari, Celia Gonzalez, Natalia Leistner, Mary Easley, Mary Provost, Krista Estrada, Ann Geier, Amy Thompson, Heather Barker, Karol Moore, Kelly Moen, Monica Atwood, Amber Wolf, Brandi Dickerson, Manyvohn Rinehart, Dina Hammine, Angela Eichler, Casey Johnson, Nathan Arthur | Johnson County Clin-Trials | Lenexa, KS |
| Veronica G. Fragoso, MD | Lisa Holloway, Cecilia McKeown-Bragas, Teresa Becker, Vicki Miller, Leena Mir, Elton Oliveira, Moez Talpur, Enya Rentas-Sherman, Gabriela Maria Becerra, Dewayne Hicks, Robert Krbashyan, Shakira Barr, Ashraf Jafri, Herman Ortiz, Zohair Harianawala, Chandra Tobin, Norma Gonzalez, Saji Perinjelil, Khorshid Amirkhosravi, Tracy Kowalski, Biman Goswami, Waheeda Sureshbabu, Amy Anderson, Berenice Ferrero, Simeen Khan, Chen-Ho Yang, Nazanin Zarinkamar, Scott Ward, Crystal Reese, Miyosha Lewis, Olga Konshina, Lorrian Yates.Joel Cano, Quiana Wilson, Kara | Texas Center for Drug Development, Inc. | Houston, TX |

| Principal Investigator | Study Team | Institution | Location |
|---|---|---|---|
| | Sikes, Diana Chehab, Joanna Quezan, Maryam Rabbani, Sadaf Batla, Abbyssinia Moges. Diego Carrington, Matthew Joseph, Laura Grissanty, Dean Jang, Dustin McFadden, Misbah Baloch, Elisa Moralez, Abdeali Dalal, Frances Saubon, William Fernandez, Jenny Toress, Blessing Felix, Zain Rizvi | | |
| Ian Frank, MD | Annet Davis, Eileen Donaghy, Nicole Sundo, Juan Ramirez, Laura Schankel, Dana Brown, Katharine Bar, Dana Brown, Christopher Chianese, Gillain Constantino, Dovie Watson, Kathleen Degnan, Helen Koenig, William Short, Petra Alexander, Eileen Mergliano, Jie Ho, Michele Wisniewski, Debora Dunbar, Liani Santini-Lopez, Rosemarie Kappes, Angela Cabassa, Tammy Chen, Berry SotoVega, Deborah Kim, Devon Cliett, Kate Kearns, Jillian Baron, Vivian Leung, Florence Momplaisir, Sarah Wood, Tameka Matthews, David Metzger, Richard Tustin | University of Pennsylvania | Philadelphia, PA |
| Sharon E. Frey, MD | Irene Graham, Getahun Abate, Daniel Hoft, Heather Douds, Cassandra Zehenny, Joan Siegner, Helay Hassas, Kim Cooper, Shirley Dettlebach, Sabrina DiPiazza, Carol Duane, Linda Eggemeyer-Sharpe, Lauren Foreman, Jerry Hutter, Ryan Kerr, Kate Liefer, Tracy Montauk, Karla Mosby, Janice Tennant, Nicole Purcell, Kiana Wilder, Kathleen Chirco, Sharon Irby-Moore, Kathleen Koehler, Melissa Loyet, Thomas Pacatte, Susan Stewart, Azra Blazevic, Tamara Blevins, Chase Colbert, Christopher Eickhoff, Lainey Mejia Jauregui, Keith Meyer, Krystal Meza, Amanda Nethington, Huan Ning, Brittany Williams, Mei Xia, Yinyi Yu, Stanley Doublin, Mary Pat Eastman, Eric Eggemeyer, Mikayla Frye, Michelle Harris, Aleshia McCoy, Donna Duncan, Gwendolyn Tatum, Nicole Purcell, Kiana Wilder, Tammy Grant, Claudia Castillo Paredes, Rong Hou, Jin Wang, Qian Wang, Sarah George | Saint Louis University | St. Louis, MO |
| Cynthia Gay, MD | David Wohl, Joseph Eron, Jr., Andrew Thorne, Michelle Floris-Moore, Christopher Hurt, David Wohl, Chidinma Okafor, Janette Goins, Ulrike Adam, Ekundayo Nylander-Thompson, Anna Furlong, XinHong Ao, Kathy Guerrero, Melinda Hart, Kathleen Loeven, Rachael Mossey, Esther Speight, Rachel White, Chloe Twomey, Kristen Gray, Miriam Chicurel-Bayard, Susanne Henderson, Patti Vasquez, April Welch, Camille O'Reilly, Maureen Furlong, Noshima Darden-Tabb, Elizabeth DuBose, Marie Oriol, Dynesha Perry, Maria Stetson, Maria Bullis, Shelby Turner, Ebony Harrington, Michael Herce, Suzanne Blevins, Alexander Bradley, Susan Pedersen, Becky Straub, Sandra Barnhart, Felicia Barriga Munante, Nazneen Howerton, Tevnan Keller, Mandy Tipton, Abigail Riddick, Kristi Kirkland, Maggie Harman, Tania Hossain, Centhla Washington, Erin Hoffman, Carolina Pastrana Medina, William Johnson, Samantha Earnhardt, Amy James Loftis, Catherine Kronk, Yaa Ofori-Marfoh, Julie A Nelson, Nicole Maponga, Lina Rosengren-Hovee, William Zhao, Jennifer Thompson, Sarah Law, Holly Milner, Jonathan Oakes, Rachel Cook, Erin Cardot, Oesa Vinesett, Victoria Rucinski, Joy Wannamaker, Tanailly Giralt Smith, Eliza DuBose, Chidinma Okafor | University of North Carolina at Chapel Hill | Chapel Hill, NC |
| Richard M. Glover, II, MD | Stacy Slechta, Troy Holdeman, Robyn Hartvickson, Amber Grant, Jennifer Bennett, Lindsey Brewer, Janelle Brown, Kelsey Burden, Melissa Burton, Brianna Burton, Jordan Danby, Sheri Duncan, Amber Grant, Robyn Hartvickson, Lisa Hemmelgarn, Sherry Henning, Jeri King, Riley King, Colton King, April Kitterman, Shannen Lassiter, Cayla Lawless, Janna Martinez, Ragene Moore, Marissa Mueller, Aaron Nguyen, Justin Phillips, Jordan Reheis, Rebecca Ring, Katherine Saengerhausen, Shannon Thomas, Dylan Thomas, Cindy Thome, Denae Villines, Amber Wenzel, Eilleen Wilbert, Avi Woods, Caressa Presley, Brianna Newport, Olivia Allen, Miranda Santiago, Cheryl Sauerwein, Jill Longstaff, Sadie Allen, Candace Heckart | Alliance for Multispecialty Research | Newton, KS |
| Gregory Mark Gottschlich, MD | Melissa Gottschlich, Steven Anderson, Gregory Mark Gottschlich II, Mary Woeste, Kate Harden, Cindy Young, Michael Pordy, Audrius Ruksenas, Lacy Baird, Kim Krogman, Lori Stanton, Melissa Fuson, Mason Urban, Christine Watson, Richard Powell, Mary Smith, Jacob Sekinger, Diamond Russell, Nicole Lim, Mylene Asmar-Rios, Yusef Museitif, Craig Mitchell, Tarik Whitham, Zachary Rutledge, Troy Porter, Andrea Newlands, Jami Ramsey, Mary Frances Curry, Nishay Holloman, Crystal Barket, Michelle Spear, Shelley Mahan, Taeleigha Greene, Zachary Eardley, Gen Moussa, Mary Ann Gottschlich | New Horizons Clinical Research | Cincinnati, OH |
| Sinikka L. Green, MD | Julie Hamilton, Alex Fuller, Jeanette Dickhaus, Colleen Jacobson, Triny Cooper, Michelle Jackson, Taylor Evans, Tabitha Judd, Kathryn Alexander, Megan Rosallo, Sikhongi Phungwayo, Robin Dotson, Dana Finley, Michael Vasquez, Cyndi Foster, Gregg Lucksinger, Sarah Smiley, Jayasree Krishnankutty, Ray Coon, Grishma Dhimmer, Melanie Wilkerson, Tatum Shawver, Marcedes Coffman, Devin Teal, Laura Crenshaw | Advanced Clinical Research | Cedar Park, TX |
| Carl P. Griffin, MD | William Schnitz, Andrea Romero, Kim Hamilton, Raymond Cornelison, Angela Genovese, Shelly Brunson, April Green, Lacey Dietz, Kim Calloway, Chris Hyatt, Destiny Heinzig-Cartwright, Chalimar Rojo, Sharee Wright, Kathi Shaw, Michael Pojezny, Avery Keller, Krystal Hightower, Dalia Tovar, Shanda Gower | Lynn Health Science Institute | Oklahoma City, OK |
| Milton Haber, MD | Maria Candelario, Martha Bunnell-Pollak, Lauren Wade, Jackie Ziegler, Deena Ramirez, Perla Avalos, Maria Drada, Jasmine Ali, Jessica McDowell, Kehinde Busari, Patricia Church, Ronald Meza, Marco Vela, Esteban Zurita, Chris Connolly, Ruben | Laguna Clinical Research Associates | Laredo, TX |

| Principal Investigator | Study Team | Institution | Location |
|---|---|---|---|
| | Del Bosque, Alisha Lutat, Chelsea Fleming, Brett Potthoff, Anita Suri, Cynthia Priester, Brenda Hernandez, Veronica Procasky, Eva Cerreta, Matt Honold, Melinda Rodriguez, Maria Regalado, Jordan Stauffer | | |
| Greg Hachigian, MD | Michael Cancilla, Ricardo Castellanos, Angela Cuellar, Yaman Darmarathne, Shaila Faulker, Yana Gordeyeva, Michelle Hisey, Ashley Jungsten, Kristin Kiersey, Pawandeep Nagra, Nav Nagra-Kooner, Jazmin Nauta, Masaru Oshita, Kenneth Quick, Julie Raygoza, Amanny Sadek, Melisa Tinder, Jhoana Torres, Deborah Murray, Kristen Kiersey | Benchmark Research | Sacramento, CA |
| Laurie J. Han-Conrad, MD | Brandon Baldwin, Lucian Cappoli, Tenisha Garcia, Ella Grach, Brenda Grande, Nicolle Mendez, Natalie Moy, Matthew Musikanth, Karen Mylerberg, Brennan Opanasenko, Mark Pulera, Patti Sanchez-Emery, Mireles Sarah, Todd Simmons, Denis Tarakjian | WR-Medical Center for Clinical Research | San Diego, CA |
| Wayne Lee Harper, MD | Toni Bland, Lori Bridges, Lucian Cappoli, Lisa Cohen, Leah Corts, Annie Craft, James Earnhardt, Lynn Eckert, Aubrey Farray, Laura Hoer, Matthew Hong, Chris Hoyle, Jenee Jiggetts, Brian Joseph, Bradley Killebrew, Kendra Lisec, Lucie Mangala, David Musante, Adnan Nasir, Amanda Olsen, Brennan Opanasenko, Marci Parks, Marion Peoples, Katherine Schuch, Judith Shand, Sabine Ucik, Douglas Wadeson, Barbara Wheeler | M3 Wake Research | Raleigh, NC |
| Ripley R. Hollister, MD | Jeremy Brown, Brandy Ball, Jeremy Brown, Valerie Dyster, Dalia Jeronimo, Shelby Pickle, Michael Pojezny, Melody Ronk, Kathi Shaw, Bobbi Shofner, Jami Wagner, Meghan York, Jill York | Lynn Institute of the Rockies | Colorado Springs, CO |
| Lisa A. Jackson, MD, MPH | Marilyn Nguyen, Maya Dunstan, Barbara Carste, Sarah Friend, Diana McFeters, Lynn Gross, Mohamed Ajenah, Jana Ffitch, Audra Mccoy, David Skatula, Susan Lasicka, Kimberly Brinker, Karen Sherwin, Melissa Scheer, Paula Lins, Roger Calvert, Roxanne Erolin, Stella Lee, Vi Tran, Stephanie Pimienta, Bruce Douglas, Lee Barr, Colin Fields, Erika Kiniry, Joe Choe, Janice Suyehira, Joyce Benoit, Michael Witte, Rebecca Lau | Kaiser Permanente Washington Health Research Institute | Seattle, WA |
| Spyros Andrews Kalams, MD | Greg Wilson, Kyle Rybczyk, Katie Crumbo, Carly Griffin, Latoya Hannah, Amy Kerrigan, Valerie Mitchell, Jenna Caserta, Mary Downey, Nicole Swindle, Shonda Sumner, Amber Massey, Trudy Sullivan, Rita Smith, Cindy Nochowicz, Eric Olson, Christian Warren, Josh Simmons, Dana King, Gwendolyn Rees, Matt Donio, Jesse Case, Keith Richardson, Jarissa Greenard | Vanderbilt University Medical Center | Nashville, TN |
| Colleen Kelley, MD, MPH | Valeria D. Cantos, Sheetal Kandiah, Carlos del Rio, Christina Bacher, Hannah Huston, Juliet Brown, Divya Bhamidipati, Nithin Gopalsamy, Brittany Lynn Speigel, Elizabeth (Betsy) Hall, Brandon Spratt, Kiran Dhillon, Caitlin Moran, Michael Chung, Felecia Wright, Marcia Peters, Rondell Jaggers, Vanessa Soliman, Ron Gaston, Christopher Foster, Sarah Wiatrek, Bezuayehu Mandefro, Pamela Weizel, Pamela Lankford-Turner, Anandi Sheth, John Gharbin, Catherine Abrams, Philip Powers, Paulina Rebolledo, Christin Root, Tiraje Lester, Sha Yi, Damien Swearing, Fred Ede, Isaac Perez, Kelly Likos, Meen Dhir, Aastha KC, Gabriela Georgial, Tucker Colvin, Nabeel Yar Khan, Valarie Hunter, D'Jamel Young, Felecia Atkinson | Emory University<br>Emory University – Ponce de Leon Clinical Research Site | Atlanta, GA |
| Christina Kennelly, MD | Jacob Coleman, Brittany Bundeff, Melissa C. Hennessey, Kenneth Owen, Caroline Wilds Wilds, Jennifer Womack, Susan Martello, Chiedza Hooker, Robert Brownlee, Melissa James, Deborah Wesley-Farrington, Lori Whiteheart, Hala Webster, David Framm, Cortney Fretz, Gwyn Gibson, Susan Donahue, Kelly Woodell, Linda McCarty, Jim Vesely, Scott Chatterton, Andrew Ottesen, Enrico Belgrave, Krishna Shah, James Chester Alexander, Brittain Callahan | Javara | Charlotte, NC |
| Shishir Kumar Khetan, MD | Taja Adams, Tanya Alexander, Tanya Alexaner, Sydney Barmoy, Jake Bart, Kira Bell, Ira Berger, Jemario Blackwell, Priscilla Buahin, Bounphone Chanthavong, Juliana DeVito, Azure Erskine, Brandon Essink, Laura Falcone, Debra Gabrielson, Beau Garland, Barb Geiger, Tiana Oliver, Courtney Heisey, Sucharita Katikala, Andrew Kimball, Heather Lang, Jeanette Lee, Asefa Mekonnen, Devan Myers, Kimberly Nieves, Allison O'Brien, Oyebisi Olanrewaju, Nicole Osborn, Adunola Oshiyoye, Rahul Patel, Alan Pollack, April Poole, Collin Smith, Kathryn Stoddard, Chao Wang, Sean Whelan, Jonathan Whelan, Graciela Zapata, Nan Zhai | Meridian Clinical Researc | Rockville, MD |
| Murray A. Kimmel, DO | Alexa Diec, Ann Riley, Bette Denmat, Bram Swarr, Christina Raidl, Dania Billman, Denise Dixon, Donald Dawson, Elaine Crudo, James Crowley, Katrina Carlson, Kaylie Worzick, Laura Worth, Lisbeth Gordon, Marion Oliver, Robert Holt, Simmy Pinto, Taylor Atkinson, Traci Mitchell, Lana Ghomrawi, Norma Rokoff | Optimal Research | Melbourne, FL |
| Judith L. Kirstein, MD | Jared Bradshaw, Krista Forster, Jeanette Dickhaus, Marcia Bernard, Erica Sanchez, Nikki Abels, Cynthia Kunakom, Vanessa Vandergoot, Jessica Fisher, Carol Remigio, Jourdan Manfred, Frederick Lloyd, Tiffany Williams, Clarisse Baudelaire, Lovette Cherelle, Nolan Mackey, Alan Valenzuela, Theodore Wyman, Alyssa Taber, Karen Myers, Craig Koch | Rancho Paseo Medical Group | Banning, CA |
| Michael J. Koren, MD | Shannon Trull, Amanda Elwood, Mary Strickland, Ivy Gulliermo, Chistopher Ganzhorn, Sonia Gerardo, Taylor Johnson, Victoria Kaposchansky, Cassie Lawler, Laura Little, Amanda Pratt, Sheldon Warren, Andrea West, Emery Noles, Nathanial | Jacksonville Center for Clinical Research | Jacksonville, FL |

| Principal Investigator | Study Team | Institution | Location |
|---|---|---|---|
| | Grant, Jillian Agnew, Lori Alexander, Brenda Anderson, Deirdre Arrington, Sara Benner, Lisa Carl, Allison Crain, Nafisa Ishaku, Robert Nix, Sharon Smith, Amber Devries, Sandy Salceiro, Opara Chukwudi, Mikaela Karney-Trull, Ramil Castillo, David Graham, Gail Lowe, Alexander Hill, Carolyn Tran, Jeffry Jacqmein, Darlene Bartilucci, Alpa Patel, Janet Garvey, Mitchell Rothstein, Kenneth Aung-Din, Margaret Gannaway, Arman Mughal, Sandra Fuit, Jolenne Wolfer, Erin Schelhorn, Jacob Wolfer, Madison Martinez, Melissa Parks, Patricia Neal | | |
| Karen L. Kotloff, MD | Matthew Laurens, Milagritos Tapia, Lisa Chrisley, Cheryl Young, Barbara Albert, Robin Barnes, Shernel Barrett, Andrea Berry, Melissa Billington, Shannon Bittner, Colleen Boyce, Faith Pa'Ahana Brown, James Campbell, Regina Carpenter, Jamonie Carter, Ginny Cummings, Brenda Dorsey, Jorge Flores, DeAnna Friedman-Klabanoff, Shirley George, Nancy Greenberg, Hassan Haji, Elizabeth Hammershaimb, Susan Holian, Leslie Howe, Myounghee Lee, Alyson Kwon, Kirsten Lyke, Alma Valle Maldonado, Jennifer Marron, Kaitlin Mason, Monica McArthur, Rosa McBryde, Sherry McCammon, Sandra Molina, Kathleen Neuzil, Daniele Nitkowski , Justin Ortiz, Rekha Rapaka, Mardi Reymann, Toni Robinson, Wanda Somrajit, Mark Travassos | University of Maryland, School of Medicine | Baltimore, MD |
| Mark E. Kutner, MD | Amanda Colina, Isett Caro, Frances Beltran, Jessie Downs, Jonathan Fernandez, Mariete Renden, Mirnaya Mujica-Alabaci, Susel Figueredo, Yanelis Dominguez, Jaime Blandon, Bryan Ruiz, Leidy Montoya, Edgardo Rodriguez, Jessie Downs, Jason Rothscheld, Janett Acle, Yaime Martinez, Soraya Ricardo, Maria Hernandez Moran, Eloisa Guerra, Heidie Perez, Claudia Rodriguez, Victoria Moreno, Vanessa Hechavarria, Saray Carvajal, Daniel Lopez, Carlos Ivircu, Neiner Enriquez, Paola Garcia, Chris Hoyle, Marianela Carvajal, Janet Mendez, Edisleidy Mesa, Marco Ramirez, Dalila Del Valle, Jennifer Ortega, Yeni Hernandez, Jhobana Vargas, Carmen Amador, Juan Delgado, Maury Santos, Meredith Arguelles, Leyanis Coello, Vanessa Ansorena, Jorge Caso, Stacy Machado, Raydel Valdes, Giann Lightbourn, Dayami Dovales, Alain Chang | Suncoast Research Group | Miami, FL |
| Mimi Van Der Leden, MD, PhD | Chrishea Harvey, Tricia Oyeyemi, Aicha Moutanni, Stephanie Melton, Peta-Gay Jackson Booth, Jennifer Yoon, Gloria Kim, Atanas Filev, Francis Uwandi, Meyling Lopez, Janice Spreitzer, Courtney Gennes, Xiangfei Cheng, Matthew Van Sickle, Nick Bart, Brianne Okunji, Frank Maloba | Optimal Research | Rockville, MD |
| Michael L. Levin, MD | Brennan Opanasenko, Yajaira Ramos, Shonda Lester, Rebecca Boucher, Shawn Harrell, Shon Boucher, Patti Sanchez, Nina Scharbach, Alex Sanchez, Shyane Raniello, Wendy Guerra, Krystal Tyner, Kimberly Temple, Ruby Ortiz, Daniel Terreault, Amy Kill, Jade Odynski, Adolfo DeLeon, Debbie Carter, Eduardo Rodriguez, Julia Gass, Sara Esparza, Sierra Dansbee, Tammy Harrison, Marcy Kulic, Lucian Cappoli, Mora KIm, Matthew Fenner, Heather Jimenez, Shraddha Dubal, Julie Hussey | WR-Clinical Research Center of Nevada | Las Vegas, NV |
| Michael Lewis, MD | Nancy Mohler, Mai Pham, Ron Waldorf, Elham Ghadishah, Samantha Feril, Stella Lee, Dzuyen Nguyen, Ruoxiang Wang, Justine Velandria, Benjamin Dreskin, Joseph Yusin, Lauren Vigil, Sara Wong, Suchi Tiwari, Joseph Pisegna, Sunita Dergalust, Wayman Lee, Krissa Caroff | VA Greater Los Angeles Healthcare System | Los Angeles, CA |
| Gregg H. Lucksinger, MD | Jaleh Ostovar, Craig Koch, Danuel Hamlin, Kelly Chase, Jeanette Dickhaus, Edward Kerwin, Frederick Forde, Allison Alvord, Dawn Stewart, Dan Hamlin, Kevin Parks, Ryan Israelsen, Kary Kelly, Tiffany Smith, Melissa Myers, Ryan Rackley, Audrey Kuehl, Savannah Peterson, Hannah Hall, Jay Weisbart, Alison Dodenhoff, Emily Kelly | Velocity Clinical Research | Medford, OR |
| Mary Beth Manning, MD | Carol Salango, Alec Ireland, Lisa Hoagland, Jeanette Dickhaus, Toby Briskin, Joan Rothenberg, Michael Gaston, Sharita Tedder-Edwards, Denise Roadman, Megan Sokolowski, Tina Shickluna, Katherine Bielanski, Samantha Hood, Talia Chandler, Brianna Arman, Melinda DeLong, Naqib Ahmad, Karly Tarase, Jade Svoboda, Lisle Merriman, Melisa Sebera, Emma Landskroner, Amy Maroun, Brooke Glivar, Jennifer Gaston, Sarah Dzigiel, Cassiandra Uminski, Karol Sabol, Devan Patel, Nick Zarbo, Briana Jackson, Brian Sharpe, Nicole, Baitt, Kaitlyn Duffy, Gabrielle Jacobs, Ann Czuprun, Tracee Cash, Diamond Ivey, Kaitlyn Rubell | Rapid Medical Research | Cleveland, OH |
| Kristen Marks, MD | Grant Ellsworth, Tina Wang, Timothy Wilkin, Mary Vogler, Carrie Johnston, Marshall Glesby, Roy Gulick, Ole Vielemeyer, Rebecca Fry, Todd Stroberg, Caitlin Rhoades, Noah Goss, Shaun Barcavage, Valery Hughes, Jonathan Berardi, Caroline Greene, Sarah Galloway, Caique Mello, Ashley Machado, Mia Crowley, Monique Williams, Katherine Fee, Elizabeth DeJesus, Andrew Yu, Minkyung Lee, Susan Herder, Mary Ann Zweibel, Patrice Weller, Antonio Rivera-Lopez, Edward Kenny, Hetal May, Natella Fridman, Parul Shah, Ruby Lee, Venus Fernandez, Victoria Lesina, Celine Arar, Byron Bullough, Kinge-Ann Marcelin, Brian Mangano, Jessenia Fuentes, Jiamin Li, Genessi Rodriguez, Catherine Jerry, Nadi Islam, Liqun Cai, Wayne Burns, Akinbayo Caulcrick, Andrika Thomas, Barbara Batog, Guoan He, Sara Yoder, Tamara Crowder, Gianna Resso, Sophia Alvarez, Tahera Begum, Elizabeth Connolly, Roxanne Rosario, Paul Kim, Steven Wang, Vasilika Koci | Cornell Clinical Trials Units - Weill Cornell Chelsea and Uptown | New York, NY |

| Principal Investigator | Study Team | Institution | Location |
|---|---|---|---|
| Judith Martin, MD | Alejandro Hoberman, Timothy Shope, Gysella Muniz, Sonika Bhatnagar, Kumaravel Rajakumar, Anne-Marie Rick, Peri Unligil, Jennifer Nagg, Melissa Andrasko, Mary Ann Sieber, Jennifer Opal, Lalicia Roman, Spenser Kinsey, Michelle Burke, Matthew Lee, Dominic Kramer, Linette Milkovich, Emily Dougherty, Emily Carney, Shannon Mance, Nader Shaikh, Diana Kearney, Jamie Fries, Lisa Vavro, Shayla Goller | UPMC University Center | Pittsburgh, PA |
| John W. McGettigan, Jr., MD | Walter Patton, Jennifer Schnider, Riemeka Brakema, Heeten Desai, Mikell Brett Karsten, Patricia Jalomo, Cindy Finch Benoy, Karin Choquette, Jonlyn McGettigan, Yvonne De Los Reyes, Melissa Cozzens, Amanda Hermosillo, Cindy Montgomery, Susan Tarwid, Annette Elzy, Tianna Young, Saysamone Banks, Cristina Fernandez, Damaris Atondo, Zoe Sesma, Norma Barrientos, Maggie Tono, Kisha Adams, JoAnn Wilkins, Arianna Bermudez, Carol Sayer, Julie McDowell, Angelina Navarro, Mercedes Sullivan, Crystal Mata, Sheldon Gingrich, Aaliyah Sestiaga, Gia Longo | Quality of Life Medical & Research Centers | Tucson, AZ |
| Mark Montgomery McKenzie, MD | Tiffany Jewell, Zackery Harmon, Michael Elizabeth, Christy Sweet, Teresa Deese, Catherine Schon, Misti Earwood, Lou Cappoli, Brennan Opanasenko, Lisa Guider, Michelle Forgey, Justian Jarrett, Rachel Scott, Elizabeth Michael, Erica Osmundsen, Andrew Wood, Shelly Brooks, Gisela Heintz, Lilian Nukuna | WR-ClinSearch | Chattanooga, TN |
| Vicki E. Miller, MD | Sajjad Naqvi, Soofia Masood, Fredric Santiago, Sonia Guerrero, Subhash Koneru, Nirja Shah, Andrea Torres, Ramani Gali, Talha Baig, Heather Leary, Afifah Ayub, Nayab Goher, Patti Tate, Reagen Reed, Muhammad Irfan, Amy Starr, Alefiyah Motiwala, Julia Kenny, Victoria Aguilar, Jessica Arguijo, Insiya Valika, Victoria Aguilar, Jagruti Patel, Anna Pena, Faryal Mahmood, Blanca Gomez, Nancy Torres, Kristyn Latil, Tarori Mark, Laura Djampou, Lindsey Kueng, Marianne Tadros, Mohammad Millwala, Monica Murray, Murtaza Marvi, Shivani Shah, Vanessa Gonzalez, Zohair Harianawala, Zainab Rizvi, Ambily Dileep, Jaquelyn Gonzales, Ragen Powell, Carolina Deandres, Syed Fahad Ali Kazmi, Sandra Natalia Perez, Shannon Amacker, Shiela Varghese | DM Clinical Research | Tomball, TX |
| Gowdhami Mohan, MD | Rodolfo Barrera, Emma Partin, Kelly White, Ashley Rochester, Charles Thompson, Stefanie Tyson, Ashten Sheriff, Alyssa-Kay Peay, Kayla Corn, Barbara A. Richardson, Kristin Miller, Steven Clemons, Cameron King, Emma Partin, Gary Clemons, Brianna Starr, Danyel Johnson, Taylor Davis, Niki Tyson | VitaLink Research | Anderson, SC |
| Kathleen M. Mullane, DO, PharmD | David L. Pitrak, Cheryl Nuss, Karen Cornelius, Randee Estes, Amy Luckett, Michelle Moore, Judi Pi, Stephen Schrantz, Jill Stetkevych | University of Chicago | Chicago, IL |
| Joseph Lee Newberg, MD | Mary Reyes, Nicole Leahy, Victoria Andriulis, Herbert Whinna, Patricia James, Lana Ghomrawi, Carole Kempfer, Miriam Arroyo, Maria Castro, Anna Maddox, Reuben Martinez, Jacquilyn McCormick-Burks, Laura Pearlman, Rosalinda Vazquez, Shaheera Suleiman, Neha Atal, Rosalind Vazquez | Synexus Clinical Research | Chicago, IL |
| Richard M. Novak, MD | Regina Harden, Maria Schwarber, Michael Pacini, Rebeca Gansari, Margie Villarreal, Stephanie Martin, Michelle Lee, Richard Morrissy, , Taylor Ellis, Samuel Rene, Tara Cobbs, Claudia Preciado, Scott Borgetti, Maximo Brito, Olamide Jarrett, Mahesh Patel, Tracy Cable, Charity Ball, Maryann Holtcamp, Rodrigo Burgos, Sarah Michienzi, Emily Drwiega, Mikayla Johnson, Fischer Herald, Benjamin Ladner, Minseung Chu, Carolyn Dickens, Alfredo Mena Lora, Stockton Mayer, Andrea Wendrow, Habiba Sultana, Nanu Nunwar, David Chan, Marla Schwarber, Khandaker Anwar, Mahmood Ghassemi, Md Ruhul Amin, Doris Carroll, Rosa Valencia, Michelle Agnoli, Elena Llinas, Samuel Rene, Liam Morrissy, Adrian Raygoza, Addis Mekkonnen, Lisa Lindemann, Daniel Meslar, Karen Pacini, Corey Ringhisen, Amy Kennedy-Krage, Claudia Miller, Lorna Sanchez McCann, Gizelle Alvarez, Nia Moragne-Oneal, Nusirat Williams, Ian Feather, Nikki Griffith, Wardrick Nealon, Renyce Powell, Nila Safaeian, Monica Gingell, Diana Bahena, Gerald Beck, Brad Farrington, Rod Reyes, Monica Wilson, Juline Wondrasek, Kimberly Shapiro, Shannon Whitted, Victoria Roehl, Braulio Carrasco, Michael Chen, Olivia Murray, Yasiel Lacalle, Tessa Eckley, Anna Schluckebier, Kevin Cao, Elise DeBruyn | University of Illinois at Chicago - Project WISH | Chicago, IL |
| Paul Joseph Nugent, DO | Leonard Singer, Jennifer Jones, April Smith, Georgettea Geuss, Lana Ghomrawi, Christine Bennett, Norma Blevins, Linda Brotherton, Michele Byrd, Krista Doss, Victoria Holden, Christine Hull, Jean Montgomery, Nancy Cipollone, Savanah Torline, Brandon Brown, Meagan Thomas, Katie Ziska, Dana Sias, Hannah Wagner | Synexus Clinical Research | Cincinnati, OH |
| Jeffrey Scott Overcash, MD | Hanh Chu, Kia Lee, Karla Zepeda, John Rodriguez, Adam Prince, Yashveer Dubbula, Elizabeth Tomatsu Michael Voskanian, Crystle Rajania, Stephanie Ramirez, Claudia Camacho, Lauren Arnett, Kecia Darbeau, Ashley Smith, Kimberly Quillin, Cesar Ramirez, Daniel Robitaille, Erica Sanchez, Allie Davis, Michael Waters, Pat Kappen, Valerie Horne, Thao Vuong, Andrew Dennis, Nikki Abels, Dominique Panis, Richard McQuaid, Whitley Harbison, Erika Trujillo, Andrea Garcia, Jose Jacob Esparza, Carlos Vera, Raquel Taitingfong, Cathy Meza, He Pu, Jackielynn Smith, Shandel Odom, Zahira Nieves, Ashliegh Lindsay, Ariana Nasatka, Jose Cazarez, Nora Martinez, Angela Hunt, Antonio Delgado, Linda Vega, Angela Anorve, Erica Martinelli, Melania Riordan, Sylvia Lindholm, Gina Ciezkowski, Grecia Perez, Jacob Pineda, | Velocity Clinical Research, San Diego | La Mesa, CA |

| Principal Investigator | Study Team | Institution | Location |
|---|---|---|---|
| | Nathan Tyler, Ranya Salem, Amara Yilmaz, Jessica Gonzales, Zabrina Ruiz, Laura Castillo, Yajaira Contreras, Angelica Guzman, Makenna Orel, Jeffery Alvarez, Gordon Bovee, Roxana Ramirez, Joan Esquivel | | |
| James Todd Peterson, MD | Christopher Mickelson, Madeline Maldonado, Alison Charlton, Ashley Bragg, Sean Hansen, Emily Wilcox, Colby Bostock, Megan Henry, Pam Iwasaki, Bradley Young, Katelyn Walker, Joy Nguyen, Lindsey Bevan, Megan Grimmett, Madeline Grote, Heather Littell, Natalie Bee, Alexander Clark, Shana Eborn, Susan Edwards, Dan Henry, Heather Jackson, Gerald Kelty, Issac Pena-Renteria, Jacqueline Rohrer, Jack Taylor, Brooke Barrick, Ty Henry, Anna Dansie, Kenadie Hamblin | J. Lewis Research | Salt Lake City, UT |
| Paul Pickrell, MD | Susan Bonner, Blaire Graham, Staci Taggart, Hussain Malbari, Tiffany Lemuz, Ethan Shotton, Andrew Bell, Megan Malek, David Pampe, Carol Ann Linebarger, Michelle Peterson, Brandi Chalman, John Luna, Elizabeth Santellanes, Christina Martinez, Lisa Johnson, Lisa Savage, Melissa Winn, Wendi McKenzie, Eileen Euperio, Stefanie Mott, Paul Menefee, Katie Caballero, Darrell O'Brien, Morgan Schulle, Kate Jurek, Olivia Hapanowicz | Tekton Research, Inc. | Austin, TX |
| Terry L. Poling, MD | Meenakshi (Kavya) Natesan, Patricia Contreras, Denise Hole, Avi Woods, Jill Hiebert, Melissa Burton, Olivia Eagleson, Laura Holz, Terri Ford, Cindy Thome, Terry D Klein, Gregory Greer, Diandra Henriques, Tracy R Klein, Thomas C Klein, Christa Shue, Gina Young, Brenna Sprout | Alliance for Multispecialty Research | Wichita, KS |
| Bruce G. Rankin, DO | Jennifer Dittman, Lora Parahovnik, Crystal Paccione, Melissa Hodges, Katina Marchione, Matt Maxwell, Any Dominy, Diana Toney, Andrea Marrafino, Laura Isbell, Leandro Fernandez, Claxton Copeland, Michelle Tutt, Adam VanDeusen, Kevin Feldman, Clark Mason, Tifany Huertas, Over Seijas, Jennifer Cline, Christian Beierschmitt, Ryan Hobbick, Jessica Gilliam, Jeanette de Leon, Iman Mencia, Daniel Layish, Vienna Bauer, Shatonia Fields, Albert Garcia, Carrie Rycort, Tasha Brocato, Marshall Nash, Samantha Watts, Amy Houck-Dominy, Angela Hammerle, Teresa Logsdon, Erika Wierzbicki, Taylor Martin, Ranie Hutchins, Fadhel Alyunis, Gail Lavine, Jeffery Hood, Robert Duran, Michelle Jones, Ginny McClanahan, Heather Jackson, Leandra Fernandez, Douglas Winter, Antonio Rivera, Amber Vasquez, Thais Truffa, Daniel Campbell, Grace Newcomb, Elizabeth Orlando, Steven Shinn, John Hill, Christina Isbell, Dhaneshwar Oomrow, Alicia Cevera | Accel Research Sites | DeLand, FL |
| Michele Diane Reynolds, MD | Jennifer Bashour, Robert Schmidt, Cynthia Mayeux, Uvoka Huffman, Lisa Nicholson, Jacklyn Newton, Lynn Yauch, Cathy Monroe, Kathleen Carty, Angelica Banks, Taylor Werner, Pamela Echols, Pauline Jackson, Chana Hines, Lorine Cook, Cristina Puig, Patrick Brooks, Jennifer Ruiz, Deanna Bowman, Ladina Garcia | Synexus Clinical Research | Dallas, TX |
| Rambod Rouhbakhsh, MD, MBA | John Johnston, Richard Calderone, Tasha Stevenson, Tameka Fortune, Brandi Pace, Adreanna Pou, Jerrica Sullivan, Yolanda Lewis, April Rouse, Tiffany Jefferson, Elizabeth Danford, Jeff Repper, Mason Boutwell, Alexycia Washington, Krista Hirth, Meagan Grabel | MediSync Clinical Research Hattiesburg Clinic | Petal, MS |
| Nadine Rouphael, MD | Renata Dennis, Tigisty Girmay, Michelle Wiles, Sharon Curate-Ingram, Lauren Hewitt, Alexis Ahonen, Mari Hart, Sarah Bechnak, Erin Carter, Lauren Nolan, Daniel Sans Graciaa, Geoffrey Kamau, Easton Beshears, Sy Tran, Mary Atha, Mary Bower, Ghina Alaaedine, Brandy Johnson, Jacob Usher, Eileen Osinski, Erin Scherer, C. Tae Stallworth, Stephanie Ramer, Rose Pope, Esther Park, Francine Dyer, Laura Clegg, Rebecca Gonzalez, Stacey Wheeler, Susan Rogers, Vy Ngo, Vanessa Soliman, Kristen Unterberger, Bernadine Panganiban, Christopher Huerta, Juton Winston, Ali Alvarez, Jianguo Xu, Colleen Kelley, Paulina Rebolledo, Nicholas Scanlon, Jessica Traenkner, Matthew Collins, Hollie Macenczak, Cassie Grimsely-Ackerley, Tiffany Lee, Amy Anderson, Michele Paine McCullough, Hannah Huston, Daniella Carter, Lisa Harewood, Srilatha Edupuganti, Varun Phadke, Mindee Adamson, Jeanne Allen, Debbie Bartenfeld, Lily Berz, Amy Cromwell, Sergio Cruz, Fred Ede, Monica Godfrey, Evan Gutter, Angelle Ijeoma, Sara Jo Johnson, Vinit Karmali, Dean Kleinhenz, Jennifer Kleinhenz, Alexandra Koumanelis, Maranda Leary, Tiraje Lester, Juliet Alise Morales, Shashi Nagar, Julia Paine, Dilshad Rafi Ahmed, Brittany Robinson, Amanda Rosner, Renee Silver, Trevor William Simon, Talib Sirajud-Deen, Damien Swearing, Maliya Tolbert, Pamela Turner, Chia Uzuegbunam, Claire Wan, Dongli Wang, Erika Wimberly, Jean Winter, Joy Winters, Yong Xu, Sha Yi | Emory University - Hope Clinic | Decatur, GA |
| Richard Rupp, MD | Amber Stanford, Megan Berman, Laura Porterfield, Gerianne Casey, Hala Ghoson, Doreen Jones, Michael Willig, Cori Burkett, Robert Cox, Amy McMahan, Diane Barrett, Kristin Pollock | University of Texas Medical Branch | Galveston, TX |
| Jamshid Saleh, MD | Matthew Miles, Rafael Lupercio, Vicky Martin, Marla Clark, Matthew Pohlmeyer, Ruba Zanaid, Veronica Blevins, Tara Ulberg, Carlyee Chambers, Marisol Corrales, Emily Crews, Mohamed Yassin, Sarah Sandberg, Frank Chen, Mandy Swanson | Paradigm Clinical Research Center | Redding, CA |
| John W. Sanders, MD, MPH | Stacy Harpe-Hall, Jesse Hopkins, Ann Schweppe, Jaymous Fayssoux, Kathryn Bender, James Peacock, Katharine Pearsall, Brandy Snyder, Deidre Knox, Megan Thorpe, Melissa Ellingson, Brittany Bundeff, Lisa Ashworth, Meredith Hiatt, Ritu Rathee, Stacy Woodliff, Brian Strittmatter, Amanda Wright, Daisy DeWeese-Gatt, | Wake Forest University Health Sciences | Winston Salem, NC |

| Principal Investigator | Study Team | Institution | Location |
|---|---|---|---|
| | Caryn Morse, John Williamson, Samantha Wheeler, Lori Whiteheart, Susan Donahue, James Lovette, Kaitlyn Van Leuvan, Kelly Ledbetter, Scott Chatterton, Julio Nasim, Amie Sidberry, Ashley Davis, Carter Noecker, Chie Hooker, Johanna Breenan, Sam Cable, Anna Bowman, Stephanie Boothe, Shea Overcash | | |
| Howard I. Schwartz, MD | Carlos Valladares, Jocelyn Morrera, Yulexis Amestoy, Tori Wallenburg, Thelma Beltran, Terry Piedra, Monica Garces, Alexandra Galvis, Wanda Delgado, Catherine Casas, Lesly Miguel Sosa, Vivian Rosales, Jose Fernando Henriquez, Mikael Yaniz, Beatriz Rivera, Peter Ventre, Gabriella Huyke, Maria Companioni, Jessie De Vega, Brianna Gamez, Stephanie Diaz, James Jean-Mary, Americo Padilla, Nikita Notise, Yorlina Luquetta, Monifa Wilson-Morris, Kenia Gutierrez, Roilan Garcia, Karla Pentzke, Leyda Valentin, Lazara Novas, Marilein Camacho, Jazmin Henfield, Laymis Alvarez, Myriam Rosado, Maxine Bryant, Maria Pinero, Laura Raucci, Francisco Ramirez, Angelic Gamez, Mailin Perez, Yasmin Baddour, Hary Leon Joseph, Yaquelin De la Cruz, Dunia Torres, Rosaidaliz Carreira, Chanella Garcia, Surisaday Mederos, Jose Muniz, Karenda Plotka, Sara Gomez, Maria Soto, Cathy Cruz, Nelia Sanchez-Crespo, Jennifer Schwartz, Barbara Corral, Matthew Muniz, Dayana Deltejo, Ana Castro, Reem Hassan | Research Centers of America | Hollywood, FL |
| Nathan Segall, MD | Michelle Sowell, Nancy Levine, Erynn McKinley, Hannah Smith, Karen Hickson, Elizabet West, Patrizia Greene, Jon Finley, Mildred Stull, Susan Jones, Jennifer LeBrun, Pamela Talbott, Kwannda Whatley, Jeffrey Jones, Michelle Binns, Donna Toepfer, Cynthia Steele, Grace Newville, Gillian Waite, Cynthia Pinckney, Karen Yangapatty, Kiara Tyner, Kimberly Cobb, Kourtney Richardson | Clinical Research Atlanta | Stockbridge, GA |
| William Seger, MD | Kimberly Pullen, Jean Seignon, Anthony Kim, Mohammed Antwi, Allison Green, Lizzy Seger, Elizabeth Boydston, Abdur Rafay Qadri, Deborah Devlin, Tasha Todd, Oluwatosin Akingbala, Alma Guel, Tisha Davis, Melody Dufrene, Samantha Loudermilk, Virginia Loudermilk, Crystal Starr, John Villegas, Ben Seger, Katherine Hollie | Benchmark Research | Fort Worth, TX |
| Neil Parmanand Sheth, MD | Kenneth Stell, David Beckett, Enitt Gonzalez, Donna McGunigal, Amanda Burns, Nancy Wood, Shelley Miceli, Christina Avila, Rebecca Baker, Laura Vigliotti, Sarah Kading, Samer Salama | Synexus Clinical Research | Glendale, AZ |
| William B. Smith, MD | Richard L Gibson, Jennifer Winbigler, Elizabeth Parker, Madison Watts, Suzann Cloninger, Talya Thomas | Alliance for Multispecialty Research | Knoxville,TN |
| Joel Solis, MD | Martha Carmen Medina, Xavier Morales, Hank Heller, Blake Torrence, Joanna Gurrola-Mahoney, Cynthia Bueno, Heather Holloway, Irving Salinas, Joel Perez, Paola Garcia, Erica Canales, Blanca Urbina, Brancisilio Gutierrez, Carolina Cantu, Chelsea Vargas, Cindy Vasquez, Cody McIntire, Gabriela Gutierrez, Hugo Sosa, Irvin Munoz, Jessica Estrada, Jonna Lopez, Kaegan Knox, Mirella Melendez, Natalia Valle, Natalie Echavarria, Nicole Litton, Amber Victor, Nancy Torrence, Madhu Shreya, Mathew Maran, Asfak Alam, Westly Keating, Tara Green, Devora Torrence, Gerardo Sedas, Shruti Konda, Prem Jangam, Mario Echavarria, Alejandro Silva, Anne McNulty, Daniel Contreras, Daniel Gomez, Edgar Garcia, Elizabeth Weber, Luis Lopez, Samuel Ramirez, Kayla Lopez, Pedro Penalo, Angel Salinas, Jaime Solis, Shannon Moyer, Aryana Ibarra, Guadalupe Gurrola, Jenna Anastasiades, Uchechi Ehiemua, Sara Solorzano | Centex Studies, Inc. | McAllen, TX |
| Stephen A. Spector, MD | Amaran Moodley, Jill Blumenthal, Baharin Abdullah, Christina Addington, Juan Carlos Alcantar, Deyna Arellano, Bernadette Cale, Brendan Costello, Tammelita Cotlon-Pineda, Fanny Delebecque, Karen Deutsch, Aram Dimayuga, Son Do, Yasmeen Esshaki, Aileen Everhart, Cindy Ewing, Veronica Figueroa, Medardo Gaytan, Crystal Groom, Carolyn Hernandez, Heather Huitema, Benjamin Hull, Sylvia Isaac, Jaclyn Jaskowiak, Cindy Knott, Leander Lazaro, Thuan Le, Megan Loughran, Michelle Madey, Rosalva Martha-Patten, Colleen McLellan, Jeff Ledford-Mills, Asami Mimura, Patty Moraes, Jennifer Morales, Jessica Nasca, Phirum Nguyen, Marielys Padilla-Martinez, Dennis Perpetua, Mike Pizza, Shannon Ransom, Emily Rizo, Carlos Rojas, Thaine Ross, Marie Sagrado, Eugene Sato, Lisa Stangl, Ji Sun, Nancy Tang, Mina Trivedi, Rodney Trout, Donna Voss, Lindsey Woronicz | University of California, San Diego | La Jolla, CA |
| Cynthia Becher Strout, MD | Rica Santiago, Yvonne Davis, Patty Howenstine, Alison Bondell, Jaime Robertson, Anissa Moussa, Geronimo Feria Garzon, Sierra Bennett, Marlena Petrie | Coastal Carolina Research Center | Mount Pleasant, SC |
| Shobha Swaminathan, MD | Amesika Nyaku, Tilly Varughese, Rondalya Deshields, Michelle L DallaPiazza, Elise Lewis, Jennifer Punsal, Mario Portilla, Malithi Desilva, Christina Daliani, Susana Rivera, Aidan Ziobro, Andressa Rebellatto, Brian Murloy, Christina Ninan, Ernest Pianim, Eunice Wang, Merit Henen, Muhammad Usman, Rebecca Kim, Shiao Wang, Gener Eric Cruz, Bethany Birago, Joyell Arscott, Dina Meawad, Christie Lyn Costanza, Francesca Escaleira, Zoraida Cruz-Barahona, Jared Khan, Valeria Cadorett, Jamir Tuten, Travis Love, Eric Asencio, Sukhwinder Singh | Rutgers New Jersey Medical School | Newark, NJ |

| Principal Investigator | Study Team | Institution | Location |
|---|---|---|---|
| Ramy Joseph Toma, MD | Olivia Graves, Josiah Robinson, Patricia Hammonds, Lana Ghomrawi, Kara Quinnelly, Shaun O'Conor, Michael Lambe, Rachell Stewart, William Kirby, Pink Folmar, Rachel Culbreth, Heidi Leblanc, Julie McDaniel, Rian Montgomery, Andrea Woodle, Samantha Williams, Hunter Russell, Shereen Lowe, Maureen Mayer, Hollis Ryan, Elaine Reese | Synexus Clinical Research | Birmingham, AL |
| Timothy P. Vachris, MD | Mark Hutchens, Stephen Daniels, Margaret Wells, Sandra Clancy, Rebecca Martinez, Jessica Buot, Merissa Daugherty, Julie Hamilton, Kimberly Hernandez, Ashli Alejandro, Amy Collins, Monique Gawlik, Patricia Johnson, Maria Moreno, Ashley Washington, Tina Rountree, Daniel Dore, Ravi Davuluri, Ashlee Brunaugh, Jorge Martinez, James Hermon, Vianai Carreno, Mia Rountree, Colleen Coelho | Optimal Research | Austin, TX |
| Keith William Vrbicky, MD | Charles Harper, Chelsie Nutsch, Wendell Lewis III, Cathy Laflan, Linden DeBoer, Kayla Andal, Misty Appeldorn, Jenniger Grebe, Russell Herstein, Catherine King, Samantha Wieseler, Alisha Kiepke, Christy Lee, Kelsey Kelley, Kelli James, Ashley Frisch, Courney Green, Taysha Hingst, Jeni Hoppe, Kimber Breeden, Debra Gabrielson, Ginny McNew | Meridian Clinical Research | Norfolk, NE |
| Larkin T. Wadsworth III, MD | Ashley Dale, Christy Schultz, Rebecca Munsch, Anya Penly, Liz Garner, Stephanie Tesson, George Cherniawski, Angie Kean, Dan Reed, Courtney Kubiak, Maureen Dempsey, Heather Cherniawski, Breanna Galibert, Kristin Branson, Laura Hartupee, Karen Knapp, Horacio Marafioti, Lyly Dang, Jennifer Berry, Lauren Clement, Megan Dandurand | Sundance Clinical Research | St. Louis, MO |
| Jordan L. Whatley, MD | Patricia Whatley, Christopher Dedon, Anika Payne, Amie Shannon, Kristen Losavio, Nicole Harrell, Mary Margaret Dobson, Lindsey Hall, Chaney Bennett, Crystal Rowell, Mimi Dimmick, Amy Thomassie, Kimber Breeden, Cody LaFleur, Makaylea Truitt, Taryn Collett, Emily Best, Alexandra Caillouet, | Meridian Clinical Research | Baton Rouge, LA |
| Judith White, MD | Amy Edridge, Chelsea Montalvo, Eugenia Clark, Lisa Russell, Zahra Somji, Lesli Leimer, Robert Meyer, Christine Murphy, Prity Patel, Sejal Patel, Ruben Moliere, Samantha Merveillard, Yarnick Mirjah, Bryn Walls, Joey Cruz, Aaron Cooper, Jessica Bienaime, Ashley Gilcrist, Alisa Petit, Tyler Knightly, Kimberly Stokes, Christina Rosario, Talhia Matos, Ilona Boggs, Nicholas Weber, Felix Busot, Linda Colon, Heather Gillenwater, Cristina Kaplun, Melissa Caputi, Shayna Siplin, Daminee Shah, Samuel Martin, Alexis Waldorf, Vihar Upadhyay, Adolfo Henriquez, Saskia Singh, Maria Roberts, John Caporelli, Shirley Salvador, Quevina Scarver, Vanessa Garcia, Taylor Moore, Jayasen Singh, Curshinda Galvin-Burch, Mary Kesner, Jasmin Gil, Shay Gray, Steven Monsegur, Michele Steinmetz, Michael Lambe, Heather Powell, Sandra Torres, Shaban Katbeh, Taylor Wilson | Synexus Clinical Research | Orlando, FL |
| Priyantha N. Wijewardane, MD | Natalie Johnson, Martha Evans, Sondra Wright, Richard Pellegrino, Lastida Burns, Natasha Williams, Haylee Rowe, Kayla Graham, Amanda Horn, Eric Bravo, Jeffrey Thessing, A. Michele Maxwell, Amy Cooper, Lauren Evans, Tonya Cato, Haylee Tucker, Lesa Gann, Hannah Jones, Amanda May, Tiffany Walker, A. LeiAn Diaz, Laura Khalil, Lydia Purcell, Timothy Campbell, Charlotte Garcia-Velez, Andrea Scarborough, Beatrice A. Miller, Keith Bracy, Aujania Thompson, Cassandra Johnson, Krishana Day, Freddie Hicks, Jamie Pettus | Baptist Health Center for Clinical Research | Little Rock, AR |
| Barton G. Williams, MD | Flo Abbott, Nicole Burton, Alice Cipollini, Madison Croucher, Philip Dattilo, Erin Harrelson, Kelsey Heston, James Ingram, William H Jones, Karla Lane, Brandy Lowman, Evan Lucas, Megan Marles, Morgan Mathis, Angie Northcott, Clyda Pasquantonio, Alyssa Valente, Ciara Winders, Stephanie Graham | Trial Management Associates | Wilmington, NC |
| Marcus J. Zervos, MD | Paul Kilgore, Mayur Ramesh, Jelena Verkler, Pardeep Pabla, Andrew Clark, Katrina Williams, Dee Dee Wang, Beverley Duthie, Samia Arshad, Alandra White, Anna Kern, Ashley Mattern, Bilqis Mosed, Dana Parke, Doreen Dankerlui, Dragana Spasevska, Hanah Woods, Helina Misikir, Howard Klausner, Janay Scott, Jessica Heinonen, John Zervos, Joseph Miller, Kate Zenlea, Kristin Eis, Marissa Vasquez, Maurice Slaughter, Meaghan Flynn, Michael Garcia, Michelle Sankah, Nina Paeilli, Philip Benson, Robert Devore, Stevanya Baho, Tony Eljallad, Tyler Prentiss, Yaman Ahmed, Sharon Mathys, Linda Kaljee, Jeffrey Van Laere, Claudia Hanni, Hassan Zafar, Mona Desai, Gina Maki, Mary Perri, Dora Vager, Shannon Thomas, Autumn Robinson, Isis Hamilton, Sonia Eliya, Jehan Jazrawi, Biljana Popovic, Sharon Zahul, Joshua Ruzzin, John Laguio, Ali Mathena, Bobby Cook Jr., Marlene Hesler, Rochelle Fleming, Terria Minniefield, John Simons, Sherese Henderson, Ashley Hopkins, Rebecca McFarlane, Raeshell Carson, Jonathan Williams, Katherine Reyes, Erica Herc, Indira Brar, Mayur Ramesh, John McKinnon, Lacquis Duncan, Tim Asmar, Margaret Beyer, Kaleem Chaudhry, Madison Lee, Jo-Ann Rammal, Karthik Sridasyam, Siddesh Veer, Angelique Buluran, Kimberlyn Lott, Jeremiah Rooker, Alayna Wilder, Kathleen Wilson, Allison Weinmann, Hassan Mourtada | Henry Ford Health System | Detroit, MI |

**United States Government (USG)/Coronavirus Prevention Network (CoVPN) Biostatistics Team.**
(PubMed listed, and ordered alphabetically by institution affiliation)

| Affiliation | Team Members |
|---|---|
| Biomedical Advanced Research and Development Authority (BARDA), Washington, DC | Di Lu, James Zhou |
| Department of Biostatistics and Bioinformatics, Rollins School of Public Health, Emory University | David C. Benkeser |
| Vaccine and Infectious Disease Division, Fred Hutchinson Cancer Research Center, Seattle, WA | Jessica Andriesen, Bhavesh Borate, Lindsay N. Carpp, Andrew Fiore-Gartland, Youyi Fong*, Peter B. Gilbert*, Ying Huang*, Yunda Huang, Ellis Hughes, Ollivier Hyrien, Holly E. Janes*, Michal Juraska, Yiwen Lu, April K. Randhawa, Brian Simpkins, Brian D. Williamson, Lars W.P. van der Laan, Chenchen Yu |
| Biostatistics Research Branch, NIAID, NIH, Bethesda, MD | Michael P. Fay, Dean Follmann, Martha Nason |
| Division of Biostatistics, School of Public Health, University of California, Berkeley, CA | Nima S. Hejazi |
| Department of Biostatistics, University of Washington, Seattle, WA | Marco Carone, Kendrick Li, Wenbo Zhang |
| Department of Statistics, University of Washington, Seattle, WA | Alex Luedtke |
| Department of Population Health Sciences, Weill Cornell Medical College,New York, New York | Iván Díaz |

*YF, PBG, YH, and HEJ are also affiliated with the Department of Biostatistics, University of Washington, Seattle, WA.

**Figure S1. Timing of mRNA-1273 doses, blood sampling, and the two time periods for diagnosis of COVID-19 endpoints ("Intercurrent" and "Post Day 57").** The schematic applies to baseline SARS-CoV-2 negative per-protocol recipients of two doses of mRNA-1273.

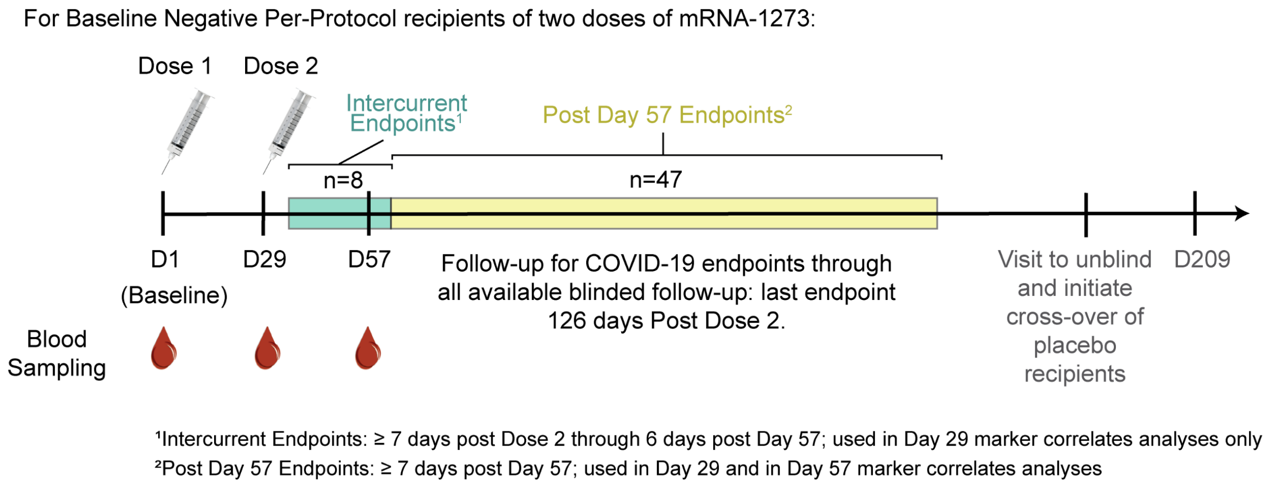

**Figure S2. Flowchart of study participants from enrollment to the case-cohort set of baseline SARS-CoV-2 negative per-protocol participants.**

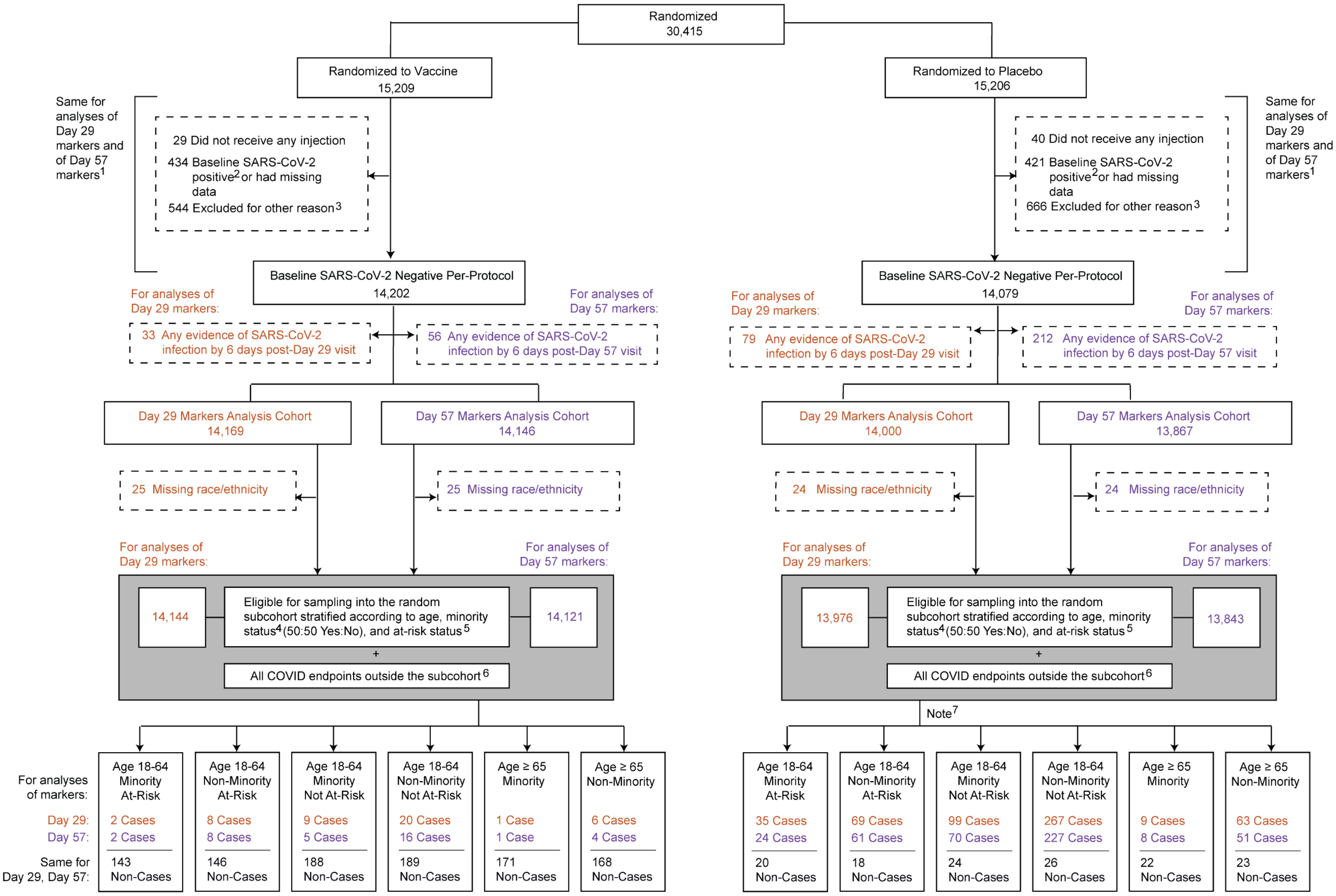

1 If a participant had multiple reasons for exclusion from the Per-Protocol Set, they were only counted in the earliest-listed reason.
2 Baseline SARS-CoV-2 positive is defined as immunologic or virologic evidence of prior COVID-19 (i.e. a positive nasopharyngeal swab and/or binding antibodies against SARS-CoV-2 nucleocapsid above the limit of detection or above the lower limit of quantification) at Day 1 before the first dose of investigational product (same definition as in Baden et al. 2021).
3 The other reasons for exclusion were: did not receive second dose; received incorrect vaccine; received dose 2 out of window; other major protocol deviations; were living with HIV; adjudicated COVID-19 cases up to Day 29 visit (dose 2 date).
4 Minority includes Blacks or African Americans, Hispanics or Latinos, American Indians or Alaska Natives, Native Hawaiians, and other Pacific Islanders. Non-Minority includes all other races with observed race (Asian, Multiracial, White, Other) and observed ethnicity Not Hispanic or Latino.
5 Participants 18-64 were categorized as "At-Risk" (of severe COVID-19 illness) if they had at least one of the following risk factors: chronic lung disease (e.g., emphysema, chronic bronchitis, idiopathic pulmonary fibrosis, cystic fibrosis, or moderate-to-severe asthma); cardiac disease (e.g., heart failure, congenital coronary artery disease, cardiomyopathies, or pulmonary hypertension); severe obesity (BMI ≥40); diabetes (type 1, type 2, or gestational); liver disease; or HIV infection (same as in Baden et al. 2021).
6 Correlates analyses of Day 29 markers start counting COVID-19 endpoints at 7 days post Day 29 visit, and correlates analyses of Day 57 markers start counting COVID-19 endpoints at 7 days post Day 57 visit.
7 Almost all of the antibody levels for placebo recipients were negative, as expected given restriction to participants without evidence of prior SARS-CoV-2 infection. These antibody data were not included in correlates analyses as they do not contribute value except for veritying low false positive rates of the assays.

**Supplementary Text 1: Additional details on the immunoassays**

**Solid-phase electrochemiluminescence S-binding IgG immunoassay (ECLIA)**

MSD SECTOR® plates were precoated by MSD with SARS-CoV-2 spike (S-2P), receptor binding domain (RBD) protein, Nucleocapsid (N) protein and a Bovine Serum Albumin (BSA) control in each well in a specific spot-designation for each antigen. The assay was performed with a Beckman Coulter Biomek based automation integration platform including the Biotek 405TS Plate Washer. Serum samples were heat-inactivated for 30 minutes at 56°C prior to assay. Plates were blocked for 60 minutes at room temperature (RT) with MSD blocker A solution without shaking. Plates were washed and MSD reference standard (calibrator), QC test sample (pool of COVID-19 convalescent sera) and human serum test samples were added to the precoated wells in duplicates in an 8-point dilution series. Reference standard was added in triplicates. MSD Control sera (low, medium and high) were added undiluted in triplicates as per validated assay format. Additional assay controls might be added in triplicates. Samples were incubated at RT for 4 hours with shaking on a Titramax Plate shaker (Heidolph) at 1500 rpm. SARS-CoV-2 specific antibodies present in the sera or controls bound to the coated antigens. Plates were washed to remove unbound antibodies. Antibodies bound to the SARS-CoV-2 viral proteins were detected using an MSD SULFO-TAG™ anti-human IgG detection antibody incubated for 60 minutes at RT and with shaking. Plates were washed and a read solution (MSD GOLD™ read buffer) containing electrochemiluminescence (ECL) substrate was applied to the wells, and the plate was entered into the MSD MESO Sector S 600 detection system. An electric current was applied to the plates and areas of well surface which form antigen-anti human IgG antibody SULFO-TAG™ complex emitted light in the presence of the ECL substrate.

The MSD MESO Sector S 600 detection system quantitates the amount of light emitted and reports the ECL unit response as a result for each test sample, control sample and reference standard of each plate. Analysis was performed with the MSD Discovery Workbench software, Version 4.0. Calculated ECLIA parameters to measure binding antibody activities include interpolated concentrations or assigned arbitrary units (AU/mL) read from the standard curve. Recently the arbitrary units were bridged to the WHO International Standard and a conversion factor was calculated and confirmed. Parallelism was established for all three antigens between the MSD provided reference standard and the WHO provided international standard. Concentration assignments were performed and then confirmed both at MSD and as part of a multi-site confirmation study. Sample results reported here have been converted to international units (IU/mL).

**Neutralization assay**

The neutralization assay was performed as detailed in Shen et al.[1] Briefly, spike-pseudotyped virus was prepared by transfection in 293T cells using a lentivirus backbone vector, a spike-expression plasmid, a TMPRSS2 expression plasmid and a firefly Luc reporter plasmid. A pre-titrated dose of pseudovirus was incubated with eight serial 5-fold dilutions of serum samples (1:10 start dilution) in duplicate in 96-well flat-bottom poly-L-lysine-coated culture plates for 1 hr at 37° C prior to adding 293T/ACE2 cells. One set of eight wells received cells + virus (virus control) and another set of eight wells received cells only (background control). Luminescence was measured after 66-72 hr of incubation using Promega 1X lysis buffer and Bright-Glo luciferase reagent. Neutralization titers are the inhibitory dilution of serum samples at which relative luminescence units (RLUs) were reduced by either 50% (ID50) or 80% (ID80) compared to virus control wells after subtraction of background RLUs. Serum samples were heat-inactivated for 30 min at 56°C prior to assay.

### Calibrated neutralization titers (cID50 and cID80)

Results from the neutralization assay are reported as calibrated ID50 (cID50) and calibrated ID80 (cID80) titers. The calibration was conducted for the First WHO International Standard for Anti-SARS-CoV-2 Immunoglobulin (20/136) in the SARS-CoV-2 spike-pseudotyped virus neutralization assay in 293T/ACE2 cells. The reagent is intended to be used in part to normalize neutralization titers across multiple SARS-CoV-2 neutralization assays (SOP CFAR02-A0026 "Measuring Neutralizing Antibodies Against SARS-CoV2 Using Pseudotyped Virus and 293T/ACE2 Cells" from the Duke Montefiori lab). The calibration work was performed by the "Neutralizing Antibody Core" Laboratory in the Surgical Oncology Research Facility, under the GCLP oversight of the Quality Assurance for Duke Vaccine Immunogenicity Programs (QADVIP).

*Reagent description.* In December 2020, the WHO released a well-characterized international standard for the purpose of improving comparability of results among different assays in different laboratories and reducing interlaboratory variability of anti-SARS-CoV-2 antibody assays.[2] As described in the user instructions provided by the National Institute for Biological Standards and Controls (NIBSC): "The First WHO International Standard for anti-SARS-CoV-2 immunoglobulin is the freeze-dried equivalent of 0.25 mL of pooled plasma obtained from eleven individuals recovered from SARS-CoV-2 infection. The preparation was evaluated in a WHO International Collaborative study. The intended use of the International Standard is for the calibration and harmonization of serological assays detecting anti-SARS-CoV-2 neutralizing antibodies. The preparation can also be used as an internal reference reagent for the harmonization of binding antibody assays. The preparation has been solvent-detergent treated to minimize the risk of the presence of enveloped viruses".[3]

*Reagent preparation and storage.* The reagent was shipped as a lyophilized powder by the NIBSC on December 23, 2020 and was received at Duke on January 4, 2021. A second shipment of the lyophilized reagent was received from the NIBSC on March 15, 2021. The reagent was stored at -80°C upon arrival. Each vial of lyophilized reagent was reconstituted with 0.25 mL of sterile distilled water (Invitrogen, Cat No. 10977015, Ultra-pure DNase, RNase free, Lot 2186762, Exp 30-Aug-2022) as described in the instruction packet[3] and stored at 4°C until use (no longer than 4 weeks).

*Protocol and assay results.* Neutralization assays were conducted in accordance with SOP CFAR02-A0026 "Measuring Neutralizing Antibodies Against SARS-CoV2 Using Pseudotyped Virus and 293T/ACE2 Cells" using the spike pseudotyped virus CoV-2 VRC7480.D614G.1[CMVΔR8.2]/293T/17. The assays were performed between January 5, 2021 and April 16, 2021 by four operators. All assays used either a 1:20 or 1:30 start dilution and a 5-fold dilution series for a total of 8 dilutions. One vial of the standard was reconstituted on January 5 and assayed once on each of 10 plates in a single setting by a single operator (EY) on January 5, 2021; the plates were read on January 8, 2021 (Exp ID EY18-134 in Table 1). These 10 assay results were described in Duke-02-MVR-COVID0001.2. In response to FDA/CBER recommendations (MF 026862, comments dated March 1, 2021), additional assays were performed over different operators and days to yield more precise estimates of the calibration factors. Additional vials of the reagent were reconstituted on March 20, 2021 and assayed in quadruplicate by two operators on two days. A third operator assayed the reconstituted standard in quadruplicate on 3 days. All assays were set up within 24 days of reconstitution. The WHO assigned an arbitrary unitage of 250 IU/ampoule (1000 IU/mL) for neutralizing activity. For calibration purposes, ID50 and ID80 titers may be converted to IUs by dividing 1000 IU/mL by either the mean, median or geometric mean ID50 and ID80 titer as a dilution factor. Thus, the calibration factor for mean ID50 is 1000 ÷ 4135 = 0.242. The calibration factor for mean ID80 is 1000 ÷ 666 = 1.502. The calibration factor for median ID50 is 1000 ÷ 2422 = 0.413. The calibration factor for median ID80 is 1000 ÷ 489 = 2.045. The calibration factor for geometric mean ID50 is 1000 ÷ 3047 = 0.328. The calibration factor for geometric mean ID80 is 1000 ÷ 567 = 1.764. The calibration factors are summarized in the following table:

| SUMMARY VALUES FOR WHO ANTI-SARS-COV-2 IgG (20/136) | | |
|---|---|---|
| **Virus: SARS-CoV-2 D614G Variant** | | **Calibration Factor** |
| Arithmetic mean ID50 | 4135 | 0.242 |
| Arithmetic mean ID80 | 666 | 1.502 |
| Median ID50 | 2422 | 0.413 |
| Median ID80 | 489 | 2.045 |
| Geometric Mean ID50 | 3047 | 0.328 |
| Geometric Mean ID80 | 567 | 1.764 |

The three types of calibration factors (based on arithmetic mean, median, and geometric mean) were compared head-to-head on validation data from Day 29 and Day 57 ID50 and ID80 values from 30 recipients of the mRNA-1273 vaccine, with the common samples conducted by both the Duke Montefiori neutralization assay used for the COVE trial immune correlates study and the Monogram PhenoSense neutralization assays. The results showed that using arithmetic mean of the WHO IS sample for calibration yielded the highest agreement between the calibrated vaccine responses from the two labs, based on the concordance correlation coefficient (manuscript in preparation). Based on this validation experiment, the arithmetic mean calibration factors were selected for use in reporting results in terms of cID50 and cID80 titers.

**Calibration of ID50 and ID80 titers between the Duke neutralization assay on COVE trial samples and the Monogram PhenoSense neutralization assay performed on AZD12222[4] samples**

Using the WHO First Anti-SARS CoV-2 Immunoglobulin International Standard (20/136): 1000 IU/mL, PhenoSense SARS CoV-2 nAb titers can be converted to IU/ml by multiplying the nAb titer (ID50 or ID80) by the appropriate conversion factor, where conversion factors were estimated both for the D614 pseudovirus vaccine strain and for the D614G pseudovirus mutation strain. For each strain, conversion factors were calculated using the mean, geometric mean and median based on 24 replicate tests of the 20/136 standard (6 replicates per day x 4 days). Based on the validation data noted above the arithmetic mean conversion factor was used. For the D614 strain, the conversion factors for ID50 and ID80 were 0.1428 and 0.4585, respectively. For the D614G strain, the conversion factors for ID50 and ID80 were 0.0653 and 0.2281, respectively. In the main article, these conversion factors were first applied as follows: the ID50 values reported in Table 2 of Feng et al.[4] corresponding to 70% and 90% estimated vaccine efficacy were multiplied by 0.1428, where the D614 strain factor was used because the AZD12222 performed the Monogram PhenoSense pseudovirus neutralization assay on participant samples using the D614 strain. Specifically, the ID50 values 57, 183, 982, 303 reported in Table 2 of Feng et al. were each multiplied by 0.1428 to obtain cID50 values 8, 26, 140, 43, respectively; these values are reported in the Discussion in the main article.

In addition, because the Duke neutralization assay performed on COVE samples used the D614G strain, the main article also reports a sensitivity analysis which assumes that D614G is more sensitive to neutralization by vaccine recipient sera than D614. The multiplicative factor defining more sensitive was taken to be (0.1428/0.0653) = 2.19, the ratio of conversion factors (D614 vs. D614G) calculated in the Monogram study that defined the conversion factors. This factor was applied to each cID50 value from the AZD12222 study, which means that each AZD12222 ID50 value on its original scale was multiplied by 0.1428*2.19 = 0.31. Therefore, in this sensitivity analysis, the ID50 values 57, 183, 982, 303 reported in Table 2 of Feng et al. were each multiplied by 0.31 to obtain the cID50 values 18, 57, 307, 95, respectively.

**Table S1. Meso-Discovery (MSD) and pseudovirus neutralization assay limits of the four antibody markers evaluated as immune correlates.**

| | MSD Binding Assay (VRC) | | | PsV nAb (Duke) | |
|---|---|---|---|---|---|
| Reported units | IU/ml* | | | Calibrated titers** | |
| | Spike | RBD | N | cID50 | cID80 |
| Positivity Cutoff | 10.8424 | 14.0858 | 23.5 | 2.42 | 15.02 |
| LOD | 0.3076 | 1.5936 | 0.09 | 2.42 | 15.02 |
| LLOQ | 1.7968 | 3.43 | 4.49 | 4.477 | 21.4786 |
| ULOQ | 10,155.95 | 16,269 | 575 | 10919 | 15368 |

*AU/ml units converted to IU/ml units for all data analysis.

**Original titers calibrated to the WHO anti-SARS-CoV-2 immunoglobulin International Standard (NIBSC code: 20/136) for all data analyses.[2,3] cID50 = calibrated ID50 titer; cID80 = calibrated ID80 titer.

**Supplementary Text 2: Baseline covariates adjusted for in immune correlates analyses, including the baseline COVID-19 risk score**

In addition to adjusting for the at-risk indicator (a stratification factor used in the COVE trial randomization) and the indicator of membership in community of color, all correlates analyses adjust for a baseline COVID-19 risk score that was developed through machine learning of the baseline SARS-CoV-2 negative per-protocol placebo arm data in the COVE trial. This supplementary text summarizes the COVID-19 risk score, with the Statistical Analysis Plan providing additional details.

**Table S2** lists the input variables that were included in the machine learning to build a model predicting the COVID-19 endpoint, where cases are COVID-19 endpoints starting 7 days post Day 57 visit and non-cases are participants with follow-up beyond 7 days post Day 57 visit and that never registered a COVID-19 endpoint. The risk score is defined as the logit of the predicted COVID-19 outcome probability from the predictive regression model, estimated using the ensemble algorithm superlearner (i.e. stacking), where this logit predicted outcome is scaled to have empirical mean zero and empirical standard deviation one.

**Table S2. Individual baseline variables input into the Superlearner model for predicting occurrence of COVID-19 in baseline SARS-CoV-2 negative per-protocol placebo recipients.[1]**

| Variable Name | Definition | Total missing values |
|---|---|---|
| MinorityInd | Baseline covariate underrepresented minority status (1=minority, 0=non-minority) | 0/14079 (0.0%) |
| EthnicityHispanic | Indicator ethnicity = Hispanic (0 = Non-Hispanic) | 0/14079 (0.0%) |
| EthnicityNotreported | Indicator ethnicity = Not reported (0 = Non-Hispanic) | 0/14079 (0.0%) |
| EthnicityUnknown | Indicator ethnicity = Unknown (0 = Non-Hispanic) | 0/14079 (0.0%) |
| Black | Indicator race = Black (0 = White) | 0/14079 (0.0%) |
| Asian | Indicator race = Asian (0 = White) | 0/14079 (0.0%) |
| NatAmer | Indicator race = American Indian or Alaska Native (0 = White) | 0/14079 (0.0%) |
| PacIsl | Indicator race = Native Hawaiian or Other Pacific Islander (0 = White) | 0/14079 (0.0%) |
| Multiracial | Indicator race = Multiracial (0 = White) | 0/14079 (0.0%) |
| Other | Indicator race = Other (0 = White) | 0/14079 (0.0%) |
| Notreported | Indicator race = Not reported (0 = White) | 0/14079 (0.0%) |
| Unknown | Indicator race = unknown (0 = White) | 0/14079 (0.0%) |
| HighRiskInd | Baseline covariate high risk pre-existing condition (1=yes, 0=no) | 0/14079 (0.0%) |
| Sex | Sex assigned at birth (1=female, 0=male) | 0/14079 (0.0%) |
| Age | Age at enrollment in years, between 18 and 85 | 0/14079 (0.0%) |
| BMI | BMI at enrollment (kg/m^2) | 79/14079 (0.6%) |

[1]The per-protocol group for immune correlates analysis is slightly different than that for the primary vaccine efficacy analysis,[5] due in part to a data cutoff for the primary vaccine efficacy analysis of November 25, 2020[5] vs a data cutoff for the immune correlates analysis of March 26, 2021, and also in part due to the inclusion of participants

with HIV in the per-protocol set for vaccine efficacy analysis[5] vs. the exclusion of participants with HIV from the per-protocol set for the immune correlates analysis

The following details were used in the implementation of superlearner of the baseline SARS-CoV-2 negative per-protocol placebo arm:

- All of the selected learners (i.e., regression methods for classifying whether a participant has a COVID-19 outcome or not) were coded into the SuperLearner R package available on CRAN, and the analysis was done using this R package.
- Each quantitative and ordinal variable was pre-scaled to have empirical mean 0 and standard deviation 1.
- 5-fold cross-validation was used with no more than max(20,floor(np/20)) input variables included in each model, where np is the number of evaluable placebo arm cases.
- High-correlation variable screening was used, not allowing any pair of input variables to have Spearman rank correlation r > 0.9.
- Two levels of cross-validation (CV) were used. The outer level computed CV-AUC over 5-fold cross-validation, and the inner level used 5-fold CV.
- Results for comparing classification accuracy of different models were based on point and 95% confidence interval estimates of cross-validated area under the ROC curve (CV-AUC).[6,7] Results are presented as forest plots of point and 95% confidence interval estimates similar to those used in Figure 3B of Neidich et al.[8] and in Figure 2 of Magaret, Benkeser, and Williamson et al.[9] CV-AUC was estimated using the *vimp* R package[10] available on CRAN.

**Table S3** lists the learning algorithms that were applied to estimate the conditional probability of the COVID-19 outcome based on the input variables listed in **Table S2**. Some of the algorithms are non-data-adaptive type learning algorithms, such as parametric regression models (e.g., generalized linear models [glms]), which are simple and stable. Data-adaptive type algorithms are also included, for increasing flexibility of modeling and reducing the risk of model misspecification: SL.randomForest, SL.gam, SL.polymars, and SL.xgboost. All of the selected learners are coded into the SuperLearner R package.

**Table S3. Learning algorithm-screen combinations (14 in total) used as input to the Superlearner model in baseline negative per-protocol placebo recipients.**

| Learner | Screen* |
|---|---|
| SL.mean | all |
| SL.glm | all |
| | glmnet |
| | univar_logistic_pval |
| | highcor_random |
| SL.glm.interaction | glmnet |
| | univar_logistic_pval |
| | highcor_random |
| SL.glmnet | all |
| SL.gam | glmnet |
| | univar_logistic_pval |
| | highcor_random |
| SL.xgboost | all |
| SL.ranger.imp | all |

*Screen details:
all: includes all variables
glmnet: includes variables with non-zero coefficients in the standard implementation of SL.glmnet that optimizes the lasso tuning parameter via cross-validation
univar_logistic_pval: Wald test 2-sided p-value in a logistic regression model < 0.10
highcor_random: if pairs of quantitative variables with Spearman rank correlation > 0.90, select one of the variables at random

**Table S4** shows the weights that the Superlearner ensemble model applied to each of the individual prediction algorithms in the modeling of the placebo arm.

**Table S4. Weights assigned by Superlearner to each individual learner in the modeling of the placebo arm**

| Learner | Screen | Weight |
|---|---|---|
| SL.gam | screen_highcor_random | 0.369 |
| SL.gam | screen_univariate_logistic_pval | 0.291 |
| SL.ranger.imp | screen_all | 0.165 |
| SL.mean | screen_all | 0.112 |
| SL.xgboost | screen_all | 0.062 |
| SL.glm | screen_all | 0.000 |
| SL.glmnet | screen_all | 0.000 |
| SL.glm | screen_glmnet | 0.000 |
| SL.glm | screen_univariate_logistic_pval | 0.000 |
| SL.glm | screen_highcor_random | 0.000 |
| SL.glm.interaction | screen_glmnet | 0.000 |
| SL.glm.interaction | screen_univariate_logistic_pval | 0.000 |
| SL.glm.interaction | screen_highcor_random | 0.000 |
| SL.gam | screen_glmnet | 0.000 |

To inform about which individual input variables were most important for the risk score, **Table S5** shows the predictors used in the learners that were assigned positive weight by the Superlearner model, with multiple metrics reported that can be used for ranking variable importance. BMI and age were ranked as the two most important variables for predicting COVID-19.

**Table S5. Predictors in learners assigned positive weight by Superlearner in the modeling of the placebo arm.**

| Learner | Screen | Weight | Predictors | Coefficient | Odds Ratio | Importance | Feature | Gain | Cover | Frequency |
|---|---|---|---|---|---|---|---|---|---|---|
| SL.gam | screen_highcor_random | 0.369 | (Intercept) | -3.109 | 0.045 | NA | NA | NA | NA | NA |
| SL.gam | screen_highcor_random | 0.369 | s(Age,2) | -0.280 | 0.756 | NA | NA | NA | NA | NA |
| SL.gam | screen_highcor_random | 0.369 | s(BMI,2) | 0.174 | 1.190 | NA | NA | NA | NA | NA |
| SL.gam | screen_highcor_random | 0.369 | MinorityInd | -0.042 | 0.959 | NA | NA | NA | NA | NA |
| SL.gam | screen_highcor_random | 0.369 | EthnicityHispanic | 0.027 | 1.028 | NA | NA | NA | NA | NA |
| SL.gam | screen_highcor_random | 0.369 | EthnicityNotreported | -0.046 | 0.955 | NA | NA | NA | NA | NA |
| SL.gam | screen_highcor_random | 0.369 | EthnicityUnknown | -0.056 | 0.945 | NA | NA | NA | NA | NA |
| SL.gam | screen_highcor_random | 0.369 | Black | -0.250 | 0.779 | NA | NA | NA | NA | NA |
| SL.gam | screen_highcor_random | 0.369 | Asian | -0.047 | 0.954 | NA | NA | NA | NA | NA |
| SL.gam | screen_highcor_random | 0.369 | NatAmer | -0.059 | 0.943 | NA | NA | NA | NA | NA |
| SL.gam | screen_highcor_random | 0.369 | PacIsl | -0.442 | 0.643 | NA | NA | NA | NA | NA |
| SL.gam | screen_highcor_random | 0.369 | Multiracial | -0.143 | 0.866 | NA | NA | NA | NA | NA |
| SL.gam | screen_highcor_random | 0.369 | Other | 0.018 | 1.018 | NA | NA | NA | NA | NA |
| SL.gam | screen_highcor_random | 0.369 | Notreported | 0.033 | 1.034 | NA | NA | NA | NA | NA |
| SL.gam | screen_highcor_random | 0.369 | Unknown | 0.029 | 1.030 | NA | NA | NA | NA | NA |
| SL.gam | screen_highcor_random | 0.369 | HighRiskInd | 0.007 | 1.007 | NA | NA | NA | NA | NA |
| SL.gam | screen_highcor_random | 0.369 | Sex | 0.054 | 1.056 | NA | NA | NA | NA | NA |
| SL.gam | screen_univariate_logistic_pval | 0.291 | (Intercept) | -3.084 | 0.046 | NA | NA | NA | NA | NA |
| SL.gam | screen_univariate_logistic_pval | 0.291 | s(Age,2) | -0.274 | 0.761 | NA | NA | NA | NA | NA |
| SL.gam | screen_univariate_logistic_pval | 0.291 | s(BMI,2) | 0.183 | 1.201 | NA | NA | NA | NA | NA |
| SL.gam | screen_univariate_logistic_pval | 0.291 | EthnicityHispanic | 0.010 | 1.010 | NA | NA | NA | NA | NA |
| SL.gam | screen_univariate_logistic_pval | 0.291 | Black | -0.268 | 0.765 | NA | NA | NA | NA | NA |
| SL.gam | screen_univariate_logistic_pval | 0.291 | Multiracial | -0.150 | 0.861 | NA | NA | NA | NA | NA |
| SL.ranger.imp | screen_all | 0.165 | MinorityInd | NA | NA | 3.296 | NA | NA | NA | NA |
| SL.ranger.imp | screen_all | 0.165 | EthnicityHispanic | NA | NA | 3.844 | NA | NA | NA | NA |
| SL.ranger.imp | screen_all | 0.165 | EthnicityNotreported | NA | NA | 1.069 | NA | NA | NA | NA |
| SL.ranger.imp | screen_all | 0.165 | EthnicityUnknown | NA | NA | 0.433 | NA | NA | NA | NA |
| SL.ranger.imp | screen_all | 0.165 | Black | NA | NA | 2.608 | NA | NA | NA | NA |
| SL.ranger.imp | screen_all | 0.165 | Asian | NA | NA | 2.318 | NA | NA | NA | NA |
| SL.ranger.imp | screen_all | 0.165 | NatAmer | NA | NA | 0.971 | NA | NA | NA | NA |
| SL.ranger.imp | screen_all | 0.165 | PacIsl | NA | NA | 0.133 | NA | NA | NA | NA |
| SL.ranger.imp | screen_all | 0.165 | Multiracial | NA | NA | 1.394 | NA | NA | NA | NA |
| SL.ranger.imp | screen_all | 0.165 | Other | NA | NA | 2.155 | NA | NA | NA | NA |

| | | | | | | | | | | |
|---|---|---|---|---|---|---|---|---|---|---|
| SL.ranger.imp | screen_all | 0.165 | Notreported | NA | NA | 1.390 | NA | NA | NA | NA |
| SL.ranger.imp | screen_all | 0.165 | Unknown | NA | NA | 1.682 | NA | NA | NA | NA |
| SL.ranger.imp | screen_all | 0.165 | HighRiskInd | NA | NA | 4.380 | NA | NA | NA | NA |
| SL.ranger.imp | screen_all | 0.165 | Sex | NA | NA | 6.329 | NA | NA | NA | NA |
| SL.ranger.imp | screen_all | 0.165 | Age | NA | NA | 50.511 | NA | NA | NA | NA |
| SL.ranger.imp | screen_all | 0.165 | BMI | NA | NA | 109.263 | NA | NA | NA | NA |
| SL.xgboost | screen_all | 0.062 | NA | NA | NA | NA | BMI | 0.664 | 0.796 | 0.75 |
| SL.xgboost | screen_all | 0.062 | NA | NA | NA | NA | Age | 0.238 | 0.153 | 0.17 |
| SL.xgboost | screen_all | 0.062 | NA | NA | NA | NA | Sex | 0.025 | 0.006 | 0.01 |
| SL.xgboost | screen_all | 0.062 | NA | NA | NA | NA | Black | 0.022 | 0.018 | 0.01 |
| SL.xgboost | screen_all | 0.062 | NA | NA | NA | NA | MinorityInd | 0.018 | 0.009 | 0.01 |
| SL.xgboost | screen_all | 0.062 | NA | NA | NA | NA | HighRiskInd | 0.014 | 0.006 | 0.01 |
| SL.xgboost | screen_all | 0.062 | NA | NA | NA | NA | EthnicityHispanic | 0.011 | 0.004 | 0.00 |
| SL.xgboost | screen_all | 0.062 | NA | NA | NA | NA | Multiracial | 0.007 | 0.004 | 0.00 |
| SL.xgboost | screen_all | 0.062 | NA | NA | NA | NA | Asian | 0.001 | 0.003 | 0.00 |
| SL.xgboost | screen_all | 0.062 | NA | NA | NA | NA | Other | 0.000 | 0.001 | 0.00 |

**Figure S3** Panel A shows the cross-validated receiver operating characteristic curve for the 2 top-performing learners, Superlearner, and the Discrete Superlearner models classifying COVID-19 outcome status in baseline SARS-CoV-2 negative per-protocol placebo recipients, with predictive performance summarized by cross-validated area under the receiver operating characteristic curve (CV-AUC). The point estimate of CV-AUC for the Superlearner was 0.612 with 95% CI 0.591, 0.633.

**Figure S3** Panel B shows the receiver operating characteristic curve for the Superlearner model built from baseline SARS-CoV-2 negative per-protocol placebo recipients applied to baseline SARS-CoV-2 negative per-protocol vaccine recipients, for which the point estimate of AUC for classifying COVID-19 outcome status was 0.614.

**Figure S3. (A) Cross-validated receiver operating characteristic curve for the 2 top-performing learners, Superlearner, and the Discrete Superlearner models classifying COVID-19 outcome occurrence in baseline SARS-CoV-2 negative per-protocol placebo recipients, with cross-validated area under the curve (CV-AUC) summarizing classification performance. (B) Receiver operating characteristic curve for the Superlearner upon applying the model built from placebo recipients to baseline SARS-CoV-2 negative per-protocol vaccine recipients, with AUC in parentheses summarizing classification performance.**

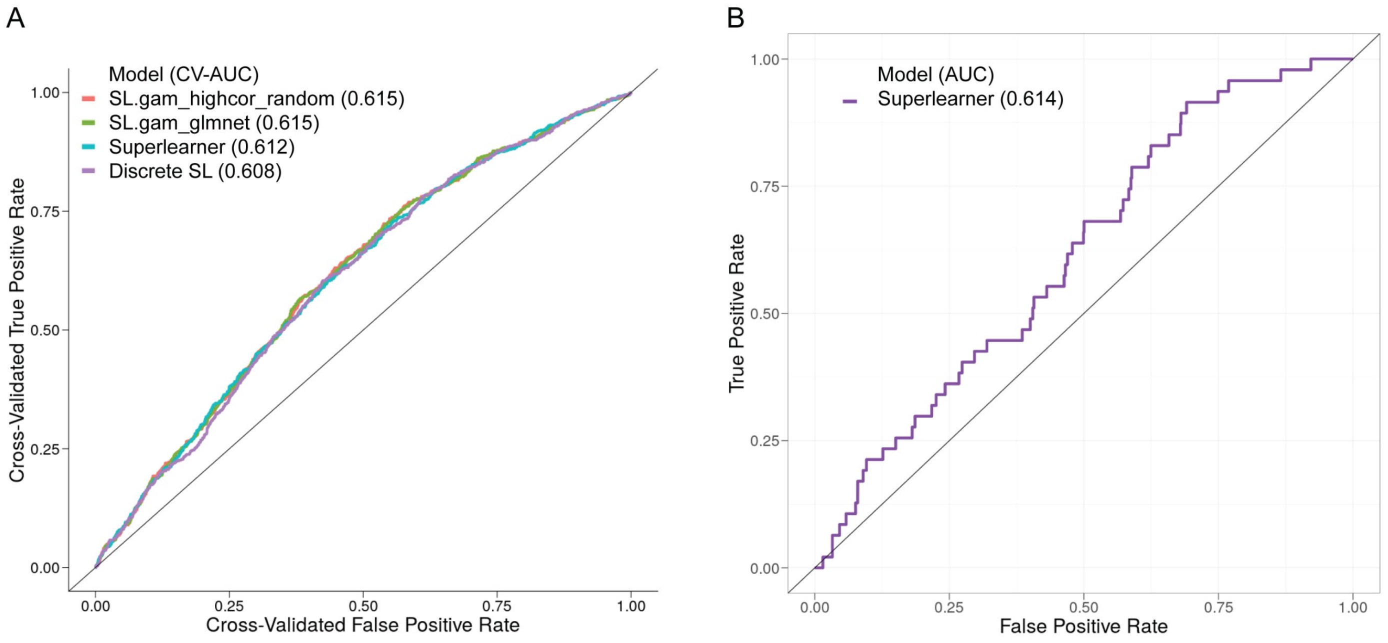

**Figure S4** shows Superlearner model predicted probabilities of the COVID-19 endpoint starting 7 days post Day 57 visit by case vs. non-case status, for baseline SARS-CoV-2 negative per-protocol vaccine recipients in the case-cohort set.

**Figure S4. Superlearner model predicted probabilities of the COVID-19 endpoint starting 7 days post Day 57 visit by case vs. non-case (i.e., control) status, for baseline SARS-CoV-2 negative per-protocol vaccine recipients in the case-cohort set.**

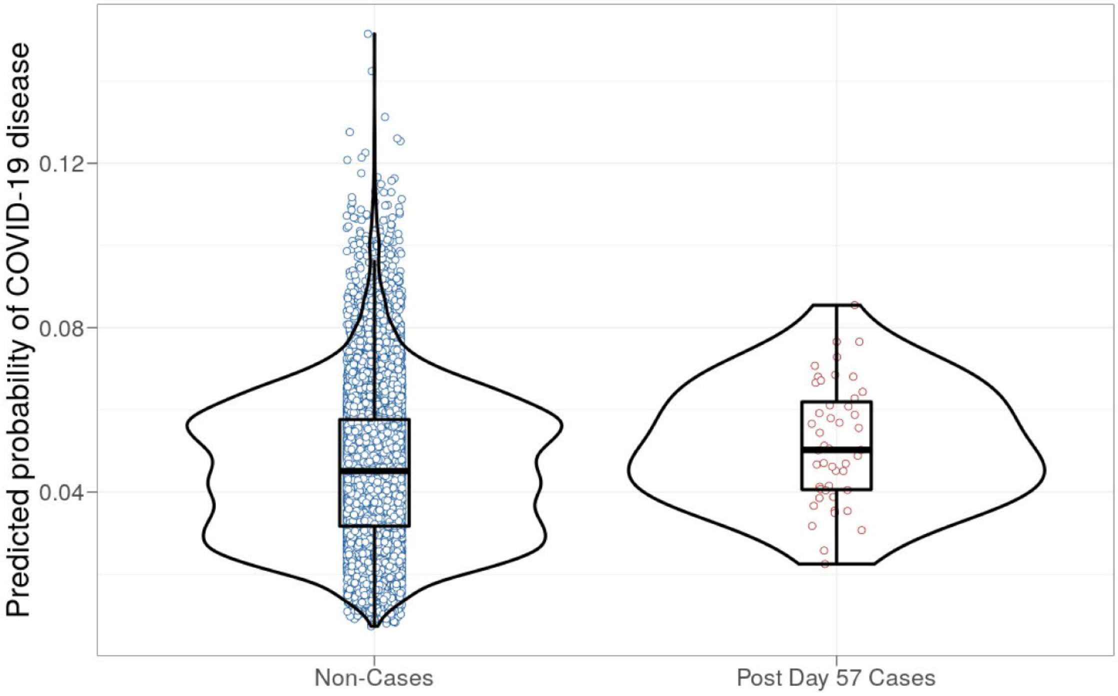

**Table S6. Numbers of participants in the case-cohort set by baseline sampling strata.** Case-cohort set = baseline SARS-CoV-2 negative per-protocol participants sampled into the immunogenicity subcohort or acquired the COVID-19 primary endpoint: all such participants have Day 1, 29, 57 antibody data, except for Day 29 marker correlates analyses intercurrent cases may not have Day 57 antibody data and Day 57 antibody data are not used.

| | Baseline Sampling Strata of SARS-CoV-2 Negative Participants | | | | | | |
|---|---|---|---|---|---|---|---|
| | 1 | 2 | 3 | 4 | 5 | 6 | Total |
| Vaccine | | | | | | | |
| Day 29 Cases | 1 | 6 | 2 | 8 | 9 | 20 | 46 |
| Day 57 Cases | 1 | 4 | 2 | 8 | 5 | 16 | 36 |
| Non-Cases | 171 | 168 | 143 | 146 | 188 | 189 | 1005 |
| Placebo | | | | | | | |
| Day 29 Cases | 9 | 63 | 35 | 69 | 99 | 267 | 542 |
| Day 57 Cases | 8 | 51 | 24 | 61 | 70 | 227 | 441 |
| Non-Cases | 22 | 23 | 20 | 18 | 24 | 26 | 133 |

Demographic covariate strata:

1. Age ≥ 65 Minority
2. Age ≥ 65 Non-Minority
3. Age < 65 At-risk Minority
4. Age < 65 At-risk Non-Minority
5. Age < 65 Not At-risk Minority
6. Age < 65 Not At-risk Non-Minority

Minority includes Blacks or African Americans, Hispanics or Latinos, American Indians or Alaska Natives, Native Hawaiians, and other Pacific Islanders.
Non-Minority includes all other races with observed race (Asian, Multiracial, White, Other) and observed ethnicity Not Hispanic or Latino.
Unknown includes unknown, unreported race or ethnicity.

Cases for Day 29 marker correlates analyses are baseline SARS-CoV-2 negative per-protocol vaccine recipients with the symptomatic infection COVID-19 primary endpoint diagnosed starting 7 days after the Day 29 study visit. Cases for Day 57 marker correlates analyses are baseline SARS-CoV-2 negative per-protocol vaccine recipients with the symptomatic infection COVID-19 primary endpoint diagnosed starting 7 days after the Day 57 study visit. Non-cases are baseline SARS-CoV-2 negative per-protocol participants sampled into the immunogenicity subcohort with no COVID-19 primary endpoint up to the time of data cut and no evidence of SARS-CoV-2 infection up to six days post Day 57 visit.

**Table S7. Demographics and clinical characteristics of baseline SARS-CoV-2 negative per-protocol trial participants in the immunogenicity subcohort and thus have Day 1, 29, 57 antibody marker data.**

| Characteristics | Vaccine (N = 1010) | Placebo (N = 137) | Total (N = 1147) |
|---|---|---|---|
| **Age** | | | |
| Age < 65 | 670 (66.3%) | 91 (66.4%) | 761 (66.3%) |
| Age ≥ 65 | 340 (33.7%) | 46 (33.6%) | 386 (33.7%) |
| Mean (Range) | 54.6 (18.0, 87.0) | 53.4 (19.0, 85.0) | 54.4 (18.0, 87.0) |
| **BMI** | | | |
| Mean ± SD | 30.9 ± 7.6 | 31.3± 9.0 | 30.9± 7.8 |
| **Risk for Severe COVID-19** | | | |
| At-risk | 396 (39.2%) | 57 (41.6%) | 453 (39.5%) |
| Not at-risk | 614 (60.8%) | 80 (58.4%) | 694 (60.5%) |
| **Age, Risk for Severe COVID-19** | | | |
| Age < 65 At-risk | 291 (28.8%) | 40 (29.2%) | 331 (28.9%) |
| Age < 65 Not at-risk | 379 (37.5%) | 51 (37.2%) | 430 (37.5%) |
| Age ≥ 65 | 340 (33.7%) | 46 (33.6%) | 386 (33.7%) |
| **Sex Assigned at Birth** | | | |
| Female | 476 (47.1%) | 63 (46.0%) | 539 (47.0%) |
| Male | 534 (52.9%) | 74 (54.0%) | 608 (53.0%) |
| **Hispanic or Latino Ethnicity** | | | |
| Hispanic or Latino | 322 (31.9%) | 44 (32.1%) | 366 (31.9%) |
| Not Hispanic or Latino | 685 (67.8%) | 93 (67.9%) | 778 (67.8%) |
| Not reported and unknown | 3 (0.3%) | 0 (0.0%) | 3 (0.3%) |
| **Race** | | | |
| White | 735 (72.8%) | 99 (72.3%) | 834 (72.7%) |
| Black or African American | 182 (18.0%) | 24 (17.5%) | 206 (18.0%) |
| Asian | 25 (2.5%) | 6 (4.4%) | 31 (2.7%) |
| American Indian or Alaska Native | 17 (1.7%) | 2 (1.5%) | 19 (1.7%) |
| Native Hawaiian or Other Pacific Islander | 5 (0.5%) | 0 (0.0%) | 5 (0.4%) |
| Multiracial | 12 (1.2%) | 4 (2.9%) | 16 (1.4%) |
| Other | 25 (2.5%) | 2 (1.5%) | 27 (2.4%) |
| Not reported and unknown | 9 (0.9%) | 0 (0.0%) | 9 (0.8%) |
| White Non-Hispanic | 468 (46.3%) | 61 (44.5%) | 529 (46.1%) |
| Communities of Color | 542 (53.7%) | 76 (55.5%) | 618 (53.9%) |

This table summarizes the baseline SARS-CoV-2 negative per-protocol immunogenicity subcohort, which was randomly sampled within 12 strata defined by enrollment characteristics: Assigned treatment arm × Baseline SARS-CoV-2 naïve vs. non-naïve status (defined by serostatus and NAAT testing) × Randomization strata (Age < 65 and at-risk, Age < 65 and not at-risk, Age ≥ 65)× Community of color (Yes/No) defined by White Non-Hispanic vs. all others (same as in Baden et al.[5]).

**Figure S5. For each antibody marker, correlations of Day 29 levels with Day 57 levels in baseline SARS-CoV-2 negative per-protocol vaccine recipients in the immunogenicity subcohort.** Corr = baseline variable adjusted Spearman rank correlation.

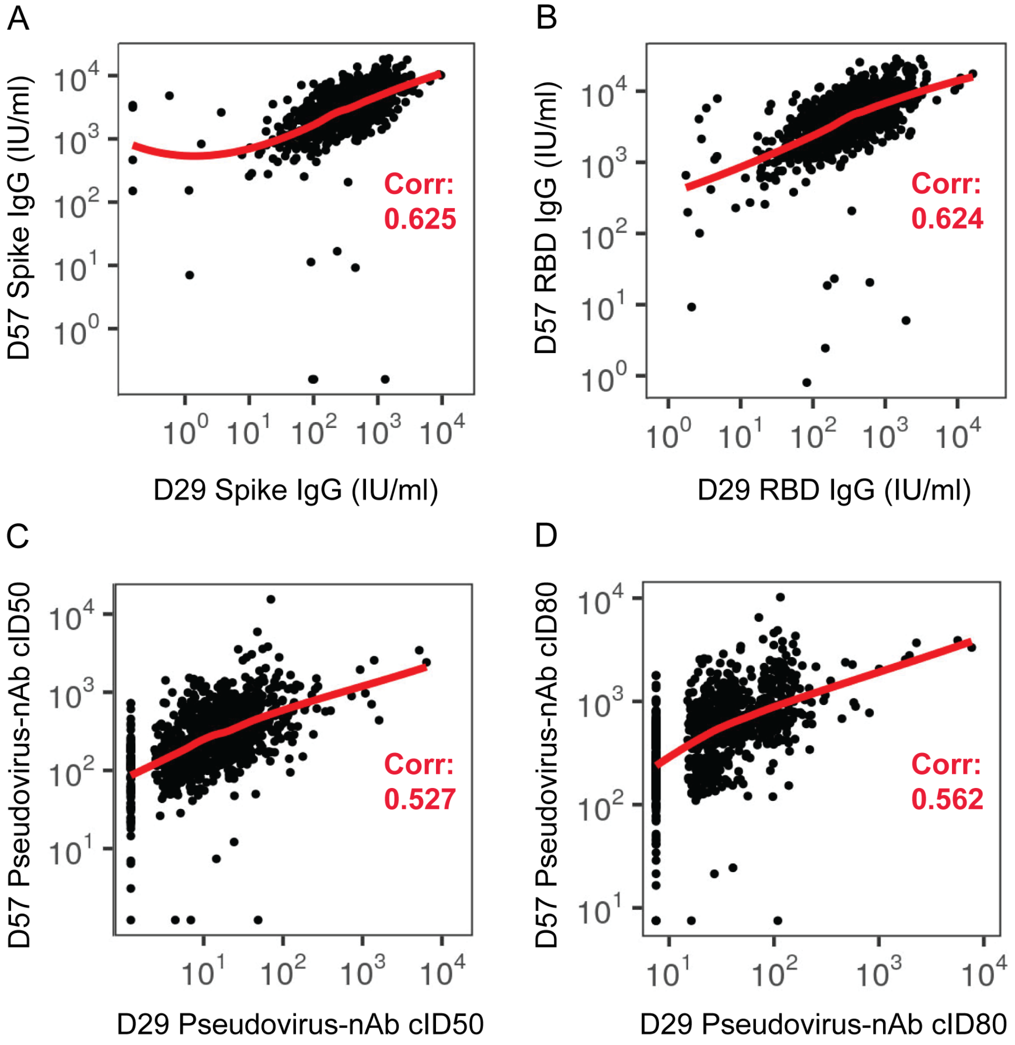

**Figure S6. Correlations of Day 29 antibody markers in baseline SARS-CoV-2 negative per-protocol vaccine recipients in the immunogenicity subcohort.** cID50, cID80: calibrated ID50, ID80 titer. Corr = baseline variable adjusted Spearman rank correlation.

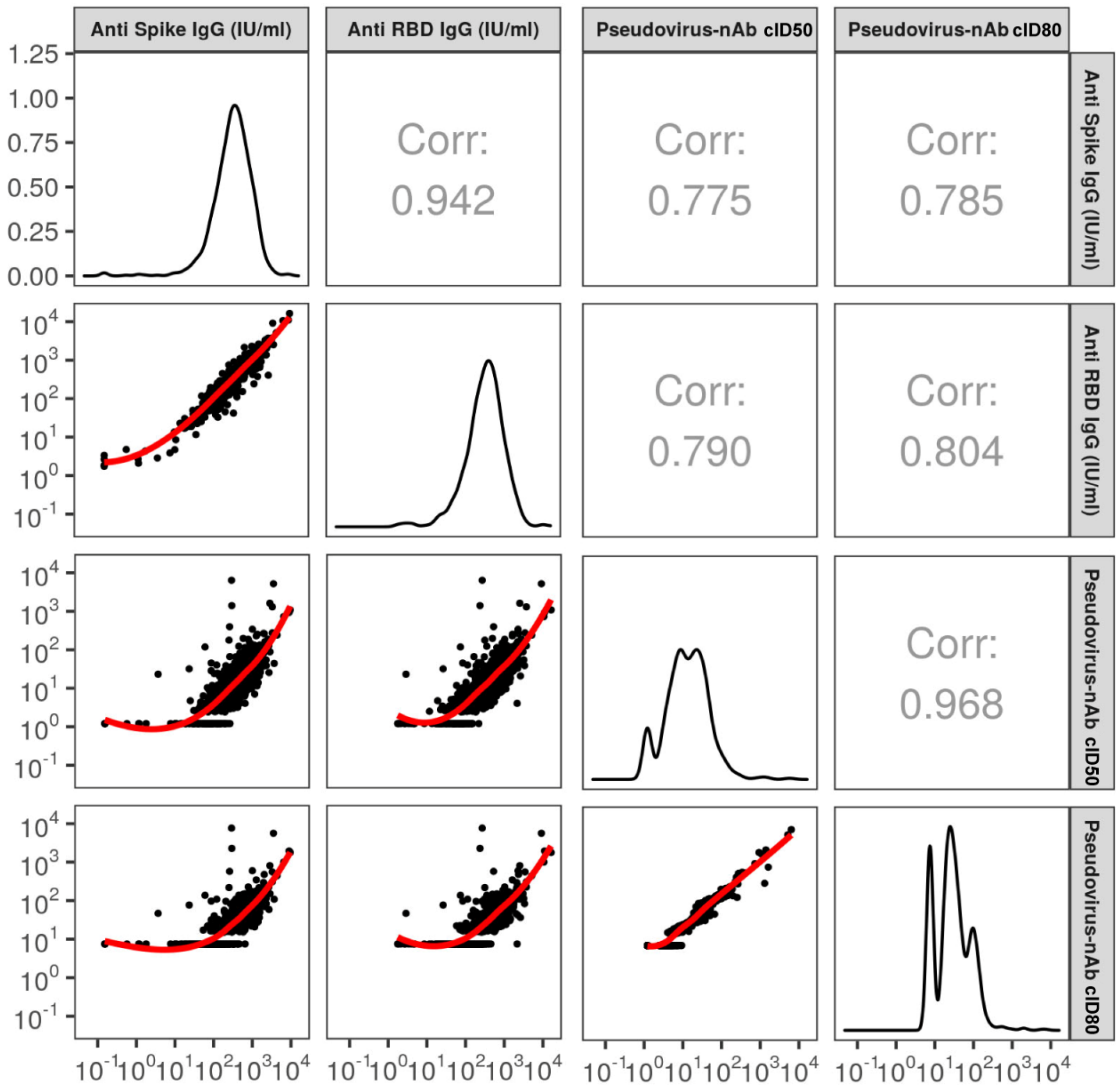

**Figure S7. Correlations of Day 57 antibody markers in baseline SARS-CoV-2 negative per-protocol vaccine recipients in the immunogenicity subcohort.** cID50, cID80: calibrated ID50, ID80 titer. Corr = baseline variable adjusted Spearman rank correlation.

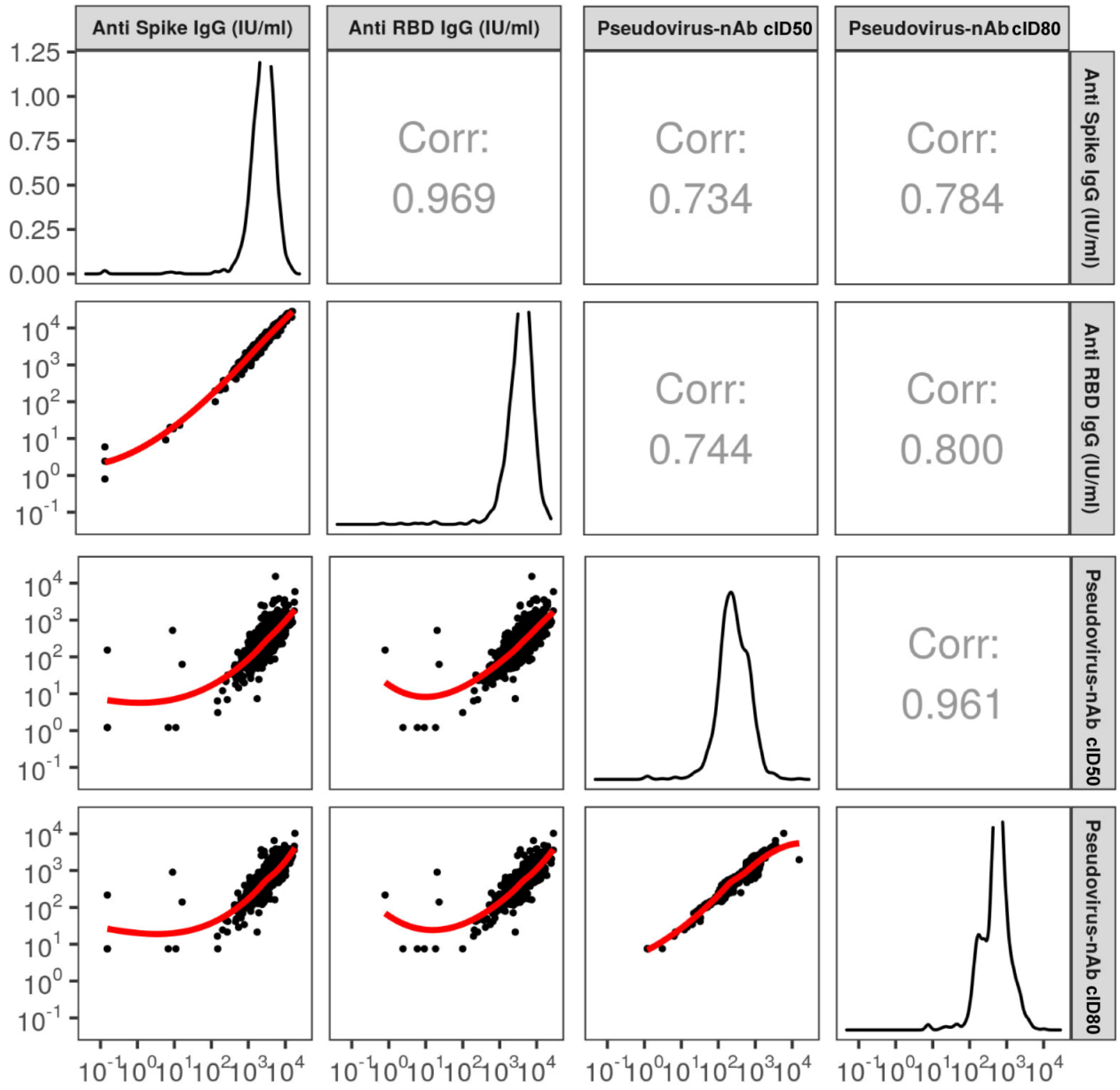

**Figure S8. A) Anti-RBD IgG concentration and B) pseudovirus neutralization cID80 titer by COVID-19 outcome status.** Data points are from baseline SARS-CoV-2 negative per-protocol vaccine recipients selected into the case-cohort set. Pos.Cut, Positivity cut-off. LoD, limit of detection. ULoQ, upper limit of quantitation; ULOQ = 15,368 for cID80 (above all data points). Post Day 57 cases are COVID endpoints starting 7 days post Day 57 visit; Intercurrent cases are COVID endpoints starting 7 days post Day 29 visit through 6 days post Day 57 visit.

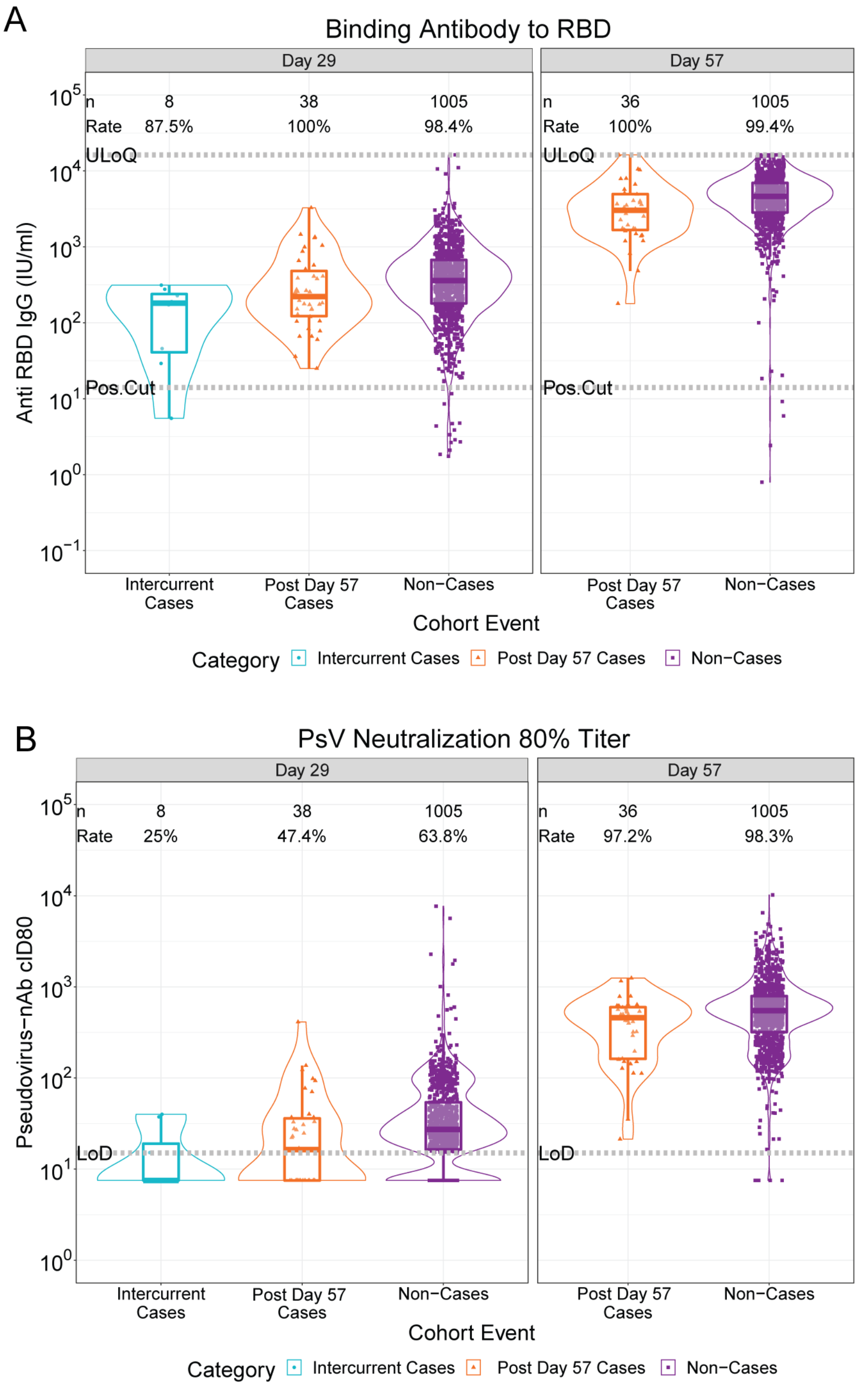

**Figure S9. Marker values (Spike IgG, RBD IgG, cID50, cID80) by COVID-19 outcome status in placebo recipients.** Data points are from baseline SARS-CoV-2 negative per-protocol placebo recipients selected into the case-cohort set. Positive response rates are computed with Inverse Probability Sampling (IPS) weighting. Pos.Cut, Positivity cut-off. LoD, limit of detection. ULoQ, upper limit of quantitation.

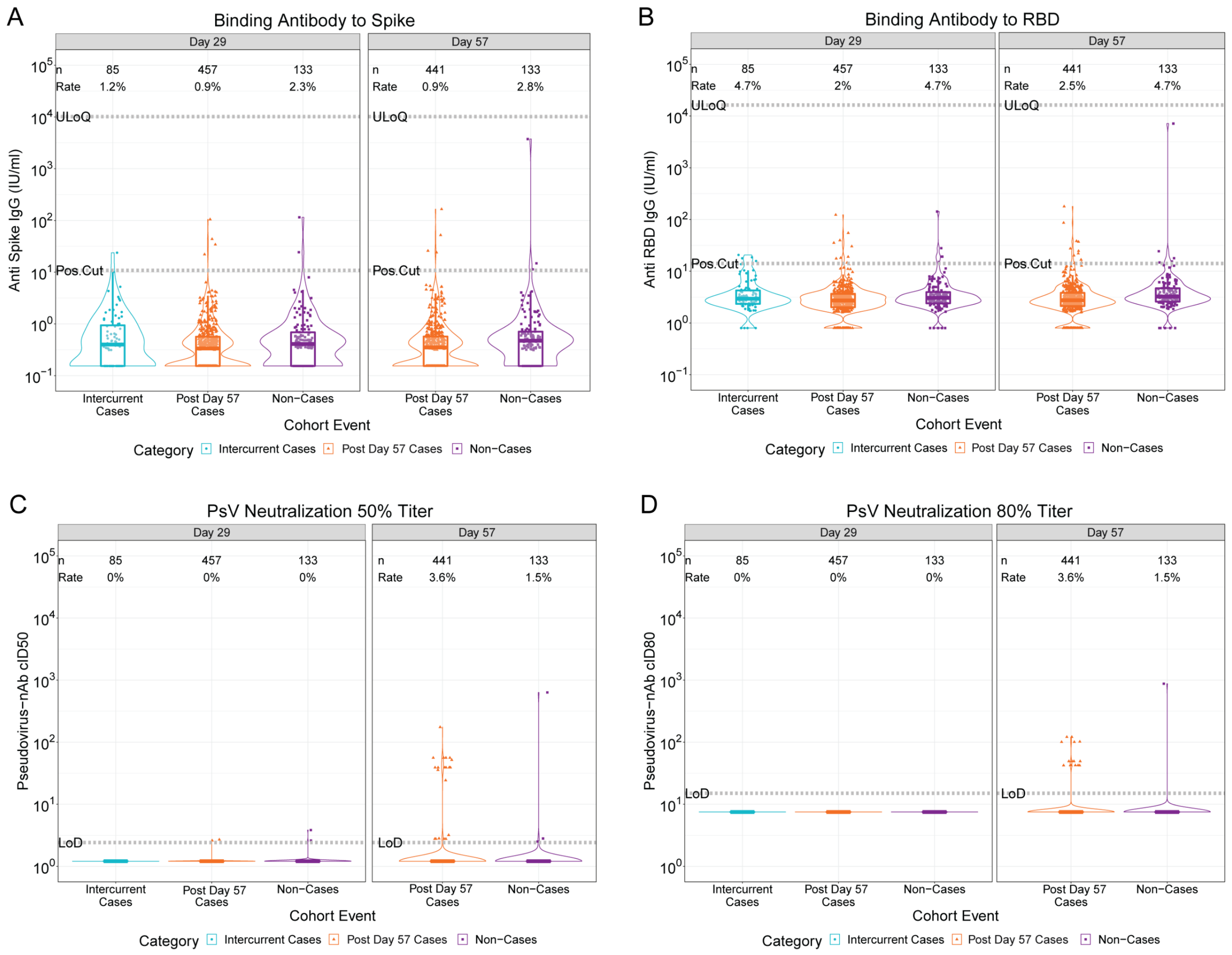

**Figure S10. Day 29 marker data points from baseline SARS-CoV-2 negative per-protocol vaccine recipients selected into the case-cohort set.** Non-cases are represented by orange dots, intercurrent cases by large blue circles, and post Day 57 cases by large purple squares.

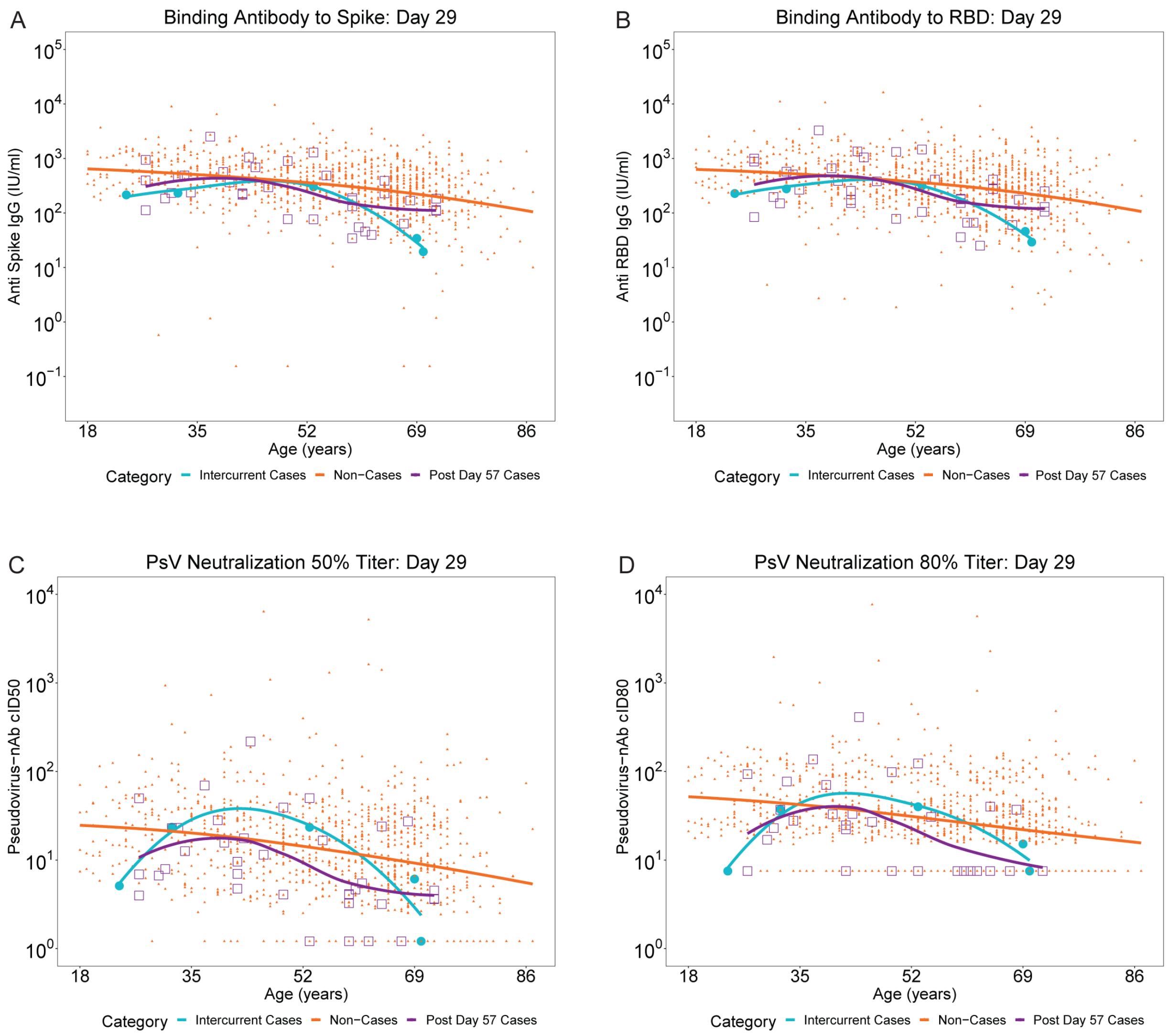

**Figure S11. Day 57 marker data points from baseline SARS-CoV-2 negative per-protocol vaccine recipients selected into the case-cohort set.** Non-cases are represented by orange dots, intercurrent cases by large blue circles, and post Day 57 cases by large purple squares.

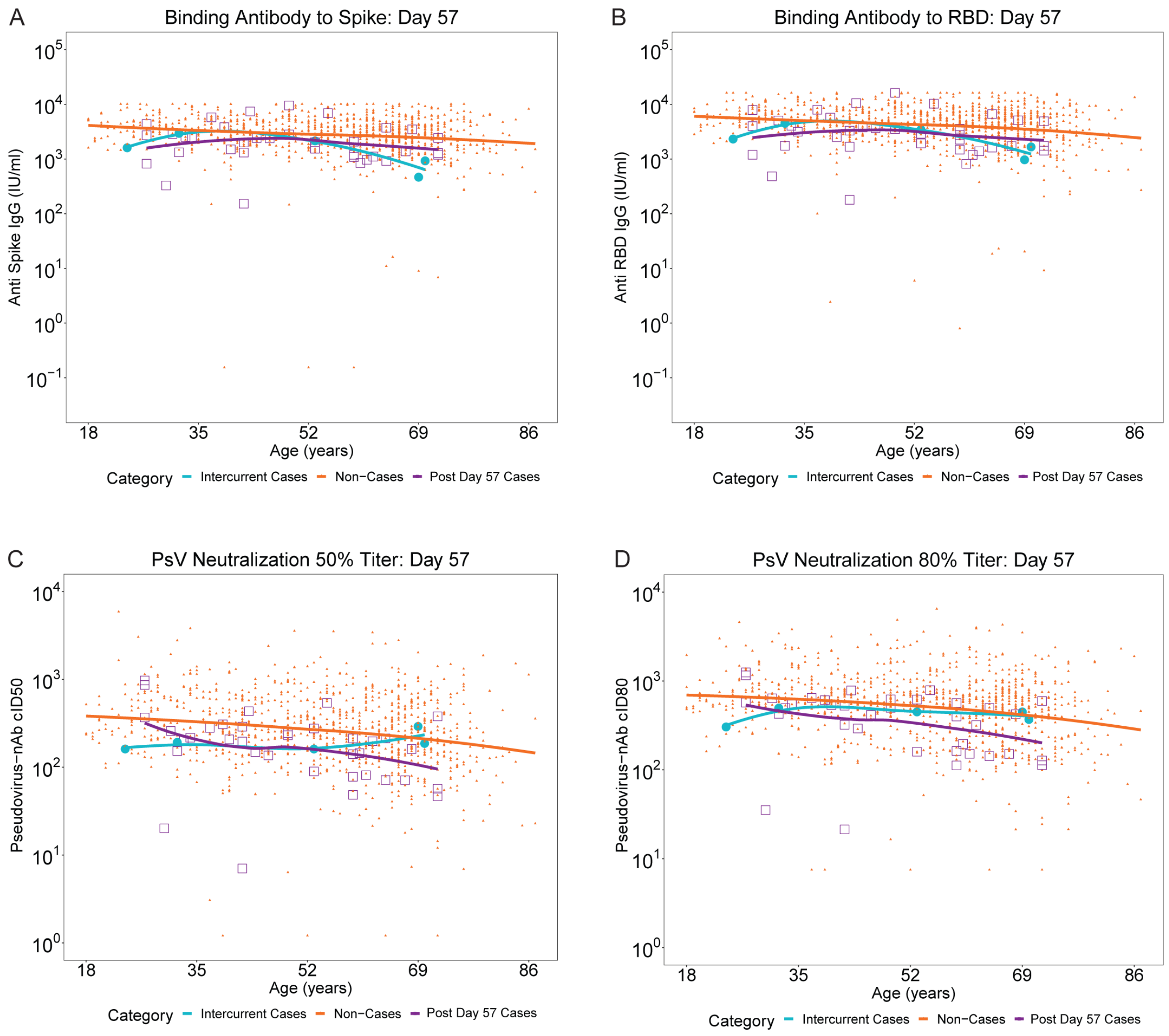

**Figure S12. Inverse probability sampling (IPS)-weighted empirical reverse cumulative distribution function curves for each Day 57 marker (Spike IgG, RBD IgG, cID50, cID80) and application of the Siber (2007) method[11] for estimating a threshold of perfect vs. no protection. cID50, cID80: calibrated ID50, ID80 titer.**

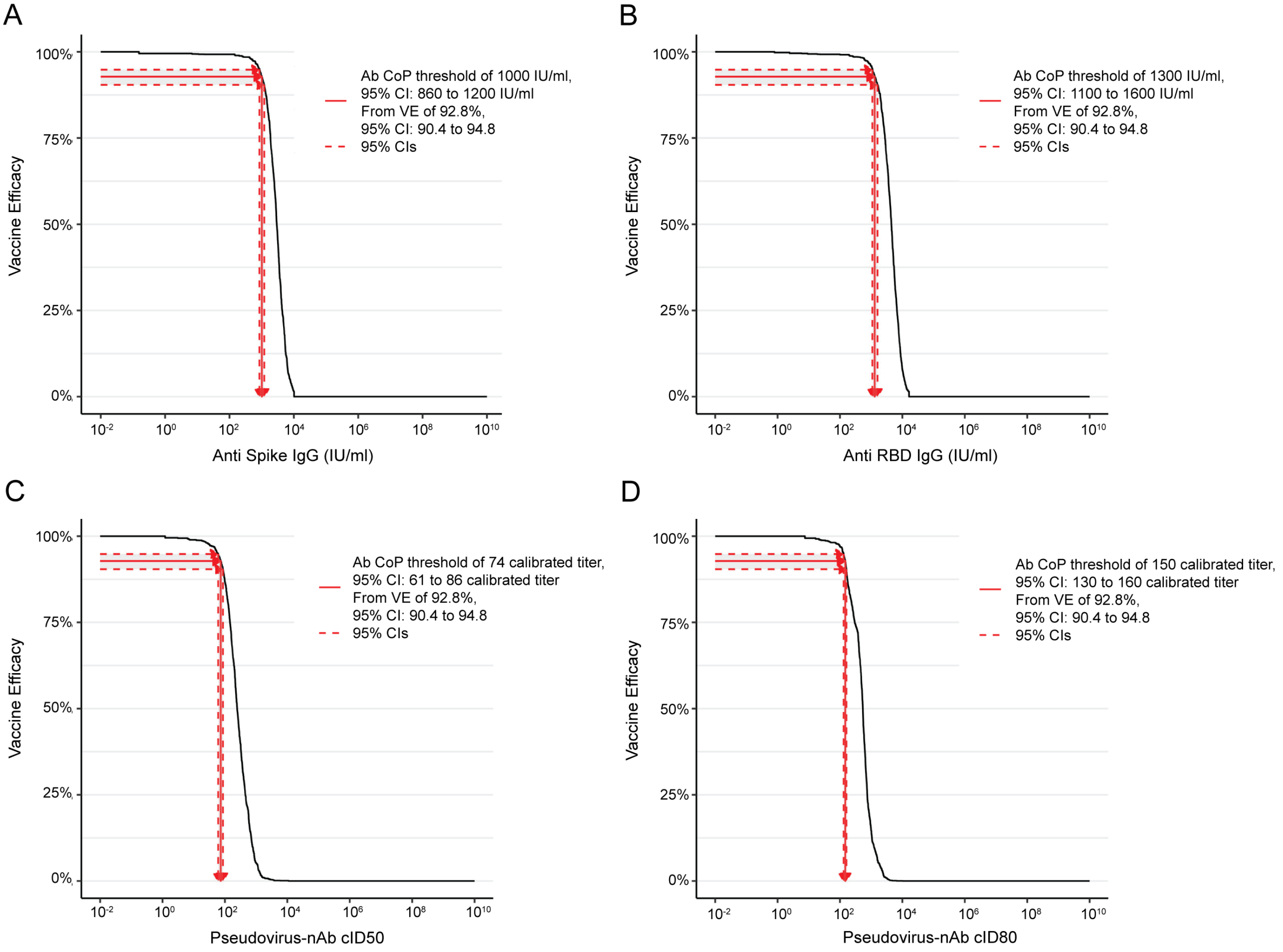

**Figure S13. Inverse probability sampling (IPS)-weighted empirical reverse cumulative distribution function curves for each Day 29 marker (Spike IgG, RBD IgG, cID50, cID80) and application of the Siber (2007) method[11] for estimating a threshold of perfect vs. no protection. cID50, cID80: calibrated ID50, ID80 titer.**

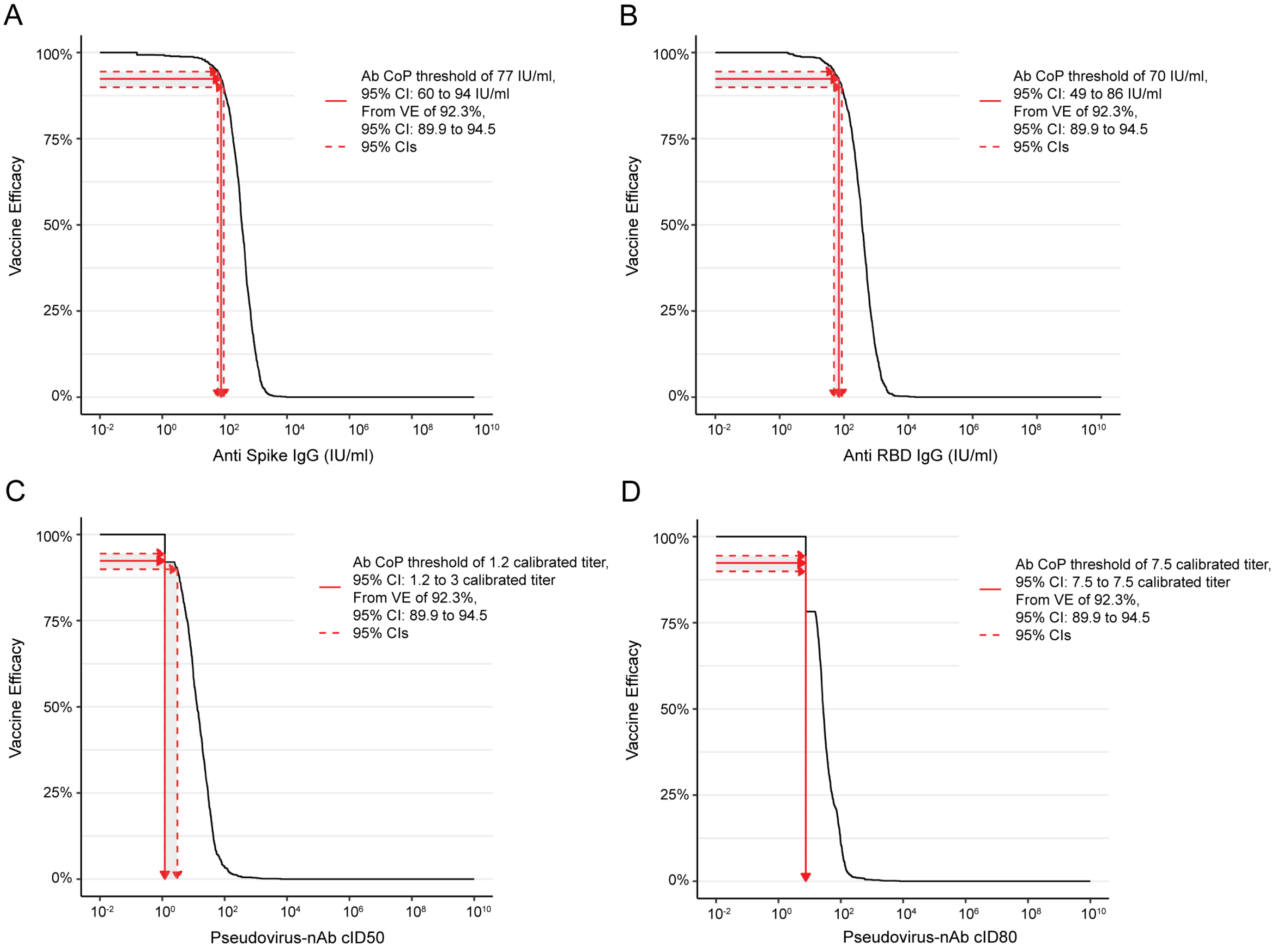

**Figure S14. Day 29 and Day 57 antibody markers (Spike IgG, RBD IgG, cID50, cID80) vs. number of days from Day 29 visit until COVID-19 primary endpoint diagnosis for per-protocol baseline SARS-CoV-2 negative vaccine recipient breakthrough cases.** Blue circles are Day 29 marker values for Intercurrent Cases. Orange triangles are Day 29, Day 57 marker values for Post Day 57 cases, with paired marker values from the same participant joined by a vertical line segment, with Day 57 marker value always the top triangle. cID50, cID80: calibrated ID50, ID80 titer.

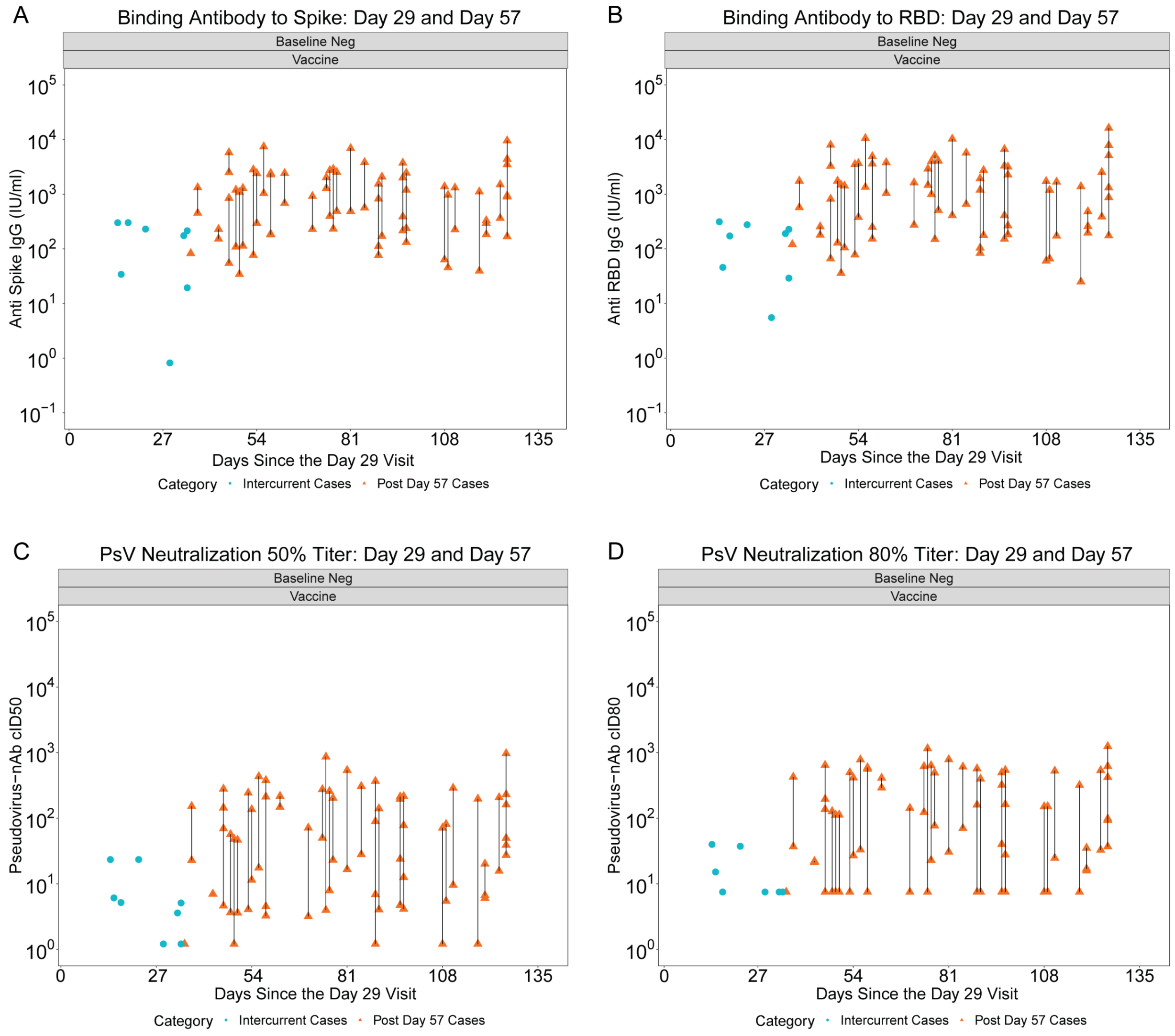

**Figure S15: Covariate-adjusted cumulative incidence of COVID-19 by Low, Medium, High tertile of Day 57 IgG concentration or pseudovirus neutralization titer. (A) Anti-RBD IgG concentration; (B) cID80 titer.**

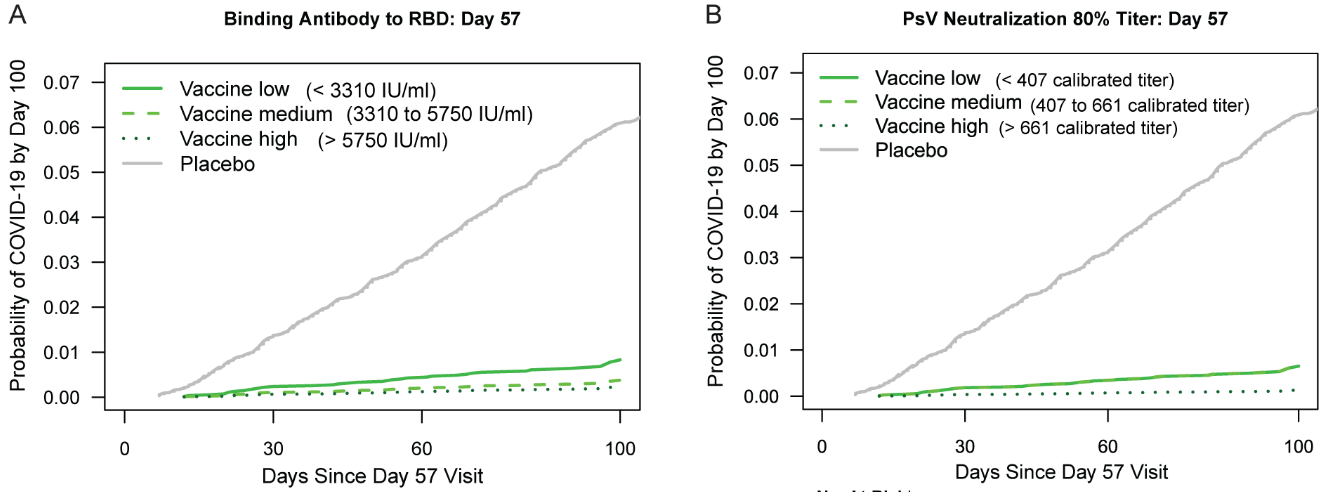

(A)

**No. At-Risk***

| | 0 | 30 | 60 | 100 |
|---|---|---|---|---|
| Low: | 4620 | 4590 | 4245 | 1434 |
| Med: | 4745 | 4696 | 4283 | 1763 |
| High: | 4699 | 4663 | 4344 | 1728 |
| Plac: | 13758 | 13218 | 11165 | 3364 |

**Cumulative No. of COVID-19 Endpoints****

| | 0 | 30 | 60 | 100 |
|---|---|---|---|---|
| Low: | 0 | 8 | 13 | 25 |
| Med: | 0 | 7 | 12 | 13 |
| High: | 0 | 3 | 5 | 9 |
| Plac: | 0 | 187 | 409 | 646 |

(B)

**No. At-Risk***

| | 0 | 30 | 60 | 100 |
|---|---|---|---|---|
| Low: | 4742 | 4717 | 4379 | 1367 |
| Med: | 4715 | 4665 | 4222 | 1555 |
| High: | 4607 | 4567 | 4271 | 2003 |
| Plac: | 13758 | 13218 | 11165 | 3364 |

**Cumulative No. of COVID-19 Endpoints****

| | 0 | 30 | 60 | 100 |
|---|---|---|---|---|
| Low: | 0 | 8 | 12 | 20 |
| Med: | 0 | 8 | 14 | 22 |
| High: | 0 | 1 | 4 | 5 |
| Plac: | 0 | 187 | 409 | 646 |

*No. At-Risk = estimated number in the population for analysis: baseline negative per-protocol vaccine recipients not experiencing the COVID-19 endpoint through 6 days post Day 57 visit.
**Cumulative No. of COVID-19 Endpoints = estimated cumulative number of this cohort with a COVID-19 endpoint.

**Figure S16. Covariate-adjusted cumulative incidence of COVID-19 by Low, Medium, High tertile of Day 29 IgG concentration or pseudovirus neutralization titer.** (A) Anti-Spike IgG concentration; (B) cID50 titer; (C) IgG (Spike, RBD) and (cID50, cID80). The overall p-value is from a generalized Wald test for whether the COVID-19 hazard differed across Low, Medium, and High subgroups.

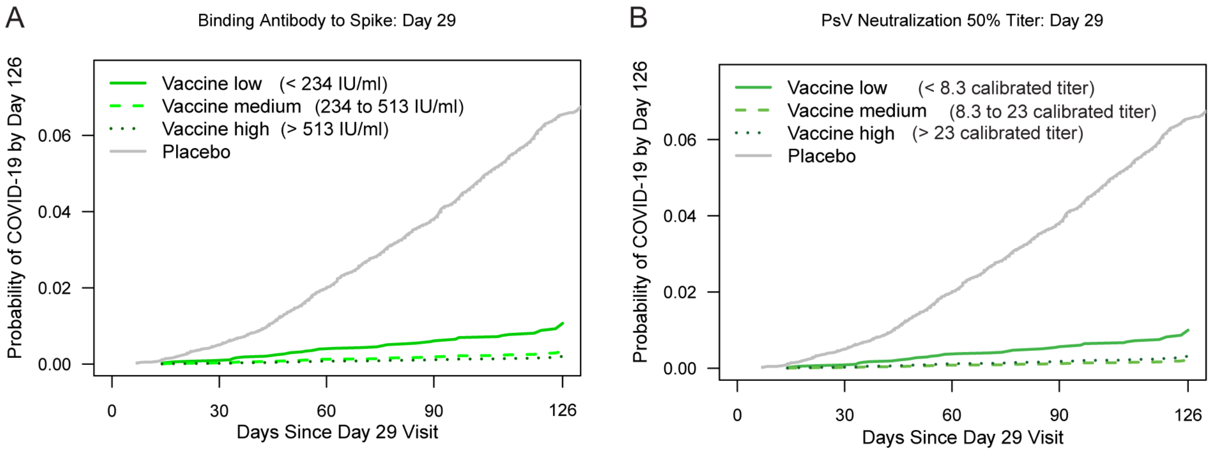

(A)

**No. At-Risk***

| | 0 | 30 | 60 | 90 | 126 |
|---|---|---|---|---|---|
| Low: | 4509 | 4506 | 4462 | 4182 | 1664 |
| Med: | 4897 | 4894 | 4878 | 4434 | 1926 |
| High: | 4735 | 4735 | 4688 | 4207 | 1939 |
| Plac: | 13972 | 13856 | 13250 | 11117 | 3963 |

**Cumulative No. of COVID-19 Endpoints****

| | 0 | 30 | 60 | 90 | 126 |
|---|---|---|---|---|---|
| Low: | 0 | 4 | 18 | 24 | 33 |
| Med: | 0 | 2 | 5 | 8 | 13 |
| High: | 0 | 0 | 2 | 6 | 8 |
| Plac: | 0 | 71 | 278 | 507 | 734 |

(B)

**No. At-Risk***

| | 0 | 30 | 60 | 90 | 126 |
|---|---|---|---|---|---|
| Low: | 4688 | 4684 | 4635 | 4371 | 1687 |
| Med: | 4778 | 4778 | 4737 | 4185 | 1917 |
| High: | 4675 | 4673 | 4656 | 4266 | 1925 |
| Plac: | 13972 | 13856 | 13250 | 11117 | 3963 |

**Cumulative No. of COVID-19 Endpoints****

| | 0 | 30 | 60 | 90 | 126 |
|---|---|---|---|---|---|
| Low: | 0 | 4 | 18 | 25 | 33 |
| Med: | 0 | 0 | 4 | 5 | 8 |
| High: | 0 | 2 | 4 | 8 | 13 |
| Plac: | 0 | 71 | 278 | 507 | 734 |

*No. At-Risk = estimated number in the population for analysis: baseline negative per-protocol vaccine recipients not experiencing the COVID-19 endpoint through 6 days post Day 29 visit.
**Cumulative No. of COVID-19 Endpoints = estimated cumulative number of this cohort with a COVID-19 endpoint.

C

| COVE Immunologic Marker | Tertile¶ | No. cases / No. at-risk§ | Attack rate | Haz. Ratio Pt. Est. | Haz. Ratio 95% CI | P-value (2-sided) | Overall P-value | Overall q-value† | Overall FWER |
|---|---|---|---|---|---|---|---|---|---|
| Anti Spike IgG (IU/ml) | Low | 33/4,509 | 0.0073 | 1 | N/A | N/A | <0.001 | <0.001 | <0.001 |
| | Medium | 13/4,897 | 0.0027 | 0.31 | (0.15,0.65) | 0.002 | | | |
| | High | 8/4,735 | 0.0017 | 0.19 | (0.08,0.44) | <0.001 | | | |
| Anti RBD IgG (IU/ml) | Low | 30/4,559 | 0.0066 | 1 | N/A | N/A | 0.002 | 0.004 | 0.003 |
| | Medium | 14/4,803 | 0.0029 | 0.40 | (0.19,0.84) | 0.016 | | | |
| | High | 11/4,779 | 0.0023 | 0.28 | (0.13,0.60) | 0.001 | | | |
| Pseudovirus-nAb cID50 | Low | 33/4,688 | 0.0070 | 1 | N/A | N/A | <0.001 | 0.001 | 0.001 |
| | Medium | 8/4,778 | 0.0017 | 0.22 | (0.09,0.53) | <0.001 | | | |
| | High | 13/4,675 | 0.0028 | 0.32 | (0.15,0.69) | 0.003 | | | |
| Pseudovirus-nAb cID80 | Low | 31/4,709 | 0.0066 | 1 | N/A | N/A | 0.001 | 0.003 | 0.002 |
| | Medium | 16/4,827 | 0.0033 | 0.44 | (0.21,0.90) | 0.025 | | | |
| | High | 8/4,604 | 0.0017 | 0.22 | (0.09,0.51) | <0.001 | | | |
| Placebo | | 734/13,972 | 0.0525 | | | | | | |

Baseline covariates adjusted for: baseline risk score, at risk or not, community of color or not.
Maximum failure event time 126 days post Day 29 visit.
¶Tertiles:
Spike IgG: Low is < 234 IU/ml, Medium is 234 to 513 IU/ml, High is > 513 IU/ml.
RBD IgG: Low is < 234 IU/ml, Medium is 234 to 537 IU/ml, High is > 537 IU/ml.
ID50: Low is < 8.3, Medium is 8.3 to 23, High is > 23 (all in calibrated titer).
ID80: Low is < 21, Medium is 21 to 40, High is > 40 (all in calibrated titer).
§No. at-risk = estimated number in the population for analysis: baseline negative per-protocol vaccine recipients not experiencing the COVID-19 endpoint through 6 days post Day 29 visit; No. cases = estimated number of this cohort with an observed COVID-19 endpoint.
† q-value and FWER (family-wide error rate) are computed over the set of p-values both for quantitative markers and categorical markers using the Westfall and Young permutation method (10000 replicates).

**Figure S17: Covariate-adjusted cumulative incidence of COVID-19 by Low, Medium, High tertile of Day 29 IgG concentration or pseudovirus neutralization titer. (A) Anti-RBD IgG concentration; (B) cID80 titer.**

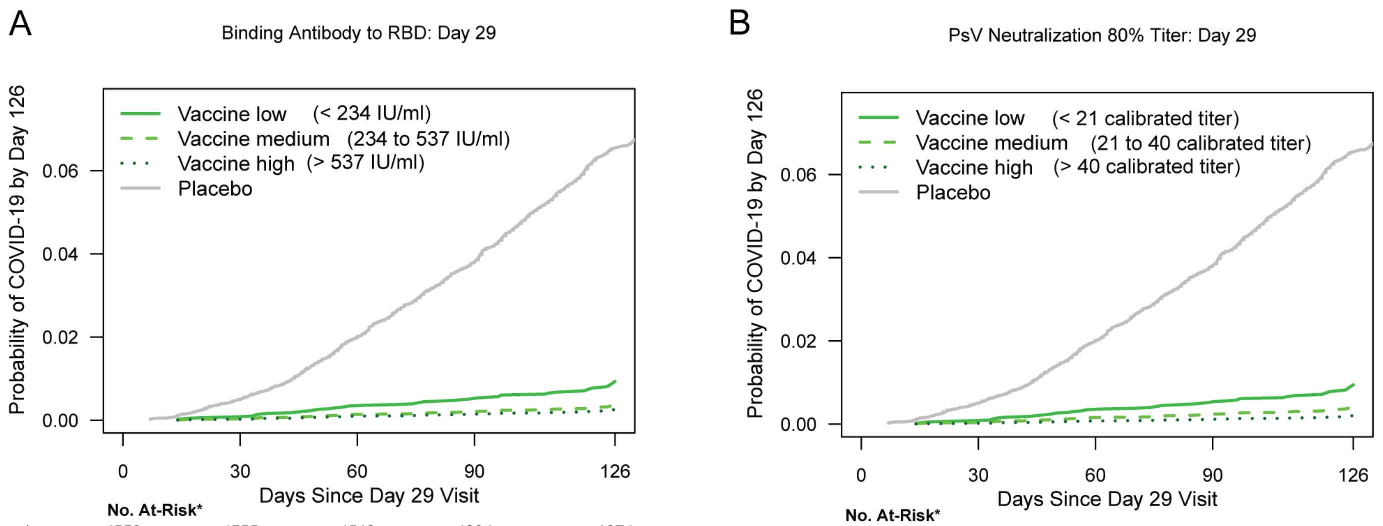

(A) **No. At-Risk***

| | 0 | 30 | 60 | 90 | 126 |
|---|---|---|---|---|---|
| Low: | 4559 | 4555 | 4519 | 4234 | 1674 |
| Med: | 4803 | 4800 | 4783 | 4378 | 1884 |
| High: | 4779 | 4779 | 4726 | 4210 | 1971 |
| Plac: | 13972 | 13856 | 13250 | 11117 | 3963 |

(A) **Cumulative No. of COVID-19 Endpoints****

| | 0 | 30 | 60 | 90 | 126 |
|---|---|---|---|---|---|
| Low: | 0 | 4 | 16 | 20 | 30 |
| Med: | 0 | 2 | 6 | 10 | 14 |
| High: | 0 | 0 | 4 | 8 | 11 |
| Plac: | 0 | 71 | 278 | 507 | 734 |

(B) **No. At-Risk***

| | 0 | 30 | 60 | 90 | 126 |
|---|---|---|---|---|---|
| Low: | 4709 | 4706 | 4658 | 4363 | 1710 |
| Med: | 4827 | 4825 | 4783 | 4242 | 1918 |
| High: | 4604 | 4604 | 4587 | 4218 | 1900 |
| Plac: | 13972 | 13856 | 13250 | 11117 | 3963 |

(B) **Cumulative No. of COVID-19 Endpoints****

| | 0 | 30 | 60 | 90 | 126 |
|---|---|---|---|---|---|
| Low: | 0 | 4 | 17 | 23 | 31 |
| Med: | 0 | 2 | 7 | 10 | 16 |
| High: | 0 | 0 | 1 | 6 | 8 |
| Plac: | 0 | 71 | 278 | 507 | 734 |

*No. At-Risk = estimated number in the population for analysis: baseline negative per-protocol vaccine recipients not experiencing the COVID-19 endpoint through 6 days post Day 29 visit.
**Cumulative No. of COVID-19 Endpoints = estimated cumulative number of this cohort with a COVID-19 endpoint.

**Figure S18. Covariate-adjusted hazard ratios of COVID-19 per 10-fold increase in each Day 29 antibody marker in baseline SARS-CoV-2 negative per-protocol vaccine recipients overall and in subgroups.** (A) Inferences for IgG (Spike, RBD) and (cID50, cID80); (B) Forest plots for anti-Spike IgG concentration; (C) Forest plots for cID50.

A

| COVE Immunologic Marker | No. cases / No. at-risk* | HR per 10-fold incr. Pt. Est. | 95% CI | P-value (2-sided) | q-value ** | FWER |
|---|---|---|---|---|---|---|
| Anti Spike IgG (IU/ml) | 55/14,141 | 0.54 | (0.40,0.74) | <0.001 | <0.001 | <0.001 |
| Anti RBD IgG (IU/ml) | 55/14,141 | 0.46 | (0.30,0.70) | <0.001 | 0.001 | 0.001 |
| Pseudovirus-nAb cID50 | 55/14,141 | 0.33 | (0.17,0.65) | 0.001 | 0.003 | 0.002 |
| Pseudovirus-nAb cID80 | 55/14,141 | 0.19 | (0.07,0.56) | 0.003 | 0.004 | 0.003 |

*No. at-risk = estimated number in the population for analysis: baseline negative per-protocol vaccine recipients not experiencing the COVID-19 endpoint through 6 days post Day 29 visit; No. cases = estimated number of this cohort with an observed COVID-19 endpoint starting 7 days post Day 29 visit.
** q-value and FWER (family-wide error rate) are computed over the set of p-values both for quantitative markers and categorical markers using the Westfall and Young permutation method (10000 replicates).

B

Binding Antibody to Spike: Day 29

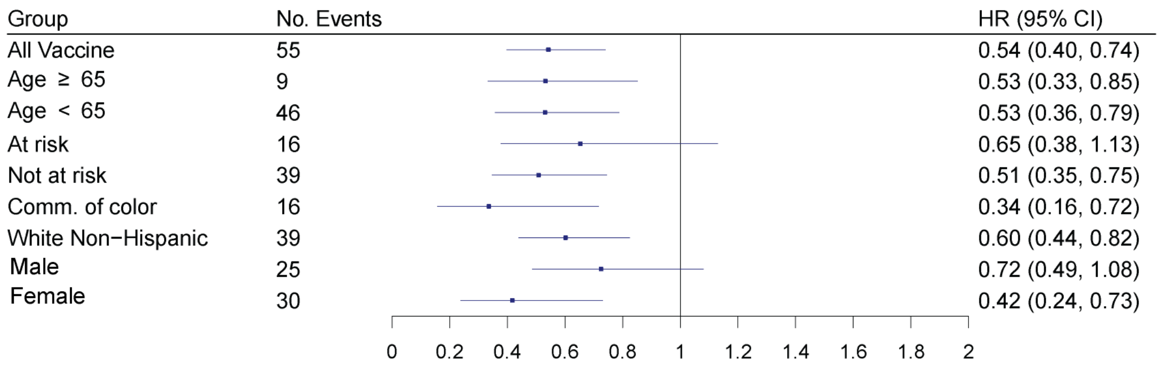

C

PsV Neutralization 50% Titer: Day 29

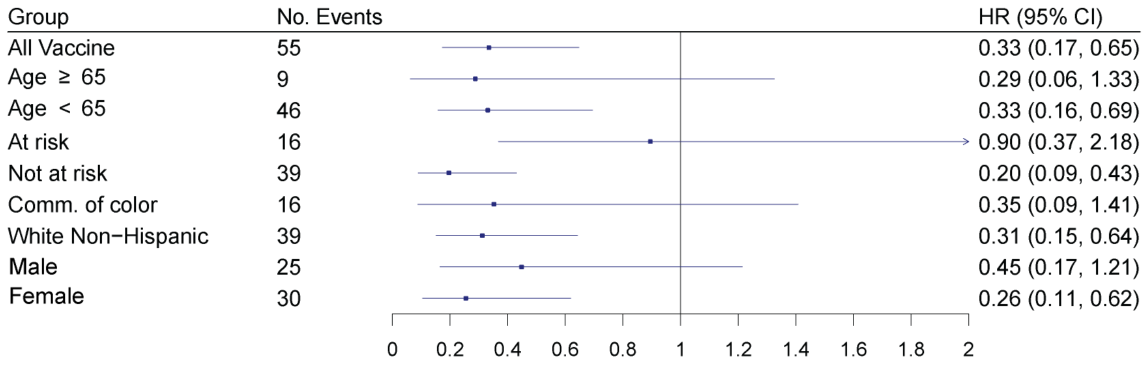

**Figure S19. Covariate-adjusted risk of COVID-19 by the level of each Day 57 marker (Spike IgG, RBD IgG, cID50, cID80), estimated with a generalized additive model.**

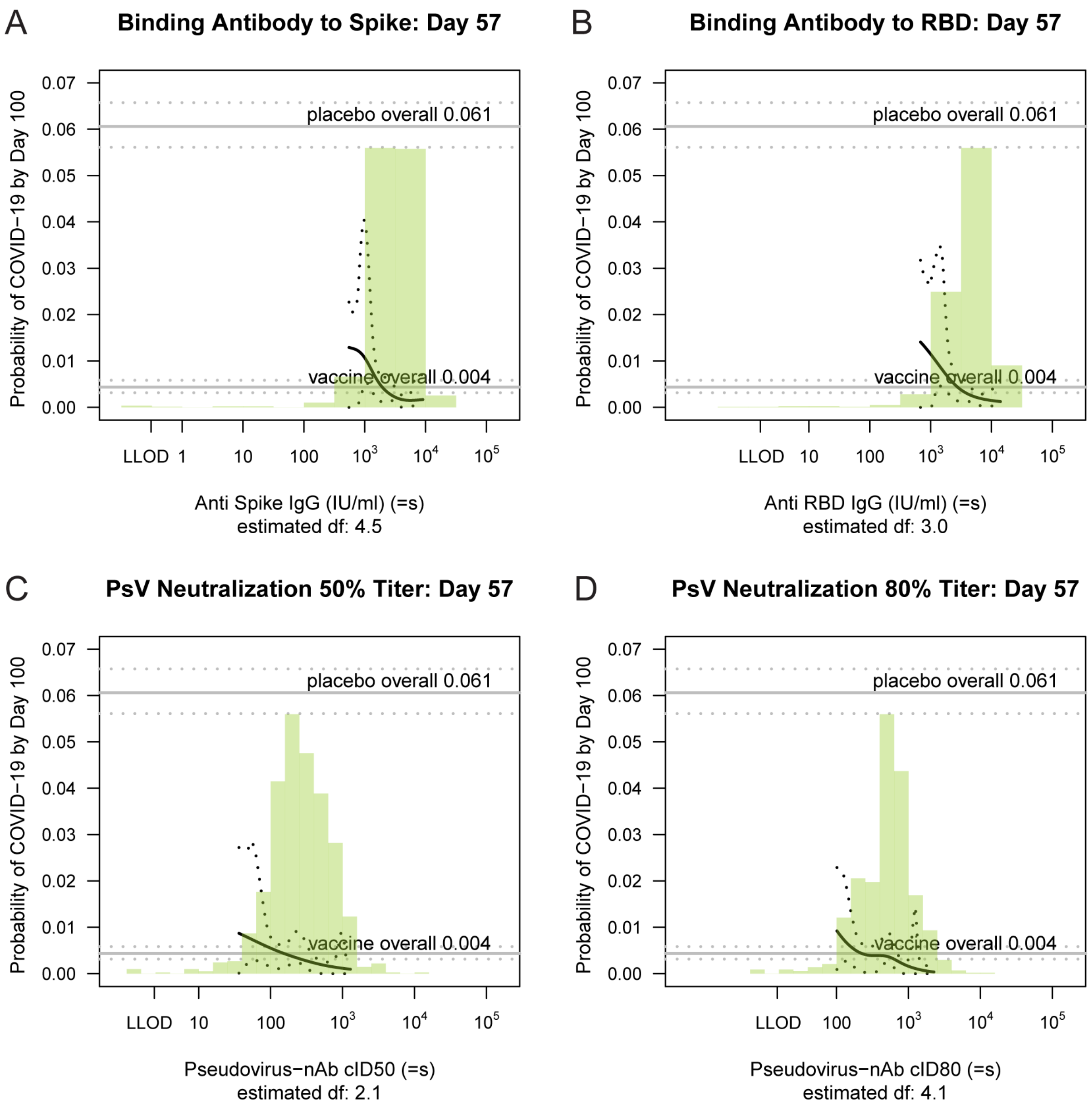

**Figure S20. Covariate-adjusted risk of COVID-19 by the level of each Day 29 marker (Spike IgG, RBD IgG, cID50, cID80), estimated with a generalized additive model.**

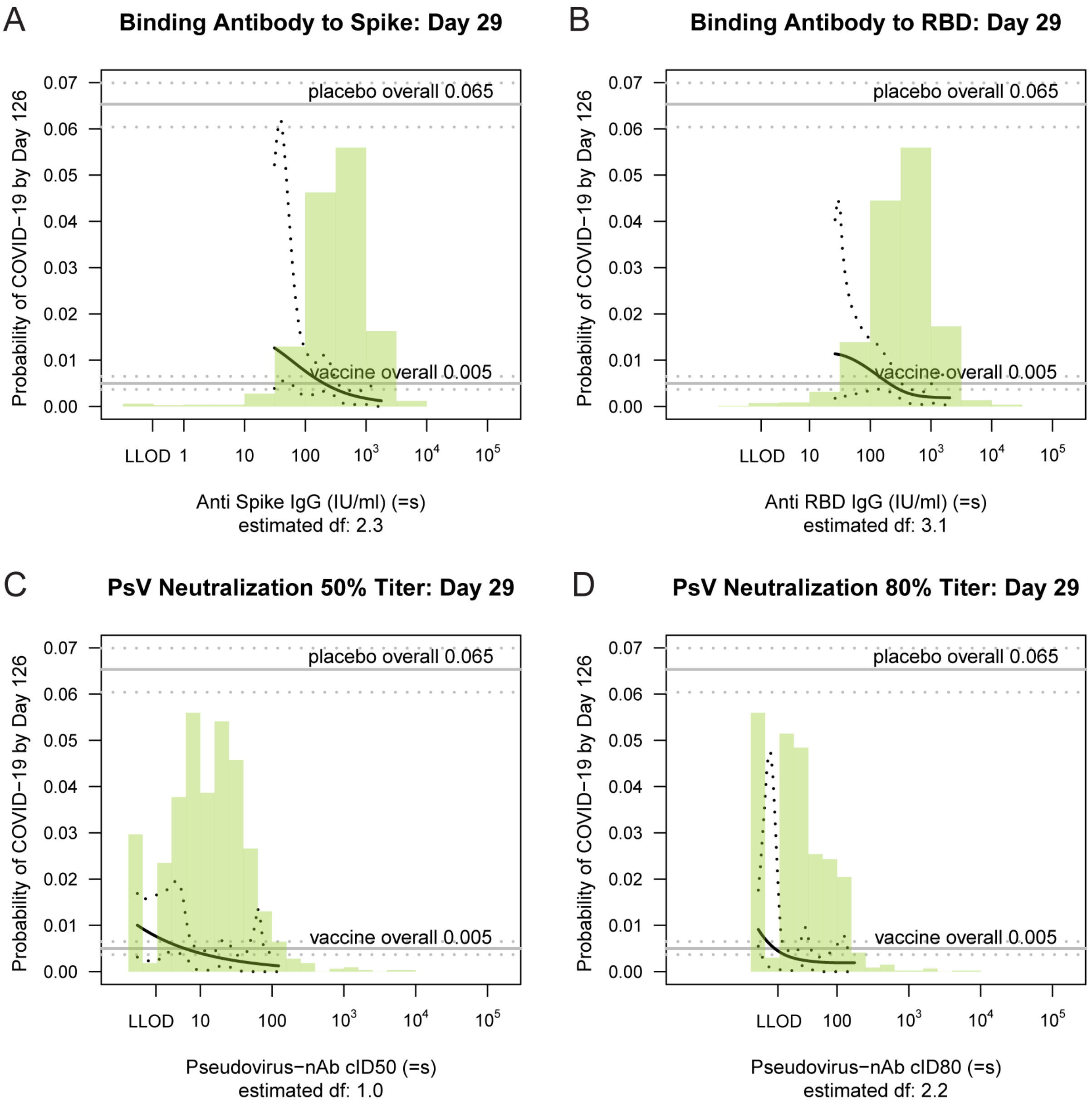

**Figure S21. (A) Covariate-adjusted risk of COVID-19 by subgroups defined by Day 57 cID80 level above a threshold, with reverse cumulative distribution function of Day 57 cID80 level overlaid in green; (B) Covariate-adjusted cumulative incidence of COVID-19 by 100 days post Day 57 by Day 57 cID80 level; (C) Vaccine efficacy by Day 57 cID80 level.** In (A), the gray shaded area is pointwise 95% confidence intervals (CIs). The upper boundary of the green shaded area is the estimate of the reverse cumulative distribution function of the marker in baseline SARS-CoV-2 negative per-protocol vaccine recipients. In (B), the dotted lines indicate bootstrap point-wise 95% CIs. In (C), vaccine efficacy estimates were obtained using the method of Gilbert, Fong, and Carone.[12] In (B) and (C), the green histograms are an estimate of the density of marker level in baseline negative per-protocol vaccine recipients. LOD, limit of detection. cID80: ID80 nAb titer calibrated to the WHO International Standard.

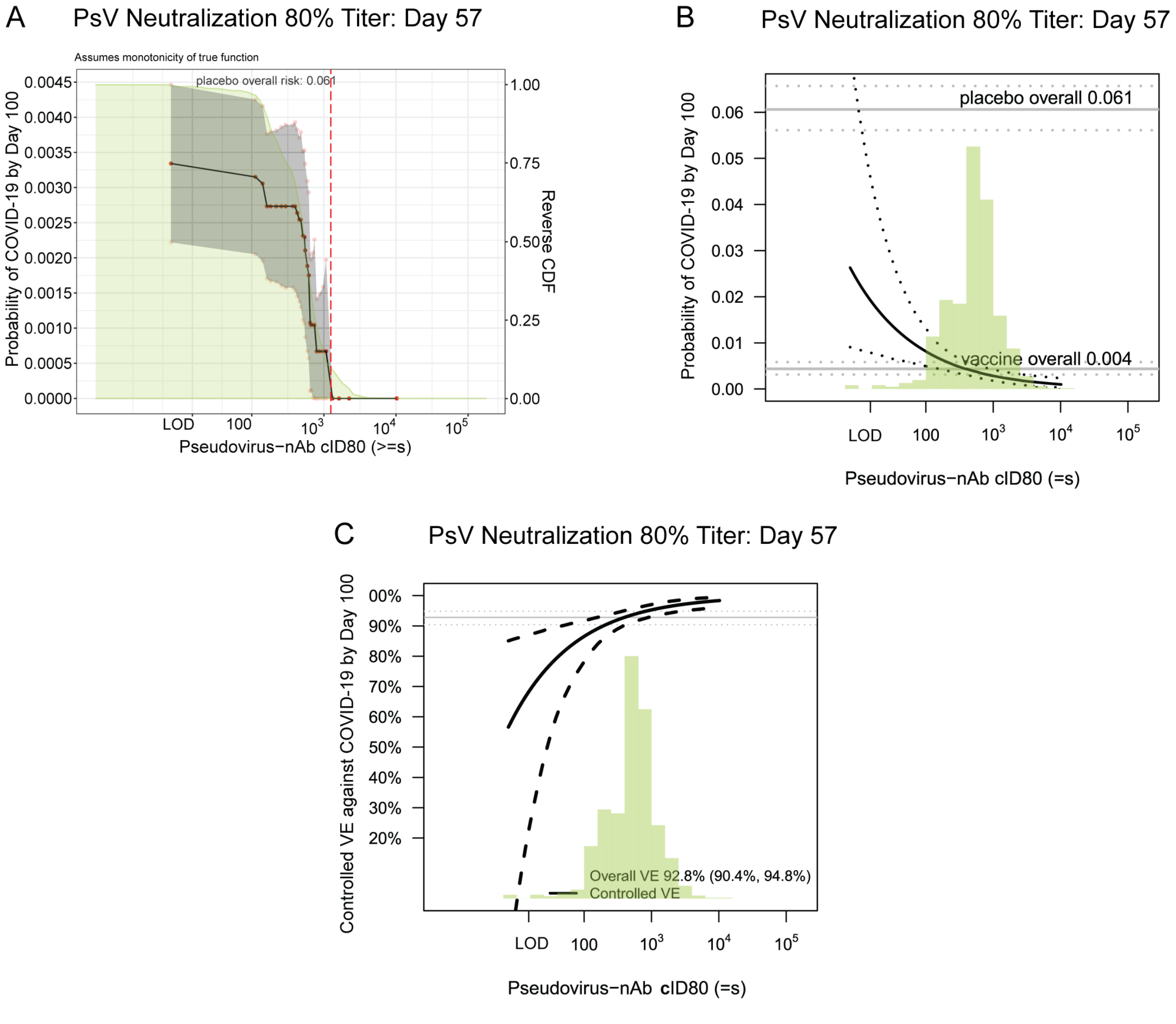

**Figure S22. (A) Covariate-adjusted risk of COVID-19 by subgroups defined by Day 57 Anti-Spike IgG level above a threshold, with reverse cumulative distribution function of Day 57 Anti-Spike IgG level overlaid in green; (B) Covariate-adjusted cumulative incidence of COVID-19 by 100 days post Day 57 by Day 57 Anti-Spike IgG level; (C) Controlled vaccine efficacy by Day 57 Anti-Spike IgG level.** In (A), the gray shaded area is pointwise 95% confidence intervals (CIs). The upper boundary of the green shaded area is the estimate of the reverse cumulative distribution function of the marker in baseline SARS-CoV-2 negative per-protocol vaccine recipients. In (B), the dotted lines indicate bootstrap point-wise 95% CIs. In (C), vaccine efficacy estimates were obtained using the method of Gilbert, Fong, and Carone.[12] In (B) and (C), the green histograms are an estimate of the density of marker level in baseline negative per-protocol vaccine recipients. LOD, limit of detection.

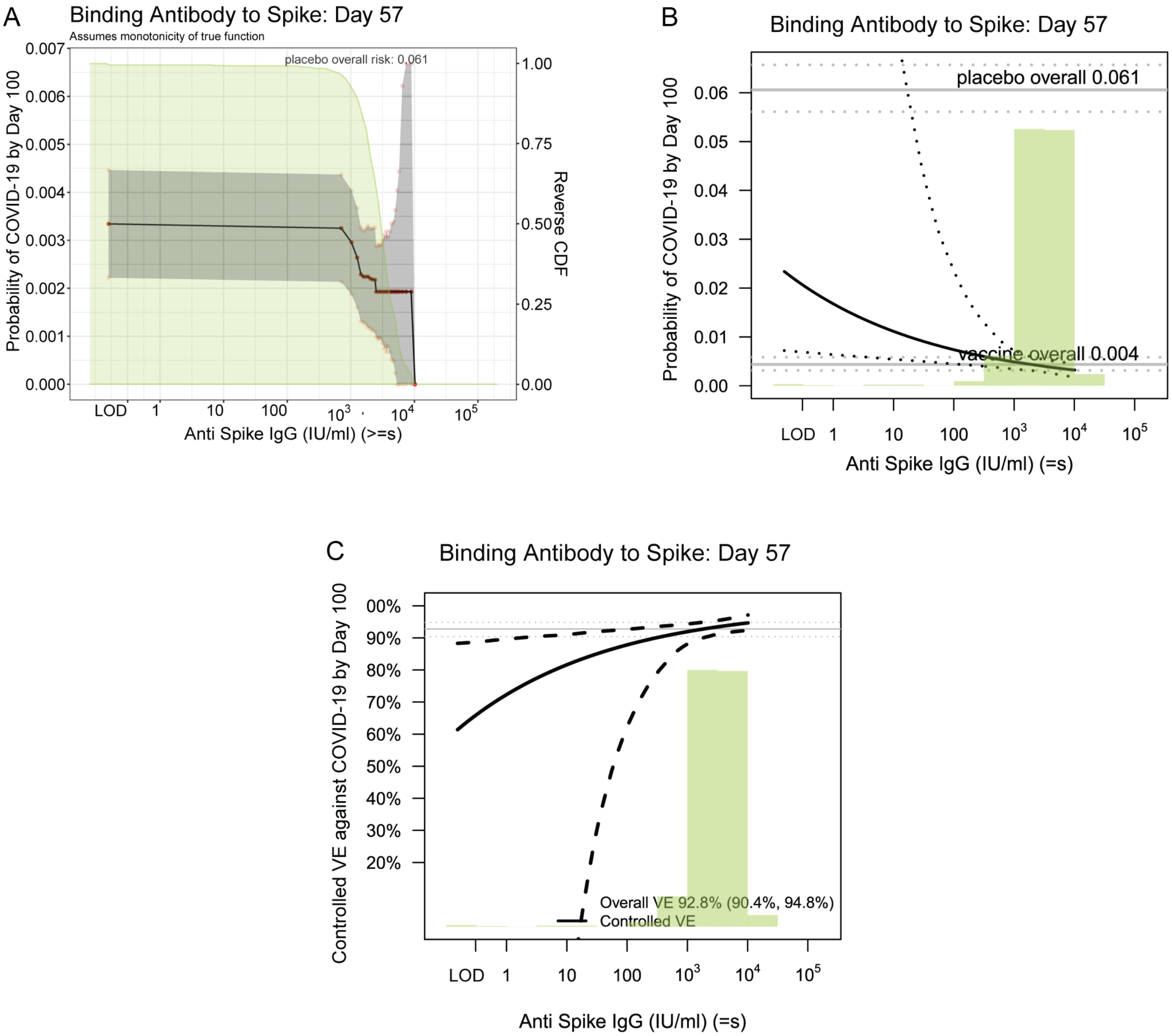

**Figure S23. (A) Covariate-adjusted risk of COVID-19 by subgroups defined by Day 57 Anti-RBD IgG level above a threshold, with reverse cumulative distribution function of Day 57 Anti-RBD IgG level overlaid in green; (B) Covariate-adjusted cumulative incidence of COVID-19 by 100 days post Day 57 by Day 57 Anti-RBD IgG level; (C) Controlled vaccine efficacy by Day 57 Anti-RBD IgG level.** In (A), the gray shaded area is pointwise 95% confidence intervals (CIs). The upper boundary of the green shaded area is the estimate of the reverse cumulative distribution function of the marker in baseline SARS-CoV-2 negative per-protocol vaccine recipients. In (B), the dotted lines indicate bootstrap point-wise 95% CIs. In (C), vaccine efficacy estimates were obtained using the method of Gilbert, Fong, and Carone.[12] In (B) and (C), the green histograms are an estimate of the density of marker level in baseline negative per-protocol vaccine recipients. LOD, limit of detection.

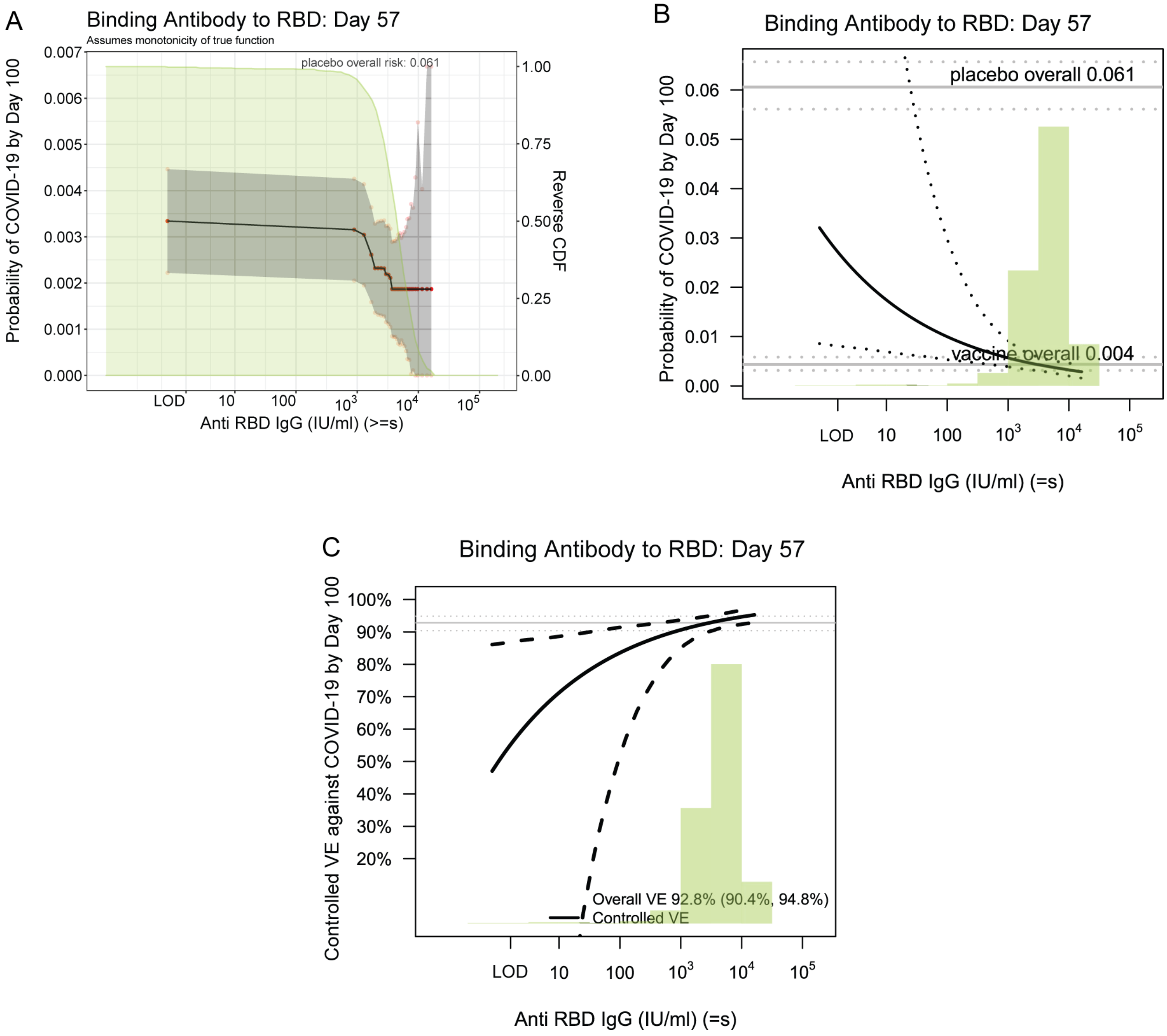

**Figure S24. (A) Covariate-adjusted risk of COVID-19 by subgroups defined by Day 29 cID50 level above a threshold, with reverse cumulative distribution function of Day 29 cID50 level overlaid in green; (B) Covariate-adjusted cumulative incidence of COVID-19 by 126 days post Day 29 by Day 29 cID50 level; (C) Controlled vaccine efficacy by Day 29 cID50 level.** In (A), the gray shaded area is pointwise 95% confidence intervals (CIs). The upper boundary of the green shaded area is the estimate of the reverse cumulative distribution function of the marker in baseline SARS-CoV-2 negative per-protocol vaccine recipients. In (B), the dotted lines indicate bootstrap point-wise 95% CIs. In (C), vaccine efficacy estimates were obtained using the method of Gilbert, Fong, and Carone.[12] In (B) and (C), the green histograms are an estimate of the density of marker level in baseline negative per-protocol vaccine recipients. LOD, limit of detection. cID50: ID50 nAb titer calibrated to the WHO International Standard.

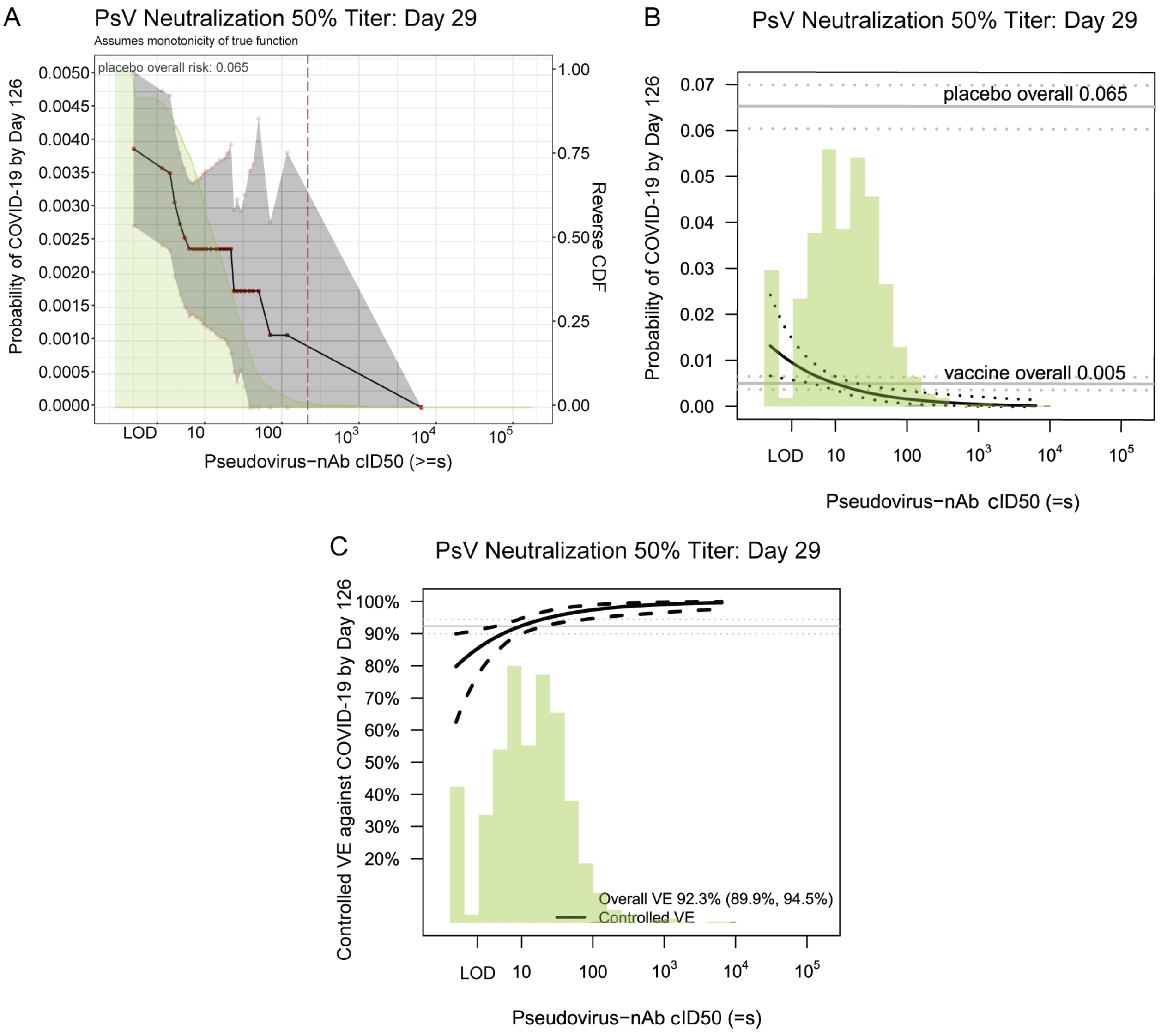

**Figure S25. (A) Covariate-adjusted risk of COVID-19 by subgroups defined by Day 29 cID80 level above a threshold, with reverse cumulative distribution function of Day 29 cID80 level overlaid in green; (B) Covariate-adjusted cumulative incidence of COVID-19 by 126 days post Day 29 by Day 29 cID80 level; (C) Controlled vaccine efficacy by Day 29 cID80 level.** In (A), the gray shaded area is pointwise 95% confidence intervals (CIs). The upper boundary of the green shaded area is the estimate of the reverse cumulative distribution function of the marker in baseline SARS-CoV-2 negative per-protocol vaccine recipients. In (B), the dotted lines indicate bootstrap point-wise 95% CIs. In (C), vaccine efficacy estimates were obtained using the method of Gilbert, Fong, and Carone.[12] In (B) and (C), the green histograms are an estimate of the density of marker level in baseline negative per-protocol vaccine recipients. LOD, limit of detection. cID80: ID80 nAb titer calibrated to the WHO International Standard.

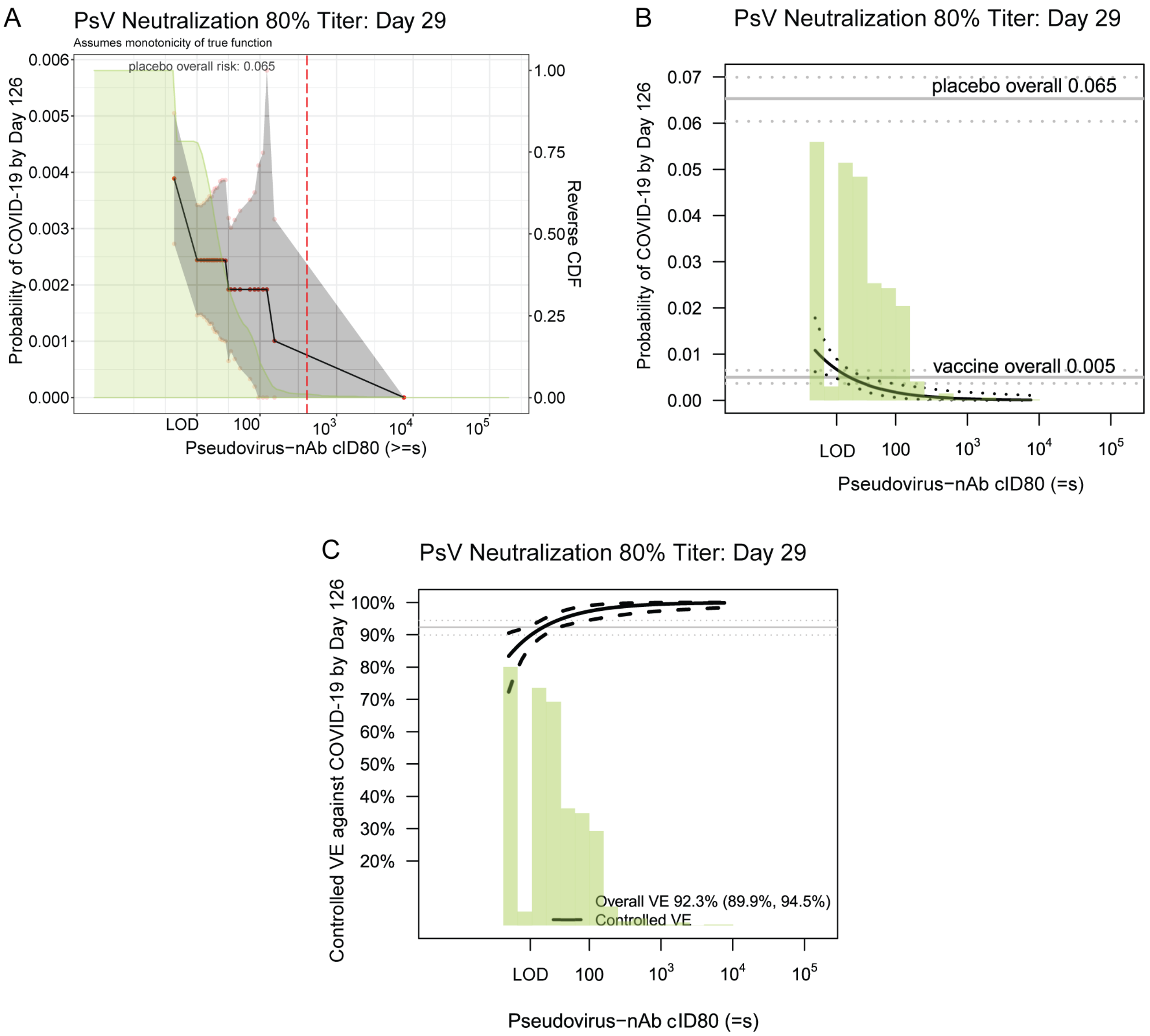

**Figure S26. (A) Covariate-adjusted risk of COVID-19 by subgroups defined by Day 29 Anti-Spike IgG level above a threshold, with reverse cumulative distribution function of Day 29 Anti-Spike IgG level overlaid in green; (B) Covariate-adjusted cumulative incidence of COVID-19 by 126 days post Day 29 by Day 29 Anti-Spike IgG level; (C) Controlled vaccine efficacy by Day 29 Anti-Spike IgG level.** In (A), the gray shaded area is pointwise 95% confidence intervals (CIs). The upper boundary of the green shaded area is the estimate of the reverse cumulative distribution function of the marker in baseline SARS-CoV-2 negative per-protocol vaccine recipients. In (B), the dotted lines indicate bootstrap point-wise 95% CIs. In (C), vaccine efficacy estimates were obtained using the method of Gilbert, Fong, and Carone.[12] In (B) and (C), the green histograms are an estimate of the density of marker level in baseline negative per-protocol vaccine recipients. LOD, limit of detection.

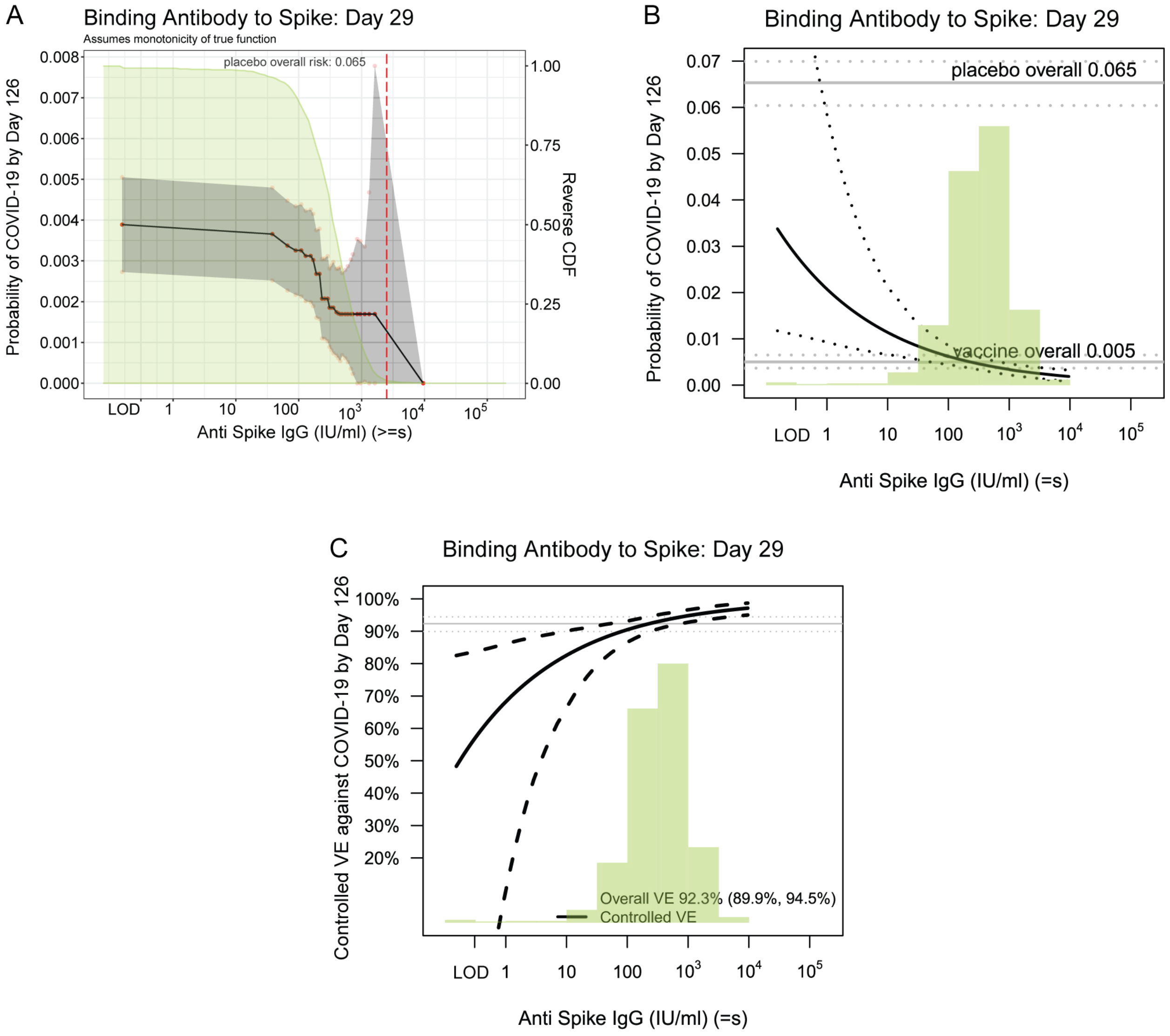

**Figure S27. (A) Covariate-adjusted risk of COVID-19 by subgroups defined by Day 29 Anti-RBD IgG level above a threshold, with reverse cumulative distribution function of Day 29 Anti-RBD IgG level overlaid in green; (B) Covariate-adjusted cumulative incidence of COVID-19 by 126 days post Day 29 by Day 29 Anti-RBD IgG level; (C) Controlled vaccine efficacy by Day 29 Anti-RBD IgG level.** In (A), the gray shaded area is pointwise 95% confidence intervals (CIs). The upper boundary of the green shaded area is the estimate of the reverse cumulative distribution function of the marker in baseline SARS-CoV-2 negative per-protocol vaccine recipients. In (B), the dotted lines indicate bootstrap point-wise 95% CIs. In (C), vaccine efficacy estimates were obtained using the method of Gilbert, Fong, and Carone.[12] In (B) and (C), the green histograms are an estimate of the density of marker level in baseline negative per-protocol vaccine recipients. LOD, limit of detection.

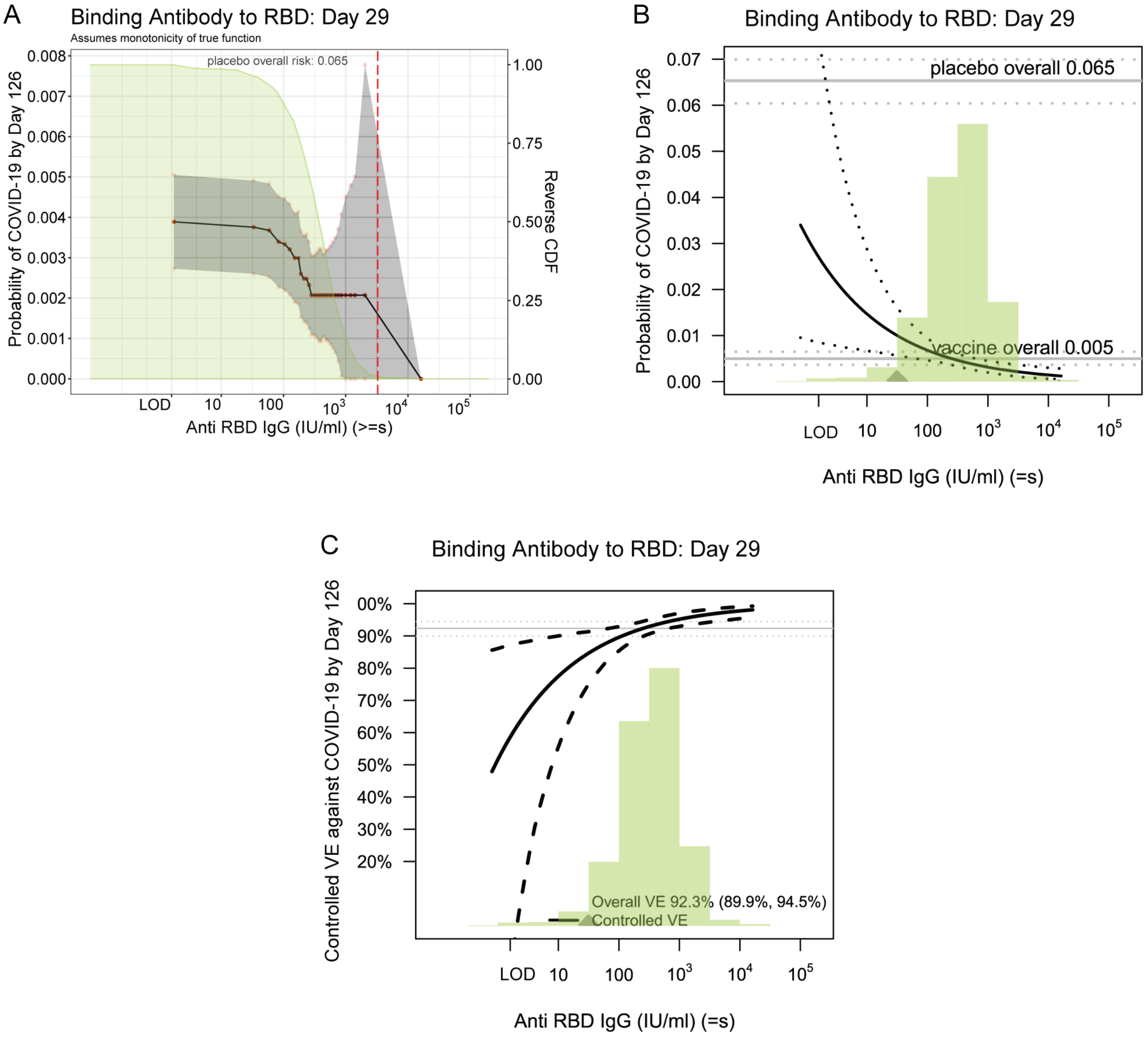

**Supplementary Text 3: Controlled vaccine efficacy sensitivity analysis to assess robustness of the effect of antibody marker on preventing COVID-19 to unmeasured confounding**

**Figure 2** reports results for estimation and inference for baseline covariate-marginalized COVID-19 risk ratios comparing vaccine recipients with Day 57 antibody marker in the third tertile vs. in the first tertile; denote this marginalized risk ratio by $RR_M(0,1)$, where 1 indicates the third tertile and 0 indicates the first tertile. As described in the SAP and Gilbert, Fong, and Carone,[12] under the assumption of no unmeasured confounding of the effect of the antibody marker on COVID-19, the COVID-19 risk ratio $RR_M(0,1)$ equals a causal effect parameter $RR_C(0,1)$, the controlled effects risk ratio, which in turn equals (1 – CVE(1))/(1 – CVE(0)), where CVE(1) is controlled vaccine efficacy for vaccine recipients with third tertile marker level and CVE(0) is controlled vaccine efficacy for vaccine recipients with first tertile marker level. In sensitivity analysis, we allow for some unmeasured confounding that makes $RR_M(0,1) < RR_C(0,1)$. In particular, as specified in the SAP the first part of the sensitivity analysis reports E-values, both for the point estimate of $RR_C(0,1)$ and for the upper 95% confidence limit of $RR_C(0,1)$. The E-value is the minimum strength of association, on the risk ratio scale, that an unmeasured confounder would need to have with both the antibody marker (exposure) and the COVID-19 outcome in order to fully explain away a specific observed exposure-outcome association, conditional on the measured covariates.[13] Here 'fully explain away' means that $RR_C(0,1) =1$, or equivalently CVE(0)=CVE(1), i.e., controlled vaccine efficacy does not differ by upper vs. lower tertile, which means no correlate of protection. Many epidemiologists recommend that E-values are always reported (or alternative sensitivity metrics) for causal inferences, given that it is not possible to guarantee that all confounders are controlled for in the analysis; thus by using E-values we are implementing best practice.

In addition, we apply the sensitivity analysis technique developed by Ding and Vanderweele (2016),[14] as implemented in the SAP and in Gilbert, Fong, and Carone[12] for the COVE trial correlates application, to estimate $RR_C(0,1) = (1 – CVE(1))/(1 – CVE(0))$ under a specified scenario of unmeasured confounding that makes it harder to conclude a correlate of protection. This sensitivity analysis specifies an amount of unmeasured confounding defined by $RR_{UD}(0,1)=RR_{EU}(0,1)=2$, where these parameters are defined in Ding and Vanderweele (2016)[14] and in our SAP and Gilbert et al. If the 95% confidence interval for $RR_C(0,1) = (1 – CVE(1))/(1 – CVE(0))$ is less than 1 after accounting for this confounding that pushes the confidence interval upwards, it provides evidence for a correlate of protection with a degree of robustness to possible unmeasured confounding.

**Table S8** shows results for the eight Day 57 and Day 29 antibody markers categorized by upper vs. lower tertile that were assessed as correlates. For example, the E-value of 9.3 for the Day 57 cID80 marker means that a marginalized risk ratio $RR_M(0,1)$ at the observed value 0.20 could be explained away (i.e., $RR_C(0,1) = 1.0$) by an unmeasured confounder associated with both the exposure and the outcome by a marginalized risk ratio of 9.3-fold each, after accounting for the measured potential confounders (risk score, at-risk status, community of color indicator), but that weaker confounding could not do so. In addition, the E-value of 3.3 for the upper confidence limit UL indicates the strength of unmeasured confounding at which statistical significance of the inference that CVE(1) > CVE(0) would be lost.

**Table S8. Sensitivity analysis to assess Day 57 and Day 29 antibody markers categorized as upper vs. lower tertiles as controlled vaccine efficacy CoPs against COVID-19**

| | Marginalized Risk Ratio $RR_M(0,1)$ | | Controlled Risk Ratio = (1-CVE(1))/(1-CVE(0))[1] | | E-values[2] | |
|---|---|---|---|---|---|---|
| Antibody Marker | Point Est. | 95% CI | Point Est. | 95% CI | For Point Est. | For 95% CI UL |
| Day 57 Spike IgG | 0.24 | 0.06, 0.56 | 0.32 | 0.09, 0.75 | 7.9 | 3.0 |
| Day 57 RBD IgG | 0.28 | 0.08, 0.62 | 0.38 | 0.11, 0.83 | 6.5 | 2.6 |
| Day 57 PsV cID50 | 0.31 | 0.08, 0.72 | 0.42 | 0.11, 0.96 | 5.9 | 2.1 |
| Day 57 PsV cID80 | 0.20 | 0.03, 0.51 | 0.27 | 0.05, 0.68 | 9.3 | 3.3 |
| Day 29 Spike IgG | 0.19 | 0.06, 0.40 | 0.26 | 0.08, 0.53 | 9.8 | 4.5 |
| Day 29 RBD IgG | 0.29 | 0.10, 0.59 | 0.38 | 0.13, 0.79 | 6.5 | 2.8 |
| Day 29 PsV cID50 | 0.33 | 0.13, 0.65 | 0.44 | 0.17, 0.86 | 5.5 | 2.5 |
| Day 29 PsV cID80 | 0.22 | 0.07, 0.46 | 0.30 | 0.10, 0.61 | 8.5 | 3.8 |

[1]Conservative (upper bound) estimate assuming unmeasured confounding at level $RR_{UD}(0, 1) = RR_{EU}(0, 1) = 2$ and thus $B(0, 1) = 4/3$.
[2]E-values are computed for upper tertile ($s = 1$) vs. lower tertile ($s = 0$) biomarker subgroups after controlling for baseline risk score, at risk or not, community of color or not; UL = upper limit.

A second sensitivity analysis was conducted to assess robustness of findings on correlates of vaccine efficacy as a function of the quantitative antibody marker level varying over its whole range as studied in **Figure 4C** for cID50 titer and in **Figures S21-S27** for the other seven antibody markers. With details in the SAP and Gilbert, Fong, and Carone,[12] the sensitivity analysis was done similar to the analysis above for categorized markers, except now instead of specifying $RR_{UD}(0,1)$ and $RR_{EU}(0,1)$ for third tertile vs. first tertile, we specified $RR_{UD}(s^{15}, s^{85}) = 2$ and $RR_{EU}(s^{15}, s^{85}) = 2$ for $s^{15}$ the 15th percentile of the antibody marker and $s^{85}$ the 85th percentile of the antibody marker. We then specified unmeasured confounding across the whole range of antibody marker levels through the model

$$\log(RR_{UD}(s1, s2)) = [(s2 - s1)/ (s^{85} - s^{15})] \log(RR_{UD}(s^{15}, s^{85})) \text{ for all } s1 \leq s2,$$

with the same model used for $RR_{EU}(s1, s2)$. The result of the sensitivity analysis is point estimates of controlled vaccine efficacy CVE(s) over the range s of antibody marker levels, with 95% bootstrap pointwise confidence intervals, which build in robustness to unmeasured confounding by supposing bias that makes the estimate of the curve CVE(s) flatter.

**Figures S28 and S29** show the results for the eight antibody markers.

**Figure S28. Vaccine efficacy with sensitivity analysis by Day 57 (A) Anti-Spike IgG level, (B) Anti-RBD IgG level, (C) cID50 level, or (D) cID80 level.** Vaccine efficacy estimates were obtained using the method of Gilbert, Fong, and Carone.[12] The upper boundary of the green shaded area is the estimate of the reverse cumulative distribution function of the marker in baseline SARS-CoV-2 negative per-protocol vaccine recipients. The pink solid line is point estimates assuming no unmeasured confounding; the dashed lines are bootstrap point-wise 95% CIs. The red solid line is point estimates assuming unmeasured confounding in a sensitivity analysis (dashed lines are bootstrap point-wise 95% CIs); see Supplementary Text 3 for details of the sensitivity analysis. LOD, limit of detection.

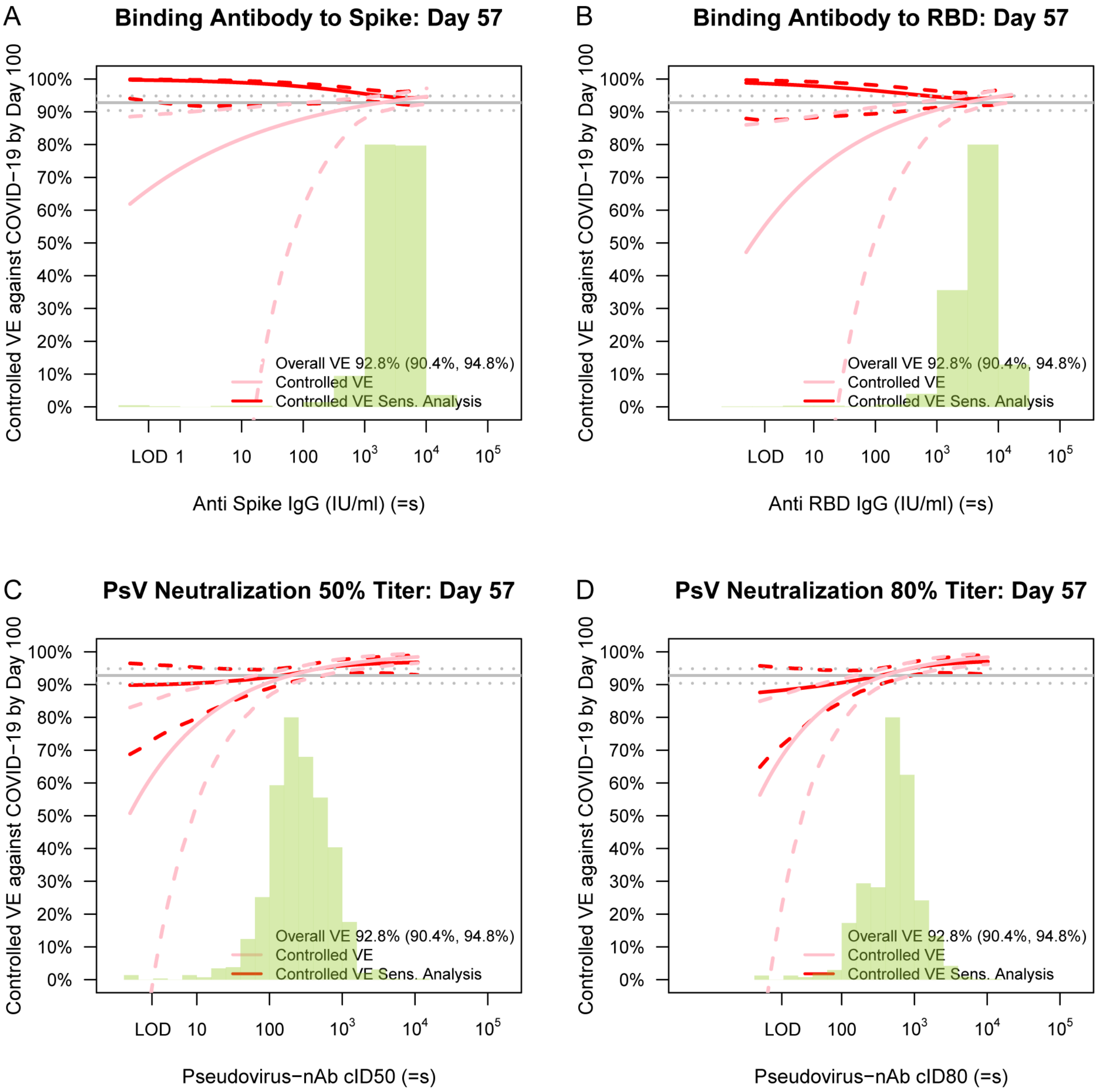

**Figure S29. Vaccine efficacy with sensitivity analysis by Day 29 (A) Anti-Spike IgG level, (B) Anti-RBD IgG level, (C) cID50 level, or (D) cID80 level.** Vaccine efficacy estimates were obtained using the method of Gilbert, Fong, and Carone.[12] The upper boundary of the green shaded area is the estimate of the reverse cumulative distribution function of the marker in baseline SARS-CoV-2 negative per-protocol vaccine recipients. The pink solid line is point estimates assuming no unmeasured confounding; the dashed lines are bootstrap point-wise 95% CIs. The red solid line is point estimates assuming unmeasured confounding in a sensitivity analysis (dashed lines are bootstrap point-wise 95% CIs); see Supplementary Text 3 for details of the sensitivity analysis. LOD, limit of detection.

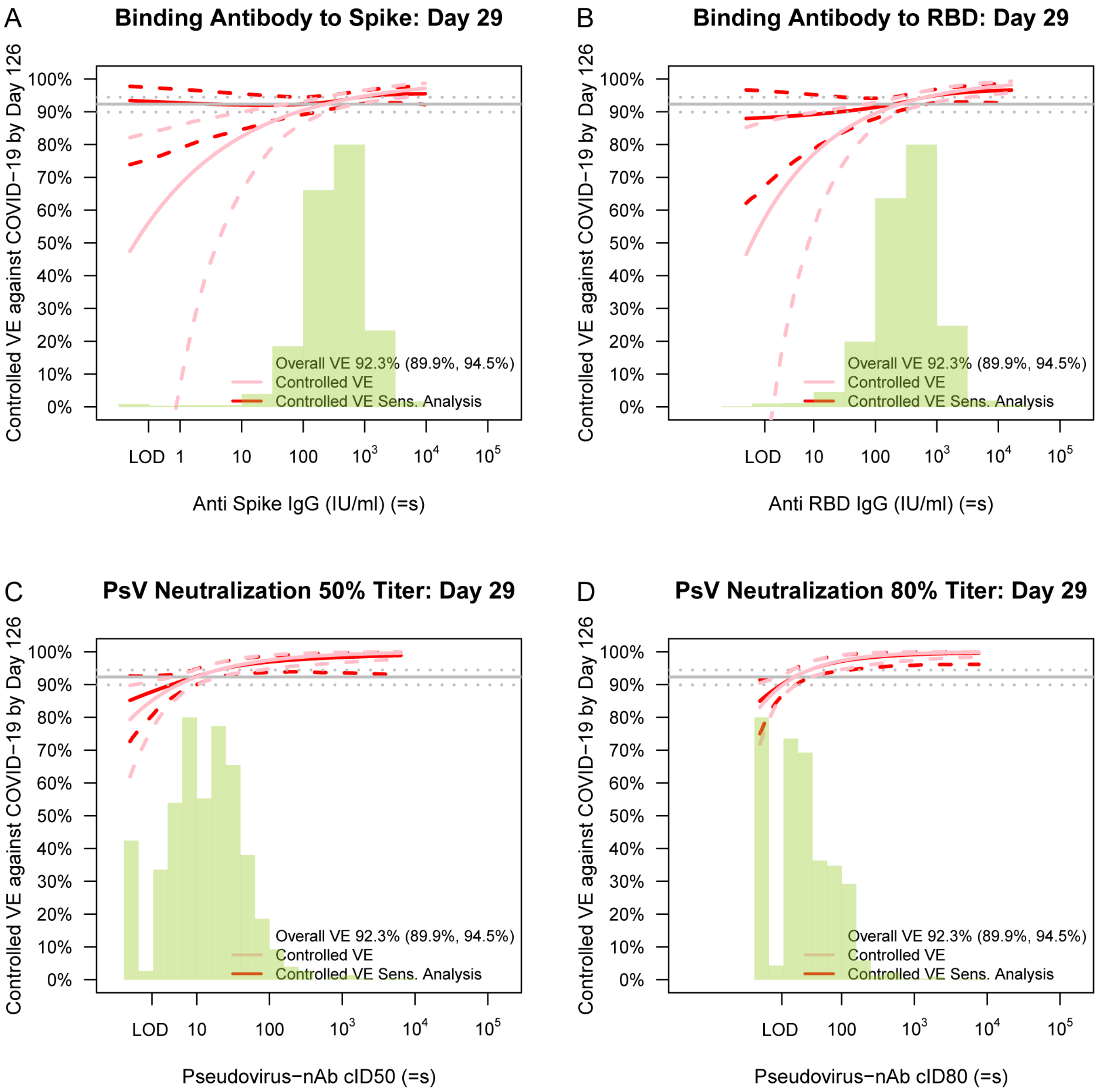

**Table S9. Table of mediation effect estimates for quantitative markers with 95% confidence intervals.**

Direct VE = VE comparing vaccine vs. placebo with marker set to distribution in placebo.
Indirect VE = VE in vaccinated comparing observed marker vs. hypothetical marker under placebo.
Prop. mediated = fraction of total risk reduction from vaccine attributed to antibody response.

| Time | Assay | Direct VE | Indirect VE | Prop. mediated |
|---|---|---|---|---|
| Day 29 | PsV Neutralization 50% Titer (Calibrated, cID50) | 0.560 (0.422, 0.665) | 0.832 (0.769, 0.878) | 0.685 (0.585, 0.784) |
| Day 29 | PsV Neutralization 80% Titer (Calibrated, cID80) | 0.739 (0.601, 0.829) | 0.717 (0.597, 0.801) | 0.485 (0.345, 0.624) |
