## Supplementary material for "Immune Correlates Analysis of the mRNA-1273 COVID-19 Vaccine Efficacy Trial": Statistical Analysis Plan

### Statistical Analysis Plan for Assessing Immune Correlates in the Coronavirus Efficacy (COVE) Phase 3 Trial of the mRNA-1273 COVID-19 Vaccine

USG COVID-19 Response Team / Coronavirus Prevention Network (CoVPN) Biostatistics Team

Peter B. Gilbert[1,2*], Youyi Fong[1,2], David Benkeser[3], Jessica Andriesen[1], Bhavesh Borate[1], Marco Carone[2], Lindsay N. Carpp[1], Iván Díaz[4], Michael P. Fay[5], Andrew Fiore-Gartland[1], Nima S. Hejazi[6], Ying Huang[1,2], Yunda Huang[1], Ollivier Hyrien[1], Holly E. Janes[1,2], Michal Juraska[1], Kendrick Li[2], Alex Luedtke[7], Martha Nason[5], April K. Randhawa[1], Lars van der Laan[6], Brian D. Williamson[1], Wenbo Zhang[2], Dean Follmann[5]

[1]Vaccine and Infectious Disease and Public Health Sciences Divisions, Fred Hutchinson Cancer Research Center, Seattle, Washington

[2]Department of Biostatistics, University of Washington, Seattle, Washington

[3]Department of Biostatistics and Bioinformatics, Rollins School of Public Health, Emory University, Atlanta, Georgia

[4]Department of Population Health Sciences, Weill Cornell Medical College, New York, New York

[5]National Institute of Allergy and Infectious Diseases, Bethesda, Maryland

[6]Division of Biostatistics, School of Public Health, University of California, Berkeley, California

[7]Department of Statistics, University of Washington, Seattle, Washington

August 9, 2021

Contents

**List of Tables** **5**

**List of Figures** **6**

**1 Introduction** **7**

**2 Antibody Assays and Day 29 and Day 57 Markers** **7**

**3 Study Cohorts and Endpoints** **12**

**7 Baseline Risk Score (Proxy for SARS-CoV-2 Exposure)** **28**

**8 Correlates Analysis Descriptive Tables by Case/Non-Case Status** **29**

**9 Correlates of Risk Analysis Plan** **30**

**10 Correlates of Protection: Generalities** **50**

**11 Correlates of Protection: Correlates of Vaccine Efficacy Analysis Plan** **50**

**12 Correlates of Protection: Interventional Effects** **54**

**13 Summary of the Set of CoR and CoP Analyses and Their Requirements and Contingencies, and Synthesis of the Results, Including Reconciling Any Possible Contradictions in Results 68**

13.1 Synthesis Interpretation of Results . . . . . . . . . . . . . . . . 70

13.2 Multiple Hypothesis Testing Adjustment for CoP Analysis . . 75

**14 Estimating a Threshold of Protection Based on an Established or Putative CoP (Population-Based CoP) 76**

**15 Considerations for Baseline SARS-CoV-2 Positive Study Participants 77**

**16 Avoiding Bias with Pseudovirus Neutralization Analysis due to Use of Anti-HIV Antiretroviral Drugs 77**

**17 Accommodating Crossover of Placebo Recipients to the Vaccine Arm 78**

4 Two-stage correlates analysis. Stage 1 consists of analyses of Day 29 and Day 57 markers as correlates of risk and of protection of the primary endpoint and potentially also of some secondary endpoints, and includes antibody marker data from all COVID and SARS-CoV-2 infection cases (COV-INF) through to the time of the data lock for the first correlates analyses. Stage 2 consists of analyses of Day 29 and Day 57 markers as correlates of risk and of protection of longer term endpoints and analyses of longitudinal markers as outcome-proximal correlates of risk and of protection, and includes antibody marker data from all subsequent COVID and COV-INF cases. Stage 1 measures Day 1, 29, 57 antibody markers and COV-INF and COVID diagnosis time point markers; Stage 2 measures antibody markers from all sampling time points and COV-INF plus COVID diagnosis sampling time points not yet assayed. The same immunogenicity subcohort is used for both stages. . 82

### 1 Introduction

This SAP describes the statistical analysis of antibody markers measured at Day 29 and at Day 57 as immune correlates of risk and as immune correlates of protection against the COVID primary endpoint in the Coronavirus Efficacy (COVE) phase 3 trial of the mRNA-1273 COVID-19 vaccine. In this trial, estimated efficacy of the mRNA-1273 vaccine against symptomatic COVID illness was 94.1% (95% confidence interval, 89.3 to 96.8%) [Baden et al. (2021)].

### 2 Antibody Assays and Day 29 and Day 57 Markers

The antibody markers of interest are measured using two different humoral immunogenicity assays [more detail on assay type (2) can be found in Sholukh et al. (2020)]:

(1) **bAbs**: **Binding antibodies** to the vaccine insert SARS-CoV-2 proteins;

and (2) **Pseudovirus-nAbs**: **Neutralizing antibodies** against viruses **pseudotyped** with the vaccine insert SARS-CoV-2 proteins.

The Supplementary text in the article provides details of the assays. We include the necessary statistical details below.

(1) **bAb assay**: The MSD-ECL Multiplex Assay (MSD-ECL = meso scale discovery-electrochemiluminescence assay).

The MSD assay measures binding antibody to antigens corresponding to: Spike (an engineered version of the Spike protein harboring a double proline substitution (S-2P) that stabilizes it in the closed, prefusion conformation [McCallum et al. (2020)]); the Receptor Binding Domain (RBD) of the Spike protein; and Nucleocapsid protein (N), which is not contained in any of the COVID-19 vaccines.

The bAb assay readouts are in units AU/ml, where AU stands for arbitrary units from a standard curve. The process of validating the assay defined a lower limit of detection (LLOD), an upper limit of detection (ULOD), a lower limit of quantitation (LLOQ), an upper limit of quantitation (ULOQ), and a positivity cut-off for each antigen that defines positive vs. negative response. These values are as follows:

- bAb Spike:
  - Pos. Cutoff = 1204.71 AU/ml
  - LLOD = 34.18 AU/ml
  - ULOD = 19,136,250 AU/ml
  - LLOQ = 199.64 AU/ml
  - ULOQ = 1,128,438.87 AU/ml
- bAb RBD:
  - Pos. Cutoff = 517.86 AU/ml
  - LLOD = 58.59 AU/ml
  - ULOD = 8,201,250 AU/ml
  - LLOQ = 125.9678 AU/ml
  - ULOQ = 598,133.3615 AU/ml
- N:
  - Pos. cutoff = 9779.62 AU/ml
  - LLOD = 39.06 AU/ml
  - ULOD = 21,870,000
  - LLOQ = 1870.70 AU/ml
  - ULOQ = 239,449.31

The Vaccine Research Center established factors for converting the MSD assay readouts from AU/ml to WHO International Units/ml. For the three binding antibody variables CoV-2 Spike IgG, CoV-2 RBD IgG, and CoV-2 N IgG, these conversion factors are 0.0090, 0.0272, and 0.0024, respectively. These conversion factors are applied, such that all binding Ab readouts are reported in WHO International Units/ml (IU/ml), for all analyses. These conversion factors are also applied to yield the LLOD, ULOD, LLOQ, and ULOQ on the WHO IU/ml scale. The following shows the assay limits on the IU/ml scale:

- bAb Spike:
  - Pos. Cutoff = 10.8424 IU/ml
  - LLOD = 0.3076 IU/ml
  - ULOD = 172,226.2 IU/ml
  - LLOQ = 1.7968 IU/ml
  - ULOQ = 10,155.95 IU/ml
- bAb RBD:
  - Pos. Cutoff = 14.0858 IU/ml
  - LLOD = 1.593648 IU/ml
  - ULOD = 223,074 IU/ml
  - LLOQ = 3.4263 IU/ml
  - ULOQ = 16,269.23 IU/ml
- bAb N:
  - Pos. Cutoff = 23.4711 IU/ml
  - LLOD = 0.093744 IU/ml
  - ULOD = 52,488 IU/ml
  - LLOQ = 4.4897 IU/ml
  - ULOQ = 574.6783 IU/ml

All values below the LLOD are assigned the value LLOD/2. For immunogenicity reporting, values greater than the ULOQ are not given a ceiling value of the ULOQ, the actual readouts are used. For the immune correlates analyses, values greater than the ULOQ are assigned the value of the ULOQ.

(2) **Pseudovirus-nAb assay**: A firefly luciferase (ffLuc) reporter neutralization assay for measuring neutralizing antibodies against SARS-CoV-2 Spike-pseudotyped viruses.

Based on the assay in the Duke lab of David Montefiori, serum inhibitory dilution 50% titer (ID50) and serum inhibition dilution 80% titer (ID80) values are estimated based on a starting serum dilution of 1:10, with eight 5-fold dilutions. Thus 1:10 is the LLOD on the scale of the assay. The process of validating the assay defined the LLOD, LLOQ, and ULOQ for ID50 and ID80 as follows:

- ID50:
  - LLOD = 10
  - LLOQ = 18.5
  - ULOQ = 45118
- ID80:
  - LLOD = 10
  - LLOQ = 14.3
  - ULOQ = 10232

ID50 and ID80 values below the LLOD are assigned the value 10/2 = 5. Values between the LLOD and the LLOQ are taken as their actual numeric value. For immunogenicity reporting, values greater than the ULOQ are not given a ceiling value of the ULOQ, the actual readouts are used. For the immune correlates analyses, values greater than the ULOQ are assigned the value of the ULOQ.

ID50 and ID80 values are reported in international units based on the report from David Montefiori "Reagent Calibration Report: First WHO International Standard for SARS-CoV-2 Immunoglobulin in a Neutralization Assay" (May, 2021). This report derived calibration factors based on arithmetic means:

- Calibration factor ID50: 0.242

- Calibration factor ID80: 1.502

The original readouts are calibrated to the IU scale by multiplying each original ID50 value by 0.242, and multiplying each original ID80 value by 1.502, and units are reported as calibrated ID50 (cID50) and calibrated ID80 (cID80). Consequently, the LLOD, LLOQ and ULOQ for cID50 and cID80 are as follows in International Units:

- cID50:
  - LLOD = 2.42
  - LLOQ = 4.477
  - ULOQ = 10919
- cID80:
  - LLOD = 15.02
  - LLOQ = 21.4786
  - ULOQ = 15368

Based on each immunoassay applied to triples of serum samples collected from participants on Day 1 (baseline, first dose of vaccination visit), Day 29 (second dose of vaccination visit), and Day 57 (post-vaccination visit), the following set of antibody markers was defined for immunogenicity and immune correlates analyses.

- For bAb: $\log_{10}$ IgG concentration (IU/ml) at each time point, the difference in $\log_{10}$ concentration (Day 29 minus Day 1) representing $\log_{10}$ fold-rise in IgG concentration from baseline to dose two, and the difference in $\log_{10}$ concentration (Day 57 minus Day 1) representing $\log_{10}$ fold-rise in IgG concentration from baseline to 28 days post dose two. These markers are defined for each antigen Spike, RBD, and N.
- For PsV nAb: $\log_{10}$ serum inhibitory dilution 50% titer (cID50) and serum inhibition dilution 80% titer (cID80) at each time point, as well as the $\log_{10}$ fold-rise of these markers over Day 1 to Day 29, and over Day 1 to Day 57.

#### 3 Study Cohorts and Endpoints

##### 3.1 Study Cohort for Correlates Analyses

The analysis cohort for the correlates analysis is baseline SARS-CoV-2 negative participants in the per-protocol cohort, with the per-protocol cohort defined as those who received both planned vaccinations without any specified protocol deviations, and who were SARS-CoV-2 negative at the terminal vaccination visit. We refer to this cohort representing the primary population for correlates analysis as the Per-Protocol Baseline Negative Cohort. The definition of baseline negative and per-protocol are the same as in Baden et al. (2021).

As the primary analysis of vaccine efficacy is conducted in baseline negative individuals, correlates of risk (CoR) and correlates of protection (CoP) analyses are only done in baseline negative individuals, and the analysis of data from baseline positive individuals is for purposes of immunogenicity characterization, given too-few anticipated vaccine breakthrough study endpoints for CoR/CoP assessment (although if there are many baseline positive vaccine breakthrough endpoint cases that baseline positive subgroup analyses may be considered). In baseline negative individuals, antibody marker data in placebo recipients is relevant for verifying the expectation that almost all Day 29 and Day 57 marker responses will be negative, given the lack of SARS-CoV-2 antigen exposure.

##### 3.2 Study Endpoints

Endpoints for correlates analyses of Day 57 markers are included if they occur at least 7 days after the Day 57 visit, to help ensure that the endpoint did not occur prior to Day 57 antibody measurement. Similarly, endpoints for correlates analyses of Day 29 markers are included if they occur at least 7 days after the Day 29 visit, again to help ensure that the endpoint did not occur prior to Day 29 antibody measurement.

Figure 1 defines five study endpoints assessed in COVID-19 vaccine efficacy trials, where COVID (symptomatic infection) is used as the primary endpoint in the COVE trial. Only the COVID endpoint is assessed in the current manuscript. For the correlates analysis, all available follow-up for participants is included through to the time of the data base lock for the correlates analysis, for every CoR and CoP analysis that is conducted. This means that the time of right censoring for a given failure time endpoint is the first event of loss to follow-up or the date of administrative censoring defined as the last date of available follow-up. For CoP analyses, which use both vaccine and placebo recipient data and leverage the randomization, follow-up is censored at the time of unblinding. In general for the current manuscript all blinded follow-up is included and no post-unblinding follow-up is included.

### 4 Objectives of Immune Correlates Analyses of a Phase 3 Trial Data Set

#### 4.1 Correlates of Risk and Correlates of Protection

We broadly classify the proposed analyses into two related categories: correlates of risk (CoR) and correlates of protection (CoP) analyses. CoR analyses seek to characterize correlations/associations of markers with future risk of the outcome amongst vaccinated individuals in the study cohort. CoP analyses seek to formally characterize causal relationships among vaccination, antibody markers and the study endpoint, and use data from both vaccine and placebo recipients. Table 1 summarizes these objectives and statistical frameworks that are commonly used to these ends; while the table focuses on Day 57 markers, the same objectives are of interest for Day 29 markers.

The advantage of CoR analyses it that it is possible to obtain definitive answers from the phase 3 data sets, that is one can credibly characterize associations between markers and outcome. The advantage of CoP analyses is that the effects being estimated have interpretation directly in terms of how an antibody marker can be used to reliably predict vaccine efficacy (the criterion for use of a non-validated surrogate endpoint for accelerated approval, Fleming and Powers, 2012). The disadvantage of CoR analyses are that a CoR may fail to be a CoP, for example due to unmeasured confounding, lack of transitivity where a vaccine effect on an antibody marker occurs in different individuals than clinical vaccine efficacy, or off-target effects (VanderWeele, 2013). The disadvantage of CoP analyses is that statistical inferences rely on causal assumptions that cannot be completely verified from the phase 3 data, such that compelling evidence may require multiple phase 3 trials and external evidence on mechanism of protection (e.g., from adoptive transfer or vaccine challenge trials). Our approach presents results for both CoR and CoP analyses, seeking clear exposition of how to interpret results, the assumptions undergirding the validity of the results, and diagnostics of these assumptions and assessment of robustness of findings to violation of assumptions.

Table 1: Correlates of Risk (CoRs) and Correlates of Protection (CoPs) Objectives for Day 57 Markers

| **Objective Type** | **Objective** |
|---|---|
| **CoRs (Risk Prediction Modeling)** | **To assess Day 57 markers as CoRs in vaccine recipients**<br>a. Relative risks of outcome across marker levels<br>b. Absolute risk of outcome across marker levels<br>c. Machine learning risk prediction for multivariable markers |
| **CoP: Correlates of VE** | **To assess Day 57 markers as correlates of VE in vaccine recipients**<br>a. Principal stratification effect modification analysis<br>b. Assesses VE across subgroups of vaccine recipients defined by Day 57 marker level in vaccine recipients |
| **CoP: Controlled Effects on Risk and VE** | **To assess Day 57 markers for how assignment to vaccine and a fixed marker value would alter risk compared to assignment to placebo** |
| **CoP: Stochastic Interventional Effects on Risk and VE** | **To assess Day 57 markers for how stochastic shifts in their distribution would alter mean risk and VE** (Hejazi et al., 2020) |
| **CoP: Mediators of VE** | **To assess Day 57 markers as mediators of VE**<br>a. Mechanisms of protection via natural direct and indirect effects<br>a. Estimate the proportion of VE mediated by a marker or markers |

##### 4.2 Synthesis of the Phase 3 Correlates Analyses for Decisions

Establishment of an immunologic biomarker for approval/bridging applications is generally not based on pre-fabricated criteria nor a single type of correlates analysis. Therefore, the goal of the correlates analysis is to generate evidence about correlates from many perspectives, and to synthesize the evidence to support certain decisions. Consequently, we believe there is value in assessing all of the types of correlates presented in Table 1, given that the analyses address distinct questions. Obtaining a set of results from multiple distinct approaches that provide complementary and coherent support may increase the rigor and robustness of an evidence package supporting potential use of an antibody marker as a validated surrogate (for traditional approval) or as a non-validated surrogate (for accelerated approval) (Fleming and Powers, 2012); these uses of a biomarker are summarized below. However, the assumptions needed for valid inferences are somewhat different across the methods, and some of these assumptions have testable implications; therefore examination of the assumptions may lead to favoring some methods over others, and affect the synthesis and interpretation of results, and moreover if diagnostics support that some necessary assumptions are infeasible then certain analyses will be canceled, as described below. Section 13 summarizes the approach that is used and the interpretation of the set of multiple correlates of protection methods.

#### 5 Case-cohort Sampling Design for Measuring Antibody Markers

Figure 3 illustrates the case-cohort (Prentice, 1986) sampling design that is used for measuring Day 1, 29, 57 antibody markers in a random sample of trial participants. The random sample is stratified by the key baseline covariates: assigned randomization arm, baseline SARS-CoV-2 status (defined by serostatus and NAAT and/or RNA PCR testing, Baden et al., 2021), and randomization strata (defined by age and heightened COVID at-risk status). Because the design uses a stratified random sample instead of the simple random sample proposed by Prentice (1986), the design may also be referred to as a "two-phase sampling design" (Breslow et al., 2009b,a), where "phase one" refers to variables measured in all participants and "phase two" refers to variables only measured in a subset (thus the "case-cohort sample" constitutes the phase-two data).

The case-cohort design enables obtaining marker data (Day 1, 29, 57) for the immunogenicity subcohort during early trial follow-up in real-time batches, thereby accelerating the time until final data set creation and hence data analysis and results on Day 29 and Day 57 marker correlates. The design allows using the same immunogenicity subcohort to assess correlates for multiple endpoints, relevant for the COVID-19 VE trials with multiple endpoints (Figure 1). This makes the design operationally simpler than a case-control sampling design.

##### 5.1 Immunogenicity subcohort

The immunogenicity subcohort was sampled from the subset of participants in the Full Analysis Set (FAS) cohort used in the primary analysis of vaccine efficacy against the primary endpoint (with the FAS defined as all randomized participants who received at least one dose of investigational product) for whom all of the following information was available: baseline SARS-CoV-2 status; age, race/ethnicity (needed to define Minority status as described below), and heightened COVID at-risk status; and Day 1, Day 29, and Day 57 samples collected.

Table 2 summarizes the planned size of the immunogenicity subcohort, by the baseline factors used to stratify the random sampling. In this subcohort 6 baseline demographic strata are used. A 50:50 balance is specified by minority status Yes:No. The subcohort sampling is implemented to create representative sampling across the entire period of enrollment.

For the sampling, Minority includes Blacks or African Americans, Hispanics or Latinos, American Indians or Alaska Natives, Native Hawaiians, and other Pacific Islanders. Non-Minority includes all other races with observed race (Asian, Multiracial, White, Other) and observed ethnicity Not Hispanic or Latino. Therefore Unknown and Not reported have missing values for this sampling stratum variable.

"At-risk" refers to participants considered to be at heightened risk of severe COVID-19 illness. Only participants 18-64 were categorized as either "At-risk" or "Not at-risk". Specifically, participants 18-64 were categorized as "at-risk" if they had at least one of the following risk factors: chronic lung disease (e.g., emphysema, chronic bronchitis, idiopathic pulmonary fibrosis, cystic fibrosis, or moderate-to-severe asthma); cardiac disease (e.g., heart failure, congenital coronary artery disease, cardiomyopathies, or pulmonary hypertension); severe obesity (BMI $\geq$40); diabetes (type 1, type 2, or gestational); liver disease; or HIV infection.

Table 2: Planned Immunogenicity Subcohort Sample Sizes by Baseline Strata for Antibody Marker Measurement

| | Baseline SARS-CoV-2 Negative[2] | | | | | | Baseline SARS-CoV-2 Positive[3] | | | | | |
|---|---|---|---|---|---|---|---|---|---|---|---|---|
| Bas. Cov. Strata[1] | 1 | 2 | 3 | 4 | 5 | 6 | 1 | 2 | 3 | 4 | 5 | 6 |
| Vaccine | 150 | 150 | 150 | 150 | 150 | 150 | 50 | 50 | 50 | 50 | 50 | 50 |
| Placebo | 20 | 20 | 20 | 20 | 20 | 20 | 50 | 50 | 50 | 50 | 50 | 50 |

[1]Sampling was stratified within 6 baseline covariate strata:
1 = Age 18-64 Minority At-risk; 2 = Age 18-64 Non-Minority At-risk; 3 = Age 18-64 Minority Not At-risk; 4 = Age 18-64 Non-Minority Not At-risk; 5 = Age $\geq$ 65 Minority; 6 = Age $\geq$ 65 Non-Minority
[2]The vaccine group baseline negative strata are assigned large sample sizes because the correlates of risk analysis focuses on baseline negative vaccine recipients. The placebo group baseline negative strata are assigned small sample sizes given the expectation that almost all Day 57 bAb and nAb readouts will be negative/zero given the absence of prior exposure to SARS-CoV-2 antigens.
[3]Equal stratum sizes are assigned for the vaccine and placebo groups in order to compare bAb and nAb responses in previously infected persons, studying potential differences in natural+vaccine-elicited responses vs. natural-elicited responses.

If certain strata do not have enough eligible participants available for sampling, then additional sampling is done from other strata to keep the total immunogenicity subcohort sample size close to 1620 or somewhat higher.

Figure S1 and Table S3 in the Supplementary Material describe the actual numbers of participants sampled into the baseline negative portion of the immunogenicity subcohort – the relevant portion given the focus of correlates analyses on baseline negative participants.

##### 5.2 Correlates Objectives Addressed in Two Stages

Figure 4 depicts the two stages of the immune correlates analyses. Stage 1 includes antibody marker data from all COVID and infection (COV-INF) cases diagnosed through to the last date of: (1) the time that at least 25 evaluable vaccine breakthrough COVID endpoint cases are available for analysis; and (2) the time of a data-cut at or after the primary analysis used to define the data base for the first correlates analysis. Only Day 1, 29, 57 antibody markers, and COVID and COV-INF diagnosis time point antibody markers, are measured in Stage 1. The objectives of Stage 1 correlates analyses focus on Day 29 and Day 57 markers, which are the objectives listed in Table 1. Stage 1 focuses on Day 57 markers because in general validated or non-validated surrogate endpoints for approved vaccines are based on the peak antibody time point, and this approach fits the priority to develop a validated or non-validated surrogate endpoint as rapidly as possible. Stage 1 also focuses on Day 29 markers because if a correlate based on this time point is found to perform as well as a Day 57 correlate, then it may be preferred given the practical advantage to be measured earlier and to not require a Day 57 post-vaccination visit and blood draw. Another advantage of an earlier measurement is providing opportunity to include additional breakthrough COVID endpoint cases (intercurrent endpoints) in the correlates analyses.

Stage 2 includes antibody marker data from all COVID and COV-INF cases diagnosed after the Stage 1 cases through to the end of the trial, including all available sampling time points (6–7 time points). For immunogenicity subcohort participants, the antibody markers at all available time points other than Day 1, 29, 57 are measured for Stage 2 correlates analyses (4–5 additional time points). The Stage 2 clinical endpoint data and antibody marker data enable assessment of longitudinal antibody markers as outcome-proximal correlates of instantaneous endpoint risk and as various types of outcome-proximal correlates of protection.

The manuscript restricts to assessment of Stage 1 correlates.

### 6 Unsupervised Feature Engineering of Antibody Markers (Stage 1: Day 1, 57)

#### 6.1 Descriptive Tables and Graphics

##### 6.1.1 Antibody marker data

Binding antibody titers to full length SARS-CoV-2 Spike protein, to the RBD domain of the Spike protein, and to the Nucleocapsid (N) protein will be measured in all participants in the immunogenicity subcohort (augmented with infected cases). N-specific binding antibody titers are not used for correlates analyses or for graphical reporting; these data are only used for tabular reporting. Binding antibody IgG Spike, IgG RBD, IgG N, as well as fold-rise in these three markers from baseline, are measured at each pre-defined time point. Indicators of 2-fold rise and 4-fold rise in IgG concentration (fold rise [post/pre] $\geq 2$ and $\geq 4$, 2FR and 4FR) are measured at each pre-defined post-vaccination timepoint. Binding antibody responders to a given antigen at each pre-defined timepoint are defined as participants with value above the antigen-specific positivity cut-off. Binding antibody IgG 2FR (4FR) at each pre-defined timepoint to a given antigen are defined as participants who had baseline values below the LLOQ with IgG concentration at least 2 times (4 times) above the assay LLOQ, or as participants with baseline values above the LLOQ with at least a 2-fold (4-fold) increase in IgG concentration.

Pseudovirus neutralizing antibody cID50 and cID80 titers, as well as fold-rise in cID50 and cID80 titers from baseline, are measured at each pre-defined time point. Indicators of 2-fold rise and 4-fold rise in cID50 titer (fold rise [post/pre] $\geq 2$ and $\geq 4$, 2FR and 4FR) are measured at each pre-defined post-vaccination timepoint. Neutralization responders at each pre-defined timepoint are defined as participants who had baseline values below the LLOD with detectable cID50 neutralization titer above the assay LLOD, or as participants with baseline values above the LLOD with a 4-fold increase in neutralizing antibody titer. Neutralization 2FR (4FR) at each pre-defined timepoint are defined as participants who had baseline values below the LLOQ with cID50 at least 2 times (4 times) above the assay LLOQ, or as participants with baseline values above the LLOQ with at least a 2-fold

(4-fold) increase in neutralizing antibody titer. While quantitative fold-rise is shown for both cID50 and cID80, response above LLOD, 2FR and 4FR responder status are shown only for cID50. (However, for superlearner analysis of multivariable CoRs, 2FR and 4FR responder status variables are included for each of pseudovirus-nAb cID50 and cID80, given the objectives of more comprehensive analysis in building the estimated optimal surrogate.)

Note that for defining positive response, 2FR, and 4FR, a reason why values below the LLOD are set to half the LLOD before calculating the indicator of response, is to ensure that a vaccine recipient that has an unusually low antibody readout at baseline and a post-vaccination value below or near the LLOD is not erroneously counted as a responder.

The following list describes the antibody variables that are measured from immunogenicity subcohort and infection case participants. (The pre-defined time points are Day 1, 29, 57.)

1. Individual anti-Spike antibody concentration at each pre-defined time point
2. Individual anti-Spike antibody fold-rise concentration post-vaccination relative to baseline at each pre-defined post-vaccination time point
3. Individual anti-RBD antibody concentration at each pre-defined time point
4. Individual anti-RBD antibody fold-rise post-vaccination relative to baseline at each pre-defined post-vaccination time point
5. Individual anti-N antibody concentration at each pre-defined time point
6. Individual anti-N antibody fold-rise post-vaccination relative to baseline at each pre-defined post-vaccination time point
7. 2-fold-rise and 4-fold rise (fold rise in anti-Spike antibody concentration [post/pre] $\geq 2$ and $\geq 4$, 2FR and 4FR) at each pre-defined post-vaccination time point
8. 2-fold-rise and 4-fold rise (fold rise in anti-RBD antibody concentration [post/pre] $\geq 2$ and $\geq 4$, 2FR and 4FR) at each pre-defined post-

vaccination time point

9. 2-fold-rise and 4-fold rise (fold rise in anti-N antibody concentration [post/pre] $\geq 2$ and $\geq 4$, 2FR and 4FR) at each pre-defined post-vaccination time point

10. Pseudovirus-nAb responders, at each pre-defined timepoint defined as participants who had baseline values below the LLOQ with detectable pseudovirus-nAb cID50 titers above the assay LLOQ or as participants with baseline values above the LLOQ with a 4-fold increase in pseudovirus-nAb cID50 titers

Summaries of the immunogenicity data will be reported in tables. In particular, the tables will include, for each pre-defined post-baseline time point:

1. For each binding antibody marker, the estimated percentage of participants defined as responders, and with concentrations $\geq$ 2x LLOQ or $\geq$ 4 x LLOQ, will be provided with the corresponding 95% CIs using the Clopper-Pearson method.

   In addition, the estimated percentage of participants defined as responders, participants with 2-fold rise (2FR), and participants with 4-fold rise (4FR) will be provided with the corresponding 95% CIs using the Clopper-Pearson method.

2. For the cID50 pseudo-virus neutralization antibody marker, the estimated percentage of participants defined as responders, participants with 2-fold rise (2FR), and participants with 4-fold rise (4FR) will be provided with the corresponding 95% CIs using the Clopper-Pearson method

3. Geometric mean titers (GMTs) and geometric mean concentrations (GMCs) will be summarized along with their 95% CIs using the t-distribution approximation of log-transformed concentrations/titers (for each of the four Spike-targeted marker types including pseudovirus-nAb cID50 and cID80, as well as for binding Ab to N).

4. Geometric mean titer ratios (GMTRs) or geometric mean concentration ratios (GMCRs) are defined as geometric mean of individual titers/concentration ratios (post-vaccination/pre-vaccination for each injection)

5. GMTRs/GMCRs will be summarized with 95% CI (t-distribution approximation) for any post-baseline values compared to baseline, and post-Day 57 values compared to Day 57
6. The ratios of GMTs/GMCs will be estimated between groups with the two-sided 95% CIs calculated using t-distribution approximation of log-transformed titers/concentrations [the groups compared are vaccine recipient Non-Cases vs. vaccine recipient breakthrough cases used for Day 57 marker correlates analyses (Post Day 57 cases) and vaccine recipient Non-Cases vs. vaccine recipient breakthrough cases used for Day 29 marker correlates analyses (Intercurrent cases and Post Day 57 cases)].
7. The differences in the responder rates, 2FRs, 4FRs between groups will be computed along with the two-sided 95% CIs by the Wilson-Score method without continuity correction (Newcombe, 1998) (the groups for comparison are as described in the previous bullet).

All of the above point and confidence interval estimates will use inverse probability of antibody marker sampling weighting in order that estimates and inferences are for the population from which the whole study cohort was drawn. In two-phase sampling data analysis nomenclature, the “phase 1 ptids” are the per-protocol individuals excluding individuals with a COVID failure event or any other evidence of SARS-CoV-2 infection $<$ 7 days post Day 57 visit. The “phase 2 ptids” are then the subset of these phase 1 ptids in the immunogenicity subcohort with Day 1 and Day 29 and Day 57 Ab marker data available. Thus, marker data for the COVID endpoint cases outside the subcohort will not be used in immunogenicity analyses; these cases are excluded from immunogenicity analyses. Similarly, for Day 29 marker correlates analyses the “phase 1 ptids” are the per-protocol individuals excluding individuals with a COVID failure event or any other evidence of SARS-CoV-2 infection $<$ 7 days post Day 29. The “phase 2 ptids” are then the subset of these phase 1 ptids in the immunogenicity subcohort with Day 1 and Day 29 Ab marker data available. Thus again, marker data for the COVID endpoint cases outside the subcohort will not be used in immunogenicity analyses; these cases are excluded from immunogenicity analyses.

The estimated weight $\hat{w}_{subcohort.57x}$ is the inverse sampling probability weight, calculated as the empirical fraction (No. Day 57 phase 1 ptids / No. Day 57 phase 2 ptids) within each of the baseline strata [(vaccine, placebo) × (baseline negative, baseline positive) × (demographic strata)]. For individuals outside the phase 1 ptids, $\hat{w}_{subcohort.57x}$ is assigned the missing value code NA. All other individuals have a positive value for $\hat{w}_{subcohort.57x}$, including cases not in the subcohort. This weight is only used for case outcome-status blinded immunogenicity inferential analyses. Note that $\hat{w}_{subcohort.57x}$ is used for all immunogenicity analyses, which are based solely on the immunogenicity subcohort, for Day 1, Day 29, and Day 57 markers. (Not used for correlates analyses.)

Tables will be provided separately for (1) baseline negative individuals, (2) baseline positive individuals, (3) baseline negative individuals by subgroup defined as in Table 3, and (4) baseline positive individuals by the same subgroups as in (3). Each table will show data for all available time points and for each of the vaccine and placebo arms.

Table 3: Baseline Subgroups that are Analyzed[1].

| |
|---|
| **Age:** $< 65$, $\geq 65$ |
| **Heightened Risk for Severe COVID:** At risk, Not at risk |
| **Age x Risk for Severe COVID:** |
| $< 65$ At risk, $< 65$ Not at risk, $\geq 65$ At risk, $\geq 65$ Not at risk |
| **Sex Assigned at Birth:** Male, Female |
| **Age x Sex Assigned at Birth:** |
| $< 65$ Male, $< 65$ Male, $\geq 65$ Female, $\geq 65$ Female |
| **Hispanic or Latino Ethnicity:** Hispanic or Latino, Not Hispanic or Latino |
| **Race or Ethnic Group:** |
| White Non-Hispanic[2], Black, Asian, American Indian or Alaska Native (NatAmer) |
| Native Hawaiian or Other Pacific Islander (PacIsl), Multiracial, |
| Other, Not reported, Unknown |
| **Underrepresented Minority Status in the U.S.:** |
| Communities of color (Comm. of color), White[2] |
| **Age x Underrepresented Minority Status in the U.S.:** |
| Age $\geq 65$ Comm. of color, Age $< 65$ Comm. of color, Age $\geq 65$ White, Age $\geq 65$ White |

[1]All analyses are done within strata defined by randomization arm and baseline positive/negative status, such that these variables are not listed here as subgroups for analysis.

[2]White Non-Hispanic is defined as Race=White and Ethnicity=Not Hispanic or Latino. All of the other Race subgroups are defined solely by the Race variable, with levels Black, Asian, American Indian or Alaska Native, Native Hawaiian or Other Pacific Islander, Multiracial, Other, Not reported, Unknown. Communities of color is defined by the complement of being known White Non-Hispanic.

For comparing antibody levels between groups, the following groups are compared:

- Baseline negative vaccine vs. baseline negative placebo
- Baseline positive vaccine vs. baseline positive placebo
- Baseline negative vaccine vs. baseline positive vaccine
- Within baseline negative vaccine recipients, compare each of the following pairs of subgroups listed in Table 3: Age $\geq 65$ vs. age $< 65$; risk for severe COVID: at risk vs. not at risk; age $\geq 65$ at risk vs. age $\geq 65$ not at risk; age $< 65$ at risk vs. age $< 65$ not at risk; male vs. female; Hispanic or Latino ethnicity: Hispanic or Latino vs. Not Hispanic

or Latino; Underrepresented minority status: Communities of color vs. White Non-Hispanic (within the U.S.).

The entire immunogenicity analysis is done in the per-protocol cohort with Day 1, Day 29, and Day 57 marker data available (the two-phase sample).

###### 6.1.2 Graphical description of antibody marker data

The Day 1, 29, 57 antibody marker data collected from the immunogenicity subcohort participants will be described graphically. These data are representative of the entire study cohort. Importantly, only antibody data from the immunogenicity subcohort are included (i.e., no data from cases outside the subcohort are included). This makes the analyses unsupervised (independent of case-control status), enabling interrogation and optimization of the antibody biomarkers prior to the inferential correlates analyses.

Plots are developed for the following purposes. All of the analyses are done separately within each of the four subgroups defined by randomization arm cross-classified with baseline negative/positive status. In addition, many of the descriptive analyses will also be done separately for each demographic subgroup of interest listed above. For descriptive plots of individual marker data points that pool over one or more of the baseline strata subgroups, plots show all observed data points.

For each antibody marker readout, both Day 57 and baseline-subtracted Day 57 readouts are of interest. We will refer to the latter as 'delta.' All readouts, including delta, will be plotted on the $\log_{10}$ scale, with plotting labels on the natural scale. As such, delta is $\log_{10}$ fold-rise in the marker readout from baseline.

The following descriptive graphical analyses are done.

1. The distribution of each antibody marker readout at Day 1, Day 29, and Day 57 will be described with plots of empirical reverse cumulative distribution functions (rcdfs) and boxplots (including individual data points) within each of the four groups defined by randomization arm (vaccine, placebo) and baseline positivity stratum (seronegative, seropositive). In-

verse probability of sampling into the subcohort weights ($\hat{w}_{subcohort.57x}$) are used in the estimation of the rcdf curves; henceforth we refer to these weights as "inverse probability of sampling" (IPS) weights. Analyses of Day 1 markers always pool across vaccine and placebo recipients given that the two subgroups are the same at baseline.

2. Plots are arranged to compare each Day 29 or Day 57 marker readout between randomization arms within each of the baseline seropositive and baseline seronegative subgroups.

3. Plots are also arranged to compare each Day 29 or Day 57 marker readout between baseline serostatus groups within each randomization arm.

4. The correlation of each antibody marker readout among Day 1, Day 29, and Day 57, and between Day 1 and fold-rise to Day 29 and to Day 57 (delta), is examined within each randomization arm and baseline positivity stratum. Pairs plots/scatterplots will be used, annotated with baseline strata-adjusted Spearman rank correlations, implemented in the PResiduals R package available on CRAN. For calculating the correlation within each randomization arm and baseline positivity stratum, because PResiduals does not currently handle sampling weights, the correlation estimates are computed as follows: For each re-sampled data set in the second approach to graphical plotting, the covariate-adjusted Spearman correlation is calculated. The average of the estimated correlations across re-sampled data sets is reported.

5. The correlation of each pair of Day 1 antibody marker readouts are compared within each baseline positivity stratum, pooling over the two randomization arms. Pairs plots/scatterplots and baseline-strata adjusted Spearman rank correlations are used, with covariate-adjusted Spearman rank correlations computed as described above. The same analyses are done for each pair of Day 29 antibody marker readouts and for each pair of Day 57 antibody marker readouts.

6. Point estimates of Day 57 marker positive response rates for each randomization arm within each baseline positivity stratum are provided. The point and 95% CI estimates include all of the data and use IPS

weights. The same analyses are done for Day 29 marker positive response rates.

##### 6.2 Methods for Positive Response Calls for bAb and nAb Assays

As noted above, binding antibody responders at each pre-defined timepoint are defined as participants with concentration above the specified positivity cut-off, with a separate cut-off for each antigen Spike, RBD, N (10.8424, 14.0858, and 23.4711, respectively, in IU/ml). This approach is used for each of the Spike and RBD and N protein antigen targets.

Pseudovirus neutralization responders at each pre-defined timepoint are defined as participants who had baseline cID50 values below the LLOD with detectable cID50 neutralization titer above the assay LLOD, or as participants with baseline values above the LLOD with a 4-fold increase in neutralizing antibody titer. Otherwise a value is negative for pseudovirus neutralization. The same approach is used based on cID80 titer.

##### 6.3 SARS-CoV-2 Antigen Targets Used for bAb and nAb Markers

The homologous vaccine strain antigens are used for the immune correlates analyses for the bAb markers, whereas the homologous vaccine strain with D614G mutation is used for the pseudovirus nAb markers.

##### 6.4 Score Antibody Markers Combining Information Across Individual bAb and/or nAb Readouts

For each time point Day 29 and Day 57 separately, score antibody markers that combine information across the five individual markers are defined and included in the multivariable CoR machine learning analyses. In particular, five score variables are studied:

1. Maximum signal-diversity score calculated as described in He and Fong (2019).
2. First two linear principal components PCA1 and PCA2

3. Nonlinear extensions of principal components FSDAM1 and FSDAM2 calculated as in Fong et al. (2020).

The purpose of these score markers is to seek to maximally capture the main immune response signal and to study whether there are more than one distinct signals that are associated with the COVID outcome, and to study whether score markers can provide strengthened association with COVID compared to the individual assay markers. The score markers are included as input features in the machine learning (superlearning) prediction modeling (multivariable CoR objective).

#### 7 Baseline Risk Score (Proxy for SARS-CoV-2 Exposure)

The list of baseline covariates potentially relevant for SARS-CoV-2 exposure and risk of COVID was specified (Table S4 in Supplementary Material). Based on these covariates, a baseline risk score is developed and controlled for in correlates analyses to adjust for potential confounding. The risk score is developed using placebo arm data only, restricting to baseline negative per-protocol placebo recipients. The risk score is defined as the logit of the predicted outcome probability from a regression model estimated using the ensemble algorithm superlearner (i.e. stacking), where this logit predicted outcome is scaled to have empirical mean zero and empirical standard deviation one. The settings of superlearner (i.e., loss function, cross-validation technique, library of learners) that are used for implementation of superlearner for building a baseline risk score are described in Section 9.5. For predictive modeling of the COVID endpoint, cases are COVID endpoints starting 7 days post Day 57 visit and non-cases are participants with follow-up beyond 7 days post Day 57 visit and never registered a COVID endpoint.

Independent of the superlearner risk score, important individual risk factors are also specified for inclusion as adjustment factors in correlates analyses. In particular, in addition to the risk score the at-risk indicator and the communities of color indicator are adjusted for in all correlates analyses. This choice is justified by the epidemiological data showing that these two indicators are strong infection and COVID-19 risk factors, and making use of the flexibility of super learner to develop a model for how age relates to risk.

Henceforth we refer to the baseline variables that are adjusted for in correlates analyses as "baseline factors" which, depending on the risk score results and performance, will consist of only the individual key risk factors, or key individual risk factors plus the baseline risk score.

### 8 Correlates Analysis Descriptive Tables by Case/Non-Case Status

The key table summarizing the distribution of each of the five antibody markers at the Day 1, 29, and 57 times points is listed below. For each time point Day 1, Day 29, and Day 57 separately, the positive response rate with 95% CI, and the GMT or GMC with 95% CI, is reported for each of of the case and non-case groups. In addition, the point and 95% CI estimate of the difference in positive response rate (non-cases vs. cases) and the GMT or GMC ratio (non-cases/cases), is reported. Two cases vs. non-cases comparisons are done: Post Day 57 cases vs. Non-cases, and Intercurrent + Post Day 57 cases vs. Non-cases, with Post Day 57 cases and Intercurrent cases defined below. The same set of non-cases is used in each comparison.

- Immunogenicity table: Antibody levels in the baseline SARS-CoV-2 negative per-protocol cohort (vaccine recipients). Post Day 57 cases are baseline negative per-protocol vaccine recipients with the symptomatic infection COVID-19 primary endpoint diagnosed starting 7 days after the Day 57 study visit. Intercurrent cases are baseline negative per-protocol vaccine recipients with the symptomatic infection COVID-19 primary endpoint diagnosed starting 7 days after the Day 29 study visit and before 7 days post Day 57 study visit. Non-cases/Controls are baseline negative per-protocol vaccine recipients sampled into the immunogenicity subcohort with no COVID primary endpoint up to the time of data cut and no evidence of SARS-CoV-2 infection up to six days post Day 57 visit.

The point and confidence interval estimates are computed using inverse probability sampling weights $\hat{w}_{subcohort.57x}$ for Post Day 57 cases and for Non-cases, and using $\hat{w}_{29.x}$ for Intercurrent + Post Day 57 cases combined, as defined in Section 9.3.1.

### 9 Correlates of Risk Analysis Plan

This analysis plan for CoRs and CoPs focuses on the COVID primary endpoint, with its continuous failure times (failure time defined by the day of the event) and no competing risks.

#### 9.1 CoR Objectives

The following CoR objectives are assessed in baseline seronegative per-protocol vaccine recipients:

1. **Univariable CoR** To assess each individual Day 29 and Day 57 antibody marker as a CoR of outcome in vaccine recipients, adjusting for baseline factors (See Section 7)
2. **Multivariable CoR** To build models predictive of outcome based on a set of Day 29 and Day 57 antibody marker readouts, adjusting for baseline factors (See Section 7)

#### 9.2 Outline of the Set of CoR Analyses

The univariable CoR objective is addressed by Cox proportional hazards regression and nonparametric threshold regression. The multivariable CoR objective is addressed by superlearning. All of these analyses are implemented in automated and reproducible press-button fashion.

In addition, supportive exploratory analyses of the univariable CoR objective are conducted using flexible parametric regression modeling: generalized additive model regression.

#### 9.3 Day 29 and Day 57 Markers Assessed as CoRs and CoPs

The following four markers at Day 29 and at Day 57 are assessed as CoRs and CoPs, usually as quantitative variables and in some analyses as ordered trinary variables or binary variables, all of which do not subtract Day 1 (baseline) values:

1. binding Ab to Spike (IgG IU/ml)
2. binding Ab to RBD (IgG IU/ml)
3. pseudovirus neutralization cID50
4. pseudovirus neutralization cID80

For all univariable CoR analyses (first objective), the non-baseline subtracted versions of the Day 29 and Day 57 antibody markers are studied; the baseline-subtracted versions are not studied given that the analyses are done in the baseline negative cohort for which Day 1 readouts will generally be negative. The multivariable machine learning CoR analyses include synthesis markers that combine information across the individual markers listed above, as well as including 2FR and 4FR versions of variables.

###### 9.3.1 Inverse probability sampling weights used in CoR analyses

In section 6.1, estimated inverse probability sampling (IPS) weights $\hat{w}_{subcohort.57x}$ were defined for per-protocol immunogenicity subcohort members, for the purpose of immunogenicity analyses. This section describes the two IPS weights, one used for Day 57 marker correlates analyses ($\hat{w}_{57.x}$) and the other used for Day 29 marker correlates analyses ($\hat{w}_{29.x}$).

Consider the correlates analyses of Day 57 markers. For baseline sampling stratum $x$ [(vaccine, placebo) $\times$ (demographic strata)], the IPS weight $w_{57.x}$ assigned to a non-case participant in stratum $x$ is defined by $\hat{w}_{57.x} = 1/\hat{\pi}_{57}(x) = N_x/n_x$, where $N_x$ is the number of stratum $x$ vaccine recipient non-cases in the Per-Protocol Baseline Negative (PPBN) cohort and $n_x$ is the number of these participants that also have Day 1, 29, and 57 marker data available, where participants with any evidence of SARS-CoV-2 infection before 7 days post Day 57 visit are excluded from the counts $N_x$ and $n_x$. For non-case participant $i$ in the immunogenicity subcohort, $\hat{w}_{57.i} = 1/\hat{\pi}_{57}(X_i)$ denotes the weight $\hat{w}_{57.x}$ for this individual's sampling stratum. All Post Day 57 cases are assigned sampling weight $N_1/n_1$ where $N_1$ is the total number of vaccine recipient cases in the PPBN cohort restricting to cases with event time starting 7 days post Day 57, and $n_1$ is the number of these participants that also had the Day 1, 29, and 57 markers measured, and again participants with any evidence of SARS-CoV-2 infection $<$ 7 days post Day 57 visit are excluded from the counts $N_x$ and $n_x$.

In terms of two-phase sampling data analysis nomenclature, for the Day 57 marker analyses "phase 1 ptids" are defined as the entire PPBN cohort except excluding participants with any evidence of SARS-CoV-2 infection $<$ 7 days post Day 57 visit. The "phase 2 ptids" are then the subset of these phase 1 ptids with Day 1, 29, and 57 Ab marker data available. Thus the weight $\hat{w}_{57.x}$ is the inverse sampling probability weight, calculated as the empirical fraction (No. phase 1 ptids / No. phase 2 ptids) within each of the baseline negative strata (14 strata defined by PPBN vaccine group cases, PPBN placebo group cases, PPBN vaccine group non-cases divided into the 6 demographic strata, and PPBN placebo group non-cases divided into the 6 demographic strata). For baseline negative individuals outside the phase 1 ptids, $\hat{w}_{57.x}$ is assigned the missing value code NA. All other individuals have a positive value for $\hat{w}_{57.x}$.

Next consider the correlates analyses of Day 29 markers. For baseline sampling stratum $x$ [(vaccine, placebo) $\times$ (demographic strata)], the IPS weight $w_{29.x}$ assigned to a non-case participant in stratum $x$ is defined by $\hat{w}_{29.x} = 1/\hat{\pi}_{29}(x) = N_x/n_x$, where $N_x$ is the number of stratum $x$ vaccine recipient non-cases in the PPBN cohort and $n_x$ is the number of these participants that also have Day 1 and Day 29 marker data available, where participants with any evidence of SARS-CoV-2 infection before 7 days post Day 29 visit are excluded from the counts $N_x$ and $n_x$. For non-case participant $i$ in the immunogenicity subcohort, $\hat{w}_{29.i} = 1/\hat{\pi}_{29}(X_i)$ denotes the weight $\hat{w}_{29.x}$ for this individual's sampling stratum. All Intercurrent and Post Day 57 cases are assigned sampling weight $N_1/n_1$ where $N_1$ is the total number of vaccine recipient cases in the PPBN cohort restricting to cases with event time starting 7 days post Day 29, and $n_1$ is the number of these participants that also had the Day 1 and Day 29 markers measured, and again participants with any evidence of SARS-CoV-2 infection $<$ 7 days post Day 29 visit are excluded from the counts $N_x$ and $n_x$.

In terms of two-phase sampling data analysis nomenclature, for the Day 29 marker analyses "phase 1 ptids" are defined as the entire PPBN cohort except excluding participants with any evidence of SARS-CoV-2 infection $< 7$ days post Day 29 visit. The "phase 2 ptids" are then the subset of these phase 1 ptids with Day 1 and Day 29 Ab marker data available. Thus the weight $\hat{w}_{29.x}$ is the inverse sampling probability weight, calculated as the empirical fraction (No. phase 1 ptids / No. phase 2 ptids) within each of the baseline negative strata (14 strata defined by PPBN vaccine group cases, PPBN placebo group cases, PPBN vaccine group non-cases divided into the 6 demographic strata, and PPBN placebo group non-cases divided into the 6 demographic strata). For baseline negative individuals outside the phase 1 ptids, $\hat{w}_{29.x}$ is assigned the missing value code NA. All other individuals have a positive value for $\hat{w}_{29.x}$. In sum, the weights $\hat{w}_{29.x}$ are calculated in the same way as the weights $\hat{w}_{57.x}$, except the relevant time window for evidence of infection or COVID is at least 7 days post Day 29 visit instead of at least 7 days post Day 57 visit.

###### 9.3.2 Choice of regression methods

Time-to-event methods of Day 57 marker correlates analyses use the Day 57 visit date as the time origin. Similarly, time-to-event methods of Day 29 marker correlates analyses use the Day 29 visit date as the time origin.

The IPWCC Cox regression model designed for case-cohort sampling designs will be used for estimation and inference on hazard ratios of outcomes by Day 29 or Day 57 marker levels, and for estimation and inference on marginalized marker-conditional cumulative incidence over time. The models will be fit using the *survey* R package available on CRAN, and will adjust for the baseline factors. We use a method from the survey package that assumes without replacement two-phase sampling and not Bernoulli sampling, which matches the sampling design and approach to weight estimation (Lumley, 2010).

The final time point $t_F$ of follow-up for correlates analyses is taken to be the latest COVID outcome event time. Let $T$ be the failure time, $S$ a Day

29 or Day 57 marker of interest, and $X$ the vector of baseline factors that are adjusted for. With $S_1(t|s,x) = P(T > t|S = s, X = x, A = 1)$, the Cox model fit yields an estimate of $S_1(t|s, X_i)$ for each individual $i$ in the phase-two sample. The marginalized conditional risk $risk_1(t|s) = E_X[P(T \leq t|s, X, A = 1)]$ through time $t$ (for all times $t$ through $t_F$ simultaneously) is estimated based on the equation

$$risk_1(t|s) \;=\; \int (1 - S_1(t|s,x))dH(x) \tag{1}$$

where $H(\cdot)$ is the distribution of $X$ in $A = 1$ individuals.

The function $risk_1(t|s)$ can be estimated by

$$\widehat{risk}_1(t|s) = \frac{\sum_{i=1}^{n} \frac{1}{\hat{\pi}(X_i)}(1 - \hat{S}_1(t|s, X_i))}{\sum_{i=1}^{n} \frac{1}{\hat{\pi}(X_i)}}, \tag{2}$$

where $n$ is the number of participants with phase-two data.

The bootstrap is used to obtain 95% pointwise confidence intervals for $risk_1(t_F|s)$.

The bootstrap process will be performed by resampling with replacement the subjects within the subcohort and the subjects outside the subcohort separately within each stratum and by resampling with replacement subjects with undetermined stratification variables. Across all bootstrap samples, the number of participants in each stratum in the immunogenicity subcohort remains fixed, but the number of cases does not stay the same.

The results of the above Cox modeling will be output in a variety of ways:

1. Plot $\widehat{risk}_1(t_F|s)$ vs. $s$ with 95% CIs for continuous $S = s$ varying over its whole range. Include on the plot the estimate of $\widehat{risk}_0(t_F)$ with a 95% CI for the placebo arm (horizontal bands), computed by a Cox model marginalizing over the same baseline factors as for the analysis of the vaccine arm.

2. Based on a fit of the Cox model to a nominal categorical antibody marker defined as the tertiles of $S$, plot $\widehat{risk}_1(t|s)$ for each category of $S$ values with 95% CIs, for all time points $t$ from Day 57 through $t_F$. If more

than 20% of vaccine recipients have $S$ below the LLOD of the assay, then the categories instead will be (1) values $\leq$ LLOD; (2) values below the median of values $>$ LLOD; (3) values above the median of values $>$ LLOD. Include on the plot the estimated curve $\widehat{risk}_0(t)$ with 95% CIs for the placebo arm, computed by a Cox model marginalizing over the same baseline factors as for the analysis of the vaccine arm.

3. Tabular reporting of the hazard ratio per 10-fold change in the quantitative Day 29 or Day 57 antibody marker with 95% confidence interval and 2-sided p-value.

4. Tabular reporting of the hazard ratio for the Middle and Upper categories of the categorical Day 57 antibody marker vs. the Lower category, with 95% confidence interval and 2-sided p-value, as well as a global generalized Wald two-sided p-value for whether the hazard rate of the endpoint varies across the three categories. The table includes the attack rate (with no. of cases / no. at risk) through $t_F$ for each of the three vaccine marker subgroups and for the placebo arm.

5. Report point and 95% CI estimates for the hazard ratio per 10-fold change in the Day 29 or Day 57 antibody marker, for the entire per-protocol baseline negative vaccine cohort and for each of the baseline demographic strata subgroups defined in Table 3 (reported via forest plotting).

6. Westfall-Young (1997) q-values and FWER-adjusted p-values for the generalized Wald tests are included in the table.

The bootstrap is used to calculate 95% pointwise CIs for $risk_1(t_F|s)$ in $s$. The 2-sided Wald p-value for testing the regression coefficient of the marker in the Cox model provides a valid test of the null hypothesis $H_0 : risk_1(t_F|s) = risk_1(t_F)$ for all $s$, and is reported.

In addition, the same Cox model analysis will be used to estimate the alternative marginalized conditional risk parameter defined by $risk_1(t|S \geq s)$ where

$risk_1(t|S \geq s) = E_X[P(T \leq t|S \geq s, X, A = 1)]$, which can be estimated by

$$\widehat{risk}_1(t|S \geq s) = \frac{\sum_{i=1}^{n} \frac{1}{\hat{\pi}(X_i)}(1 - \hat{S}_1(t|S \geq s, X_i))}{\sum_{i=1}^{n} \frac{1}{\hat{\pi}(X_i)}}.$$

This parameter is useful because typically subgroups of interest are defined by having marker response above a threshold. We will plot $\widehat{risk}_1(t_F|S \geq s)$ vs. $s$ with 95% CIs for continuous $S$ with $s$ varying over the range of $S$ in which the number of cases to estimate $\hat{S}_1(t|S \geq s, X_i)$ is 5 or more. This type of analysis is also included because it analyzes the same parameter as the nonparametric threshold estimation method described below, providing a way to address the threshold question both by Cox modeling and by nonparametric analysis.

###### 9.3.3 Univariate CoR: Nonparametric threshold regression modeling

The van der Laan et al. (2021) extension of the nonparametric CoR threshold estimation method of Donovan et al. (2019) is applied to each of the five non-baseline subtracted antibody markers, at each time point Day 29 and Day 57, using the version that defines the binary outcome $Y$ of interest as $Y = 1$ if a COVID endpoint occurred during the blinded period of follow-up and $Y = 0$ otherwise. The analyses adjust for the same baseline factors $X$ as used in the Cox model CoR analyses.

The extension adjusts for baseline covariates by estimating the conditional mean function $E[Y|S \geq s, X, A = 1]$ using discrete-SuperLearner and then empirically averaging over the baseline covariates $X$ to estimate the marginal risk $risk_1^Y(S \geq s) = E_X[P(Y = 1|S \geq s, X, A = 1)]$ for each threshold $s$ of the the antibody marker in a specified discrete set. We do not perform pooled regression across the thresholds $s$, which ensures we are totally nonparametric in estimating the threshold dependence of $risk_1^Y(S \geq s)$ on $s$. The SuperLearner library includes a range of increasingly flexible parametric learners including logistic regression (glm), bayesian logistic regression (bayesglm), and L1-penalized logistic regression (glmnet). (Two of each learner is included in the library, one with only main-term variables and another with main-term and interaction variables.) An advantage of the nonparametric CoR threshold method compared to Cox modeling that specifies a log linear hazard ratio with the marker is that it can potentially detect a threshold of very low risk. The method is implemented with and without the monotonicity constraint that $risk_1^Y(S \geq s)$ is monotone non-increasing in $s$, where the results assuming monotonicity are reported unless there is evidence for violation of this assumption.

The results are reported in the same way that Donovan et al. (2019) reports results in its Figure 2, where point estimates, pointwise 95% confidence bands, and simultaneous 95% confidence bands for $risk_1^Y(S \geq s)$ are plotted for a range of threshold values. The simultaneous confidence bands cover the entire curve in $s$ with at least 95% probability and are useful for judging whether risk varies over threshold subgroups, whereas the pointwise 95% confidence bands are useful for quantifying precision at particular threshold values. The method uses the same empirical two-phase sampling estimated weights (IPS weights) as used for the other univariable IPWCC CoR analyses. In addition, for each pre-specified risk threshold $c$ set to take values over a grid with lowest value 0, the method is applied to estimate the inverse function $s_c = inf\{s : E_X[P(Y = 1|S \geq s, A = 1, X] \leq c\}$, where $s_c$ is estimated by substitution of the marginal risk function estimate. Note that the substitution estimator of $s_c$ requires that the marginal risk function is estimated for all thresholds, which is computationally infeasible. Instead, we estimate the marginal risk function on a sufficiently large discrete set and linearly interpolate to obtain marginal risk estimates for all thresholds outside the discrete set. In order for this estimand to be well defined, we operate (for this estimand only) under the assumption that $s \mapsto risk_1^Y(S \geq s)$ is monotone. For the substitution-based estimator of the inverse function $s_c$ to be well-defined, we require the estimate of $s \mapsto risk_1^Y(S \geq s)$ to be monotone as well. If there is evidence that the function estimate is not monotone then we replace the estimate with its monotone projection, which preserves its theoretical properties (Westling, van der Laan, Carone, 2020).

A plot of point and pointwise 95% confidence interval estimates of $s_c$ (over the grid of $c$ values) is provided to help indicate marker thresholds defining subgroups with very low risk of outcome. The confidence interval estimates for $s_c$ are derived directly from the confidence interval estimates for the marginal risk function $s \mapsto risk_1^Y(S \geq s)$, and therefore its estimates are compatible with those of the marginal risk function. In addition, a plot of point and simultaneous 95% confidence interval estimates of $s_c$ (over the grid of $c$ values) is provided, where the simultaneous confidence interval estimates for $s_c$ are derived directly from the simultaneous 95% confidence band estimates for the marginal risk function $s \mapsto risk_1^Y(S \geq s)$, and therefore its estimates are compatible with those of the marginal risk function. In particular, no multiple testing adjustments are needed.

The analysis is done using targeted maximum likelihood estimation (TMLE) as described in van der Laan, Zhang, and Gilbert (2021), and the pointwise and simultaneous simultaneous confidence bands are of the Wald-type, obtained from the asymptotic distribution of the TMLE.

##### 9.4 Univariable CoR: Supportive Exploratory Flexible Parametric Risk Modeling

For each of the four non-baseline subtracted Day 57 antibody markers, flexible nonlinear modeling of outcome risk studied as a dichotomous outcome $Y$ will be conducted, as exploratory supportive analyses. Again, the analyses adjust for the same baseline factors $X$ as used in the Cox model CoR analyses. A generalized additive model with degree of smoothing estimated by cross-validation is employed (Wood, 2017). Two-phase sampling designs are accounted for through inverse probability weighting and confidence intervals are obtained through the same bootstrap scheme as the Cox proportional hazard model bootstrap inference.

###### 9.4.1 P-values and Multiple hypothesis testing adjustment for CoR analysis

In general, p-values are only reported from pre-specified and automated (press-button) analyses. For the CoR analyses, p-values are reported for the univariable Cox regression analyses of the four specified Day 57 antibody marker variables. Two-sided p-values for hypothesis testing of a Day 57 marker CoR are calculated both for the Cox regression of quantitative markers (two-sided Wald tests), and for the Cox regression of markers binned into tertiles (two-sided Generalized Wald tests). Therefore a total of eight 2-sided p-values for Day 57 CoRs are calculated.

It is not completely clear whether to perform multiple hypothesis testing adjustment, given the expectation that the correlations among the markers are high, and possibly very high, meaning that multiplicity correction could incur a relatively high cost on the false negative error rate. However, given that robust evidence supporting an antibody marker as a CoR will be required for qualifying a marker, we will conduct multiplicity adjustment for CoR analysis, as the ability to make an inference that a marker passed pre-specified multiplicity adjusted criteria should aid an overall evidence package for establishing a validated or non-validated surrogate endpoint. Therefore, multiplicity adjustment is performed across the eight 2-sided p-values.

A permutation-based method (Westfall et al., 1993) will be used for both family-wise error rate (Holm-Bonferroni) and false-discovery rate (q-values; Benjamini-Hochberg) correction. $10^4$ replicates of the data under the null hypotheses will be created by randomly resampling the immunologic biomarkers with replacement. For each Cox regression CoR analysis the unadjusted p-value, the FWER-adjusted p-value, and the q-value is reported for whether there is a covariate-adjusted association, where all p-values and q-values are 2-sided. The FWER-adjusted p-values and q-values are computed pooling over both the quantitative marker and tertilized marker CoR analyses. As a guideline for interpreting CoR findings, markers with FWER-adjusted p-value $\leq 0.05$ are flagged as having statistical evidence for being a CoR. Additionally, markers with unadjusted p-value $\leq 0.05$ and q-value $\leq 0.10$ are flagged as having a hypothesis generated for being a CoR.

The multiplicity adjustment analyses described above for Day 57 marker CoR analyses are repeated (conducted separately) for Day 29 marker CoR analyses.

##### 9.5 Multivariable CoR: Superlearning of Optimal Risk Prediction Models

###### 9.5.1 Objectives

The multivariable CoR objective is addressed through two sub-objectives: first to build an 'estimated optimal surrogate' (Price et al., 2018), a model that best predicts the outcome from Day 57 antibody markers and baseline factors. The second sub-objective is estimation and inference on variable importance measures for each Day 57 antibody marker, for ranking of antibody markers by their importance/influence on predicting risk. The analysis plan is patterned off of the analysis of the HVTN 505 HIV-1 vaccine efficacy trial (Neidich et al., 2019). This objective also builds models for predicting outcome from Day 29 antibody markers and baseline factors, and from Day 29 antibody markers, Day 57 antibody markers, and baseline factors. For these analyses both baseline-subtracted and non-baseline subtracted versions of the Day 29 and Day 57 markers are used, in a broader unbiased analysis to build models most predictive of outcome.

###### 9.5.2 Input variable sets

Day 57 antibody markers are classified into the following three antibody marker variable sets, with individual variables listed within categories:

1. Binding antibody anti-Spike (S-bAb)
   a Day 57 anti-Spike IgG concentration
   b delta (Day 57 - Day 1) anti-Spike IgG concentration
   c indicator 2FR anti-Spike IgG concentration
   d indicator 4FR anti-Spike IgG concentration
2. Binding antibody anti-RBD (RBD-bAb)
   a Day 57 anti-RBD concentration
   b delta (Day 57 - Day 1) anti-RBD concentration
   c indicator 2FR anti-RBD concentration
   d indicator 4FR anti-RBD concentration
3. Pseudovirus neutralizing antibody anti-Spike (pseudovirus-nAb)
   a Day 57 anti-Spike cID50
   b Day 57 anti-Spike cID80

c delta (Day 57 - Day 1) anti-Spike cID50

d delta (Day 57 - Day 1) anti-Spike cID80

e indicator 2FR anti-Spike cID50

f indicator 4FR anti-Spike cID50

g indicator 2FR anti-Spike cID80

h indicator 4FR anti-Spike cID80

A second set of antibody marker variable sets is defined by replacing Day 57 above with Day 29. In addition, a third set of antibody marker variable sets is defined by replacing Day 57 antibody markers with both Day 29 and Day 57 antibody marker variables. Inclusion of these sets allow comparing classification accuracy of Day 29 markers vs. Day 57 markers, and whether including both time points improves classification accuracy.

The baseline factors without any marker data constitutes another set of variables to include in the superlearner modeling.

###### 9.5.3 Missing data

We expect a very small amount of missing data from the four antibody marker types (bAb Spike, RBD; pseudovirus-nAb cID50, cID80). However, there may be a small amount of missing data, with possibly different participants missing data for different markers. We take the following approach to handle any missing data that occurs.

First, we define the two-phase sampling indicator $\epsilon$ as taking value of one if a participant has Day 1 and Day 29 and Day 57 bAb data for both Spike and RBD, where here we assume that the MSD platform is highly robust such that it will have nearly 100% complete data for sampled participants. Second, for the other two marker types (pseudovirus-nAb cID50, cID80), for participants with $\epsilon = 1$ but the Day 1 and/or Day 29 and/or Day 57 marker value is missing, we use single imputation to fill in any missing values, ignoring the uncertainty in the imputations in the analysis, because it should have negligible impact on results given the (very) small amount of missing data.

Multiple linear regression will be used to impute missing values, separately for each antibody marker, based on the set of individuals with that antibody marker measured at Day 1, Day 29, and Day 57. Accurate imputations are possible given the high correlations of the markers, especially between cID50 and cID80 within the same immunoassay. This process means that the two-phase data set has a simple 'all-or-nothing' missing data pattern where participants with $\epsilon = 1$ have all markers with Day 1 and Day 29 and Day 57 data, and are included in IPWCC analyses, and participants with $\epsilon = 0$ have some or all markers missing and are excluded from IPWCC analyses. This means that all IPWCC data analyses can use the same empirical frequency (IPS) sampling weights, separately for correlates analyses of Day 29 markers and of Day 57 markers.

For analysis methods that use the whole cohort (phase-one plus phase-two data), the same phase-two data as described above are used. If some of the phase-one baseline factors that are adjusted for variables are missing with only a small amount of missing values, then single imputation will be used to fill in the values, and, as for the immunologic marker imputations, the uncertainty in the imputations will be ignored in the analyses. Simple average values will be used to fill in baseline covariate missing values of the baseline factors.

###### 9.5.4 Implementation of superlearner

For baseline risk score development, Superlearner is applied to the placebo arm only, as mentioned in Section 7. For multivariable immune correlates of risk/estimated optimal surrogate development, Superlearner is applied to the vaccine arm only. The following details are used in the implementation of superlearner of the vaccine arm only:

- Pre-scale each quantitative and ordinal variable to have empirical mean 0 and standard deviation 1.
- For the immune correlates analysis, the final library of learners is selected accounting for the number of phase-two endpoint cases in the vaccine arm. If the number of cases is limited, at or near 25 evaluable

endpoint cases, then the modeling will only allow learning algorithms to have a maximum of 5 antibody marker variables, and will use leave-one-out cross-validation and the negative log-likelihood loss function, a combination that tends to provide good performance in small sample size settings. This approach was used for the Moderna COVE trial given the numbers of endpoints.

- Include learning algorithms with and without screening of variables. Screens used will be: 1) glmnet (lasso) pre-screening (with default tuning parameter selection), 2) logistic regression univariate 2-sided p-value screening (at level $p < 0.10$), and 3) high-correlation variable screening (described below). The adaptive algorithms (SL.randomForest, SL.xgboost, SL.gam, SL.polymars) are only used with these screens, given that the limited number of endpoint cases may challenge use of these methods with no variable screening. Moreover, the adaptive algorithms are not used if there are only 25 (or close to it) endpoint cases, which is the case for the Moderna COVE trial. All of the selected learners are coded into the SuperLearner R package available on CRAN.

- Include high-correlation variable screening, not allowing any pair of input variables to have Spearman rank correlation $r > 0.9$. When a pair of variables has $r > 0.9$, the variable with the highest ranked signal-to-noise ratio (i.e., biological dynamic range) is selected; if these data are not available (they are not for Moderna COVE) or there is a tie then variables are selected in the following order of priority: first pseudovirus-nAb, then bAb. Given than the Spike and RBD variables have $r > 0.95$ at each time point Day 29 and Day 57, any model that would consider both Spike and RBD includes only Spike. Similarly, given than the PsV cID50 and PsV cID80 variables have $r > 0.95$ at each time point Day 29 and Day 57, any model that would consider both PsV cID50 and PsV cID80 includes only PsV cID50.

- The superlearner is conducted averaging over 10 random seeds, to make results less dependent on random number generator seed.

- All of the learners are implemented with IPS weighting, using the weights

$\hat{w}_{57.x}$ defined in Section 9.3.1 to account for the two-phase sampling design. Note that these weights are used even for models that include Day 29 markers but not Day 57 markers, because only Post Day 57 cases (starting 7 days post Day 57 visit) are included in the multivariable CoR analyses.

- Two levels of cross-validation are used:
  - Outer level: CV-AUC computed over 5-fold cross-validation repeated 10 times to improve stability
  - Inner level: leave-one-out CV used to estimate ensemble weights (if $n_v$ is near 25) and 5-fold CV if $n_v$ is larger. (For Moderna COVE leave-one-out is used.)
- Results for comparing classification accuracy of different models are based on point and 95% confidence interval estimates of cross-validated area under the ROC curve (CV-AUC) and difference in CV-AUC as a predictiveness metric (Hubbard et al., 2016; Williamson et al., 2020). Results are presented as forest plots of point and 95% confidence interval estimates similar to those used in Neidich et al. (2019) (Figure 3) and Magaret et al. (2019). CV-AUC is estimated using the R package *vimp* available on CRAN, including the IPS weights that are used for other data analyses.

For the baseline risk score SuperLearner analysis of the placebo arm (Section 7), the same approach is used, with the following modifications: (1) 5-fold cross-validation will be used with no more than max(20, floor($n_p$/20)) input variables included in each model, where $n_p$ is the number of evaluable placebo arm cases; (2) no IPS weighting is needed; (3) the adaptive learning algorithms are included.

Table 4 lists the learning algorithms that are applied to estimate the conditional probability of the outcome based on the input variable sets considered above. Most of the algorithms are non-data-adaptive type learning algorithms, such as parametric regression models (e.g., generalized linear models [glms]), which are simple, stable, and advantageous for an application with a limited number of endpoint events. Data-adaptive type algorithms are also included if the number of endpoint events is high enough, for increasing flexibility of modeling and reducing the risk of model misspecification: SL.ranger, SL.gam, and SL.xgboost. All of the selected learners are coded into the SuperLearner R package.

Before fitting the superlearner models to the vaccine arm data, a decision is made on how to define the "baseline risk factors" input variable set, based on prediction-accuracy results of the Superlearner analysis that built the baseline behavioral risk score based on the placebo arm, as well as on external knowledge of important individual risk factors. For Moderna COVE the baseline factors are defined as the baseline risk score, the indicator of being at heightened risk for COVID (a randomization factor), and the indicator of being a member of community of color.

For the immune correlates objective the superlearner model is fit to each of the following 28 variable sets, with immunological variables listed in Section 9.5.2:

1. Baseline risk factors
2. Baseline risk factors and the Day 57 bAb anti-Spike markers
3. Baseline risk factors and the Day 57 bAb anti-RBD markers
4. Baseline risk factors and the Day 57 pseudovirus-nAb cID50 markers
5. Baseline risk factors and the Day 57 pseudovirus-nAb cID80 markers
6. Baseline risk factors and the Day 57 bAb markers and the pseudovirus-nAb cID50 markers
7. Baseline risk factors and the Day 57 bAb markers and the pseudovirus-nAb cID80 markers
8. Baseline risk factors and the Day 57 bAb markers and the combination scores across the four markers [PCA1, PCA2, FSDAM1/FSDAM2 (the first two components of nonlinear PCA), and the maximum signal diversity score He and Fong (2019)].

9. Baseline risk factors and all individual Day 57 marker variables
10. Baseline risk factors and all individual Day 57 marker variables and all combination scores (full model of Day 57 markers)
11. Baseline risk factors and the Day 29 bAb anti-Spike markers
12. Baseline risk factors and the Day 29 bAb anti-RBD markers
13. Baseline risk factors and the Day 29 pseudovirus-nAb cID50 markers
14. Baseline risk factors and the Day 29 pseudovirus-nAb cID80 markers
15. Baseline risk factors and the Day 29 bAb markers and the pseudovirus-nAb cID50 markers
16. Baseline risk factors and the Day 29 bAb markers and the pseudovirus-nAb cID80 markers
17. Baseline risk factors and the Day 29 bAb markers and the combination scores across the four markers [PCA1, PCA2, FSDAM1/FSDAM2 (the first two components of nonlinear PCA), and the maximum signal diversity score He and Fong (2019)].
18. Baseline risk factors and all individual Day 29 marker variables
19. Baseline risk factors and all individual Day 29 marker variables and all combination scores (full model of Day 29 markers)
20. Baseline risk factors and the Day 29 and Day 57 bAb anti-Spike markers
21. Baseline risk factors and the Day 29 and Day 57 bAb anti-RBD markers
22. Baseline risk factors and the Day 29 and Day 57 pseudovirus-nAb cID50 markers
23. Baseline risk factors and the Day 29 and Day 57 pseudovirus-nAb cID80 markers
24. Baseline risk factors and the Day 29 and Day 57 bAb markers and the pseudovirus-nAb cID50 markers
25. Baseline risk factors and the Day 29 and Day 57 bAb markers and the pseudovirus-nAb cID80 markers

26. Baseline risk factors and the Day 29 and Day 57 bAb markers and the combination scores across the eight markers [PCA1, PCA2, FS-DAM1/FSDAM2 (the first two components of nonlinear PCA), and the maximum signal diversity score He and Fong (2019)].

27. Baseline risk factors and all individual Day 29 and Day 57 marker variables

28. Baseline risk factors and all individual Day 29 and Day 57 marker variables and all combination scores (full model of Day 29 and Day 57 markers)

Therefore in total, 28 variable sets are studied. The reason to include the baseline risk factors only variable set is to investigate how much incremental improvement in predicting outcome is obtained by adding antibody marker variables on top of baseline demographic/exposure factors. The other variable sets are designed to compare the three immunoassay types by their predictiveness, to compare the two pseudovirus neutralization readouts cID50 and cID80 for their predictiveness, to compare the two time points of marker measurement for their predictiveness, and to investigate incremental predictive value in using multiple immunoassays and time points. The final variable set is included as the full model that considers all variables together, which serves as another reference model.

Table 4: Learning Algorithms in the Superlearner Library of Estimators of the Conditional Probability of Outcome, for Building the Baseline Risk Score Based on the Placebo Arm[1].

| Algorithms | Screens/ Tuning Parameters |
|---|---|
| SL.mean | None |
| SL.glm | Low-collinearity and (All, Lasso, LR)[2] |
| SL.glm.interaction | Low-collinearity and (Lasso, LR) |
| SL.glmnet | (alpha=1; All) |
| SL.gam | Low-collinearity and (Lasso, LR) |
| SL.xgboost[3] | All and (maxdepth,shrinkage,balance)= (4, 0.1, no) |
| SL.ranger[3] | All and balance = no |

[1]All continuous and ordinal covariates are pre-standardized to have empirical mean 0 and standard deviation 1.
[2]**All** = include all variables; **Lasso** = include variables with non-zero coefficients in the standard implementation of SL.glmnet that optimizes the lasso tuning parameter via cross-validation; **Low-collinearity** = do not allow any pairs of quantitative variables with Spearman rank correlation $> 0.90$; **LR** = Univariate logistic regression Wald test 2-sided p-value $< 0.10$.
[3]Covariate balancing (if requested) is done using option `scale_pos_weight` in SL.xgboost and option `case.weights` in SL.ranger.

Table 5: Learning Algorithms in the Superlearner Library of Estimators of the Conditional Probability of Outcome: Simplified Library in the Event of Fewer than 50 Vaccine Breakthrough Cases for an Analysis, for Use in Multivariable CoR Analysis of Moderna COVE[1].

| Algorithms | Screens/ Tuning Parameters |
|---|---|
| SL.mean | None |
| SL.glm | Low-collinearity and (All, Lasso, LR)[2] |
| SL.glmnet | alpha=0, 1 |
| SL.xgboost | (maxdepth,shrinkage,balance[3])= (2, 0.1, yes) (2, 0.1, no) (4, 0.1, yes) (4, 0.1, no) |
| SL.ranger | balance = (yes, no) |

[1]All continuous and ordinal covariates are pre-standardized to have empirical mean 0 and standard deviation 1.
[2]**All** = include all variables; **Lasso** = include variables with non-zero coefficients in the standard implementation of SL.glmnet that optimizes the lasso tuning parameter via cross-validation; **Low-collinearity** = do not allow any pairs of quantitative variables with Spearman rank correlation $> 0.90$; **LR** = Univariate logistic regression Wald test 2-sided p-value $< 0.10$.
[3]Covariate balancing (if requested) is done using option `scale_pos_weight` in SL.xgboost and option `case.weights` in SL.ranger.

Given the class-imbalance issue, with many more non-case than case records, all of the cross-validation for the multivariable immune CoR objective is done stratified by case/non-case status.

In order to evaluate the relative performance of the superlearner estimated models for each of the 28 variable sets, derived using the learning algorithms specified in Table 4, the CV-AUC is estimated with a 95% confidence interval (Hubbard et al., 2016; Williamson et al., 2020). The point and 95% confidence interval estimates of CV-AUC are reported in a forest plot, which provide a way to discern which antibody assays and readouts/markers provide the most information in predicting COVID or other outcomes. As noted above CV-AUC is estimated using the R package *vimp* available on CRAN, which uses augmented inverse probability weighting to properly estimate CV-AUC accounting for the two-phase sampling design.

If there are fewer than 50 vaccine breakthrough cases included in a correlates analysis, then the libary of learners will be simplified to that specified in Table 5.

In addition, for selected variable sets, similar forest plots will be made comparing performance of the various estimated models (e.g., by individual learning algorithm types such as lasso), including discrete superlearner and superlearner models. The plot will be examined to determine which individual learning algorithm types are performing the best. If there is an interpretable algorithm that has performance close to the best-performing algorithm (which is most likely to be the superlearner), then it will be fit on the entire data set of vaccine recipients and the estimated model presented in a table.

Cross-validated ROC curves are plotted for the superlearner estimated models for each of the input variable sets. In addition, boxplots of cross-validated estimated probabilities of outcome by case-control status (as estimated from the superlearner models) are plotted.

#### 10 Correlates of Protection: Generalities

In general, for all of the correlate of protection analyses, the same antibody markers are assessed that were analysed as correlates of risk: the Day 29 and Day 57 antibody markers not subtracting for the Day 1 baseline readout are used. Each of the eight Day 29 and Day 57 antibody biomarkers are separately studied as CoPs by the different analysis approaches summarized below.

We describe the CoP methods for Day 57 antibody markers; the same methods are applied to Day 29 antibody markers.

#### 11 Correlates of Protection: Correlates of Vaccine Efficacy Analysis Plan

For each of the Day 57 antibody markers, the method of Gilbert, Blette, Shepherd, and Hudgens (2020) will be used to estimate $VE(1)$, $VE(0)$, and $VE(1) - VE(0)$, each with a 95% confidence interval and a 95% estimated uncertainty interval (EUI), where $VE(1)$ is vaccine efficacy for the subgroup of vaccine recipients with Day 57 marker if assigned vaccine $S(1)$ above a specified cut-point value $s_{cut}$, and $VE(0)$ is vaccine efficacy for the subgroup of vaccine recipients with Day 57 marker if assigned vaccine $S(1)$ not greater than $s_{cut}$. That is,

$$VE(1) = 1 - \frac{P(Y(1) = 1|S(1) > s_{cut})}{P(Y(0) = 1|S(1) > s_{cut})}$$
$$VE(0) = 1 - \frac{P(Y(1) = 1|S(1) \leq s_{cut})}{P(Y(0) = 1|S(1) \leq s_{cut})}$$

The analysis will be done under the **NEH** assumption ("no early harm") of Gilbert et al. (2020). The cut point is defined as the percentile equal to one minus the estimated vaccine efficacy in the primary analysis, with logic that a maximally simple version of a perfect CoP would have binary marker with $S = 1$ corresponding to protection and $S = 0$ corresponding to no protection. If the estimated vaccine efficacy is high (say 90% or higher), it is possible that this cutpoint will not yield stable results, because of sparse cells; in this situation we will repeat the analysis using two additional cut-points that creates greater balance in frequencies of $S = 1$ and $S = 0$ in the vaccine group immunogenicity subcohort: 20th and 40th percentiles. If the estimated vaccine efficacy is moderate (between 50% and 80%), we will also use the two additional cut-points the 20th and 40th percentiles. This analysis method does not require closeout placebo vaccination (CPV) (Follmann, 2006) or a good baseline immunogenicity predictor of the Day 57 antibody marker. The method is implemented using Bryan Blette's R package "psbinary" posted at his Github repository. Based on the Moderna COVE data, the analyses are done using the 8th, 20th, and 40th percentiles of markers.

A limitation of the Gilbert et al. method is that it only assesses a binary biomarker. Other analyses will be considered to estimate $VE(s)$ over biomarker values $s$ over the entire range, treating $S$ as a quantitative or categorical variable, and gaining efficiency by incorporating CPV and/or putative baseline immunogenicity predictors (BIPs). Based on earlier simulation studies (Follmann, 2006; Huang et al., 2013, e.g.,), methods that only leverage CPV data tend to have low power relative to methods that leverage BIP data alone (BIP-only methods) or both BIP and CPV data (BIP+CPV methods). Therefore, the key for improving efficiency will be the availability of a BIP. VE curve analysis for continuous $S$ will thus be conducted contingent on the availability of a BIP that satisfies the $R^2$ criterion outlined in Table 7. It is anticipated that post-crossover immune response marker data will not be available in early correlates analyses, and so BIP-only methods will be used in these initial analyses. When CPV data becomes available, new BIP+CPV analyses will be conducted that incorporate this new information. Details of the BIPs used can be found at the end of this section.

Let $Y(a)$ denote the potential binary outcome of interest if receiving intervention $a$, with $a = 1, 0$ standing for assignment to vaccine and placebo, respectively. Let $S(a)$ denote the potential biomarker value if receiving intervention $a$. The vaccine efficacy curve (Follmann, 2006; Gilbert and Hudgens, 2008) is defined as the curve of vaccine efficacy as a function of the immune response biomarker if assigned vaccination (i.e., $S(1)$): $VE(s) = 1 - P(Y(1) =$

$1|S(1) = s)/P(Y(0) = 1|S(1) = s)$. It characterizes the percentage reduction in clinical risk under vaccine assignment compared to under placebo assignment conditional on $S(1)$ and informs about the magnitude of potential immune response associated with certain levels of VE. Consider the existence of BIPs $X$ correlated with $S(1)$ and/or a CPV component in the trial where a subset of placebo recipients free of the outcome are vaccinated and have their immune response biomarkers measured as substitutes for $S(1)$. Under the NEE assumption and assuming the set of participants with $S(1)$ available is nested within the set of participants with BIP measures, the pseudo-score estimation method (Huang et al., 2013; Zhuang et al., 2019) based on discrete BIP measures allowing for adjustment of $X$ will be adopted for estimating the risk model $P(Y(z) = 1|S(1), X)$ and subsequently $VE(s) = 1 - \int P(Y(1) = 1|S(1), x)dF_X(x|S(1))/\int P(Y(0) = 1|S(1), x)dF_X(x|S(1))$. Hypothesis testing will be conducted for testing the null hypothesis that the VE curve is constant (Zhuang et al., 2019). Estimated parametric (Gilbert and Hudgens, 2008), semiparametric (Huang and Gilbert, 2011), or nonparametric (Li and Luedtke, 2020) likelihood estimators of VE curves will be applied to continuous BIPs. In scenarios where some BIPs are not measured from all trial participants, VE curve estimators accounting for this monotone missingness in $X$ and $S(1)$ will be adopted (Huang, 2018). If the data support positive vaccine efficacy before Day 57, sensitivity analysis approaches will be conducted for VE curve estimation under the NEH assumption. In the presence of multiple candidate biomarkers and when a CPV component is present, a multiple imputation approach as proposed in Dasgupta and Huang (2019) will be utilized to impute missing $S(1)$ data for selecting markers from multiple candidates and deriving a univariate marker score for VE curve estimation.

Finally, for scenarios with very rare events such that methods described above lack precision even with a CPV component but where the available BIP still satisfies the $R^2$ criterion outlined in Table 7, we will adopt sensitivity analysis methods that model the placebo risk conditional on the counterfactual $S(1)$ based on a sensitivity parameter that varies over some pre-specified range.

Among different strategies to identify BIPs, the following will be tried. First, for vector vaccines, we will study Day 1 bAb or nAb response to the vector as a BIP for the Day 57 markers of interest (not relevant for Moderna COVE). Second, we will check whether Day 1 bAb or nAb to Nucleocapsid protein is a BIP for the anti-Spike/anti-RBD Day 57 markers of interest. The rationale for this latter analysis is that some studies have shown cross-reactive responses to Nucleocapsid protein and to common circulating human coronaviruses.

We will also evaluate using a multivariate BIP that corresponds to all of these aforementioned candidate univariate BIPs, which may help to achieve the target $R^2$ (see Table 7). When doing this, a separate BIP $W$ will be used for each vaccine-induced immune response marker $S(1)$. Let $Y(a)$ be the counterfactual outcome of interest — e.g., a COVID disease endpoint by a prespecified time — if randomization assignment had been set to $A = a$. The analyses conducted will provide unbiased estimates of the estimands of interest when $Y(a) \perp W|S(1)$ for $a \in \{0, 1\}$. The BIP $W$ will be a learned function of baseline covariates $L$ — that is, $W = f(L)$ for a function $f$ that will be learned based on the available data. All available baseline covariates will be considered for inclusion in $L$, including age, BMI, and Day 1 bAb or nAb to Nucleocapsid protein. If available, measurements of prior immune response to the vaccine vector will always be included in $L$.

If the trial has more than 100 events on the vaccine arm in the subgroup of interest, then $f$ will be chosen to be an estimate of the following population-level optimization problem:

$$\begin{aligned} &\text{minimize } E[\{S - f(L)\}^2 | A = 1] \\ &\text{subject to } f(L) \perp Y | A = 1, S. \end{aligned}$$

The rationale for choosing $f$ to (approximately) solve this optimization problem is that the BIP should be maximally predictive of $S$, while also satisfying the needed conditional independence assumption $Y(a) \perp W|S(1)$ when $a = 1$. Moreover, the needed conditional independence assumption $Y(a) \perp W|S(1)$ for the case that $a = 0$ is most plausible when this assumption is also satisfied for the case that $a = 1$. Also, because $W = f(L)$ for some function $f$, $Y(0) \perp W|S(1)$ is always more plausible than $Y(a) \perp L|S(1)$.

The solution to the above optimization problem is given by:

$$f(\ell) := \theta(\ell) - \frac{E[\theta(L)r(L)]}{E[r(L)^2]} r(\ell)$$

where $\theta(\ell) := E\{S|A = 1, L = \ell\}$, $r(\ell) := \frac{m(\ell)}{E[m(L)]} - \frac{1-m(\ell)}{1-E[m(L)]}$ and $m(\ell) := E[Y|A = 1, L = \ell]$. The following strategy is used to estimate this solution:

1. Obtain an estimate $\hat{\theta}$ of the function $\theta$ by running a Superlearner of $S$ against $L$ in the vaccine arm, where inverse probability of sampling weights are used to account for two-phase sampling of the marker.
2. Obtain an estimate $\hat{m}$ of $m$ by using Superlearner to regress $Y$ against $L$ in the vaccine arm.
3. Obtain an estimate $\hat{r}$ via a plug-in estimator, where $E[m(L)]$ is estimated by taking the empirical mean of $\hat{m}(L)$.
4. The final estimate $\hat{f}$ of $f$ is given by

$$\hat{f}(\ell) := \hat{\theta}(\ell) - \frac{\hat{E}[\hat{\theta}(L)\hat{r}(L)]}{\hat{E}[\hat{r}(L)^2]} \hat{r}(\ell),$$

   where $\hat{E}$ denotes an empirical expectation.

Each Superlearner will be run using the same library and settings described in Table 6. If the trial has fewer than 100 events on the vaccine arm, then the function $f$ will be learned via Step 1 above only — that is, we will take $\hat{f} = \hat{\theta}$. All standard errors will be obtained via the bootstrap, with the above fitting of $\hat{f}$ redone within each bootstrap sample.

#### 12 Correlates of Protection: Interventional Effects

In these analyses, we seek to understand whether, how, and to what extent Day 57 antibody markers impact vaccine efficacy in causal ways. We describe three approaches to this problem. Each involves consideration of a binary counterfactual outcome $Y(a, s)$ (e.g., indicator of the COVID disease endpoint by a pre-specified time) under a hypothetical intervention that both sets randomization assignment $A = a$ and sets the Day 57 immunologic marker $S$ to a fixed value or based upon a random draw from a analyst-specified distribution. Below, we assume that $S$ is scalar-valued, but some of the approaches below naturally extend to the case where a vector of immunologic markers are considered (currently such analyses are not planned). Given the central goal to develop a parsimonious surrogate endpoint based on a single immunoassay, the main analysis will use each of the methods to assess each of the four quantitative readouts (not baseline-subtracted) separately as CoPs, adjusting for the same set of baseline covariates as used in the CoR analyses previously described in Section 9.

The current COVE immune correlates manuscript does not include correlates of vaccine efficacy analyses, given the number of vaccine breakthrough cases.

##### 12.1 CoP: Controlled Vaccine Efficacy

We first describe the controlled vaccine efficacy curve defined as

$$\text{CVE}(s) = 1 - \frac{P(Y(1,s)=1)}{P(Y(0)=1)} .$$

The value CVE($s$) takes represents the relative decrease in endpoint frequency achieved by administering vaccine and setting Day 57 immunologic marker level to $s$ compared to the placebo control intervention. Under our approach, the value of CVE($s$) is assumed to be monotone non-decreasing in $s$; in other words, vaccine efficacy can only potentially be improved by setting greater marker levels. The extent to which the marker plays a role in determining vaccine efficacy can be determined by the degree of flatness of the graph of CVE($s$) versus $s$.

In addition, because the primary study cohort for correlates analysis is naive to SARS-CoV-2, each of the Day 57 markers $S$ has no variability in the placebo arm [all values are 'negative,' below the assay lower limit of detection (LLOD)]. Therefore, advantageously in this setting $CVE(s)$ has a special connection to the mediation literature, where $CVE(s = LLOD)$ is the natural direct effect, and vaccine efficacy is 100% mediated through $S$ if and only if $CVE(s = LLOD) = 0$. Thus inference on $CVE(s = LLOD)$ evaluates full mediation.

Since $P(Y(0) = 1) = P(Y = 1 \mid A = 0)$ in view of vaccine versus placebo randomization, the controlled vaccine efficacy CVE($s$) at level $s$ can be identified using the fact that

$$P(Y(1, s) = 1) = E\left[P(Y = 1 \mid S = s, A = 1, X)\right]$$

whenever $Y(1, s)$ and $S$ are independent given $A = 1$ and a vector $X$ of covariates, and $P(S = s \mid A = 1, X) > 0$ almost surely. In other words, identification of the controlled vaccine efficacy requires that a rich enough set of covariates be available so that deconfounding of the relationship between endpoint $Y$ and marker $S$ is possible in the subpopulation of vaccine recipients, and that marker level $S = s$ may occur within each subpopulation defined by values of the covariates $X$ (positivity).

###### 12.1.1 Conservative (upper bound) inference and sensitivity analysis for the Cox model correlates of risk analysis

We apply the same Cox modeling approach described in Section 9.3.2, augmented with a sensitivity analysis, which harmonizes with the CoR analysis, and sensitivity analysis is generally warranted when a no unmeasured confounders assumption is made. The sensitivity analysis quantifies the rigor of evidence for a controlled VE CoP after accounting for potential bias from unmeasured confounding.

Gilbert et al. (2021) details the inferential and sensitivity analysis approach, which was applied to the CYD14 and CYD15 dengue phase 3 data sets (Moodie et al., 2018); we plan to apply it in the same way to the COVID-19 data sets (as the structure of the problem is the same). We summarize here the essential details needed for application to the COVID-19 data sets.

We define $S$ to be a controlled risk CoP if $P(Y(1, s) = 1)$ is monotone nonincreasing in $s$ with $P(Y(1, s) = 1) > P(Y(1, s') = 1)$ for at least some $s < s'$, where point and 95% confidence interval estimates of $P(Y(1, s) = 1)$ versus $s$, with built in robustness to unmeasured confounding, describe the strength of the CoP in terms of the amount and nature of decrease. Suppose the CoR analysis based on the Cox model is conducted as described in Section 9.3.2.

Let marginalized conditional risk

$$r_M(s) = risk_1(t_F|s)$$

and controlled risk

$$r_C(s) = P(Y(1,s) = 1).$$

Given that CoR analysis is based on observational data — the biomarker value is not randomly assigned — a central concern is that unmeasured or uncontrolled confounding of the association between $S$ and $Y$ could render $r_M(s) \neq r_C(s)$, biasing estimates of the controlled risk curve $r_C(s)$ and of controlled risk ratios of interest

$$RR_C(s_1, s_2) = r_C(s_2)/r_C(s_1) \ .$$

Because we can never be certain that confounding is adequately adjusted for, sensitivity analysis is warranted, as considered in extensive literature — see, e.g., VanderWeele and Ding (2017) and references therein. Sensitivity analysis is useful to evaluate how strong unmeasured confounding would have to be to explain away an observed causal association, that is, to determine the strength of association of an unmeasured confounder between $S$ and $Y$ needed for the observed exposure-outcome association to not be causal, $r_M(s) \neq r_C(s)$. We follow the recommendation of VanderWeele and Ding (2017) to report the E-value as a summary measure of the evidence of causality, or, in our application, evidence of whether $S$ is a controlled risk CoP based on variation in the controlled risk curve. We also include other closely related measures of sensitivity.

The E-value is the minimum strength of association, on the risk ratio scale, that an unmeasured confounder would need to have with both the exposure ($S$) and the outcome ($Y$) in order to fully explain away a specific observed exposure–outcome association, conditional on the measured covariates [VanderWeele and Ding (2017); VanderWeele and Mathur (2020)]. If, as in CoP analyses, the estimated marginalized risk ratio $\widehat{RR}_M(s_1, s_2) = \widehat{r}_M(s_2)/\widehat{r}_M(s_1)$

for $s_1 < s_2$ is less than one, then the E-value for $\widehat{RR}_M(s_1, s_2)$ is calculated as

$$e_{RR}(s_1, s_2) = \frac{1 + \sqrt{1 - \widehat{RR}_M(s_1, s_2)}}{\widehat{RR}_M(s_1, s_2)} . \tag{3}$$

We include the argument $(s_1, s_2)$ in the notation, with $s_1 < s_2$ by convention, to be clear that the E-value depends on specification of two specific marker-level subgroups.

To illustrate the interpretation of an E-value, suppose $S$ is binary and regression analysis yields an estimate $\widehat{RR}_M(0, 1) = \widehat{r}_M(1)/\widehat{r}_M(0) = 0.40$ with 95% confidence interval (CI) $(0.14, 0.78)$. An E-value $e(0, 1)$ of 4.4 means that a marginalized risk ratio $RR_M(0, 1)$ at the observed value 0.40 could be explained away (i.e., $RR_C(0, 1) = 1.0$) by an unmeasured confounder associated with both the exposure and the outcome by a marginalized risk ratio of 4.4-fold each, after accounting for the vector $X$ of measured confounders, but that weaker confounding could not do so.

In addition, we follow the recommendation of VanderWeele and Ding (2017) to also report the E-value $e_{UL}(s_1, s_2)$ for the upper limit $\widehat{UL}(s_1, s_2)$ of the 95% CI for the observed marginalized risk ratio $\widehat{RR}_M(s_1, s_2)$, computed as 1 if $\widehat{UL}(s_1, s_2) \geq 1$ and, otherwise, as

$$\frac{1 + \sqrt{1 - \widehat{UL}(s_1, s_2)}}{\widehat{UL}(s_1, s_2)} ,$$

which in the example equals $e_{UL}(0, 1) = 1.88$. This E-value for the upper limit indicates, for given $s_1 < s_2$, the strength of unmeasured confounding at which statistical significance of the inference that $RR_C(s_1, s_2) < 1$ would be lost. The two E-values above are useful for judging how confident we can be that an immunologic biomarker is a controlled risk CoP, with E-values near one suggesting weak support and evidence increasing with greater E-values.

$RR_C(s_1, s_2) = (1 - CVE(s_2))/(1 - CVE(s_1))$, evidence for $RR_C(s_1, s_2) < 1$ is equivalently evidence for $CVE(s_1) < CVE(s_2)$. Thus in a placebo-controlled trial $RR_C(s_1, s_2)$ can be interpreted as the multiplicative degree of superior vaccine efficacy caused by marker level $s_2$ vs. marker level $s_1$, and E-values equivalently quantify evidence for whether $CVE(s_1)$ differs from $CVE(s_2)$.

It is also useful to provide conservative estimates of controlled risk ratios and of the controlled risk curve, accounting for unmeasured confounding. We approach these tasks based on the sensitivity analysis, or bias analysis, approach of Ding and VanderWeele (2016). We give their main result and refer readers to the paper for details. We begin by defining two (possibly context-specific) fixed sensitivity parameters. First, we set $RR_{UD}(s_1, s_2)$ to be the maximum risk ratio for the outcome $Y$ comparing any two categories of the unmeasured confounders $U$, within either exposure group $S = s_1$ or $S = s_2$, conditional on the vector $X$ of observed covariates. Second, we set $RR_{EU}(s_1, s_2)$ to be the maximum risk ratio for any specific level of the unmeasured confounder $U$ comparing individuals with $S = s_1$ to those with $S = s_2$, with adjustment already made for the measured covariate vector $X$. Thus, $RR_{UD}(s_1, s_2)$ quantifies the importance of the unmeasured confounder $U$ for the outcome, and $RR_{EU}(s_1, s_2)$ quantifies how imbalanced the exposure/marker subgroups $S = s_1$ and $S = s_2$ are in the unmeasured confounder $U$. The values $RR_{UD}(s_1, s_2)$ and $RR_{EU}(s_1, s_2)$ are always specified as greater than or equal to one. We suppose that $RR_M(s_1, s_2) < 1$ for the fixed values $s_1 < s_2$ — this is the case of interest for immune correlates.

Define the bias factor

$$B(s_1, s_2) = \frac{RR_{UD}(s_1, s_2) RR_{EU}(s_1, s_2)}{RR_{UD}(s_1, s_2) + RR_{EU}(s_1, s_2) - 1}$$

for $s_1 \leq s_2$, and define $RR_M^U(s_1, s_2)$ the same way as $RR_M(s_1, s_2)$, except marginalizing over the joint distribution of $X$ and $U$. Then, $RR_M^U(s_1, s_2) \leq RR_M(s_1, s_2) \times B(s_1, s_2)$, where $RR_M^U(s_1, s_2) = E\{r(s_2, X^*)\}/E\{r(s_1, X^*)\}$ with $X^* = (X, U)$ and $r$ conditional risk defined near equation (**??**).Ding and VanderWeele (2016)

Translating this result to our problem context, under the positivity assymption, we have that $RR_M^U(s_1, s_2) = RR_C(s_1, s_2)$ and so, it follows that

$$RR_C(s_1, s_2) \leq RR_M(s_1, s_2) \times B(s_1, s_2) \ . \quad (4)$$

This inequality states that the causal risk ratio is bounded above by the marginalized risk ratio multiplied by the bias factor. It follows that a conservative (upper bound) estimate of $RR_C(s_1, s_2)$ is obtained as $\widehat{RR}_M(s_1, s_2) \times B(s_1, s_2)$, and a conservative 95% CI is obtained by multiplying each confidence limit for $RR_M(s_1, s_2)$ by $B(s_1, s_2)$. These estimates for $RR_C(s_1, s_2)$ account for the presumed-maximum plausible amount of deviation from the no unmeasured confounders assumption specified by $RR_{UD}(s_1, s_2)$ and $RR_{EU}(s_1, s_2)$. An appealing feature of this approach is that the bound (4) holds without making any assumption about the confounder vector $X$ or the unmeasured confounder $U$.

The above approach does not directly provide a conservative estimate of the controlled risk curve $r_C(s)$, because additional information is needed for absolute versus relative risk estimation. To provide conservative inference for $r_C(s)$, we next select a central value $s^{cent}$ of $S$ such that $\widehat{r}_M(s^{cent})$ matches the observed overall risk, $\widehat{P}(Y = 1|A = 1)$. This value is a 'central' marker value at which the observed marginalized risk equals the observed overall risk. Next, we 'anchor' the analysis by assuming $r_C(s^{cent}) = r_M(s^{cent})$, where picking the central value $s^{cent}$ makes this plausible to be at least approximately true. Under this assumption, the bound (4) implies the bounds

$$r_C(s) \leq r_M(s)B(s^{cent}, s) \quad \text{if } s \geq s^{cent} \tag{5}$$

$$r_C(s) \geq r_M(s)\frac{1}{B(s, s^{cent})} \quad \text{if } s < s^{cent}. \tag{6}$$

Therefore, after specifying $B(s^{cent}, s)$ and $B(s^{cent}, s)$ for all $s$, we conservatively estimate $r_c(s)$ by plugging $\widehat{r}_M(s)$ into the formulas (5) and (6). Because $B(s_1, s_2)$ is always greater than one for $s_1 < s_2$, formula (5) pulls the observed risk $\widehat{r}_M(s)$ upwards for subgroups with high biomarker values, and formula (6) pulls the observed risk $\widehat{r}_M(s)$ downwards for subgroups with low biomarker values. This makes the estimate of the controlled risk curve flatter, closer to the null curve, as desired for a sensitivity/robustness analysis.

To specify $B(s_1, s_2)$, we note that it should have greater magnitude for a greater distance of $s_1$ from $s_2$, as determined by specifying $RR_{UD}(s_1, s_2)$ and $RR_{EU}(s_1, s_2)$ increasing with $s_2 - s_1$ (for $s_1 \leq s_2$). We consider one specific approach, which sets $RR_{UD}(s_1, s_2) = RR_{EU}(s_1, s_2)$ to the common value $RR_U(s_1, s_2)$ that is specified log-linearly: $\log RR_U(s_1, s_2) = \gamma(s_2 - s_1)$ for $s_1 \leq s_2$. Then, for a user-selected pair of values $s_1 = s_1^{fix}$ and $s_2 = s_2^{fix}$ with $s_1^{fix} < s_2^{fix}$, we set a sensitivity parameter $RR_U(s_1^{fix}, s_2^{fix})$ to some value above one. It follows that

$$\log RR_U(s_1, s_2) = \left( \frac{s_2 - s_1}{s_2^{fix} - s_1^{fix}} \right) \log RR_U(s_1^{fix}, s_2^{fix}), \quad s_1 \leq s_2.$$

We anchor the sieve analysis by setting $s_1 = s_1^{fix}$ at the $15^{th}$ percentile of the Day 57 antibody marker and $s_2 = s_2^{fix}$ at the $85^{th}$ percentile of the Day 57 antibody marker.

The sensitivity analysis is done for each of the two Cox model CoR analyses described in Section 9.3.2, first for tertiles of the Day 57 marker and second for the quantitative marker. For the former, E-values are reported for both the point estimate and the upper 95% confidence limit for $RR_C(0, 1)$, where category 1 is the upper tertile, category 0 is the lower tertile, and the intermediate middle tertile subgroup of vaccine recipients is excluded from the analysis. In addition, setting $RR_{UD}(0, 1) = RR_{EU}(0, 1) = 2$, such that $B(0, 1) = 4/3$, we report conservative estimation and inference on the causal risk ratio $RR_C(0, 1)$ and equivalently on the ratio of controlled vaccine efficacy curves $(1 - CVE(1))/(1 - CVE(0))$.

Next we repeat the analysis treating $S$ as a quantitative variable, where $P(T \leq t | S = s, X, A = 1)$ is again estimated by two-phase Cox partial likelihood regression and now $RR_M(s_1, s_2)$ is the marginalized risk ratio between $s_1$ and $s_2$. We will plot point and 95% point-wise confidence interval estimates of the observed marginalized risk and controlled risk curves, for the latter using the sensitivity analysis described in Section 12.1.1.

For validity the method requires the positivity assumption, and thus the method will only be applied if the data are reasonably supportive of the positivity assumption. To check positivity, we study the antibody marker distribution in vaccine recipients within each subgroup of the covariates $X$ that are adjusted for. For the tertiles analysis we require evidence that within each subgroup some vaccine recipients have lower tertile responses and some vaccine recipients have upper tertile responses. For the quantitative $S$ analysis, we look for evidence that $S$ varies over its full range within each level of the potential confounders that are adjusted for.

##### 12.2 CoP: Stochastic Interventional Effects on Risk and Vaccine Efficacy

Another approach to studying correlates of protection involves estimating the effect of shifting the immune response marker distribution in the vaccinated individuals (Hejazi et al., 2020a). Specifically, we can consider the effect on risk of a given endpoint of a controlled intervention that shifts the distribution of an immune response by $\delta$ units, where $\delta$ is an analyst-specified real number. Considering a counterfactual scenario in which we are able to intervene so as to modify the immune response induced by the vaccine (e.g., a hypothetical change in dose or other re-formulation of the vaccine), we take this hypothetical intervention to lead to an improved (if $\delta > 0$) or lessened immune response (if $\delta < 0$) relative to the current vaccine (at $\delta = 0$). Using this framework, we can query the counterfactual risk of the endpoint under this hypothetical vaccine. Using notation established above, this quantity can be expressed as the mean of the counterfactual variable $Y(1, S(1) + \delta)$.

This approach is similar to the controlled effects approach described in Section 12.3, but with an important distinction. In the controlled effects approach, one assumes that it is possible to set $S = s$ for *all* individuals in the population. For high values of $s$, this assumption may be unrealistic if the vaccine fails to be strongly immunogenic for some subpopulations. On the other hand, with the interventional approach, it is only required that individuals' immune responses be shifted relative to their observed immune response, which may be more plausible for some vaccines.

Under assumptions (Hejazi et al., 2020a), the main two of which being no unmeasured confounding and positivity (forms of both are also required for the Controlled VE CoP analyses), the counterfactual risk of interest $E[Y(1, S(1) + \delta)]$ is identified by

$$E[P(Y = 1 \mid A = 1, S = S + \delta, X = x) \mid A = 1, X] \ .$$

Examining this quantity across a range of $\delta$ provides insight into the relative contribution of a given immune response marker in preventing the endpoint of interest.

Hejazi et al. (2020a) proposed nonparametric estimators that rely on estimates of the outcome regression (as described above) and the conditional density of the immune response marker in vaccinated participants. Their estimators efficiently account for two-phase sampling of immune responses and are implemented in the `txshift` package (Hejazi and Benkeser, 2020) for the `R` language and environment for statistical computing (R Core Team, 2020), available via both GitHub at `https://github.com/nhejazi/txshift` and the Comprehensive `R` Archive Network at `https://CRAN.R-project.org/package=txshift`.

These estimators will be applied to each of the five Day 57 antibody markers (without baseline adjustment) controlling for the same set of baseline risk factors that are controlled for in other analyses previously discussed. As with the mediation analysis approach described in Section 12.3, the procedure will leverage low-dimensional risk factors alongside parametric regression strategies and flexible conditional density estimators for endpoints with fewer than 100 observed cases (pooling over the randomization arms); however, more flexible learning techniques will be employed for modeling the outcome process for endpoints with a greater number of observed cases.

In particular, conditional density estimates of immune response markers will be principally based on a nonparametric estimation strategy that reconstructs the conditional density through estimates of the conditional hazard of the discretized immune response marker values (Hejazi et al., 2020a,d,c); this approach is an extension of the proposal of Díaz and van der Laan (2011). A Super Learner ensemble (van der Laan et al., 2007) of variants of this nonparametric conditional density estimator and semiparametric conditional density estimators based on Gaussinization of residuals will be constructed using the `sl3` `R` package (Coyle et al., 2020). In settings with limited numbers of case endpoints, the outcome process will be modeled as a Super Learner ensemble of a library of parametric regression techniques (as recommend by

Gruber and van der Laan, 2010), while the library will be augmented with flexible regression techniques — including, for example, lasso and ridge regression (Tibshirani, 1996; Tikhonov and Arsenin, 1977; Hoerl and Kennard, 1970), elastic net regression (Zou and Hastie, 2003; Friedman et al., 2009), random forests (Breiman, 2001; Wright et al., 2017), extreme gradient boosting machines (Chen and Guestrin, 2016), light and efficient gradient boosting machines (Ke et al., 2017), multivariate adaptive polynomial and regression splines (Friedman et al., 1991; Stone et al., 1994; Kooperberg et al., 1997), and the highly adaptive lasso (van der Laan, 2017; Benkeser and van der Laan, 2016; Hejazi et al., 2020b) — as the number of endpoint cases grows. These algorithm libraries will be coordinated to match those used in other CoP analyses. For Moderna COVE the more flexible algorithms are not used given the limited number of vaccine breakthrough COVID endpoints.

Additionally, we recall that $P(Y(0) = 1) = P(Y = 1 \mid A = 0)$ (in view of vaccine versus placebo randomization, as stated previously in Section 12.1) and may be estimated in the same way as for the analysis of controlled vaccine efficacy, thus yielding an estimate of stochastic interventional VE defined by

$$SVE(\delta) = 1 - \frac{E[P(Y = 1 \mid A = 1, S = S + \delta, X = x) \mid A = 1, X]}{P(Y(0) = 1)}.$$

Output of the analyses will be presented as point and 95% point-wise confidence interval estimates of $E[Y(1, S(1) + \delta)]$ and of $SVE(s)$ over the values of $s$ for each of the Day 57 antibody markers, for each of a range of $\delta$ spanning -2 to 2 on the standard unit scale for each antibody marker.

Lastly, just as for the controlled VE CoP analyses, these analyses will only be performed if diagnostics support plausibility of the positivity assumption. Importantly, however, the positivity assumption for the stochastic interventional effects differs from that usually required. That is, where the positivity assumption for effects defined by static interventions requires a positive probability of treatment assignment across all strata defined by baseline factors (i.e., that a discretized immune response value be possible regardless of baseline factors), the positivity assumption of these effects is

$$s_i \in \mathcal{S} \implies s_i + \delta \in \mathcal{S} \mid A = 1, X = x$$

for all $x \in \mathcal{X}$ and $i = 1, \ldots n$. In particular, this positivity assumption does not require that the post-intervention exposure density, $q_{0,S}(S - \delta \mid A = 1, X)$, place mass across all strata defined by $X$. Instead, it requires that the post-intervention exposure mechanism be bounded, i.e.,

$$P\{q_{0,S}(S - \delta \mid A = 1, X)/q_{0,S}(S \mid A = 1, X) > 0\} = 1,$$

which may be readily satisfied by a suitable choice of $\delta$.

More importantly, the static intervention approach may require consideration of counterfactual variables that are scientifically unrealistic. Namely, it may be inconceivable to imagine a world where every participant exhibits high immune responses, given the phenotypic variability of participants' immune systems. This too may be resolved by considering an intervention $\delta(X)$, allowing the choice of $\delta$ to be a function of baseline covariates $X$ (Hejazi et al., 2020a; Díaz and van der Laan, 2012; Haneuse and Rotnitzky, 2013; Díaz and van der Laan, 2018).

The current COVE immune correlates manuscript does not include stochastic intervention vaccine efficacy analyses.

##### 12.3 CoP: Mediation of Vaccine Efficacy

Using mediation methods, we can decompose the overall VE into so-called *natural* direct and indirect effects. We will estimate this decomposition for each Day 57 antibody marker individually (focusing on the non-baseline subtracted markers as for the other CoP analyses described above), as well as when considering all antibody markers together (although this SAP currently restricts to analysis of the individual markers).

For simplicity, as before, we describe this approach using a binary outcome, noting that extensions to time-to-event (with competing risks) are possible. The *total* effect of the vaccine can be represented by one minus the risk ratio

$$\mathrm{RR} = \frac{P(Y(1, S(1)) = 1)}{P(Y(0, S(0)) = 1)} \ .$$

The natural direct and indirect effects are, respectively,

$$\text{RR}_{DE} = \frac{P(Y(1, S(0)) = 1)}{P(Y(0, S(0)) = 1)} \text{ and } \text{RR}_{IDE} = \frac{P(Y(1, S(1)) = 1)}{P(Y(1, S(0)) = 1)} \ .$$

Note that $\text{RR} = \text{RR}_{DE}\text{RR}_{IDE}$, showing that the total effect decomposes into the direct times indirect effect. Another quantity of interest is the proportion mediated, which we express as

$$\text{PM} = 1 - \frac{\log(\text{RR}_{DE})}{\log(\text{RR})} \ .$$

We note that PM=1 if and only if $\text{RR}_{DE} = 1$, i.e., no direct effect means that the marker fully mediates VE. We will estimate PM defined in this way.

As above, we must assume all confounders $X$ of $S$ and $Y$ have been measured. We also assume there are no confounders of the mediator-outcome relationship that are affected by treatment. Moreover, we require an overlap assumption that

$$P(S = s|A = 0, X = x) > 0 \text{ implies } P(S = s|A = 1, X = x) > 0 \qquad (7)$$

for all subgroups $X = x$ (i.e., a.e.). Under these assumptions, $P(Y(a, S(a') = 1)$ is identified by

$$E[P(Y = 1 \mid A = a, S, X)|A = a', X] \ .$$

In our immune CoP application it is expected that, for analyses restricting to baseline negative individuals, the conditional density of the immune response marker in the placebo arm will be a point mass at 0, that is with $S$ below the LLOD. In other words, we do not expect any placebo recipients to have a positive value of the immune response marker. This implies the identification result that for $a = 0, 1$, $P(Y(a, S(0)) = 1) = E[P(Y = 1 \mid A = a, S = 0, X)]$. While $P(Y(0, S(1) = 1)$ is not identified, it is not necessary to estimate this term in order for estimation of the parameters of interest (natural direct effect, natural indirect effect, PM).

For a highly immunogenic vaccine, it may be the case that the needed overlap assumption (7) will be violated. This could happen, for example if each baseline negative placebo recipient has antibody marker value below the assay's

LLOD (which is expected), and every vaccine recipient has antibody marker value above the LLOD. We will only include antibody markers for mediation analysis if at least 10% of vaccine recipients have marker value equal to the value in placebo recipients.

Benkeser et al. (2021) provide a multiply robust targeted minimum loss-based plug-in estimator of natural direct and indirect effects that is appropriate for case-cohort sampling. The estimator requires estimation of several regressions, which are used in an augmented inverse probability of treatment weighted estimator. The propensity score will be estimated by a main terms logistic regression model to account for chance imbalances across randomization arms. The sequential outcome regressions used by the approach will be based on a super learner with the 14 algorithms listed in Table 6.

Table 6: Learning Algorithms in the super learner Library for mediation methods[1].

| Algorithms | Screens[2]/ Tuning Parameters |
|---|---|
| SL.mean | All |
| SL.glm | Low-collinearity and (All, Lasso, LR) |
| SL.glm.interaction | (All, Lasso, LR) |
| SL.gam | Low-collinearity and (Lasso, LR) |
| SL.glmnet | All |
| SL.xgboost | All |
| SL.ranger | All |

[1] some nuisance parameters have binary outcomes, others quantitative. For the former, we used `family = binomial()` input to the `SuperLearner` function; for the latter, we used `family = gaussian()`.

[2] **All** = include all variables; **Lasso** = include variables with non-zero coefficients in the standard implementation of SL.glmnet that optimizes the lasso tuning parameter via 10-fold cross-validation; **Low-collinearity** = do not allow any pairs of quantitative variables with Spearman rank correlation $> 0.90$; **LR** = Univariate logistic regression Wald test 2-sided p-value $< 0.10$.

The estimator is implemented in the `natmed2` package available on GitHub (https://github.com/benkeser/natmed2). The baseline covariates $X$ adjusted for are the same as for the other analyses (e.g. of CoR and of controlled vaccine efficacy).

### 13 Summary of the Set of CoR and CoP Analyses and Their Requirements and Contingencies, and Synthesis of the Results, Including Reconciling Any Possible Contradictions in Results

Table 7 summarizes all of the Stage 1 correlates analyses of Day 29 and Day 57 antibody markers that are done, including contingencies for whether and when each analysis is done. All of the Day 29 and Day 57 markers are the versions that are not baseline subtracted, given that the cohort for analysis is baseline negative. Most of the analyses focus on univariate Day 29 and Day 57 markers. The primary reason to do this is the goal to identify a parsimonious correlate based on a single marker without needing to run the set of assays, and secondary reasons are: (1) the assay readouts are expected to be highly correlated, especially for the cID50 and cID80 readouts, and (2) there is ample precedent for univariate markers being accepted as immunological surrogate endpoints for approved vaccines (Plotkin, 2010).

Table 7: Summary of Stage 1 Day 57 Marker CoR and CoP Analyses with Requirements/Contingencies for Conduct of the Analysis (Same Considerations Apply for Day 29 Markers)

| | Structure | Requirements/Contingencies | |
|---|---|---|---|
| | of | Min No. Vaccine | |
| **Analysis** | **Day 57 Marker(s)** | **Endpoints** | **Other** |
| CoR Cox Model | Tertiles of $S^1$ | 25 | None |
| | Quant. $S = s^2$ | 25 | None |
| | Quant. $S \geq s^1$ | 25 | None |
| CoR Nonpar. threshold | Quant. $S \geq s^1$ | 35 | None |
| CoR GAM | Quant. $S = s^2$ | 35 | None |
| CoR Superlearner[3] | Quant. $S = s$, 2FR, 4FR | 35 | None |
| CoP: Correlates of VE | Binary $S$ | 50 | None |
| | Quant. $S = s$ | 50 | BIP with $R^2 \geq 0.25$ |
| CoP: Controlled VE | Quant. $S = s$ | 50 | Feasibility of positivity[4] |
| | Tertiles of $S = s$ | 50 | Feasibility of positivity[4] |
| CoP: Stoch. Interv. VE | Quant. $S = s$ | 50 | Feasibility of positivity[4] |
| CoP: Mediators of VE | Quant. $S = s$ | 50 | Feasibility of positivity[4] |

[1]These analyses are harmonized in addressing the same scientific question of how does endpoint risk vary over vaccinated subgroups defined by $S$ above a threshold.
[2]These exploratory supportive analyses are harmonized in addressing the same scientific question of how does endpoint risk vary over vaccinated subgroups defined by $S$ equal to a given marker value.
[3]Only this Superlearner analysis uses data from multiple assays and multiple readouts as input features; the other analyses consider one Day 57 biomarker at a time. [4]The positivity assumptions are as follows. Controlled VE: $P(S = s \mid A = 1, X) > 0$ almost surely. Stochastic Interventional VE: $s_i \in \mathcal{S} \implies s_i + \delta \in \mathcal{S} \mid A = 1, X = x$ for all $x \in \mathcal{X}$ and $i = 1, \ldots n$. Mediators of VE: $P(S = s \mid A = 1, X) > 0$ almost surely and $P(S = s|A = 0, X = x) >$ implies $P(S = s|A = 1, X = x) > 0$. The quantitative analysis will require that the largest value $S$ observed in the placebo is larger than the smallest value of $S$ observed in the vaccine recipients. This assumption would naturally be satisfied for the tertiles analysis. For quantitative $S$, the assumption is weaker for the Stochastic Interventional VE analysis, such that it is possible that only this analysis of the three will be done.

Some of the analyses include parametric assumptions for characterizing associations (Cox model and threshold analyses, Cox model versions of Controlled VE analyses) and others are nonparametric or approximately so (all other analyses). If parametric and nonparametric analyses of the same type (e.g., Cox model vs. nonparametric CoR analysis of the same association parameter; Controlled VE Cox model vs. nonparametric monotone dose-response) suggest contradictory results, then the interpretation from the nonparametric analysis will be prioritized, given it is more robust and less likely to be an incorrect result. The diagnostic testing of the parametric assumptions will aid this interpretation. As noted above, if the nonparametric analysis suggesting a contradictory result requires a positivity assumption, then its results will only be prioritized if diagnostics support feasibility of the positivity assumption.

##### 13.1 Synthesis Interpretation of Results

To structure the interpretation of the whole set of CoR and CoP results, we consider the Bradford-Hill criteria for supporting causality assessments:

1. Temporal sequence of association (vaccination causes generation of antibodies, which precede occurrence of the clinical disease outcome)
2. Strength of association (CoR magnitude)
3. Consistency of association (across studies and methods)
4. Biological gradient (may be interpreted as dose-response with greater Day 57 antibody corresponding to lower risk and greater VE)
5. Specificity (that the antibody marker is induced by vaccination not natural infection, and the antibody impacts the particular clinical endpoint being analyzed)
6. Plausibility [(supported by other COVID vaccines through study in efficacy trials and challenge (animal or human) trials, and by other potential studies such as natural history re-infection studies and monoclonal antibody prevention efficacy studies that could be challenge (animal or human) or field trials])
7. Coherence (the causality assumption does not appear to conflict with current knowledge)
8. Experimental reversibility (if VE wanes to a low level then the antibody marker also wanes coincidently; if the Day 57 marker is a strong correlate for outcome during the period of high VE, then it becomes a weaker

correlate against endpoints occurring during the later period of low VE; also could be supported if vaccine breakthrough cases tend to occur early in follow-up when antibody levels are known to be relatively low)

9. Analogy (supported by other respiratory virus vaccines, and natural history studies or challenge studies of other respiratory virus vaccines)

We discuss evaluation of these criteria for Day 57 markers, where the same evaluations accounting for Day 29 markers are similarly relevant.

On temporal sequence, because the analyses are done in baseline negative individuals, generally the Day 57 antibody responses must be generated by the vaccine, and if the outcome occurs well after Day 57, then there is clear temporal ordering of vaccination causing antibodies followed by outcome. The nuance is outcome cases with event times near 7 days post Day 57, some of which could have been infected with SARS-CoV-2 prior to Day 57 and have relatively long incubation periods, possibly perturbing temporal ordering by creating naturally-induced rather than vaccine-induced antibody. However, the knowledge about the distribution of the time period between SARS-CoV-2 acquisition and symptomatic COVID, and the time needed for an infection to create an adaptive immune response, suggests that this issue could only haves a minor impact, and overall the temporal sequence criterion readily holds.

On strength of association, this is directly quantified in all of the analyses as a core output of each method, quantified by point estimates and confidence interval estimates of covariate-adjusted association parameters or causal effect parameters.

On consistency of association, checking for similar estimates and inferences across the multiple vaccine efficacy trials will be relevant. The fact that all of the tested vaccines are designed to protect through induction of antibody to Spike protein suggest that consistency is plausible. The vaccine platform needs to be accounted for in this evaluation, where consistency may be expected for vaccines of a given type (e.g., mRNA vaccines, Spike protein vaccines, viral vector vaccines with a similar vector), whereas across types a consistent body of evidence would be very helpful, but not a requirement.

FDA guidance has stipulated that a surrogate endpoint for one vaccine platform is not necessarily expected to hold for another, and that evidence for one platform would not be seen on its own as support for a surrogate endpoint for another.

Moreover, consistency of association may be assessed in another sense - by studying whether the different CoR methods tend to reveal a consistent directionality and pattern of an antibody marker correlated with risk, and whether the different CoP methods tend to reveal a consistent directionality and pattern of an antibody marker connected to vaccine efficacy (as measured by the various causal effect parameters) and with different versions of vaccine efficacy. A common core element of all of the CoR and CoP methods is covariate-adjusted estimation of marker-conditional risk in vaccine recipients, e.g. of marginal conditional risk $E_X[P(T \leq t_F|S = s, A = 1, X)]$ or $E_X[P(T \leq t_F|S \geq s, A = 1, X)]$. Generally, if an estimate of this function shows strongly decreasing risk with $s$, then likely all of the CoR analyses will detect such a decrease, and the CoP analyses will detect a version of vaccine efficacy increasing in $s$. A nuance in looking for consistency of results across methods stems from the fact that different methods have different power to detect the same effect; because of this fact, consistency in magnitude (point estimate) and directionality are more important than consistency in inference/statistical significance.

The fact that all of the methods adjust for the same set of baseline covariates $X$ will aid the ability to compare the results across methods in an interpretable manner. This discussion highlights the relevance of adjusting for the same set of baseline covariates across the different efficacy trials, although our choice to do covariate-adjustment through marginalization (rather than through conditional association parameters) lends some resilience to this issue.

Our comments on consistency of association have supposed a given study endpoint, such as COVID. Another dimension of consistency evaluation could include comparing results across endpoints. On the one hand, consistency in evidence across endpoints could strengthen the case for a CoP, especially for endpoints in the same 'class' such as moderate disease and severe disease. On the other hand, the greater the difference between endpoints, the less relevant consistency may be, because the vaccine may protect through different mechanisms against each endpoint (one potential example is prevention of asymptomatic infection vs. prevention of severe disease). Thus evidence for a CoP for a given endpoint should not necessarily be down-graded based on evidence that the same marker does not appear to be a CoP for another endpoint.

On biological gradient, many of the methods are flexible and designed to detect a dose-response pattern of antibody with risk or antibody with vaccine efficacy, with tabular and graphical output of point and confidence interval estimates designed to reveal dose-response.

On specificity, as noted above antibodies generally are almost surely vaccine-induced given the analysis is done in baseline negative individuals, although with nuance that care is needed to evaluate whether some vaccine breakthrough cases may have had SARS-CoV-2 acquisition unusually early in follow-up (e.g., prior to second vaccination). In addition, the assays are validated for measuring specific anti-SARS-CoV-2 antigen response. Moreover, the Day 57 antibody markers can be verified to be negative in all or almost all baseline negative placebo recipients. Therefore, the specificity criterion should readily hold, with the proviso of the complication of the possible inclusion of unusually early infections as vaccine breakthrough cases in some analyses.

On coherence, the results will be interpreted in the light of knowledge of immune correlates of protection for the same vaccine in animal challenge studies (and human challenge studies as available), where multiple studies have demonstrated that both binding and neutralizing antibodies are a correlate of protection.

The results will also be interpreted in light of any knowledge available on passively administered SARS-CoV-2 monoclonal antibodies for prevention of SARS-CoV-2 infection or COVID disease, either in challenge studies (animals or humans) or efficacy trials. In addition, the results will be interpreted in light of results on the antibody markers as correlates of re-infection in natural history studies. Note we are cautious to not use correlates studies in already-infected individuals, because the fact of infection may readily change the nature of a correlate of protection.

On experimental reversibility, in future analyses we will evaluate whether the strength of association of the Day 57 CoRs and CoPs weakens when restricting to outcomes occurring more distal to vaccination. If the vaccine efficacy is found to wane over time, and the antibody marker wanes over time, then this decrease in the strength of association would be consistent with antibody as a correlate of protection. In contrast, if vaccine efficacy and antibody waned over time, but the strength of a Day 57 CoR and CoP was the same regardless of the timing of outcomes, it might call into question the role of the antibody marker as a CoP. The Stage 2 correlates analyses will also be helpful, where experimental reversibility could be supported simply by coincident waning of VE and waning antibody.

Experimental reversibility may also be supported by "population-level" correlates analyses, a term sometimes used in reference to meta-analysis that associates the level of VE with the population-level of a Day 57 marker across subgroups or trials; e.g. the population-level Day 57 marker response may be summarized by the geometric mean titer or geometric mean concentration. Future analyses of multiple phase 3 trial data sets will apply meta-analysis surrogate endpoint evaluation methods.

On analogy, perhaps the most relevant vaccines to consider are vaccines against other respiratory viruses, including influenza vaccine and RSV vaccines. The fact that neutralizing antibodies are a CoR and CoP for both inactivated and live virus vaccines supports that neutralizing antibodies can be a CoP for SARS-CoV-2. In addition, there is ongoing correlates of protection analysis of Novavax's Phase 3 RSV vaccine efficacy trial, that is evaluating binding antibody and neutralizing antibody CoRs and CoP correlates for severe respiratory disease in infants of vaccinated pregnant mothers (submitted). Once those results are available, they will aid in checking the analogy (and coherence) criterion.

The univariate CoR analyses assess five Day 57 antibody biomarkers. The questions arise as to how do we select which biomarker seems to be the best-supported CoP, and do we need to be concerned about multiplicity adjustment issues? Given the multifactorial nature of the assessment involving biology and statistics, we for the most part avoid an approach that tries to pre-specify a quantitative ranking system; rather our approach presents the results of each marker side by side and allows human synthesis and interpretation. To guard against errors in this subjective process, we suggest that consistent results across analyses of a given trial, and consistent results (and predictive validation) across multiple trials, will provide particularly strong guidance for interpreting results. For example, if a particular Day 57 antibody marker shows remarkably consistent results in being a strong CoR and supported CoP but the other readouts do not, it may emerge as the best-supported CoP. In addition, the superlearning CoR estimated optimal surrogate objective has a special place of importance, because it includes variable importance quantification, providing some quantitative guidance on ranking the predictivneness of markers. This variable importance will be defined both internal to a given trial and based on external validation on the other efficacy trials. The metrics of CV-AUC and AUC on new trials quantifies evidence for signal in the data in a way that is protected from risk of false positive results, by virtue of having two layers of cross-validation used to estimate CV-AUC and hence avoid over-fitting. In addition, the CoR analyses use multiple hypothesis testing adjustment to help ensure clear signals and not false positive results (see Section 9.4.1). We also need a plan for minimizing the risk of false positive results for CoP analyses, which we now address.

##### 13.2 Multiple Hypothesis Testing Adjustment for CoP Analysis

For the univariable CoP analyses of the prioritized set of Day 29 and Day 57 antibody markers among the four specified marker variables, the analysis plan seeks evidence of a CoP through four different causal effect approaches. Because of this looking for evidence through different lenses, for CoP analysis we do not focus on family-wise error rate adjustment, because FWERadjustment aims to control the risk of making even a single false rejection. Rather, in an effort to build a body of consistent evidence and to ensure that a large fraction of that evidence is reliable, for CoP analysis we focus on false discovery rate correction. To do this, we use the same permutation-based method (Westfall et al., 1993) that is used for CoR analysis. The multiplicity adjustment is performed across the Day 29 and Day 57 markers and across the set of CoP methods that are applied, in a single suite of hypothesis tests with calculation of q-values. As a guideline for interpreting CoP findings (but not meant to be a rigid gateway), markers with unadjusted p-value $\leq 0.05$ and q-value $\leq 0.10$ are flagged as having statistical evidence for being a CoP.

### 14 Estimating a Threshold of Protection Based on an Established or Putative CoP (Population-Based CoP)

For each antibody marker studied as a CoP, we will apply the Chang-Kohberger (2003) / Siber (2007) method to estimate a threshold of the antibody marker associated with the estimate of overall vaccine efficacy observed in the trial.

This method makes two simplifying assumptions: (1) that a high enough antibody marker value $s^*$ implies that individuals with $S > s^*$ have essentially zero disease risk (perfect protection) regardless of whether they were vaccinated; and (2) $P(Y = 1|S \leq s^*, A = 1)/P(Y = 1|S \leq s^*, A = 0) = 1$ (zero vaccine efficacy if $S \leq s^*$). Based on these assumptions, $s^*$ is calculated as the value equating $1 - \widehat{P}(S \leq s^*|A = 1)/\widehat{P}(S \leq s^*|A = 0)$ to the estimate of overall vaccine efficacy. This estimate is supplemented by estimating the reverse cumulative distribution function (RCDF) of $S$ in baseline negative vaccine recipients and calculating a 95% confidence interval for the threshold value $s^*$ as the points of intersection of the estimated RCDF curve with the 95% confidence interval for overall vaccine efficacy (as in the figure in Andrews and Goldblatt, 2014).

This method essentially assumes that $S$ has already been established as a CoP, and under that assumption estimates a threshold that may be considered as a benchmark / study endpoint for future immunogenicity vaccine trial applications.

It is acknowledged that this approach makes simplifying assumptions that are diagnosed to be violated in the COVE trial; nonetheless it may yield a useful benchmark and complementary information on a threshold correlate of protection.

### 15 Considerations for Baseline SARS-CoV-2 Positive Study Participants

As stated above, if enough COVID cases in baseline positive vaccine and/or placebo recipients occur, then additional correlates analyses may be planned in baseline positive individuals. For example, the same or similar correlates of risk analysis plan that is used to analyze Day 29 and Day 57 marker correlates of risk in baseline negative vaccine recipients could be applied to assess Day 1 marker correlates of risk in baseline positive placebo recipients. In addition, analyses could be done to assess how vaccine efficacy in baseline positive participants varies with Day 1 markers. It is straightforward to make this analysis rigorous because Day 1 markers are a baseline covariate, such that regression analyses are valid based on the randomization.

### 16 Avoiding Bias with Pseudovirus Neutralization Analysis due to Use of Anti-HIV Antiretroviral Drugs

Because the lentivirus-based pseudovirus neutralization assay uses an HIV backbone, the presence of anti-retroviral drugs in serum will give a false positive neutralization signal. This can be easily screened for using an MuLV pseudotype control. Therefore, Day 1, Day 29, and Day 57 samples of all study participants with data included in correlates analyses will be tested for presence of anti-retroviral drugs. Participants with any of the samples at Day 1, 29, 57 positive for antiretroviral use are excluded from analyses, for all analyses that include pseudovirus neutralization. Analyses that do not consider pseudovirus neutralization are unaffected by this issue.

#### 17 Accommodating Crossover of Placebo Recipients to the Vaccine Arm

After the primary efficacy endpoint was met per the protocol-defined interim analysis, supporting the issuance on December 18, 2020 of an Emergency Use Authorization (EUA) from the FDA for the mRNA-1273 vaccine, mRNA-1273 vaccination was offered to participants who originally received placebo so that they could have the potential benefit of vaccination against COVID-19 [Moderna (2020)].

For crossed-over placebo recipients who have study visits and blood sample storage on the same schedule as if they had originally been assigned to the vaccine arm, follow-up data from the crossed over placebo recipients will be included in the correlates of risk analyses, which is expected to yield improved power and precision given the expanded sample size of vaccine recipients.

However, correlates of protection will only be assessed over follow-up through to the point that there is no longer a placebo cohort under blinded follow-up. Moreover, if immune marker data from crossed-over placebo recipients are available, then correlate of VE CoP analyses will be conducted that leverage the additional closeout placebo vaccination data.

The current manuscript restricts to the primary blinded follow-up period.

A

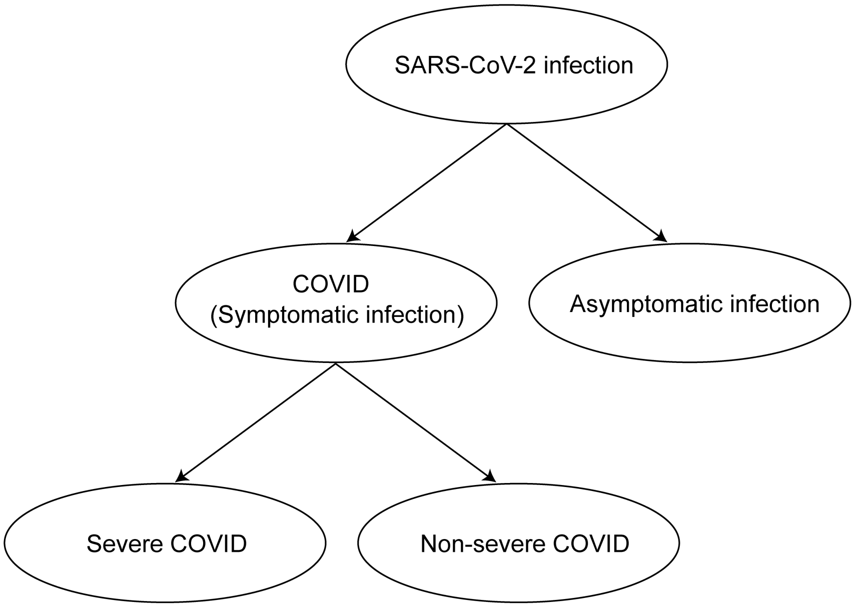

B

| **Clinical Endpoint** | **Definition** |
|---|---|
| SARS-CoV-2 infection | Positive RNA PCR test or SARS-CoV-2 seroconversion*, whichever occurs first |
| COVID (Symptomatic infection) | Meeting a protocol-specified list of COVID-19 symptoms with virological confirmation of SARS-CoV-2 infection (symptom triggered) |
| Asymptomatic infection | SARS-CoV-2 seroconversion* without prior diagnosis of the COVID endpoint¶ |
| Severe COVID | COVID endpoint with at least one protocol-specified severe disease event |
| Non-severe COVID | COVID endpoint with zero protocol-specified severe disease events |

*Seroconversion is assessed via a validated assay that distinguishes natural vs vaccine-induced SARS-CoV-2 antibodies

¶ Alternatively, the asymptomatic infection endpoint can also include an RNA PCR+ test result obtained through testing regardless of symptoms (e.g., as a requirement for travel, return to school or work, or elective medical procedures) and follow-up to confirm the participant remains asymptomatic

Figure 1: A) Structural relationships among study endpoints in a COVID-19 vaccine efficacy trial (Mehrotra et al., 2020). B) Study endpoint definitions.

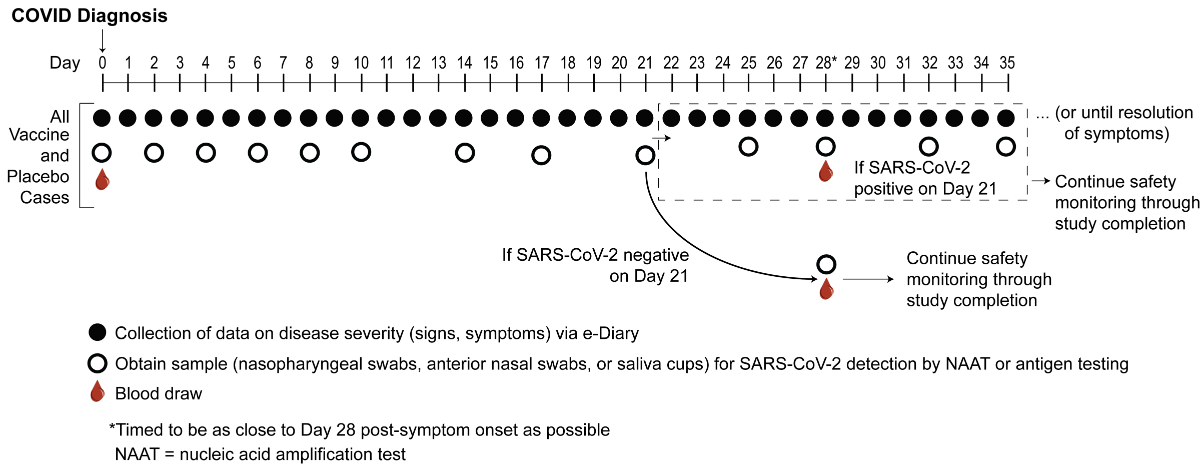

Figure 2: Example at-COVID diagnosis and post-COVID diagnosis disease severity and virologic sampling schedule, in a setting where frequent follow-up of confirmed cases can be assured. Participants diagnosed with virologically-confirmed symptomatic SARS-CoV-2 infection (COVID) enter a post-diagnosis sampling schedule to monitor viral load and COVID-related symptoms (types, severity levels, and durations).

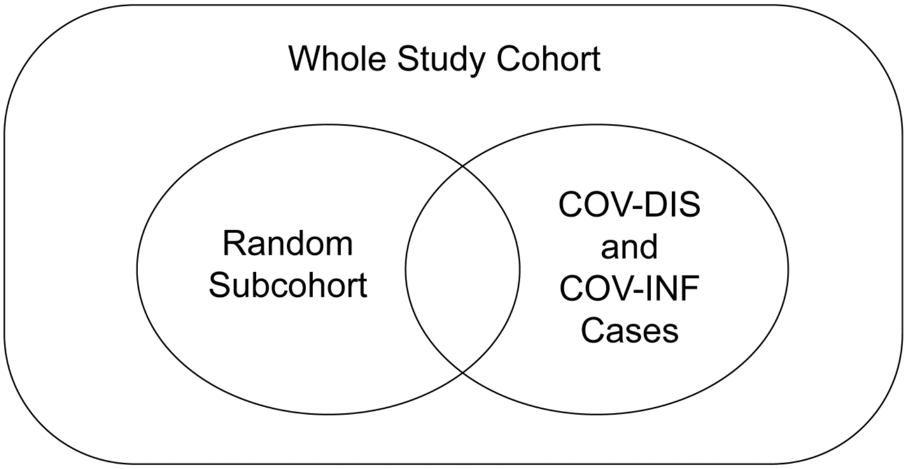

Figure 3: Case-cohort sampling design (Prentice, 1986) that measures Day 1, 29, 57 antibody markers in all participants selected into the subcohort and in all COVID and COV-INF cases occurring outside of the subcohort.

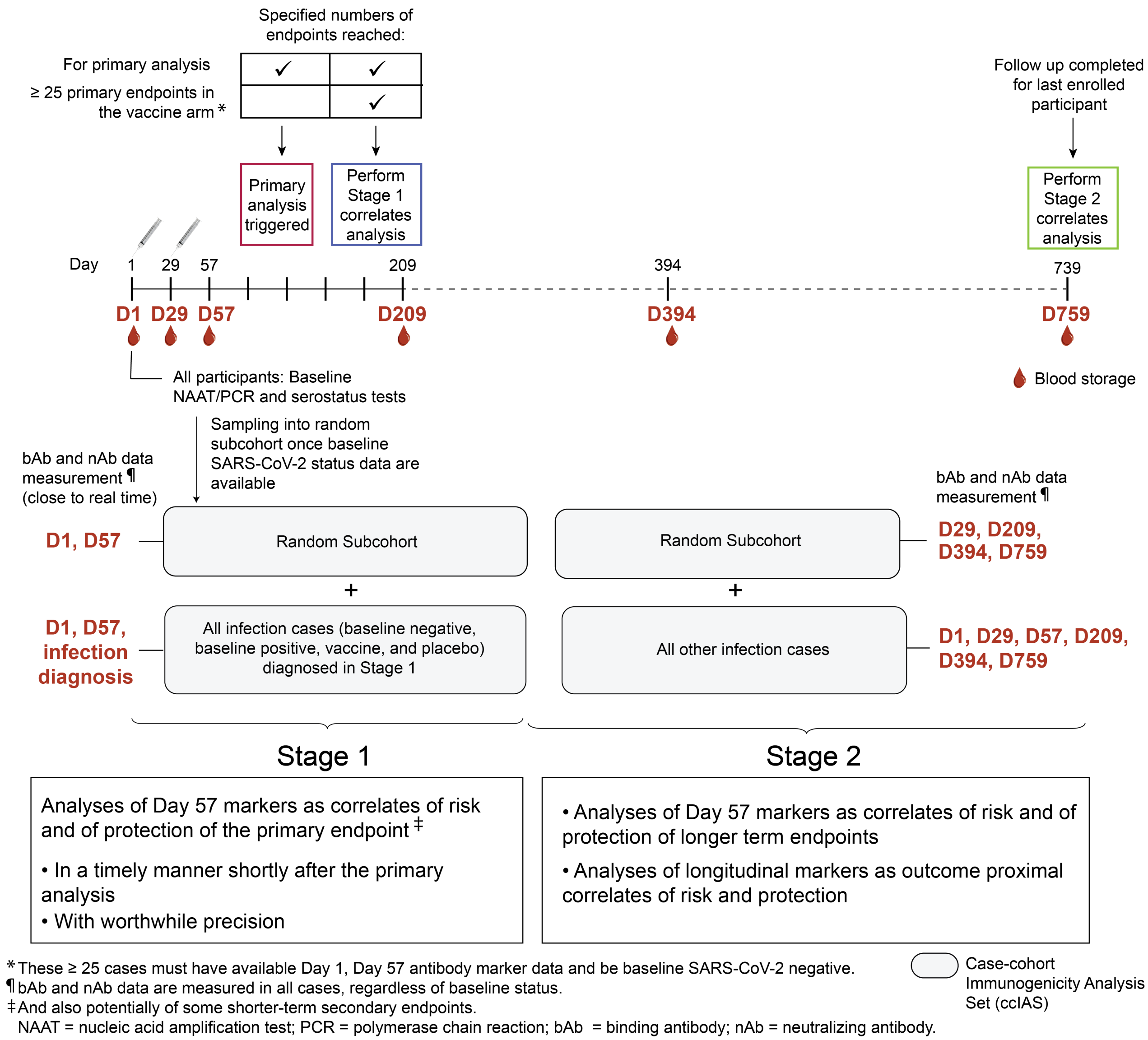

Figure 4: Two-stage correlates analysis. Stage 1 consists of analyses of Day 29 and Day 57 markers as correlates of risk and of protection of the primary endpoint and potentially also of some secondary endpoints, and includes antibody marker data from all COVID and SARS-CoV-2 infection cases (COV-INF) through to the time of the data lock for the first correlates analyses. Stage 2 consists of analyses of Day 29 and Day 57 markers as correlates of risk and of protection of longer term endpoints and analyses of longitudinal markers as outcome-proximal correlates of risk and of protection, and includes antibody marker data from all subsequent COVID and COV-INF cases. Stage 1 measures Day 1, 29, 57 antibody markers and COV-INF and COVID diagnosis time point markers; Stage 2 measures antibody markers from all sampling time points and COV-INF plus COVID diagnosis sampling time points not yet assayed. The same immunogenicity subcohort is used for both stages.
